# Faster peak alpha frequency precedes higher menstrual pain in adolescence

**DOI:** 10.64898/2026.10.20.26366046

**Authors:** Natalie R. Osborne, Frank F. Tu, Sarah E. Darnell, Matthew J. Kmiecik, Kevin M. Hellman

## Abstract

Brain differences in chronic pain are almost always measured after years of pain experience, leaving unresolved whether they create vulnerability to pain or result from it. Uniquely, menstrual pain has a developmentally scheduled onset (menarche, the first menstrual period), allowing the brain to be characterized before and after pain exposure occurs. Here we used longitudinal 64-channel EEG to measure resting brain activity in adolescents once before (n=250) and twice after (n=161, n=140) menarche. Before their first period, adolescents who later reported higher menstrual pain (≥30 on 0-100 VAS) already showed faster occipital peak alpha frequency compared to those who reported lower menstrual pain, a difference that persisted in the early (3 – 14 months) post-menarche time period. In contrast, oscillatory power and functional connectivity did not differ by menstrual pain group, suggesting that the widespread functional brain alterations reported in adults with painful periods take years to emerge. Peak alpha frequency increased, and theta power and connectivity decreased across the menarchal transition, consistent with typical adolescent maturation. However, longitudinal analyses stratified by higher and lower menstrual pain group suggested group specific timings for these neurodevelopmental changes. Overall, these findings suggest that early menstrual pain is not associated with widespread alterations in neural oscillations, but aspects of menstrual pain susceptibility may be reflected in resting peak alpha frequency. To our knowledge, this is the first neurophysiological trait shown to precede the onset of menstrual pain.

## 1. Introduction

Menarche, the first menstrual period, marks a key biological milestone of puberty. Earlier age at menarche [31] and the presence of painful menstruation (dysmenorrhea) in adolescence [55] are both associated with an increased risk for future chronic pain. The brains of pre vs. post-menarche girls are distinguishable by machine learning, suggesting menarche-specific influences on adolescent brain features [17]. While there is growing evidence for puberty-specific effects on brain maturation [10] and their potential implications for subsequent health conditions [50], it is unknown whether menarche is a *critical window* for the neurodevelopment of chronic pain. Understanding how the adolescent brain changes across the menarchal transition and the influence of early menstrual pain on functional brain organization could identify targetable mechanisms that may reduce future pain risk.

Across adolescence, peak alpha frequency (PAF) increases to reach adult levels of approximately 10 Hz [16]. Resting-state PAF is a stable neural trait [18] that has been proposed as a predictive marker of individual pain sensitivity and therefore susceptibility to chronic pain [13]. A slower PAF is linked with increased risk for developing pain after surgery [41] or an experimental prolonged pain paradigm [8], while brain stimulation targeted to increase PAF can subsequently reduce pain [9]. Slower PAF has been reported in some [14,23,38] but not all chronic pain conditions [62]. Importantly, the PAF-pain relationship has almost exclusively been studied in adults, so whether it exists in adolescence is not clear.

Although brain changes specific to menarche and the onset of menstrual pain are unknown, studies in adults with dysmenorrhea have identified structural and functional brain alterations in areas commonly implicated in chronic pain such as the periaqueductal gray, limbic, and somatosensory regions [26,30,63,66]. Moreover, an fMRI study of adolescents aged 13 to 19 with varying levels of menstrual pain found altered connectivity within and between the default mode, salience, and central executive networks [51], patterns that mirror those seen in chronic pain [22]. EEG-derived resting-state functional connectivity (FC) has not been explored in dysmenorrhea, although a recent EEG mega-analysis of multiple chronic pain conditions found that brain connectivity was a promising pain biomarker [4]. Although menstrual pain typically begins around menarche, it remains unclear whether the observed neural alterations *precede* pain onset or *result* from repeated painful experience [64].

Here we used resting-state EEG to track neurodevelopmental changes in cortical oscillatory activity over menarche, allowing us to characterize how early menstrual pain experience affects the maturing EEG characteristics of the adolescent brain for the first time. By measuring activity both before and after the typical onset of menstrual pain, we sought to clarify whether neural oscillatory differences precede menstrual pain, demonstrating a pre-existing neural vulnerability. We hypothesized that if neural traits predispose individuals to dysmenorrhea, differences in PAF, oscillatory power, or functional connectivity would be detectable prior to menarche. Alternatively, if the brain alterations seen in adults with dysmenorrhea are a consequence of cumulative menstrual pain experience, EEG signatures would be similar between pre-menarchal young adolescents who develop higher vs. lower menstrual pain.

## 2. Methods

### 2.1 Study overview

Before participation, adolescents signed assent forms and their parents signed consent forms. Participants completed a pre-menarche baseline study visit (between age 9-14), followed by at least one and sometimes two post-menarche follow-up visits (3-14 and 15-30 months after menarche). At each study visit, they completed questionnaires and EEG recordings as well as other sensory testing published elsewhere [49,64]. This study (Early Menstrual Pain Impact on Multisensory Hypersensitivity [EMPATHY]) was approved by Endeavor Health’s Institutional Review Board.

### 2.2 Participant recruitment

Pediatricians’ letters and public advertisements placed throughout northern Chicagoland were used to recruit participants from the community. Inclusion criteria were individuals who were a) assigned female sex at birth b) aged 9 to 14 and c) had not yet attained menarche. Our exclusion criteria were: a) significant medical issues or language comprehension/developmental issues that would limit assessments, b) risk factors for prolonged amenorrhea (e.g., Turner’s syndrome, severe eating disorder), c) history of chronic pain >3 months in duration, d) use of hormonal suppression, e) regular use of prescription analgesics, f) significant allergy to ultrasound or electrode gel g) a body mass index over 40, and h) mental health or medical issues that might limit study participation. The adolescent’s primary physician was contacted for their opinion in the event of doubt around exclusion criteria. Parents were asked “Does your child suffer from any chronic pain conditions or serious medical or mental health diagnoses?” to exclude participants with severe long-term pain that would interfere with our primary research question on the onset of pain during the menarchal transition. Of the 300 participants initially enrolled, 250, 161, and 140 participants met EEG data-quality criteria at the three study visits, respectively. See Fig. 1 for a detailed description of the enrollment and retention trajectories. Table 1 shows a description of the analyzed cohorts’ demographic characteristics.

**Figure 1.**
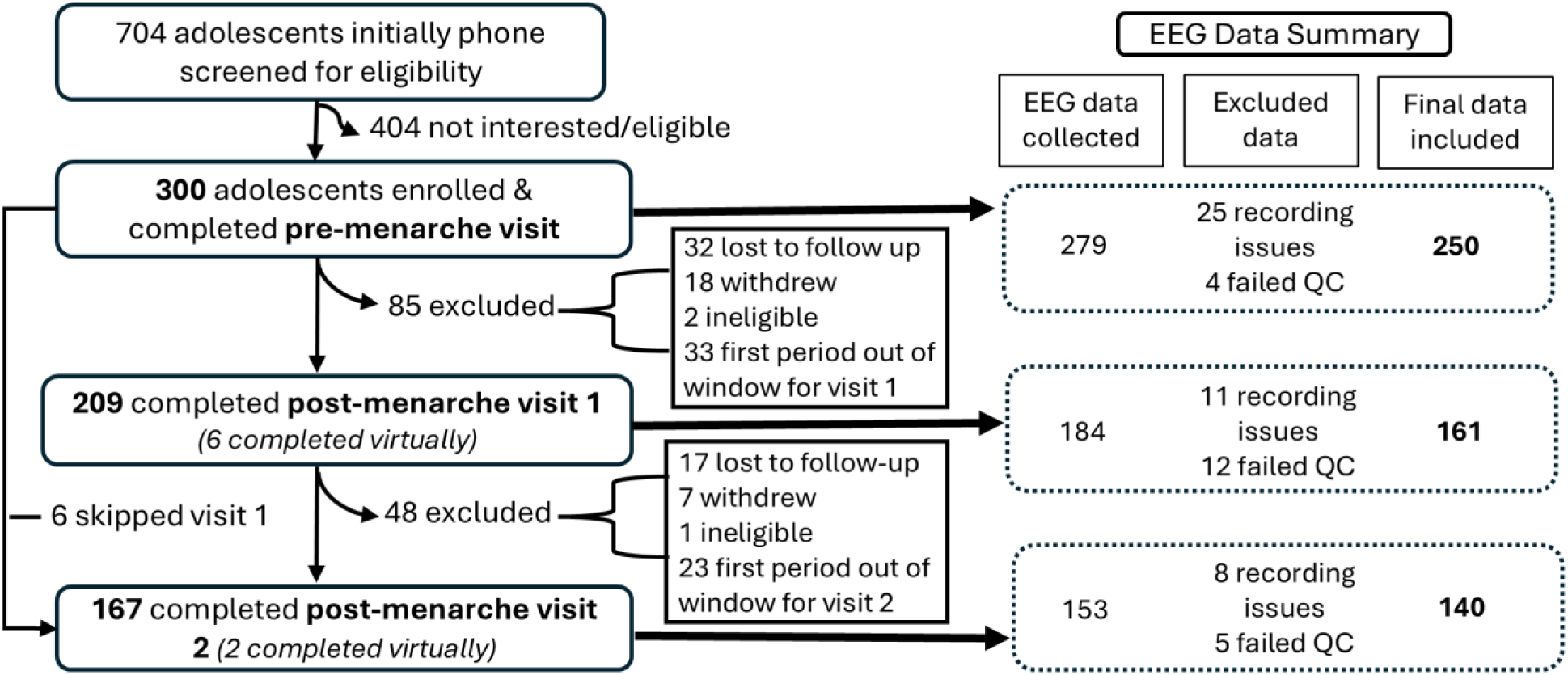
EMPATHY enrollment diagram. Flow-chart depicting screening, enrollment and participation in the Early Menstrual Pain Impact on Multisensory Hypersensitivity (EMPATHY) study. Details on the collection and cleaning of EEG data are also included. The average timespan between the pre-menarche and first post-menarche visit was 19 (*SD*: 8) months, with an average of 14 (*SD*: 4) months between the first and second post-menarche visits. At the pre-menarche, post-menarche 1, and post-menarche 2 visits, EEG data could sometimes not be acquired due to equipment malfunctions, hairstyles that made capping impractical, virtual visit completion or participant refusal, resulting in 279, 184 and 153 EEG datasets respectively. During initial preprocessing, EEG recordings had to be discarded because they were missing event markers, had incomplete recording files, or were recorded with a different reference electrode or different number of channels (i.e., 33 instead of 64), resulting in 254 pre-menarche, 173 post-menarche visit 1 and 145 post-menarche visit 2 EEG datasets suitable for entering into the DISCOVER-EEG pipeline. Participants were retained for group analyses only if they met predefined data-quality thresholds, leaving 250, 161 and 140 participants at the three visits, respectively.

**Table 1.** Demographic characteristics and pubertal status of participants.

| **Characteristic** | **Pre-menarche visit** | **Post-menarche visit 1** | **Post-menarche visit 2** |
| --- | --- | --- | --- |
| **Participants, n** | 250 | 161 | 140 |
| **Age (y), median (Q1, Q3)** | 11 (11, 12) | 13 (12, 14) | 14 (13, 15) |
| **Height (cm), median (Q1, Q3)** | 152 (146, 157) | 162 (158, 167) | 165 (160, 170) |
| **Weight (kg), median (Q1, Q3)** | 42 (36, 47) | 53 (47, 59) | 56 (50, 63) |
| **Tanner Puberty Stage - Breast, n (%)** |  |  |  |
| **I** | 24 (9.6%) | 0 (0%) | 0 (0%) |
| **II** | 111 (44.6 %) | 4 (2.5%) | 1 (0.7%) |
| **III** | 91 (36.5%) | 74 (46.8%) | 25 (18.7%) |
| **IV** | 19 (7.6%) | 60 (38%) | 59 (44%) |
| **V** | 4 (1.6%) | 20 (12.7%) | 49 (36.6%) |
| **Tanner Puberty Stage - Pubic Hair, n (%)** |  |  |  |
| **I** | 37 (14.9%) | 2 (1.3%) | 1 (0.7%) |
| **II** | 114 (45.8%) | 5 (3.2%) | 1 (0.7%) |
| **III** | 69 (27.7%) | 44 (27.8%) | 21 (15.7%) |
| **IV** | 27 (10.8%) | 72 (45.6%) | 57 (42.5%) |
| **V** | 2 (0.8%) | 35 (22.2%) | 54 (40.3%) |
| **Ethnicity, n (%)** |  |  |  |
| **Hispanic or Latino** | 18 (7.2%) |  |  |
| **Non-Hispanic or Latino** | 232 (92.8%) |  |  |
| **Race, n (%)** |  |  |  |
| **Asian** | 37 (14.8%) |  |  |
| **American Indian/Alaskan Native** | 2 (0.8%) |  |  |
| **Black or African American** | 10 (4%) |  |  |
| **White or Caucasian** | 225 (90%) |  |  |
| **Household Income, n (%)** |  |  |  |
| **Household income <\$25,000** | 2 (0.8%) | | |
| **Household income \$25,000 - <\$50,000** | 4 (1.6%) | | |
| **Household income \$50,000 - <\$75,000** | 9 (3.6%) | | |
| **Household income \$75,000 - <\$100,000** | 18 (7.2%) | | |
| **Household income \$100,000 - <\$150,000** | 44 (17.6%) | | |
| **Household income \$150,000 or greater** | 154 (61.6%) | | |
| **Household income Prefer not to answer** | 19 (7.6%) |  |  |

Our race questionnaire allowed participants to select multiple options. Tanner puberty stage scores missing from 1 pre-menarche visit participant, 3 post-menarche visit 1 participants, and 6 post-menarche visit 2 participants. Missing data excluded.

### 2.3 Questionnaires

Participants completed demographic, medical history, and psychosocial questionnaires through REDCap [20]. Adolescents completed the Short-Form Patient Reported Outcomes Measurement Information System (PROMIS) Pediatric Profile for depression (PROMIS Ped SF v2.0 - Depressive Symptoms 8b), anxiety (PROMIS Ped SF v2.0 -Anxiety 8b), and the Pediatric Psychological Stress Experiences v1.0 – Short Form 8a for stress as well as the PROMIS Pain Intensity item (9033R1r), which asked about pain intensity in the past week [2,21]. Adolescents completed a modified version of the Tanner Puberty Stage questionnaire [33] including questions on breast and pubic hair development (Supplementary Fig. 1), while parents completed the PROMIS Parent Proxy Short Form v1.0 – Sleep Disturbance 8a to assess their adolescent’s sleep. After menarche, a visual analog scale (VAS) was used to measure average menstrual pain severity, with 0 indicating no pain and 100 the worst pain imaginable. Specifically, adolescents were asked: “Think back over the last 3 months. Please use this slider to indicate the average amount of cramping pain you felt during your menstrual periods (around or below your belly button or lower back)” [64].

### 2.4 Resting state EEG

#### 2.4.1 Task acquisition

Participants were seated in a chair, where a 64-channel equidistant electrode cap (ActiCAP, CACS-64-X7, BrainVision) was fitted to their heads. Facial and eye-movement recording electrodes were placed in the middle of the brow, below the right eye and above the left eye. EEG data was recorded with a 500 Hz sampling rate, with impedances verified by research staff to be below 25 kΩ. Participants were coached to avoid excessive movement during the resting state task. To remove artifacts, we filtered at 1Hz (high-pass) and the Nyquist frequency (250 Hz, low-pass) using a 20 dB/decade Butterworth filter with a BrainVision actiCHamp 24-bit A/D amplifier and PyCorder software (BrainVision). For the recording, the room lights were turned off and the participant was instructed to look ahead at a black computer screen displaying a white fixation cross. The resting state task consisted of alternating 60 second blocks of eyes open and eyes closed rest, for a total of 8 minutes. Research staff sat with participants throughout the recording to ensure they were following task instructions.

#### 2.4.2 EEG preprocessing

At the pre-menarche, post-menarche 1, and post-menarche 2 visits, EEG data could sometimes not be acquired (e.g., equipment malfunctions, hairstyles that made capping impractical, virtual visit completion, or participant refusal) or analyzed due to not meeting data quality criteria, resulting in 250, 161, and 140 EEG datasets, respectively (see Fig. 1). Full preprocessing parameters and data quality criteria are provided in the Supplementary Methods. In brief, data were downsampled to 250 Hz; line noise was removed; and high-pass filtering, automated rejection of artefactual channels, common average referencing, ICA-based removal of ocular and muscular components, spherical-spline interpolation of removed channels, and artefact-subspace-reconstruction removal of bad time segments were applied before segmentation into 2-s epochs with 50% overlap.

#### 2.4.3 EEG feature Extraction

For each participant, the DISCOVER-EEG pipeline [15] derived global power spectra, PAF, and source-space functional connectivity; complete details are given in the Supplementary Methods and in Gil Ávila et al. (2023). Global power spectra (1–100 Hz) were computed by multitaper estimation and averaged across channels and epochs, using band definitions theta (4-7.9 Hz), alpha (8-12.9 Hz), beta (13-30 Hz) and gamma (30.1-80 Hz). PAF was estimated using a centre-of-gravity method. Because alpha power was overwhelmingly concentrated over the occipital cortex, where peak frequency estimation is most reliable, we followed up the global PAF analysis with a regional analysis restricted to occipital channels (Supplementary Methods). Functional connectivity was estimated in source space with an LCMV beamformer and the 100-region, 7-network Schaefer atlas [57], quantified for each band as the orthogonalized amplitude envelope correlation. MNI coordinates for the 100 regions of interest (ROIs) used and their corresponding network and anatomical labels are listed in Supplementary Table 1.

### 2.5 Statistical analysis

#### 2.5.1 Questionnaire data

Questionnaire data were analyzed using R (4.4.0). Because there is no official threshold of low vs. high menstrual pain in adolescents, we used a data-driven process to establish an appropriate cut-off. We performed k-medians clustering using several methods (gap, silhouette and elbow) on the VAS menstrual pain data (three-month recall) from post-menarche visit 1 and 2 to identify the optimal cluster solution. A cut-off point of 30/100 on the VAS was identified. This cut-off to distinguish between higher and lower menstrual pain groups is consistent with an earlier analysis of this data set [64] and with other studies implicating the boundary between mild and moderate pain [45]. Wilcoxon rank-sum tests with Holm-Bonferroni correction applied across the three study visits were used to compare weekly pain intensity scores between participants in the higher and lower menstrual pain groups. Group differences in PROMIS t-scores at each visit were assessed via the default one-way (Welch) ANOVA within tableone’s CreateTableOne.

#### 2.5.2 EEG features: group comparisons

For each of the EEG features examined (peak alpha frequency, power spectrum, and functional connectivity) we performed group analyses as well as several sensitivity analyses using custom R(4.4.0) and MATLAB (R2023b) scripts based in part on Gil Ávila et al.’s original DISCOVER-EEG analyses [15]. The MATLAB scripts employed functions from FieldTrip (2024-01-10), EEGLAB (2023.1) and the Brain connectivity toolbox (2019-03-03) and are available at https://doi.org/10.17605/OSF.IO/URKD7. For each EEG feature, eyes-open and eyes-closed resting state data were analyzed separately. Linear mixed-effects models (LMMs) in R were used to analyze PAF data, which consisted of one eyes-open and one eyes-closed value per participant. Alternative statistical methods (described below) were used to analyze EEG power spectral data (1–100 Hz) and functional connectivity data (100 × 100 ROI pairs) as they were too high-dimensional relative to our sample size and would be statistically underpowered to be entered into LMMs.

##### 2.5.2.1 Peak alpha frequency

Global and occipital PAF (one value per condition per participant) were analyzed with linear mixed-effects models (lme4 with lmerTest; Satterthwaite degrees of freedom; BOBYQA optimiser), fitted separately for eyes-closed and eyes-open conditions, retaining all available observations under a missing-at-random assumption. The developmental trajectory of global PAF was characterized with a full-cohort model treating visit as a linear predictor (coded 0/1/2) with by-subject random intercepts and slopes [PAF ∼ visit_num + (1 + visit_num | subject_ID)]; a likelihood-ratio test confirmed the random slope improved fit and it was retained throughout, and the average per-visit slope was obtained with the marginaleffects package. To test whether PAF differed by menstrual pain, we fitted a Group × visit_num interaction model [PAF ∼ visit_num × Group + (1 + visit_num | subject_ID)] for both global and occipital PAF. Occipital cortex was selected a priori because the resting alpha rhythm arises predominantly from this region and, consistent with this, alpha power in our cohort was maximal over occipital channels at every visit, the location where peak alpha frequency is most reliably estimated. Group differences at each visit were estimated as contrasts of estimated marginal means (emmeans) at visit_num = 0, 1 and 2, Holm-corrected across the three visits within each condition. Full model specifications, per-visit and per-group sample sizes, and the factor-coded model underlying the consecutive-visit contrasts are given in the Supplementary Methods (Supplementary Table 2).

##### 2.5.2.2 EEG power analyses

Cross-sectional group comparisons (e.g., higher vs. lower menstrual pain) of global power spectra from 1 to 100 Hz were performed for each study visit using two-tailed independent-samples cluster-based permutation tests, with Monte Carlo simulations (500 randomizations). The sum of the t-values within each cluster were used to calculate cluster-level statistics, with a cluster threshold and alpha level of 0.05. To measure how EEG power changed across the menarchal transition, dependent-samples cluster-based permutation tests were used for longitudinal within-subject comparisons between the pre-menarche visit and post-menarche visit 1, and between post-menarche visits 1 and 2. These longitudinal EEG power analyses were performed first with all participants grouped together (i.e., full cohort) and then separately for the higher and lower menstrual pain groups.

##### 2.5.2.3 Functional Connectivity

To compare functional connectivity between higher and lower menstrual pain groups at a single time point we compared the connectivity values between each pair (undirected) of the 100 Schaefer atlas ROIs for the theta, alpha, beta and gamma frequency bands. This was done using 4,950 two-sided independent samples t-tests for each connectivity matrix. These cross-sectional comparisons of connectivity were performed for each study visit. For within-subject comparisons across study visits, two-sided dependent samples t-tests were used. Like the EEG power analyses, longitudinal comparisons between the pre-menarche visit and post-menarche visit 1 and between post-menarche visits 1 and 2 were first performed in the full cohort of participants and then stratified by menstrual pain group. Multiple comparison testing was addressed by controlling the false discovery rate (FDR) to *q*<0.05. An MNI to Talairach converter [27] was used to include additional anatomical references in results tables listing significant Schaefer 7-network atlas ROI-pairs [57].

##### 2.5.2.4 Sensitivity analyses

The following cross-sectional analyses were performed for each EEG metric at each study visit to serve as controls for our group comparisons of higher and lower menstrual pain scores: 1) within-subject eyes-open vs. eyes-closed resting state EEG, 2) between-subject high vs. low weekly pain intensity and 3) between-subject high vs. low depression. Statistical tests for each EEG metric were identical to those performed in the high vs. low menstrual pain group analysis. Methodological details and results for these sensitivity analyses are discussed in the Supplementary Material and Supplementary Figs. 2 - 4.

## 3. Results

### 3.1 Menstrual pain is common in early adolescence and linked to weekly pain, but not mood

At each post-menarche visit, over half of participants reported menstrual pain ≥30 over the preceding 3 months (Fig. 2). Participants in the higher menstrual pain (≥30 on 0-100 visual analog scale) group also reported higher general weekly pain intensity (p=0.001 at post-menarche visit 1 and p=0.004 at post-menarche visit 2 – Fig. 2). At the post-menarche visits, menstrual pain severity was not associated with anxiety, depression, stress, or sleep disturbance scores (Table 2). Age at menarche and self-assessed pubertal staging (breast and pubic hair development) did not differ between menstrual pain groups at any visit (Table 1, Supplementary Materials).

**Figure 2.**
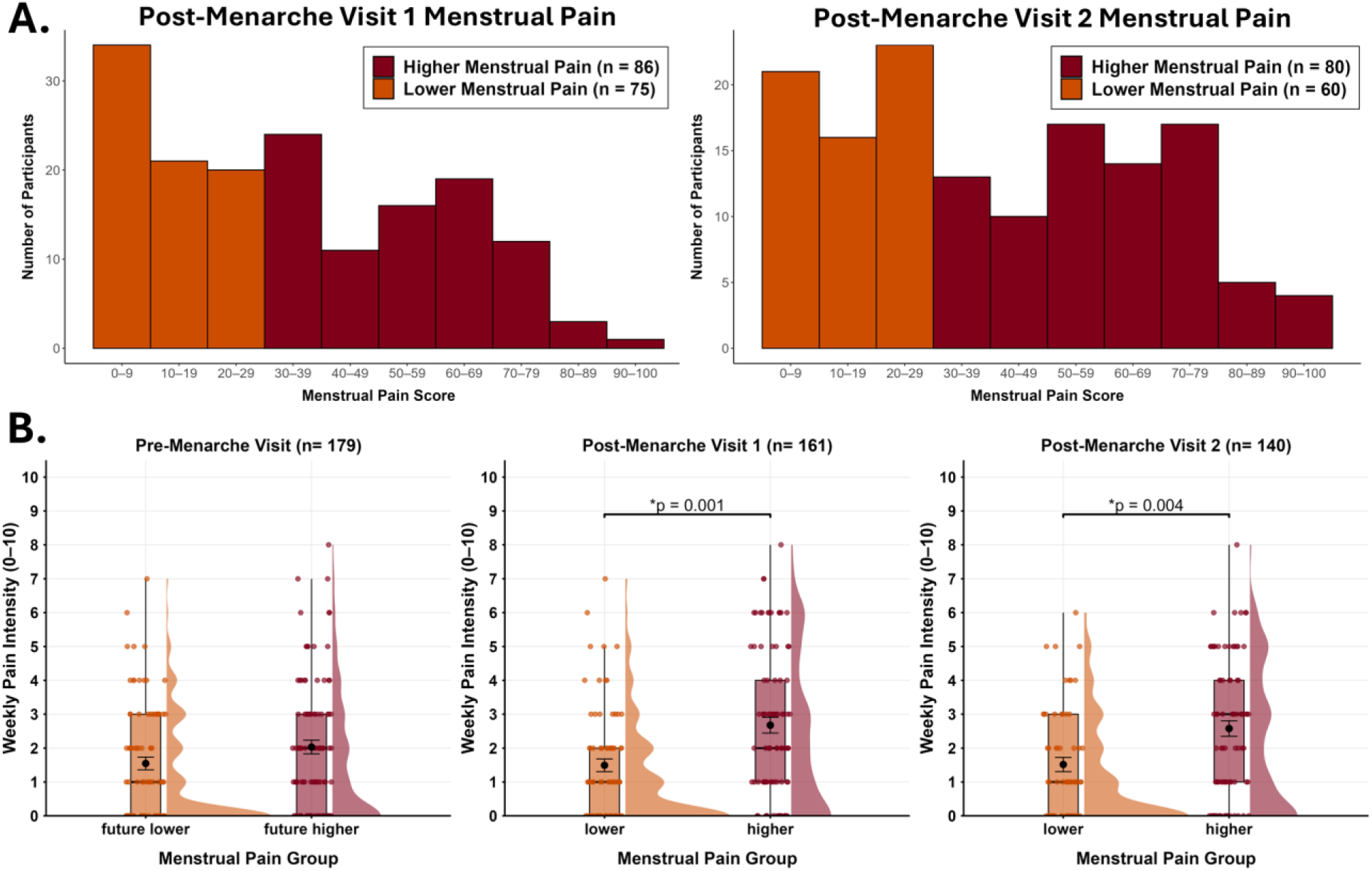
Menstrual pain ratings at the first and second post-menarche visit reveal menstrual pain is common and linked with greater weekly pain intensity. **A.** At each post-menarche visit, adolescents were asked “think back over the last 3 months. Please use this slider to indicate the average amount of cramping pain you felt during your menstrual periods (around or below your belly button or lower back)”, and then rated their pain using a visual analogue scale. At post-menarche visit 1, over half (53%) of adolescents reported menstrual pain ≥30/100. At the second post-menarche visit, 57% reported menstrual pain ≥30/100. A k-medians clustering analysis was performed on the menstrual pain scores (see supplementary material) to determine the cut-off point of 30/100 to define our lower and higher menstrual pain groups. B. Participants’ ratings on the PROMIS Pain Intensity item - which asked about pain intensity (0 -10) in the past week – organized by menstrual pain group. At the pre-menarche visit, adolescents were grouped according to their future menstrual pain status (i.e., menstrual pain ratings at the first post-menarche visit). Weekly pain intensity in this case referred to any pain the individual may be experiencing (e.g., headaches, muscle aches, stomach pain etc.,) and was not specific to menstruation. No differences in weekly pain intensity scores were seen between future higher and future lower menstrual pain groups before menarche. After menarche, adolescents with higher menstrual pain also reported higher weekly pain. * indicates Holm-corrected *p* < 0.05.

**Table 2.**
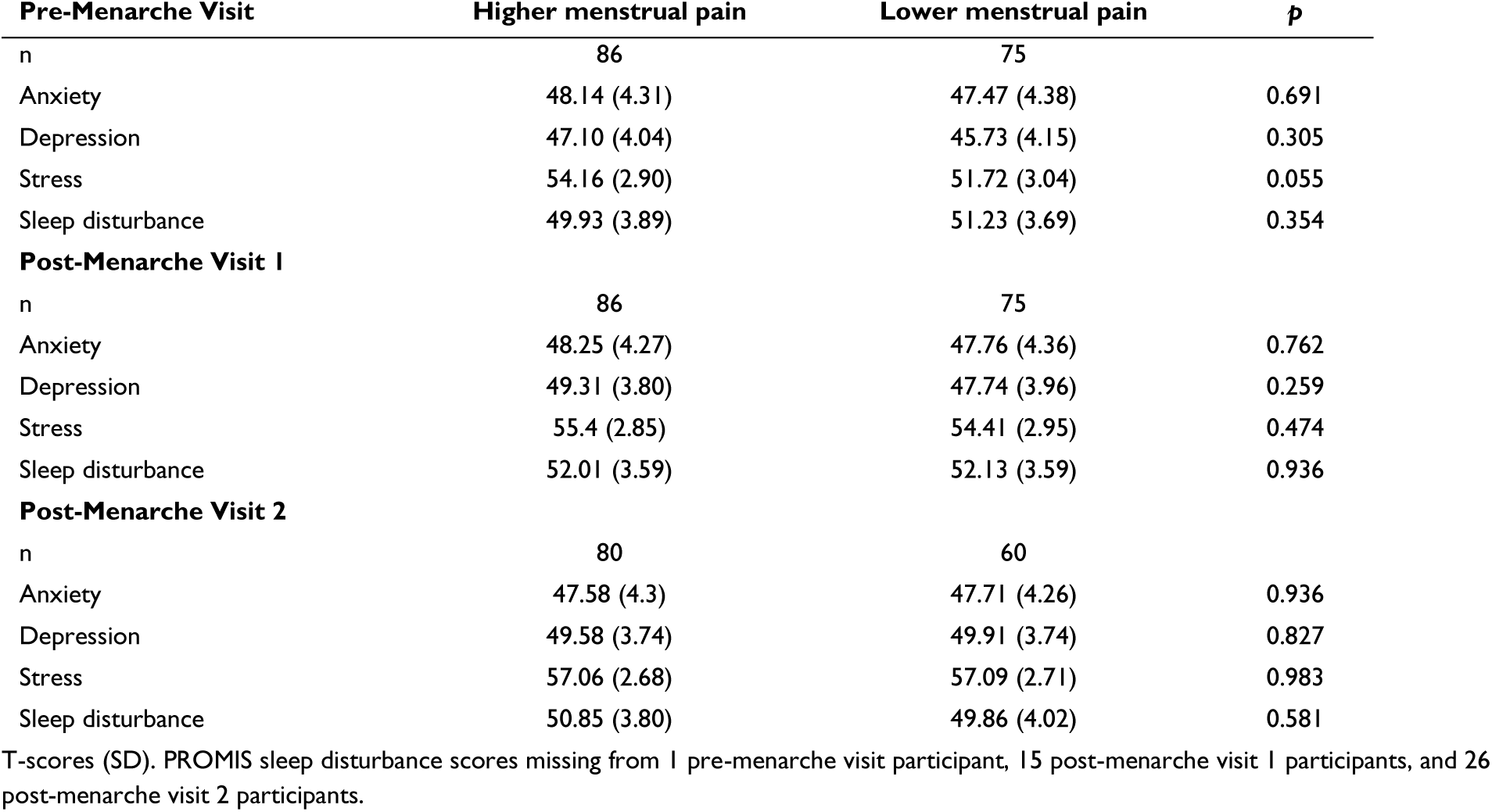
PROMIS psychological and sleep scores.

### 3.2 PAF increased across the menarchal transition

Global PAF increased steadily across the menarchal transition in the full cohort (Fig. 3A), gaining +0.056 Hz per visit in eyes-closed recording (95% CI [+0.037, +0.074], *p* < 0.001) and +0.028 Hz per visit in eyes-open recording (95% CI [+0.012, +0.045], *p* < 0.001). This developmental increase did not differ between menstrual pain groups. For global PAF, the visit × group interaction (Supplementary Table 2) was non-significant in both eyes-closed (β = −0.030 Hz/visit, 95% CI [−0.069, +0.009], *p* = 0.127) and eyes-open conditions (β = −0.024 Hz/visit, 95% CI [−0.059, +0.011], *p* = 0.185). We next restricted the PAF analysis to occipital channels, where the resting alpha rhythm predominates and its peak frequency is most reliably estimated (Fig. 3B), and the same was true for occipital PAF (eyes-closed β = −0.031, 95% CI [−0.077, +0.015], *p* = 0.182; eyes-open β = −0.029, 95% CI [−0.069, +0.011], *p* = 0.153), that is, occipital PAF increased from the pre-menarchal visit to post-menarche visit 2 at a comparable rate in both groups (Fig. 3C, Supplementary Table 2).

**Figure 3.**
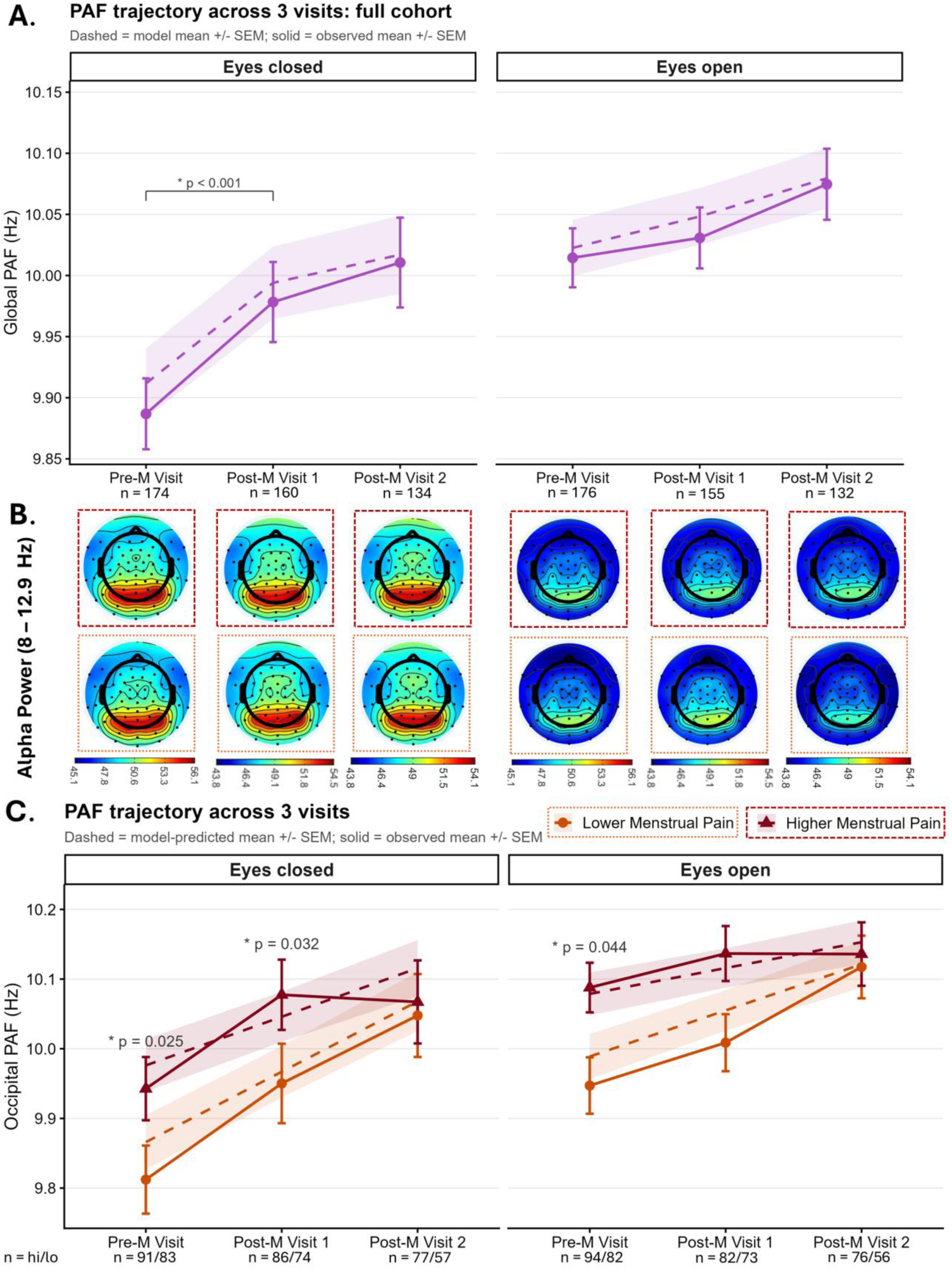
Peak alpha frequency changes across the menarchal transition and is linked with menstrual pain. **A.** Global eyes-closed and eyes-open PAF analyses revealed significant increases in PAF across the menarchal transition, with model-predicted pre-menarche visit eyes-closed/eyes-open PAF of 9.92/10.02 Hz increasing to 10.03/10.08 Hz by the second post-menarche visit. Visit-level contrasts revealed a significant increase in eyes-closed PAF from the pre-menarche to post-menarche visit 1 (Δ = +0.082 Hz, *p* <0.001). **B**. Topoplots of eyes-closed and eyes-open alpha (8 – 12.9 Hz) activity across all participants for each study visit were created in EEGLab, and the occipital cortex was identified as a region of interest for alpha power. **C.** Eyes-open and eyes-closed analyses comparing cross-sectional and longitudinal differences in occipital PAF between higher and lower menstrual pain groups similarly revealed an increase in PAF across the menarchal transition in both groups. Visit-specific contrasts revealed significantly faster pre-menarche occipital PAF in adolescents who later developed higher menstrual pain compared to those who later developed lower menstrual pain in eyes-closed [Δ = +0.110 Hz, *p* = 0.025] and eyes-open [Δ = +0.090 Hz, *p* = 0.044] conditions. Occipital eyes-closed PAF was also significantly faster in adolescents with higher vs. lower menstrual pain at post-menarche visit 1 [Δ = + 0.079 Hz, 95% CI [+0.000, +0.158], *p* = 0.032]. Topoplot colour bar units = μV^2^/Hz. PAF = peak alpha frequency. SEM = standard error of the mean. Hz = hertz. Pre-M = pre-menarche visit, Post-M1 = post-menarche visit 1, Post-M2 = post-menarche visit 2.

### 3.3 Faster peak alpha frequency is linked with higher menstrual pain

Although the two menstrual pain groups followed similar PAF trajectories (above), occipital PAF distinguished the groups, most clearly before menarche. Pre-menarche, global eyes-closed PAF was already faster in adolescents who would later develop higher menstrual pain, though this did not reach significance [Δ = +0.081 Hz, 95% CI [−0.004, +0.167], *p* = 0.066; Supplementary Table 2]. Pre-menarche occipital PAF was significantly faster in the future higher-than the future lower-pain group in both eyes-closed [Δ = +0.110 Hz, 95% CI [+0.011, +0.210], *p* = 0.025] and eyes-open [Δ = +0.090 Hz, 95% CI [+0.002, +0.178], *p* = 0.044] conditions. The eyes-closed difference remained significant at the first post-menarche visit [Δ = +0.079 Hz, 95% CI [+0.0004, +0.158], *p* = 0.032], with a non-significant difference in the same direction for eyes-open [Δ = +0.061 Hz, 95% CI [−0.008, +0.130], *p* = 0.069]. By the second post-menarche visit, the groups no longer differed in either condition (eyes-closed *p* = 0.224, eyes-open *p* = 0.348; Fig. 3C). These contrasts identify where the group difference was detectable (pre-menarche and post-menarche visit 1), but because the group x visit interaction was not significant, they should not be read as a reliable change in the effect over time.

As a positive control, global PAF showed the expected eyes-open versus eyes-closed difference (Supplementary Results, Supplementary Fig. 2), confirming the recordings were sensitive to a well-established modulation of alpha frequency. The occipital PAF effects appeared specific to menstrual pain. Neither weekly general pain intensity (Supplementary Fig. 3) nor depression score (Supplementary Fig. 4) was associated with global or occipital PAF, distinguishing the menstrual pain-PAF relationship from broader pain or mood influences.

### 3.4 Global resting power decreased after menarche

We observed no differences in the broadband power spectrum between adolescents with higher versus lower menstrual pain at each post-menarchal visit, nor between adolescents with future higher or lower menstrual pain at the pre-menarchal visit (Supplementary Fig. 5A-C). Because the normative neurodevelopmental changes occurring across early adolescence are substantially larger than any expected group difference and may overshadow it cross-sectionally, we performed separate longitudinal within-subject analyses of global power for the lower- and higher-pain groups.

In the full cohort, resting power showed pronounced developmental shifts across the menarchal transition (Fig. 4A, D). From the pre-menarche to the first post-menarche visit, power decreased across both eyes-closed [1–28.1 Hz (*p* = 0.002)] and eyes-open [1–56.9 Hz (*p* = 0.002)] conditions, reflecting broad reductions spanning theta, alpha, beta, and gamma. Further decreases in theta, alpha, and beta power occurred from the first to the second post-menarche visit in both eyes-closed (1–11.4 Hz, *p* = 0.02; 12.3–26.7 Hz, *p* = 0.02) and eyes-open conditions (1–21.8 Hz, *p* = 0.008).

**Figure 4.**
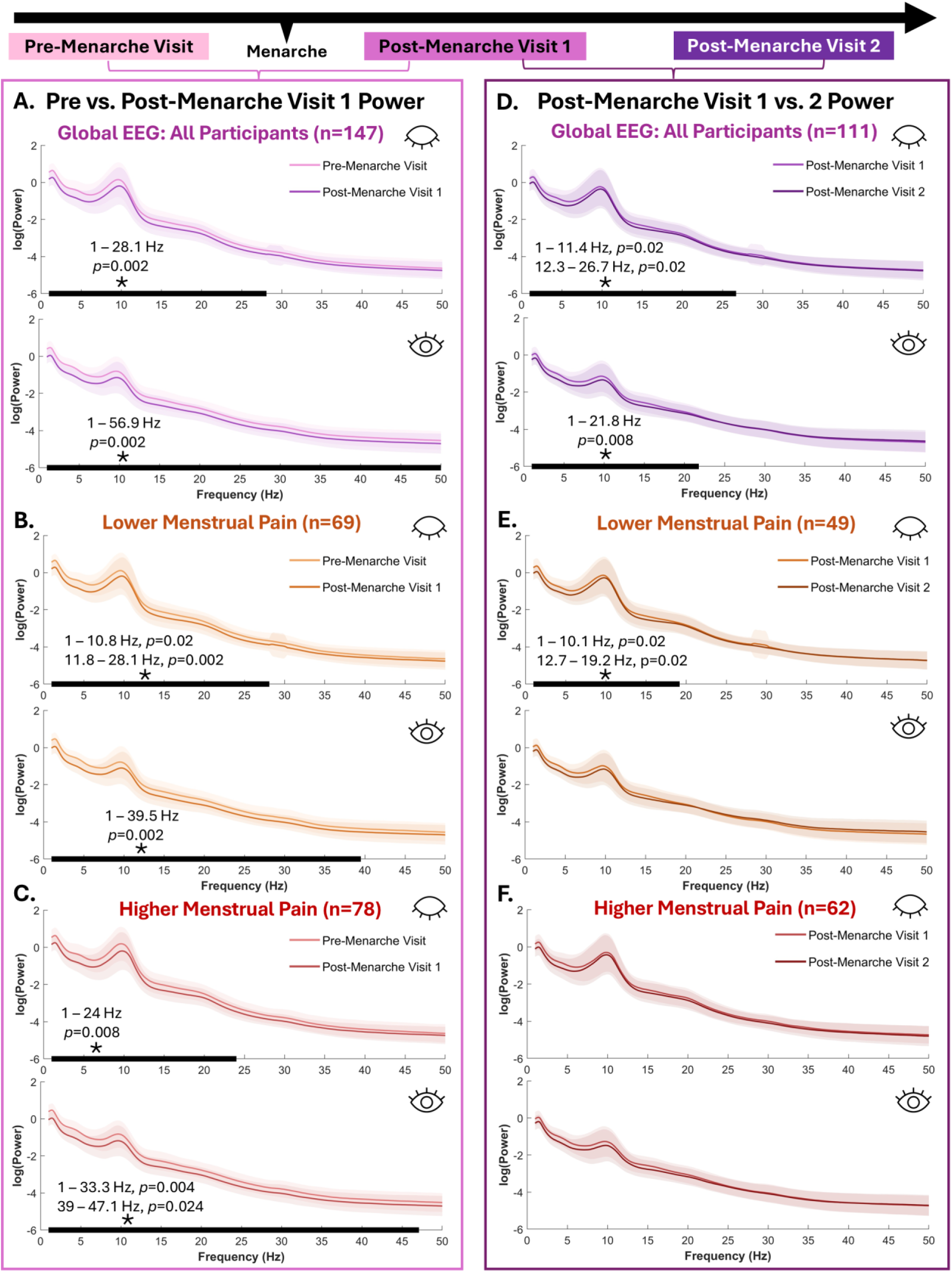
Global power decreases across the menarchal transition. **A.** Longitudinal within-subject (n=147) analysis of the global power spectrum from the pre-menarche to first post-menarche visit revealed significant reductions in eyes-closed theta, alpha, and beta power and eyes-open theta, alpha, beta and gamma power across menarche. When stratified by menstrual pain status at the first post-menarche visit, alpha, beta, theta and gamma power decreases over menarche were seen in adolescents with lower menstrual pain (**B**; n=69) and higher menstrual pain (**C**; n=78). **D.** Longitudinal within-subject analysis of all participants (n=111) from the first to second post-menarche visit revealed decreases in eyes-closed and eyes-open theta, alpha and beta power. When stratified by menstrual pain status (**E, F**), significant decreases in theta, alpha, and beta power were seen among adolescents reporting lower menstrual pain (n=49). Hz = hertz.

Analyzed separately (Fig. 4B–C, E–F), both menstrual pain groups showed significant pre- to post-menarche visit 1 decreases in eyes-open and eyes-closed theta, alpha, beta, and gamma power (*p’s* ≤ 0.024). Between the first and second post-menarche visits, the lower-pain group showed eyes-closed decreases in theta, alpha, and beta power (1–10.1 Hz, *p* = 0.02; 12.7–19.2 Hz, *p* = 0.02), whereas no significant changes were detected in the higher-pain group over this interval. These stratified group analyses suggest that power reductions in the higher-pain group were significant only in the period around menarche, while reductions in the lower-pain group extended into the later post-menarche period.

### 3.5 Theta connectivity decreases broadly across the menarchal transition

Cross-sectional comparisons of functional connectivity between higher and lower menstrual pain groups yielded no FDR-corrected differences at any visit (Supplementary Fig. 6 A-C, Supplementary Tables 7–9). Longitudinally, by contrast, connectivity changed substantially across the menarchal transition, and these changes were predominantly decreases in theta-band connectivity. Because connectivity was tested across all 4,950 ROI pairs within each band (FDR-corrected), we summarize these effects by the number of ROI-pair connections showing a significant change.

From pre-menarche to post-menarche visit 1 (Fig. 5B, Table 3, Supplementary Table 3), the most extensive change was a decrease in eyes-open theta connectivity, spanning 77 ROI-pair connections distributed across networks but concentrated on the visual and dorsal attention networks, particularly their connections with the limbic, default mode, and control networks. Eyes-closed effects over this interval were smaller and mixed in direction (3 ROI-pairs with decreased gamma connectivity, 4 with increased beta connectivity; Fig. 5B, Table 3, Supplementary Table 3). When stratified by menstrual pain group, the lower-pain group showed 20 ROI-pairs with decreased eyes-closed theta connectivity (Fig. 5B, Supplementary Table 4), most frequently involving the visual, dorsal attention, default mode, and control networks; no connections survived correction in the higher-pain group at this transition.

**Figure 5.**
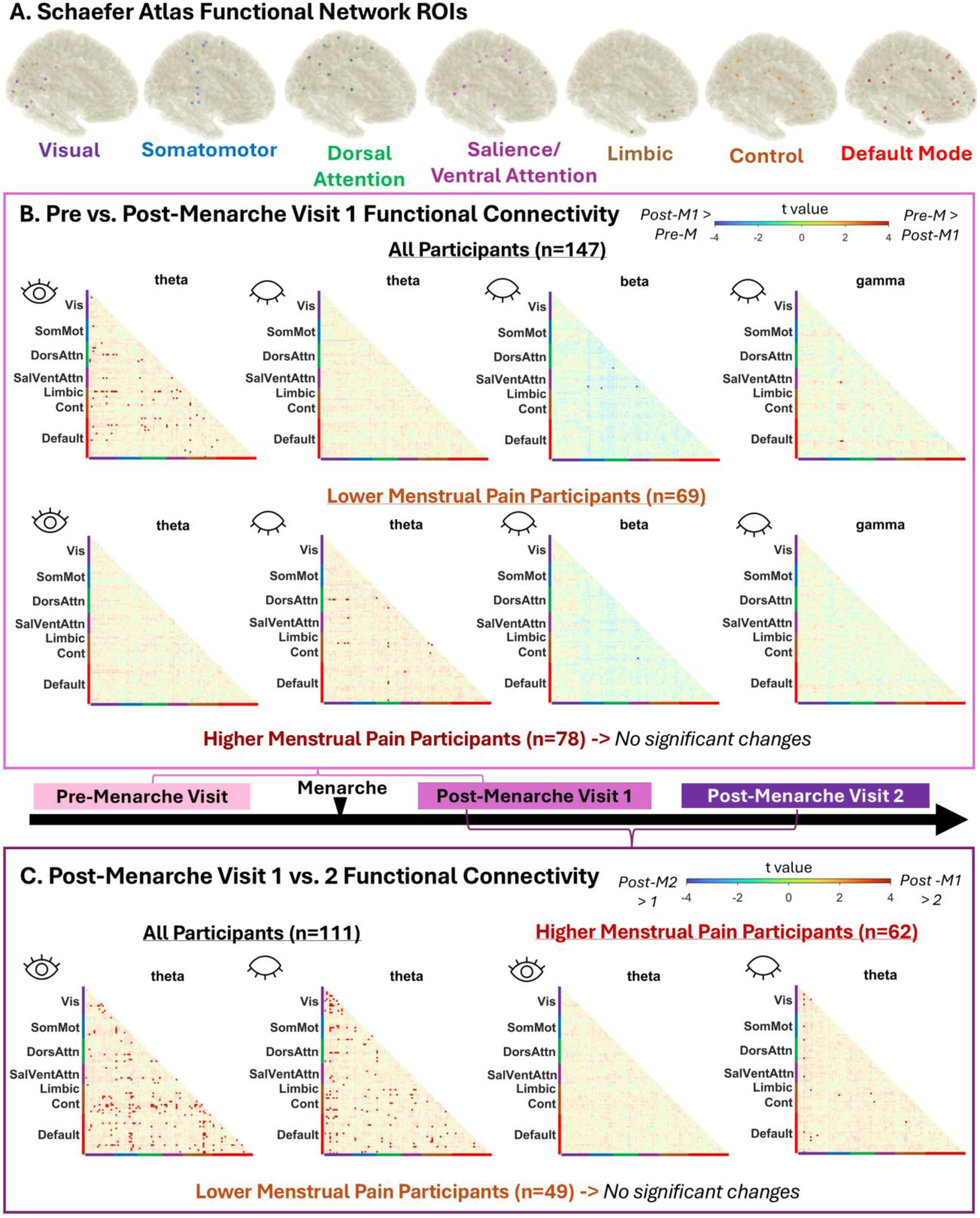
Theta functional connectivity decreases across the menarchal transition. **A.** The centroids of 100 ROIs from the 7-network version of the Schaefer atlas were used for the functional connectivity analyses, which included the which included Visual (Vis), Somato-Motor (SomMot), Dorsal attention (DorsAttn), Salience-Ventral attention (SalVentAttn), Limbic, Control (Cont), and Default Mode (Default) networks. The MNI coordinates as well as Schaefer and Talairach labels for these ROIs can be found in Table 3 and the supplementary materials. **B.** Longitudinal within-subject functional connectivity analyses from the pre to the first post-menarche visit revealed that eyes-open theta connectivity decreased across multiple brain networks in all participants (n=147), with some limited decreases in eyes-closed gamma connectivity and increases in eyes-closed beta connectivity occurring during this time period as well. When analyzed separately by menstrual pain group, adolescents with lower menstrual pain (n=69) showed decreased eyes-closed theta connectivity and one ROI pair with increased eyes-closed beta connectivity. **C.** Post-menarche visit 1 to 2 longitudinal within-subject functional connectivity analyses showed decreases in theta connectivity in all participants (n=111), this time in both eyes-open and eyes-closed conditions. When analyzed separately by menstrual pain group, adolescents with higher menstrual pain (n=62) had decreased eyes-closed theta connectivity. Correlation matrices are thresholded to *q* ≤ 0.05, false discovery rate (FDR) corrected to account for multiple comparisons. Pre-M = pre-menarche visit, Post-M1 = post-menarche visit 1, Post-M2 = post-menarche visit 2.

**Table 3.** Top functional connectivity results from full-cohort longitudinal analyses.

| **All Participants Pre-Menarche > Post-Menarche Visit 1, eyes-open theta** |  |  |  |  |  |
| --- | --- | --- | --- | --- | --- |
| **Strongest ROI-ROI Pairs** |  |  |  |  |  |
| **Schaefer label** | **Talairach label** | **t** | **q** | **Most Frequent Network Pairings** | **Most Frequent ROIs (Talairach)** |
| LH_Limbic_TempPole_2 - RH_Default_Par_1 | Left Inferior Temporal Gyrus - Right Angular Gyrus | 5.66 | 0.001 | DorsAttn -- Vis (x11) | Left Inferior Temporal Gyrus (x21) |
| LH_Vis_1 - RH_DorsAttn_Post_3 | Left Fusiform Cortex - Right Visual Motor Cortex | 5.43 | 0.001 | Limbic -- Vis (x9) | Left PreMotor & Supplementary Motor Area (x16) |
| RH_DorsAttn_Post_4 - LH_Limbic_TempPole_2 | Right Visual Motor Cortex - Left Inferior Temporal Gyrus | 5.01 | 0.005 | Default -- DorsAttn (x8) | Left Visual Association Cortex (x13) |
| LH_Limbic_TempPole_2 - RH_Cont_Par_2 | Left Inferior Temporal Gyrus - Right Angular Gyrus | 4.92 | 0.006 | Default -- Vis (x8) | Left Pars Orbitalis (x10) |
| LH_Vis_1 - RH_DorsAttn_Post_4 | Left Fusiform Cortex - Right Visual Motor Cortex | 4.75 | 0.010 | Cont -- Vis (x4) | Left Fusiform Cortex (x8) |
| **All Participants Post-Menarche Visit 1 > Post-Menarche Visit 2, eyes-open theta** |  |  |  |  |  |
| **Strongest ROI-ROI Pairs** |  |  |  |  |  |
| **Schaefer label** | **Talairach label** | **t** | **q** | **Most Frequent Network Pairings** | **Most Frequent ROIs (Talairach)** |
| RH_SalVentAttn_TempOccPar_2 - RH_Cont_PFCI_4 | Right Supramarginal Gyrus - Right Frontal Eye Fields | 5.32 | 0.004 | Cont -- Default (x24) | Right Dorsolateral Prefrontal Cortex (x50) |
| LH_SalVentAttn_FrOperIns_2 - RH_Cont_PFCI_2 | Left Insula - Right Dorsolateral Prefrontal Cortex | 5.22 | 0.004 | Default -- SomMot (x21) | Right Primary Motor Cortex (x24) |
| LH_Cont_pCun_1 - RH_Cont_PFCI_3 | Left Visual Motor cortex - Right Dorsolateral Prefrontal Cortex | 4.80 | 0.013 | Cont -- Vis (x20) | Right Frontal Eye Fields (x22) |
| RH_DorsAttn_PrCv_1 - RH_SalVentAttn_TempOccPar_2 | Right Broca's Area/Operculum - Right Supramarginal Gyrus | 4.75 | 0.013 | Cont -- SomMot (x15) | Right Broca's Area/ Pars Triangularis (x18) |
| LH_Vis_8 - RH_Vis_3 | Left Visual Association Cortex - Right Fusiform Cortex | 4.73 | 0.013 | Default -- Default (x14) | Right PreMotor & Supplementary Motor Area (x18) |
| **All Participants Post-Menarche Visit 1 > Post-Menarche Visit 2, eyes-closed theta** |  |  |  |  |  |
| **Strongest ROI-ROI Pairs** |  |  |  |  |  |
| **Schaefer label** | **Talairach label** | **t** | **q** | **Most Frequent Network Pairings** | **Most Frequent ROIs (Talairach)** |
| LH_Vis_4 - RH_Vis_3 | Left Secondary Visual Cortex - Right Fusiform Cortex | 5.35 | 0.003 | Vis -- Vis (x23) | Left Visual Association Cortex (x36) |
| RH_DorsAttn_Post_1 - RH_Limbic_OFC_1 | Right Visual Association Cortex - Right Orbitofrontal Cortex | 5.24 | 0.003 | Default -- Vis (x20) | Left Secondary Visual Cortex (x30) |
| LH_Vis_8 - LH_Vis_9 | Left Visual Association Cortex - Left Visual Association Cortex | 5.18 | 0.003 | Cont -- Vis (x12) | Right Visual Association Cortex (x17) |
| LH_Vis_6 - RH_Cont_Cing_1 | Left Primary Visual Cortex - Right Ventral Posterior Cingulate Cortex | 4.94 | 0.006 | DorsAttn -- Vis (x12) | Left Fusiform Cortex (x16) |
| LH_Vis_4 - RH_SomMot_2 | Left Secondary Visual Cortex - Right Insula | 4.92 | 0.006 | Cont -- Default (x10) | Right Ventral Posterior Cingulate Cortex (x16) |

Network labels derived from Schaefer 7-network atlas,<sup>31</sup> Talairach anatomical labels derived from MNI-Talairach converter.<sup>36</sup> Complete results available in supplementary table 3. $q \leq 0.05$ (FDR-corrected)

From post-menarche visit 1 to visit 2 (Fig. 5C, Table 3, Supplementary Table 5), theta was the only band to change significantly, decreasing in both conditions across the full cohort (205 eyes-open and 152 eyes-closed ROI-pairs). Eyes-open decreases most frequently involved the default mode, control, somatomotor, and visual networks, while eyes-closed decreases were concentrated within the visual network and between it and the default mode, control, and dorsal attention networks, as well as between the control and default mode networks. When stratified, the higher-pain group showed 18 ROI-pairs with decreased eyes-closed theta connectivity (Fig. 5C, Supplementary Table 6), particularly within the visual and default mode networks; no connections survived correction in the lower-pain group at this transition. Overall, theta connectivity decreased broadly across multiple networks over the menarchal transition. This paralleled the reduction in theta power reported above, although the two were not formally tested for association.

## 4. Discussion

We hypothesized that if neural traits predispose individuals to dysmenorrhea, distinct EEG signatures would be detectable before menarche. Consistent with this hypothesis, a faster pre-menarchal occipital PAF was linked with greater subsequent menstrual pain severity, and this faster PAF-higher menstrual pain relationship also existed in the early (3-14 months) but not late (15-30 months) post-menarche period. In contrast, oscillatory power and functional connectivity did not distinguish the groups at any visit, suggesting the widespread alterations seen in adults emerge later. To our knowledge, this is the first study describing an association between PAF and menstrual pain in adolescents, and the first time a neurophysiological trait has been linked with future menstrual pain development.

### 4.1 Menstrual pain was linked to PAF speed

PAF is thought to reflect the resonant frequency of the thalamocortical system [56] – indeed, an fMRI-EEG study in men linked PAF with resting activity in the thalamus, insula, cingulate, and sensory cortices [39]. Considered a relatively stable neural trait [18], a slower PAF has been reported in some chronic pain conditions although the findings are not always consistent [14] and can be influenced by sex and diagnosis [12]. In healthy adults, a slower resting PAF predicted increased sensitivity to prolonged [8] but not acute experimental pain [35], although the mechanism linking PAF with pain is not known. One hypothesis is that faster PAF represents a greater ability to inhibit incoming sensory input, such as nociceptive signals [36]. Others suggest PAF represents cognitive-affective aspects of pain processing, including modulation of attention and arousal [14]. Indeed, a recent study found slower PAF in patients with chronic low back pain and high fear of movement compared to patients with less fear [23], and slower PAF is associated with enhanced facilitation of nociception in response to unpleasant, threatening images [24].

In contrast, our study found a faster pre-menarche occipital PAF in adolescents who would later report higher menstrual pain. We also saw a relationship between faster PAF and current higher menstrual pain at the first post-menarche visit, but not at the second. Whether this is due to smaller post-menarche sample sizes unable to detect the modest brain-behaviour association, or a developmental shift in the PAF – pain relationship across the menarchal transition is unclear. The neurocognitive associations of PAF can change over time – for example, in young children PAF reflects sensorimotor rather than cognitive abilities [40], and PAF may vary across the menstrual cycle [5]. One cross-sectional study of adolescents found PAF increased during experimental pain in those with chronic pain, but at rest was not different from controls [48]. Faster PAF was seen during rest and pain in participants reporting higher vs. lower pain scores during a tonic pain task [34,47], which the authors attributed to increased thalamocortical feedback loop activity and greater excitability as opposed to increased sensory gating and inhibition. The underlying neurobiological explanation for why a faster PAF related to higher pain in our cohort is difficult to ascertain from this dataset, as EEG was measured at-rest and off-menses. Faster PAF could reflect greater attention to pain [25], and therefore increased likelihood of reporting painful experiences. Chronic widespread pain has also been linked with faster PAF [38], and higher menstrual pain in our cohort was previously associated with more widespread pain [49]. Overall, a faster peri-menarchal PAF may represent one neural trait in a larger physiological profile predisposing an individual to pain.

While PAF is considered relatively stable in adulthood, in adolescence it is dynamic. A developmental increase in PAF is a hallmark of a maturing brain [44] that was replicated in our cohort. The neurophysiological underpinnings of a maturing PAF are not known, although increased myelination of axons [6,65] have been proposed to lead to a faster PAF. It is possible that the adolescents who reported higher future menstrual pain experienced earlier pre-menarchal brain maturation. Stress [1,60], pain [3], and early puberty [37] can accelerate neurodevelopment in adolescence, and ABCD data recently linked advanced puberty at ages 9-10 with increased risk for dysmenorrhea at ages 12-13 [28]. However, no differences in stress, weekly pain intensity, age, or Tanner pubertal stage were seen between future menstrual pain groups at our pre-menarchal visit. Therefore, the origin of this faster occipital pre-menarchal PAF is unclear, although post-menarche PAF trajectories did not differ between higher and lower menstrual pain.

### 4.2 Oscillatory power and theta functional connectivity decreased across the menarchal transition

Global EEG power and functional connectivity changed across the menarchal transition in patterns consistent with typical neurodevelopment, with stratified analyses suggesting the timing of these changes were influenced by menstrual pain. Decreases in theta – gamma power from the pre-menarche to the first post-menarche visit mirrored overall global decreases in power reported from childhood to adolescence [58]. Adolescent decreases in resting state EEG power [68] are thought to be driven in part by the well-documented developmental decreases in gray matter [43] co-occurring with substantial synaptic pruning to create more refined, specialized neural networks [7,52]. Alpha, beta, and theta power also decreased from post-menarche visits 1 to 2 for adolescents with lower menstrual pain. Why power decreases in the later post-menarche period were not apparent in the higher menstrual pain group is unclear. Unlike PAF, it is difficult to pinpoint a “mature” pattern of oscillatory power to use as a benchmark of neurodevelopment, therefore the absence of oscillatory power decreases could hint at earlier neurocognitive development in this group or instead represent a stalled development.

We observed marked reductions in theta functional connectivity across the menarchal transition. These changes frequently involved the dorsal and ventral attention, control, and default mode networks – widely distributed networks involving prefrontal and associative cortices known to mature during adolescence [53]. Consistent with synaptic pruning, theta connectivity within the default mode, salience, and central executive networks is stronger in younger than older adolescents [32]. These same distributed networks undergoing refinement during adolescence show altered connectivity in adults with chronic pain and dysmenorrhea [29,30,51,61,66,67], therefore their development may be shaped by adolescent pain experience [3]. Stratified analyses revealed theta connectivity decreases around menarche in adolescents with lower menstrual pain and in the later post-menarche period in adolescents with higher menstrual pain – suggesting future studies should explore whether the neurobiological changes thought to underlie connectivity alterations [59] are influenced by menstrual pain.

Puberty has been proposed as a “sensitive period” [53] for neuroplasticity, and the coinciding biopsychosocial changes may be especially relevant to shaping current and future pain experience [46]. Moderate to severe menstrual pain in adolescence increases the risk of chronic pain in adulthood, indicating potential early maladaptive plasticity [55]. Indeed, a study of adolescents aged 13 to 19 years found associations between menstrual pain severity, interference, and cumulative menstrual pain exposure with functional connectivity of multiple brain regions, including the central executive, salience, and default mode networks [51]. In contrast, early menstrual pain was not associated with significant reorganization of the EEG functional connectome in our cohort, whose mean (± SD) ages at the first (12.9 ± 1.1) and second (14.1± 1.1) post-menarche visits were several years younger than the mean age (17 ± 1.8 years) of the cohort studied by Payne et al. [51]. This suggests that, apart from occipital PAF, the menstrual pain related-brain alterations seen in adults [29,61,67,69] may accumulate over adolescence rather than occur immediately surrounding menarche. Future studies characterizing how cyclic pain experiences are reflected in the mid-to-late adolescent brain could help us better understand how this chronic pain trajectory occurs, and what interventions may be able to disrupt it. Non-invasive neurostimulation for PAF has some success in decreasing pain [9], although the ability of these techniques to alter PAF in healthy individuals has been questioned [42] and more personalized approaches incorporating individual’s specific peak frequency may be more successful [11].

### 4.3 Limitations

EEG data loss was considerable owing to motion in this younger population, although stringent cleaning procedures ensured that retained data were of high quality. Estimating functional connectivity, particularly between ROIs located deeper in the brain, is more difficult with the ill-posed nature of the EEG inverse problem and the source-localization limitations of cortical EEG, even with 64 channels [19]. Our pubertal staging relied on self-reported breast and pubic hair development, which is less accurate than clinician assessment [54]. Our cohort was also predominantly White and from higher-income households, which may limit the generalizability of these findings.

## 5. Conclusion

For the first time, we identified pre-menarchal PAF as a possible neural sign of future menstrual pain intensity and linked faster peri-menarchal PAF speed with higher menstrual pain. Many of the functional brain alterations reported in adults with dysmenorrhea were absent in our adolescent cohort, suggesting that menstrual pain-related brain changes may take years to develop; nonetheless, the PAF–pain link observed here indicates that susceptibility to menstrual pain may be at least partly encoded in resting brain function even before menarche. The changes in cortical EEG oscillatory power, PAF, and functional connectivity we observed across the menarchal transition align with established patterns of adolescent neurodevelopment, and some may show menstrual pain-specific timings.

## Data Availability

Behavioural data and code for this study is available on Open Science Framework (https://doi.org/10.17605/OSF.IO/URKD7). EEG data is available upon request.

## Acknowledgements

The authors thank Drs. G.F. Gebhart and Lynn S. Walker for advice on study design and interpretation. We acknowledge our laboratory staff for their efforts in coordinating and collecting data for the EMPATHY study; Ellen Garrison, Dina Vavarutsos, Katharine Jabaay, Emily Gal, Katlyn Dillane & Genevieve Roth. We emphatically thank all the adolescents and parents/guardians for their enthusiastic participation.

This study is supported by the National Institutes of Health (NIH) grant R01HD096332.

## Supplemental digital content

Supplemental digital content associated with this article can be found online at [link].

## References

[1] Belsky J. Early-Life Adversity Accelerates Child and Adolescent Development. Curr Dir Psychol Sci 2019;28:241–246.

[2] Bevans KB, Gardner W, Pajer KA, Becker B, Carle A, Tucker CA, Forrest CB. Psychometric Evaluation of the PROMIS® Pediatric Psychological and Physical Stress Experiences Measures. J Pediatr Psychol 2018;43:678–692.

[3] Bhatt RR, Gupta A, Mayer EA, Zeltzer LK. Chronic Pain in Children: Structural and Resting-State Functional Brain Imaging Within a Developmental Perspective. Pediatr Res 2020;88:840–849.

[4] Bott FS, Zebhauser PT, Hohn VD, Turgut Ö, May ES, Tiemann L, Gil Ávila C, Heitmann H, Nickel MM, Day MA, Adhia DB, Ashar YK, Wager TD, Granovsky Y, Yarnitsky D, Jensen MP, Gross J, Ploner M. Exploring electroencephalographic chronic pain biomarkers: a mega-analysis. eBioMedicine 2025;120:105955.

[5] Brötzner CP, Klimesch W, Doppelmayr M, Zauner A, Kerschbaum HH. Resting state alpha frequency is associated with menstrual cycle phase, estradiol and use of oral contraceptives. Brain Research 2014;1577:36–44.

[6] Caffarra S, Kanopka K, Kruper J, Richie-Halford A, Roy E, Rokem A, Yeatman JD. Development of the Alpha Rhythm Is Linked to Visual White Matter Pathways and Visual Detection Performance. J Neurosci 2024;44:e0684232023.

[7] Chad JA, Lebel C. Can gray matter loss in early adolescence be explained by white matter growth? Hum Brain Mapp 2024;45:e26758.

[8] Chowdhury NS, Bi C, Furman AJ, Chiang AKI, Skippen P, Si E, Millard SK, Margerison SM, Spies D, Keaser ML, Da Silva JT, Chen S, Schabrun SM, Seminowicz DA. Predicting Individual Pain Sensitivity Using a Novel Cortical Biomarker Signature. JAMA Neurol 2025;82:237.

[9] Chowdhury NS, Taseen KJ, Chiang AK, Chang W-J, Millard SK, Seminowicz DA, Schabrun SM. A 5-day course of repetitive transcranial magnetic stimulation before pain onset ameliorates future pain and increases sensorimotor peak alpha frequency. PAIN 2025;166:1382.

[10] Duffy KA, Wiglesworth A, Roediger DJ, Island E, Mueller BA, Luciana M, Klimes-Dougan B, Cullen KR, Fiecas MB. Characterizing the effects of age, puberty, and sex on variability in resting-state functional connectivity in late childhood and early adolescence. NeuroImage 2025;313:121238.

[11] Fathi Y, Dissassuca F, Ricci G, Liberati G. Individualised Alpha-tACS for Modulating Pain Perception and Neural Oscillations: A Sham-Controlled Study in Healthy Participants. European Journal of Pain 2025;29:e70086.

[12] Fauchon C, Kim JA, El-Sayed R, Osborne NR, Rogachov A, Cheng JC, Hemington KS, Bosma RL, Dunkley BT, Oh J, Bhatia A, Inman RD, Davis KD. Exploring sex differences in alpha brain activity as a potential neuromarker associated with neuropathic pain. Pain 2022;163:1291–1302.

[13] Furman AJ, Prokhorenko M, Keaser ML, Zhang J, Chen S, Mazaheri A, Seminowicz DA. Sensorimotor Peak Alpha Frequency Is a Reliable Biomarker of Prolonged Pain Sensitivity. Cereb Cortex 2020;30:6069–6082.

[14] Garcia-Larrea L, André-Obadia N. Electroencephalography, magnetoencephalography and pain: where do we stand, and where are we going. PAIN 2025;166:S65.

[15] Gil Ávila C, Bott FS, Tiemann L, Hohn VD, May ES, Nickel MM, Zebhauser PT, Gross J, Ploner M. DISCOVER-EEG: an open, fully automated EEG pipeline for biomarker discovery in clinical neuroscience. Sci Data 2023;10:613.

[16] Gmehlin D, Thomas C, Weisbrod M, Walther S, Pfüller U, Resch F, Oelkers-Ax R. Individual analysis of EEG background-activity within school age: impact of age and sex within a longitudinal data set. International Journal of Developmental Neuroscience 2011;29:163–170.

[17] Gottschewsky N, Kraft D, Kaufmann T. Menarche, pubertal timing and the brain: female-specific patterns of brain maturation beyond age-related development. Biol Sex Differ 2024;15. doi:10.1186/s13293-024-00604-4.

[18] Grandy TH, Werkle-Bergner M, Chicherio C, Schmiedek F, Lövdén M, Lindenberger U. Peak individual alpha frequency qualifies as a stable neurophysiological trait marker in healthy younger and older adults. Psychophysiology 2013;50:570–582.

[19] Grech R, Cassar T, Muscat J, Camilleri KP, Fabri SG, Zervakis M, Xanthopoulos P, Sakkalis V, Vanrumste B. Review on solving the inverse problem in EEG source analysis. J NeuroEngineering Rehabil 2008;5:25.

[20] Harris PA, Taylor R, Minor BL, Elliott V, Fernandez M, O’Neal L, McLeod L, Delacqua G, Delacqua F, Kirby J, Duda SN, REDCap Consortium. The REDCap consortium: Building an international community of software platform partners. J Biomed Inform 2019;95:103208.

[21] Hays RD, Spritzer KL, Schalet BD, Cella D. PROMIS®-29 v2.0 Profile Physical and Mental Health Summary Scores. Qual Life Res 2018;27:1885–1891.

[22] Hemington KS, Wu Q, Kucyi A, Inman RD, Davis KD. Abnormal cross-network functional connectivity in chronic pain and its association with clinical symptoms. Brain Struct Funct 2016;221:4203–4219.

[23] Ho RLM, Park J, Wang W, Thomas JS, Cruz-Almeida Y, Coombes SA. Lower individual alpha frequency in individuals with chronic low back pain and fear of movement. Pain 2024;165:1033–1043.

[24] Huber FA, Kell PA, Shadlow JO, Rhudy JL. Cerebral peak alpha frequency: Associations with chronic pain onset and pain modulation. Neurobiol Pain 2025;18:100180.

[25] Kelardashti N, Dunkley BT, El-Sayed R, Sanmugananthan VV, Kim JA, Osborne NR, Cheng JC, Rogachov A, Bosma RL, Besik AE, Davis KD. Alpha and Theta Oscillations Associated With Behavioral Phenotypes of Pain–Attention Interaction. Brain Behav 2025;15:e70190.

[26] Kuo P-C, Chen Y-T, Chen Y-S, Chen L-F. Decoding the perception of endogenous pain from resting-state MEG. NeuroImage 2017;144:1–11.

[27] Lacadie CM, Fulbright RK, Constable RT, Papademetris X. More Accurate Talairach Coordinates for NeuroImaging using Nonlinear Registration. Neuroimage 2008;42:717– 725.

[28] Lenert ME, Smith T, Hidalgo-Lopez E, Portengen CM, Harte SE, Beltz AM, Schrepf A, Kaplan CM. Premenarche risk factors for future dysmenorrhoea: a prospective cohort study. The Lancet Child & Adolescent Health 2026;10:710–718.

[29] Liu H, Su X, Shang M, Ma L, Bai W, Wang H, Quan L, Li Y, Huang Z, He J, Dun W, Zhang Y. Abnormal dynamic functional networks during pain-free periods: Resting-state co-activation pattern analysis in primary dysmenorrhea. NeuroImage 2025;306:121009.

[30] Low I, Wei S-Y, Lee P-S, Li W-C, Lee L-C, Hsieh J-C, Chen L-F. Neuroimaging Studies of Primary Dysmenorrhea. Adv Exp Med Biol 2018;1099:179–199.

[31] Lund CI, Engdahl B, Rosseland LA, Stubhaug A, Grimnes G, Furberg A-S, Steingrímsdóttir ÓA, Nielsen CS. The association between age at menarche and chronic pain outcomes in women: the Tromsø Study, 2007 to 2016. Pain 2022;163:1790–1799.

[32] Machinskaya RI, Kurgansky AV, Lomakin DI. Age-related Trends in Functional Organization of Cortical Parts of Regulatory Brain Systems in Adolescents: an Analysis of Resting-State Networks in the EEG Source Space. Hum Physiol 2019;45:461–473.

[33] Marshall WA, Tanner JM. Variations in pattern of pubertal changes in girls. Arch Dis Child 1969;44:291–303.

[34] Martino ED, Gregoret L, Zandalasini M, Graven-Nielsen T. Slowing in Peak-Alpha Frequency Recorded After Experimentally-Induced Muscle Pain is not Significantly Different Between High and Low Pain-Sensitive Subjects. The Journal of Pain 2021;22:1722–1732.

[35] May ES, Tiemann L, Gil Ávila C, Bott FS, Hohn VD, Gross J, Ploner M. Assessing the predictive value of peak alpha frequency for the sensitivity to pain. Pain 2025;166:2076– 2090.

[36] Mazaheri A, Seminowicz DA, Furman AJ. Peak alpha frequency as a candidate biomarker of pain sensitivity: the importance of distinguishing slow from slowing. NeuroImage 2022;262:119560.

[37] McCann CF, Cheng TW, Mills KL, Silvers JA. Pubertal timing and tempo differentially influence cortical and subcortical maturation in adolescence. Dev Cogn Neurosci 2025;77:101657.

[38] McLain N, Cavaleri R, Kutch J. Peak alpha frequency differs between chronic back pain and chronic widespread pain. European Journal of Pain 2025;29:e4737.

[39] McLain NJ, Yani MS, Kutch JJ. Analytic consistency and neural correlates of peak alpha frequency in the study of pain. J Neurosci Methods 2022;368:109460.

[40] Mierau A, Felsch M, Hülsdünker T, Mierau J, Bullermann P, Weiß B, Strüder HK. The interrelation between sensorimotor abilities, cognitive performance and individual EEG alpha peak frequency in young children. Clin Neurophysiol 2016;127:270–276.

[41] Millard SK, Furman AJ, Kerr A, Seminowicz DA, Gao F, Naidu BV, Mazaheri A. Predicting postoperative pain in lung cancer patients using preoperative peak alpha frequency. Br J Anaesth 2022;128:e346–e348.

[42] Millard SK, Speis DB, Skippen P, Chiang AKI, Chang W-J, Lin AJ, Furman AJ, Mazaheri A, Seminowicz DA, Schabrun SM. Can non-invasive brain stimulation modulate peak alpha frequency in the human brain? A systematic review and meta-analysis. European Journal of Neuroscience 2024;60:4182–4200.

[43] Mills KL, Goddings A-L, Herting MM, Meuwese R, Blakemore S-J, Crone EA, Dahl RE, Güroğlu B, Raznahan A, Sowell ER, Tamnes CK. Structural brain development between childhood and adulthood: Convergence across four longitudinal samples. Neuroimage 2016;141:273–281.

[44] Miskovic V, Ma X, Chou C-A, Fan M, Owens M, Sayama H, Gibb BE. Developmental changes in spontaneous electrocortical activity and network organization from early to late childhood. NeuroImage 2015;118:237–247.

[45] Moore RA, Straube S, Aldington D. Pain measures and cut-offs - “no worse than mild pain” as a simple, universal outcome. Anaesthesia 2013;68:400–412.

[46] Nahman-Averbuch H, Li R, Boerner KE, Lewis C, Garwood S, Palermo TM, Jordan A. Alterations in pain during adolescence and puberty. Trends in Neurosciences 2023;46:307– 317.

[47] Nir R-R, Sinai A, Raz E, Sprecher E, Yarnitsky D. Pain assessment by continuous EEG: Association between subjective perception of tonic pain and peak frequency of alpha oscillations during stimulation and at rest. Brain Research 2010;1344:77–86.

[48] Ocay DD, Teel EF, Luo OD, Savignac C, Mahdid Y, Blain-Moraes S, Ferland CE. Electroencephalographic characteristics of children and adolescents with chronic musculoskeletal pain. Pain Rep 2022;7:e1054.

[49] Osborne NR, Hellman KM, Burda EM, Darnell SE, Singh L, Schrepf AD, Walker LS, Tu FF. Multimodal hypersensitivity and somatic symptoms predict adolescent postmenarchal widespread pain. Pain 2025;166:e322–e335.

[50] Paus T, Keshavan M, Giedd JN. Why do many psychiatric disorders emerge during adolescence? Nat Rev Neurosci 2008;9:947–957.

[51] Payne LA, Seidman LC, Napadow V, Nickerson LD, Kumar P. Functional connectivity associations with menstrual pain characteristics in adolescents: an investigation of the triple network model. Pain 2025;166:338–346.

[52] Petanjek Z, Judaš M, Šimic G, Rasin MR, Uylings HBM, Rakic P, Kostovic I. Extraordinary neoteny of synaptic spines in the human prefrontal cortex. Proc Natl Acad Sci U S A 2011;108:13281–13286.

[53] Piekarski DJ, Johnson CM, Boivin JR, Thomas AW, Lin WC, Delevich K, M. Galarce E, Wilbrecht L. Does puberty mark a transition in sensitive periods for plasticity in the associative neocortex? Brain Research 2017;1654:123–144.

[54] Rasmussen AR, Wohlfahrt-Veje C, Tefre de Renzy-Martin K, Hagen CP, Tinggaard J, Mouritsen A, Mieritz MG, Main KM. Validity of Self-Assessment of Pubertal Maturation. Pediatrics 2015;135:86–93.

[55] Reid-McCann R, Poli-Neto OB, Stein K, Dixon S, Cox E, Coxon L, Fazel M, Noonan M, Sharp GC, Zondervan K, Vincent K. Longitudinal association between dysmenorrhoea in adolescence and chronic pain in adulthood: a UK population-based study. The Lancet Child & Adolescent Health 2025;0. doi:10.1016/S2352-4642(25)00213-5.

[56] Robinson PA, Rennie CJ, Wright JJ, Bahramali H, Gordon E, Rowe DL. Prediction of electroencephalographic spectra from neurophysiology. Phys Rev E Stat Nonlin Soft Matter Phys 2001;63:021903.

[57] Schaefer A, Kong R, Gordon EM, Laumann TO, Zuo X-N, Holmes AJ, Eickhoff SB, Yeo BTT. Local-Global Parcellation of the Human Cerebral Cortex from Intrinsic Functional Connectivity MRI. Cereb Cortex 2018;28:3095–3114.

[58] Segalowitz SJ, Santesso DL, Jetha MK. Electrophysiological changes during adolescence: A review. Brain and Cognition 2010;72:86–100.

[59] Selemon LD. A role for synaptic plasticity in the adolescent development of executive function. Transl Psychiatry 2013;3:e238–e238.

[60] Sisk LM, Gee DG. Stress and adolescence: vulnerability and opportunity during a sensitive window of development. Curr Opin Psychol 2022;44:286–292.

[61] Su X, Li Y, Liu H, An S, Yao N, Li C, Shang M, Ma L, Yang J, Li J, Zhang M, Dun W, Huang Z-G. Brain Network Dynamics in Women With Primary Dysmenorrhea During the Pain-Free Periovulation Phase. The Journal of Pain 2024;25:104618.

[62] Ta Dinh S, Nickel MM, Tiemann L, May ES, Heitmann H, Hohn VD, Edenharter G, Utpadel-Fischler D, Tölle TR, Sauseng P, Gross J, Ploner M. Brain dysfunction in chronic pain patients assessed by resting-state electroencephalography. Pain 2019;160:2751–2765.

[63] Tu C-H, Niddam DM, Chao H-T, Chen L-F, Chen Y-S, Wu Y-T, Yeh T-C, Lirng J-F, Hsieh J-C. Brain morphological changes associated with cyclic menstrual pain. Pain 2010;150:462–468.

[64] Tu FF, Hellman KM, Darnell SE, Harber KA, Bohnert AM, Singh L, Walker LS. A multidimensional appraisal of early menstrual pain experience. Am J Obstet Gynecol 2024;230:550.e1–550.e10.

[65] Valdés-Hernández PA, Ojeda-González A, Martínez-Montes E, Lage-Castellanos A, Virués-Alba T, Valdés-Urrutia L, Valdes-Sosa PA. White matter architecture rather than cortical surface area correlates with the EEG alpha rhythm. NeuroImage 2010;49:2328– 2339.

[66] Vincent K, Warnaby C, Stagg CJ, Moore J, Kennedy S, Tracey I. Dysmenorrhoea is associated with central changes in otherwise healthy women. Pain 2011;152:1966–1975.

[67] Wei S-Y, Chao H-T, Tu C-H, Li W-C, Low I, Chuang C-Y, Chen L-F, Hsieh J-C. Changes in functional connectivity of pain modulatory systems in women with primary dysmenorrhea. Pain 2016;157:92–102.

[68] Whitford TJ, Rennie CJ, Grieve SM, Clark CR, Gordon E, Williams LM. Brain maturation in adolescence: Concurrent changes in neuroanatomy and neurophysiology. Human Brain Mapping 2007;28:228–237.

[69] Yang L, Dun W, Li K, Yang J, Wang K, Liu H, Liu J, Zhang M. Altered amygdalar volume and functional connectivity in primary dysmenorrhoea during the menstrual cycle. European Journal of Pain 2019;23:994–1005.

