## supplement for "Faster peak alpha frequency precedes higher menstrual pain in adolescence"

#### Methods

All Early Menstrual Pain Impact on Multisensory Hypersensitivity (EMPATHY) study visits took place in our experimental testing rooms located at Evanston Hospital. The pre-menarche visit took approximately 3.5 hours to complete, while the post-menarche visits took approximately 2.5 to 3 hours. Recruitment took place from January 2019 to August 2022, with in-person study visits taking place from January 2019 to August 2024. The original sample size for EMPATHY (n=300) was calculated to provide 80% power to detect a medium effect size (R2 =.33, equivalent to Cohen's d= 3.5) for EMPATHY's primary aim, which was to characterize the trajectory of dysmenorrhea in the 2 years post-menarche and determine demographic, physiological, and psychological factors associated with these trajectories.

#### Questionnaire data: Additional analyses

To determine if puberty stage differed between higher and lower menstrual pain groups and therefore could potentially contribute to observed differences in EEG characteristics as has been previously shown with structural MRI [2], self-assessed pubertal scores for breast and pubic hair were summed into one pubertal score and compared between groups at each study visit using Welch's t-tests. Similarly, the age at which participants reached menarche was also compared between the higher and lower menstrual pain groups at the post-menarche visits using Welch's t-tests, as gynecological age may influence adolescent brain function [5].

#### EEG: Preprocessing and feature extraction

##### EEG preprocessing

We split our resting state task into two separate 4 minute resting states: one with eyes closed data and the other with eyes open data. All subsequent preprocessing and analyses were run separately for the eyes open and eyes closed tasks. We used the DISCOVER-EEG pipeline [4] to preprocess our EEG data, which is described in detail at https://github.com/crisglav/discover-eeg. Briefly, data was downsampled to 250 Hz and then line noise (60 Hz) was removed using EEGLAB (v2023.1)'s *pop_cleanline()* function. Then, EEGLAB's *pop_clean_rawdata()* function was run, which applies a high pass filter with a transition band of 0.25 to 0.75 Hz using the *clean_drifts()* function. Artifactual channels were removed if they 1) flatlined for 5 seconds or more, 2) had a z-scored noise-to-signal ratio higher than 4 or 3) if the channel's timecourse could not be predicted from a randomly selected subset of remaining channels at least 70% of the recorded time. Data was then re-referenced to the average reference using the *pop_reref()* function. Independent Component Analysis (ICA) was then performed on clean channels using the function *pop_runica()*, which classified artifacts into seven categories (brain, muscle, eye, heart, line noise, channel noise and other). Components with an 80% or higher probability of being muscle or eye were removed from the data. To keep the number of channels across participants constant, bad channels that were removed with the *pop_clean_rawdata()* function were interpolated using spherical splines with the function *pop_interp()*. Bad time segments – portions of the recording where the data's variance was larger than 20 times the standard deviation of the clean "calibration" data segments - were identified by *pop_clean_rawdata()*'s artifact subspace reconstruction method and removed. The ICA, channel interpolation, and bad time segment removal steps were performed 10 times and the bad time segment mask that most resembled the average bad time segment mask across the 10 repetitions was selected. Finally, data was segmented into 2 second epochs with a 50% overlap using the function *pop_epoch()*. The 2 second epoch length was chosen by the discoverEEG authors as a balance between the longer epochs ideal for peak alpha frequency (PAF) calculation and the shorter epochs suited to measures of functional connectivity. If an epoch contained a discontinuity – e.g., due to a discarded segment, the epoch was automatically rejected.

We performed data quality analyses to decide which participants had sufficient data remaining after preprocessing to include in the group analyses. Our criteria were as follows; participants were removed from the group analyses if they had either 1) more than 50% of the 64 channels removed 2) if between 20 and 50% of their channels had been removed, we inspected the participant for focal channel loss – i.e., no more than 4 neighbouring channels could be removed and 3) more than 50% of epochs were removed. Therefore, this final step of data quality control resulted in n=250 participants with EEG data for the group analyses of the premenarche visit, n=161 participants for post-menarche visit 1 and n=140 participants for post-menarche visit 2.

After preprocessing, the discoverEEG pipeline calculates several EEG features for each participant and each task (eyes-open and eyes-closed), including power spectra from 1 to 100 Hz, PAF, and functional connectivity (in source space). These EEG feature extraction steps are described briefly below, but complete methodological details for the pipeline can be found in the accompanying publication [4].

#### Power spectrum calculation

The FieldTrip function ft_freqanalysis was used to compute power spectra between 1 and 100 Hz using Slepian multitapers with +/−1 Hz frequency smoothing. First, power spectra were computed for every channel. Then, they were averaged across the 2 second epochs and channels to produce a single global power spectrum for each individual and each task (eyes-open and eyes-closed). The DISCOVER-EEG pipeline zero-padded the epochs to 5 seconds to create a spectral resolution of 0.1 Hz that would better capture the frequency range of the bands. The frequency band limits used by the pipeline were 4 to 7.9 Hz (theta), 8 to 12.9 Hz (alpha), 13 to 30 Hz (beta) and 30.1 to 80 Hz (gamma) [11].

#### Peak alpha frequency calculation

Peak alpha frequency (PAF) was calculated from the global power spectrum in the alpha range (8 to 12.9 Hz) using the center of gravity approach, which is calculated as the weighted average of alpha frequencies with each frequency weighted by their power. Global PAF linear mixed models (LMMs) indicated a difference in PAF between higher and lower menstrual pain groups at the pre-menarche visit, but this was not significant ($p$ = 0.066). Therefore, an exploratory regional PAF analyses was performed, as both global [10] and regional [3,9] PAF have been linked to pain sensitivity and symptoms. Topoplots of alpha (8 – 12.9 Hz) activity across all participants were created in EEGLab [1] and the occipital cortex was identified as a region of interest for alpha power. Regional PAF was calculated from a selection of channels corresponding to the occipital cortex (Chs 17, 18, 47, 48 and 49 on the CACS-64-X7 acticap electrode set).

#### Peak alpha frequency statistical modeling

The full-cohort trajectory model included n = 174/176 (eyes-closed/eyes-open) at the pre-menarche visit, n = 160/155 at post-menarche visit 1, and n = 134/132 at post-menarche visit 2. Likelihood-ratio tests confirmed that adding a random slope for visit improved fit over a random-intercept-only model in both conditions (eyes-closed $\chi^2(2) = 9.07$, $p = .011$; eyes-open

$\chi^2(2) = 11.86$, $p = 0.003$). To localise the developmental increase to specific visit pairs for Figure 3A, a factor-coded model [PAF ~ visit_f + (1 + visit_num | subject_ID)] was also fitted, with consecutive contrasts (pre-menarche → post-menarche visit 1; visit 1 → visit 2) Holm-corrected across the two within-condition comparisons. In the group-comparison model, the pre-menarche visit included eyes-closed/open PAF from n = 91/94 higher- and n = 83/82 lower-pain participants; post-menarche visit 1 from n = 86/82 higher and n = 74/73 lower; and post-menarche visit 2 from n = 77/76 higher and n = 57/56 lower. Visit-specific contrasts were the higher-minus-lower difference (reverse pairwise), pooled within condition into a single family for Holm correction. Population-level trajectories for visualisation were marginal predictions (marginaleffects, re.form = NA). Significance was $p < 0.05$, two-tailed.

#### Functional connectivity calculation

First, an atlas-based beamforming approach was used to perform source reconstruction. Band-pass filtered data from sensor space was projected into source space for each frequency band with an array-gain Linear Constrained Minimum Variance (LCMV) beamformer. The centroids of 100 ROIs from the 7-network version of the Schaefer atlas were used for the source model [12], which included Visual, Somato-Motor, Dorsal attention, Salience-Ventral attention, Limbic, Control, and Default networks (see Supplementary Table 1). A conduction model based on FieldTrip's standard_bem.mat template and the source model were used to build the lead field. Then, spatial filters were created with the covariance matrices of the band-pass filtered data and the lead fields. A regularization parameter of 5% was set for the covariance matrix, and the dipole orientation was fixed to the direction of the maximum variance. Power for each source location was estimated for each participant using the spatial filter and the band-passed data.

Next, virtual time series of the 100 source locations (ROIs) were reconstructed for each frequency band by applying the band-specific spatial filters to the band-pass filtered sensor data. Functional connectivity was calculated for each frequency band using the amplitude envelope correlation (AEC) method and a custom function *compute_aec* (see https://github.com/crisglav/discover-eeg). For each 2 second epoch, a Hilbert transform was used to extract the analytical signal of the virtual time series at each of the 100 source locations. The signal at source A was orthogonalized with respect to the signal at source B for each source pair (yielding the signal A⊥B). To calculate the average connectivity between source A and B, Pearson correlations were computed between the amplitude envelope of signals B and A⊥B and between signals A and B⊥A, and the resulting correlation coefficients were averaged. This resulted in 100 X 100 connectivity matrices for each epoch, which were averaged across epochs to create an average connectivity matrix for each of the four frequency bands for each participant.

### EEG: Sensitivity analyses

The following analyses were performed for each EEG metric to serve as sensitivity analyses for our group comparisons of higher and lower menstrual pain scores; 1) within-subject eyes-open vs. eyes-closed resting state EEG, 2) between-subject high vs. low weekly pain intensity, and 3) between-subject high vs. low depression. The EEG features – power, peak alpha frequency, and functional connectivity – were calculated using the same methods as described above and in the main manuscript for the menstrual pain group comparisons.

#### Eyes-open vs. eyes-closed comparison

Eyes-closed and eyes-open resting state paradigms yield different resting EEG characteristics [6]. Collecting both conditions provides a richer characterization of resting-state brain activity in adolescents. While comparing eyes open vs. closed EEG was not the main focus of this study, it does provide an opportunity to interrogate the quality of our EEG recordings by demonstrating the expected differences in oscillatory power between these two states, e.g., alpha power is consistently found to be greater in eyes-closed vs. eyes-open resting state [6].

Global PAF was compared between the eyes-closed and open conditions for the full cohort at each study visit, paired by participant. Given that within-session EEG condition differences may not be normally distributed, a paired Wilcoxon signed-rank test was used at each visit and only participants with valid (non-missing) PAF values for both conditions at a given visit contributed to that visit's test. Raw p-values from the three Wilcoxon tests were Holm-corrected within a single family (three visits). Adjusted p-values below .05 were considered statistically significant. Cross-sectional differences in spectral power (1 – 100 Hz) were calculated with dependent-samples cluster-based permutation tests, with Monte Carlo simulations (500 randomizations). As in the main analysis, the sum of the t-values within each cluster were used to calculate cluster-level statistics, with a cluster threshold and alpha level of 0.05. For functional connectivity, the connectivity values between each pair (undirected) of the 100 Schaefer atlas regions of interest (ROIs) for the theta, alpha, beta and gamma frequency bands were compared between eyes-open and eyes-closed conditions using 4,950 two-sided dependent samples t-tests for each connectivity matrix. Multiple comparison testing was addressed by controlling the false discovery rate (FDR) to $q < 0.05$.

#### Higher vs. lower weekly pain intensity

The PROMIS Pain Intensity item - which asks about pain intensity (0 -10) in the past week – was included to capture general pain experience in our cohort and explore its effects on EEG activity. Weekly pain intensity in this case referred to any pain the individual may be experiencing (e.g., headaches, muscle aches, stomach pain etc.,) and was not specific to menstruation. The questionnaire was completed at each study visit, which in the case of the post-menarche visits took place off-menses. To match our grouping of higher vs lower menstrual pain, we split participants into higher (≥ 3/10) and lower (≤ 2/10) weekly pain and compared EEG features between these two groups.

To examine whether global or occipital PAF differed between groups at each visit, independent-samples Welch's t-tests were performed at each of the three visits within each eyes-open/closed condition, yielding six tests per grouping variable (three visits × two conditions). Because Welch's t-test was selected to accommodate potential differences in group variance. To control the familywise error rate, raw p-values were adjusted using the Holm correction applied separately within each EEG condition (i.e., corrected across the three visits within EC and across the three visits within EO as independent families). Adjusted p-values below .05 were considered statistically significant. Cross-sectional differences in spectral power (1 – 100 Hz) were calculated with independent-samples cluster-based permutation tests, with Monte Carlo simulations (500 randomizations) for each study visit. As before, the sum of the t-values within each cluster were used to calculate cluster-level statistics, with a cluster threshold and alpha level of 0.05. Functional connectivity was compared between higher and lower weekly pain groups at each study visit. Functional connectivity values between each pair (undirected) of the 100 Schaefer atlas regions of interest (ROIs) for the theta, alpha, beta and gamma frequency bands were compared between eyes-open and eyes-closed conditions using 4,950 two-sided independent samples t-tests for each connectivity matrix. Multiple comparison testing was addressed by controlling the false discovery rate (FDR) to $q < 0.05$.

#### Higher vs. lower depression

Finally, we investigated the effects of depression on resting state EEG in adolescents as a control for our pain analyses, as mood disorders have been extensively investigated in neuroimaging studies of this age group.[7] We organized participants into higher and lower depression score groups based on their responses to the PROMIS Pediatric Profile for depression (PROMIS Ped SF v2.0 - Depressive Symptoms 8a), with the top 15% of scores at each study visit classified as the "higher" group, calculated dynamically for each study visit. Therefore, the depression score cut-off for the higher depression score group was ≥ 19 at the pre-menarche visit, ≥ 20 at the first post-menarche visit and ≥ 23 at the second post-menarche visit. Group differences in global and occipital PAF, spectral power, and functional connectivity between higher and lower depression scores were calculated identically to the weekly pain intensity analyses described above.

### Results

#### Questionnaire Data

Self-assessed tanner puberty stage did not differ between the higher [average (SD); 5.03 (1.49)] and lower [4.88 (1.4)] menstrual pain groups at the pre-menarche visit ($p$ = 0.499). After menarche, there were similarly no group differences in puberty stage at the first post-menarche visit [higher = 7.42 (1.36), lower = 7.39 (1.39); $p$ = 0.891] or the second post-menarche visit [higher = 8.13 (1.73), lower = 8.41 (1.25); $p$ = 0.28]. Age at menarche did not differ between adolescents in the higher and lower menstrual pain groups at post-menarche visit 1 [average (SD) higher = 12.45 (1.07) years, lower = 12.23 (1.02) years; $p$ = 0.171] nor at post-menarche visit 2 [ higher = 12.35 (1.19) years, lower = 12.32 (0.95) years; $p$ = 0.854].

#### Resting EEG features differ when eyes are open versus closed

We compared eyes-open vs. eyes-closed resting state EEG characteristics within adolescents at each study visit (Supplementary Figure 2). The most prominent findings were 1) greater theta, alpha, and beta power in the eyes-closed condition, 2) faster peak alpha frequency in the eyes-open condition, and 3) greater alpha and beta functional connectivity in eyes-closed and greater theta connectivity in eyes-open conditions. These patterns were consistent across all three study visits. Significant power differences not shown in Supplementary Figure 2 include greater eyes-open high gamma power at the pre-menarche visit (62 – 100 Hz, $p$ = 0.004), post-menarche visit 1 (63 – 86.7 Hz, $p$ = 0.01), and post-menarche visit 2 (62.1 – 100 Hz, $p$ = 0.006). Eyes-open PAF was significantly higher than eyes-closed PAF (Supplementary Figure 2) at the pre-menarche visit ($p < 0.001$), post-menarche visit 1 ($p = 0.002$) and post-menarche visit 2 ($p = 0.002$).

#### No group differences in resting state EEG for weekly pain intensity and depression

We investigated the effect of general weekly pain experience on our resting EEG features (Supplementary Figure 3). There were no significant differences in power between adolescents who reported higher weekly pain intensity (≥ 3/10) vs. lower weekly pain intensity (≤ 2/10) at any of the study visits. Similarly, there were no significant group differences in global or occipital PAF (Supplementary Figure 3, occipital PAF results displayed).

Another sensitivity analyses we performed involved comparing EEG features in adolescents who reported higher (top 15%) vs. lower depression scores (Supplementary Figure 4, occipital PAF results displayed). There were no significant differences in power or global or occipital PAF between our depression groups.

#### No differences in power or functional connectivity in higher vs. lower menstrual pain

In addition to peak alpha frequency, we also investigated the effects of menstrual pain on resting-state power and functional connectivity at each study visit. We did not see differences in eyes-open or closed power between the future higher (≥ 30) vs. lower (≤ 29) menstrual pain groups at the pre-menarche visit nor the higher vs. lower menstrual pain groups at post-menarche visits 1 and 2 (Supplementary Figure 5). Differences in functional connectivity between higher and lower menstrual pain at the pre-menarche as well as the first and second post-menarche visits did not survive FDR correction for multiple comparisons, however we present the connectivity matrices thresholded at uncorrected $p \leq 0.01$ for illustrative purposes (Supplementary Figure 6). Future investigations of menstrual pain's influence on functional connectivity may require more directed, region-specific analyses.

### References

[1] Delorme A, Makeig S. EEGLAB: an open source toolbox for analysis of single-trial EEG dynamics including independent component analysis. J Neurosci Methods 2004;134:9–21.

[2] Duffy KA, Wiglesworth A, Roediger DJ, Island E, Mueller BA, Luciana M, Klimes-Dougan B, Cullen KR, Fiecas MB. Characterizing the effects of age, puberty, and sex on variability in resting-state functional connectivity in late childhood and early adolescence. NeuroImage 2025;313:121238.

[3] Furman AJ, Prokhorenko M, Keaser ML, Zhang J, Chen S, Mazaheri A, Seminowicz DA. Sensorimotor Peak Alpha Frequency Is a Reliable Biomarker of Prolonged Pain Sensitivity. Cereb Cortex 2020;30:6069–6082.

[4] Gil Ávila C, Bott FS, Tiemann L, Hohn VD, May ES, Nickel MM, Zebhauser PT, Gross J, Ploner M. DISCOVER-EEG: an open, fully automated EEG pipeline for biomarker discovery in clinical neuroscience. Sci Data 2023;10:613.

[5] Gottschewsky N, Kraft D, Kaufmann T. Menarche, pubertal timing and the brain: female-specific patterns of brain maturation beyond age-related development. Biol Sex Differ 2024;15. doi:10.1186/s13293-024-00604-4.

[6] Isler JR, Pini N, Lucchini M, Shuffrey LC, Morales S, Bowers ME, Leach SC, Sania A, Wang L, Condon C, Nugent JD, Elliott AJ, Friedrich C, Andrew R, Fox NA, Myers MM, Fifer WP. Longitudinal characterization of EEG power spectra during eyes open and eyes closed conditions in children. Psychophysiology 2023;60. doi:10.1111/psyp.14158.

[7] MacSweeney N, Toenders YJ, Tamnes CK. Neuroimaging insights into adolescent depression risk and development. Nat Mental Health 2025;3:772–779.

[8] Marshall WA, Tanner JM. Variations in pattern of pubertal changes in girls. Arch Dis Child 1969;44:291–303.

[9] Mazaheri A, Seminowicz DA, Furman AJ. Peak alpha frequency as a candidate biomarker of pain sensitivity: the importance of distinguishing slow from slowing. NeuroImage 2022;262:119560.

[10] McLain N, Cavaleri R, Kutch J. Peak alpha frequency differs between chronic back pain and chronic widespread pain. Eur J Pain 2025;29:e4737.

[11] Pernet C, Garrido MI, Gramfort A, Maurits N, Michel CM, Pang E, Salmelin R, Schoffelen JM, Valdes-Sosa PA, Puce A. Issues and recommendations from the OHBM COBIDAS MEEG committee for reproducible EEG and MEG research. Nat Neurosci 2020;23:1473–1483.

[12] Schaefer A, Kong R, Gordon EM, Laumann TO, Zuo X-N, Holmes AJ, Eickhoff SB, Yeo BTT. Local-Global Parcellation of the Human Cerebral Cortex from Intrinsic Functional Connectivity MRI. Cereb Cortex 2018;28:3095–3114.

### Supplementary Figures

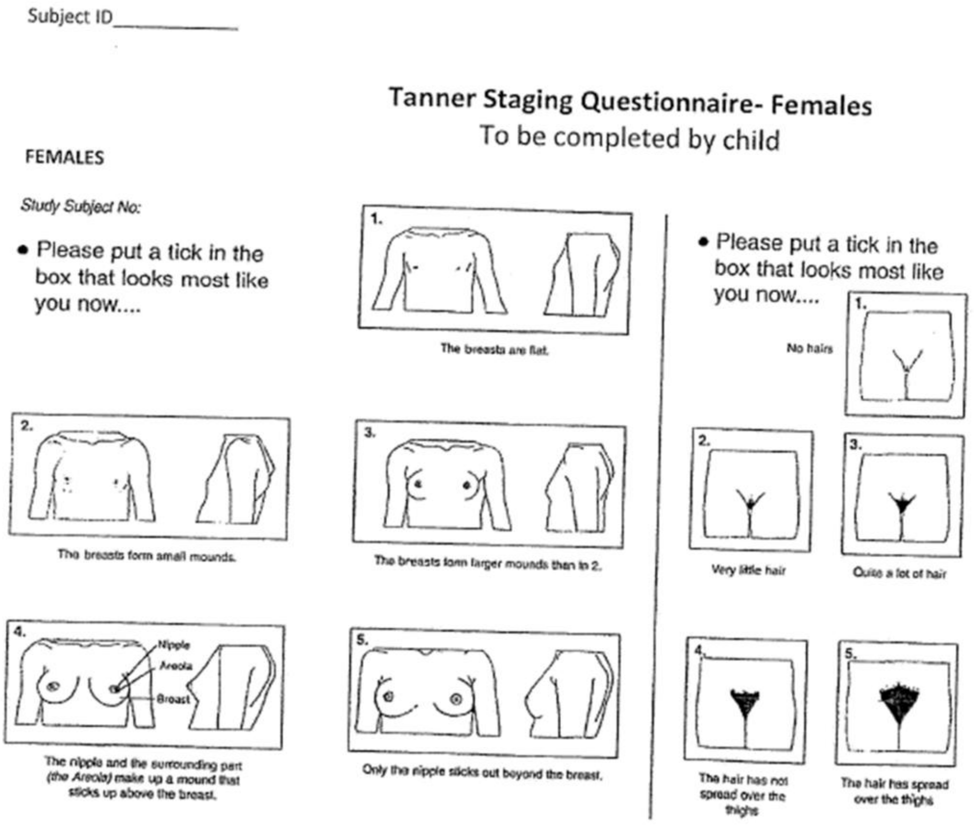

**Supplementary Figure 1. Modified Tanner Puberty Stage questionnaire.** Adolescents filled out this modified version of the Tanner Puberty Stage questionnaire [8] with questions about breast and pubic hair development at each study visit.

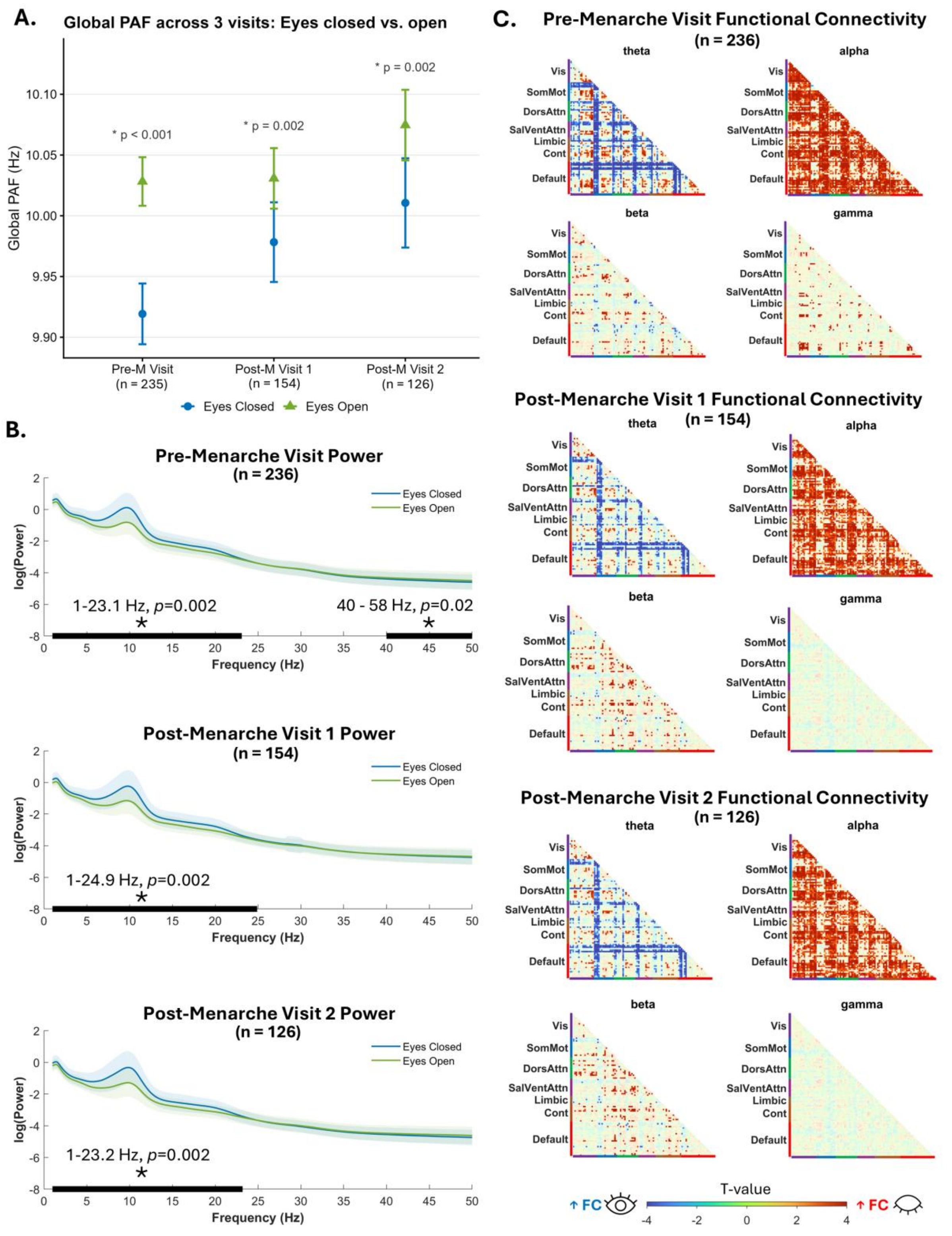

**Supplementary Figure 2. EEG characteristics of the eyes-open and eyes-closed resting state.** We compared eyes-open vs. eyes-closed resting state EEG characteristics within adolescents before and after menarche. The most pronounced findings – which were consistent at all three study visits - were 1) greater theta, alpha, and beta power in the eyes-closed condition, 2) faster PAF in the eyes-open condition, and 3) greater alpha and beta functional connectivity (FC) in eyes-closed and greater theta connectivity in eyes-open conditions. PAF values are displayed as group means ± SEM with * indicating significant ($p \leq 0.05$) Holm-adjusted p-values. Black bars and asterisks signify significant differences between conditions. Connectivity matrices are thresholded at FDR-corrected $q \leq 0.05$. Pre-M Visit = Pre-menarche visit, Post-M1 visit = Post-menarche visit 1, Post-M2 Visit = Post-menarche visit 2.

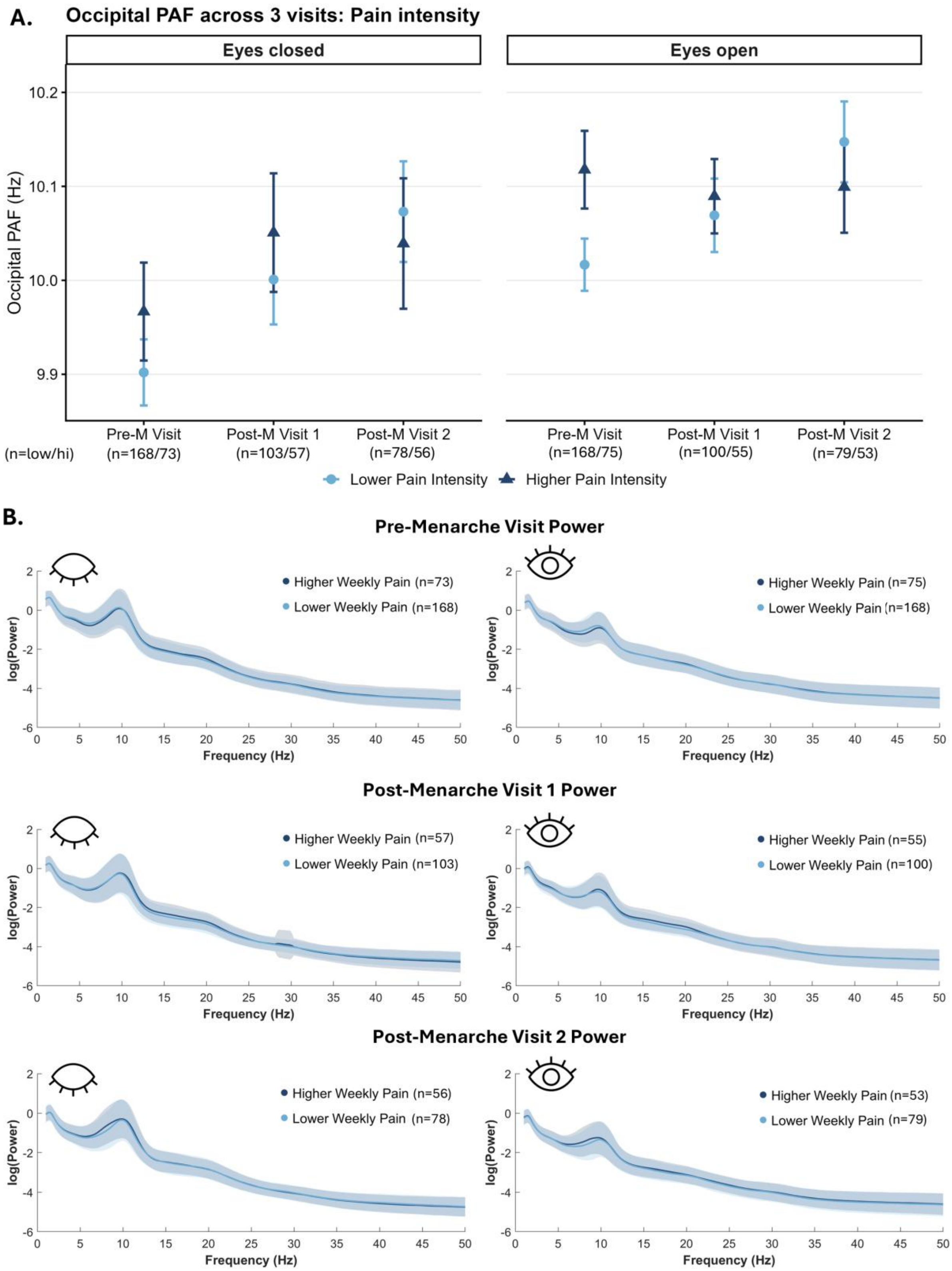

**Supplementary Figure 3. Influence of weekly pain intensity on resting-state EEG.** There were no significant differences in eyes-open or closed resting EEG spectral power or occipital peak alpha frequency (PAF) between adolescents who reported higher weekly pain intensity (≥ 3/10) vs. lower weekly pain intensity (≤ 2/10) at the pre-menarche or post-menarche study visits. PAF values are displayed as group means ± SEM. Pre-M Visit = Pre-menarche visit, Post-M1 visit = Post-menarche visit 1, Post-M2 Visit = Post-menarche visit 2.

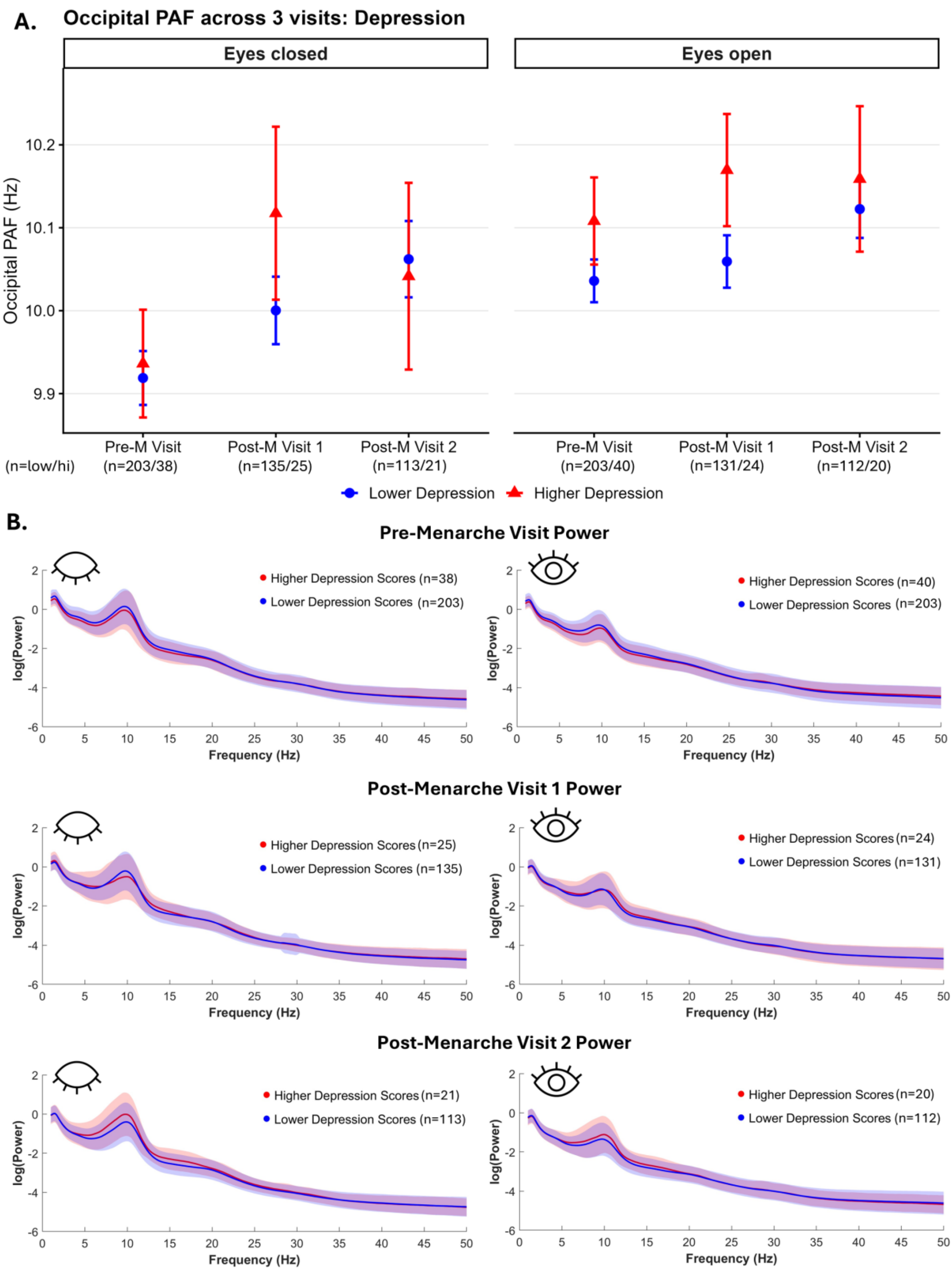

**Supplementary Figure 4. Influence of depression scores on resting-state EEG.** Adolescents were organized at each study visit into higher (top 15% of scores) and lower depression score groups to investigate the effect of depressed mood symptoms on resting EEG features shown to be influenced by menstrual pain status. We did not see differences in power or occipital peak alpha frequency (PAF) between the higher and lower depression score groups at any study visit. PAF values are displayed as group means ± SEM. Pre-M Visit = Pre-menarche visit, Post-M1 visit = Post-menarche visit 1, Post-M2 Visit = Post-menarche visit 2.

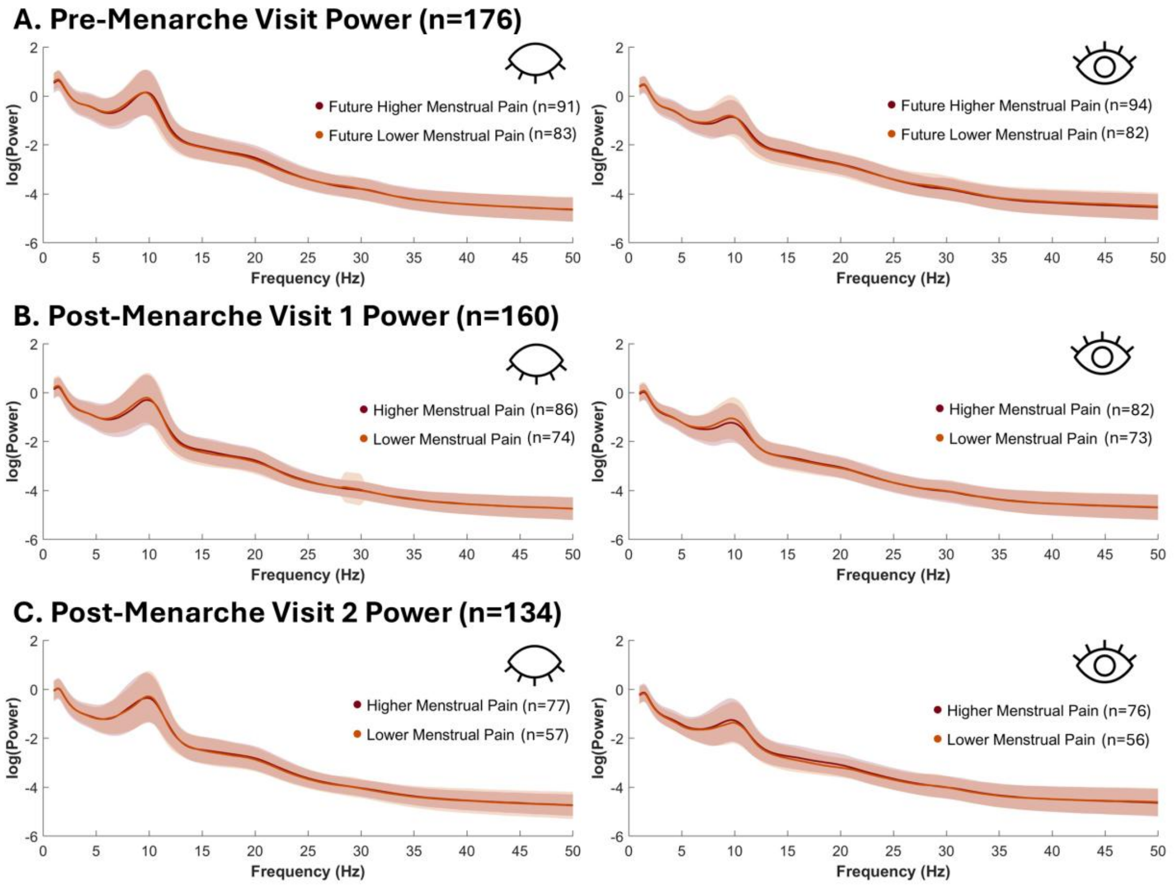

**Supplementary Figure 5. No differences in power between higher and lower menstrual pain.** Cross-sectional differences between adolescents with future (pre-menarche) and current (post-menarche visits 1 & 2) higher vs. lower menstrual pain were investigated for resting EEG power. No differences in eyes-open or closed power were seen at any study visit.

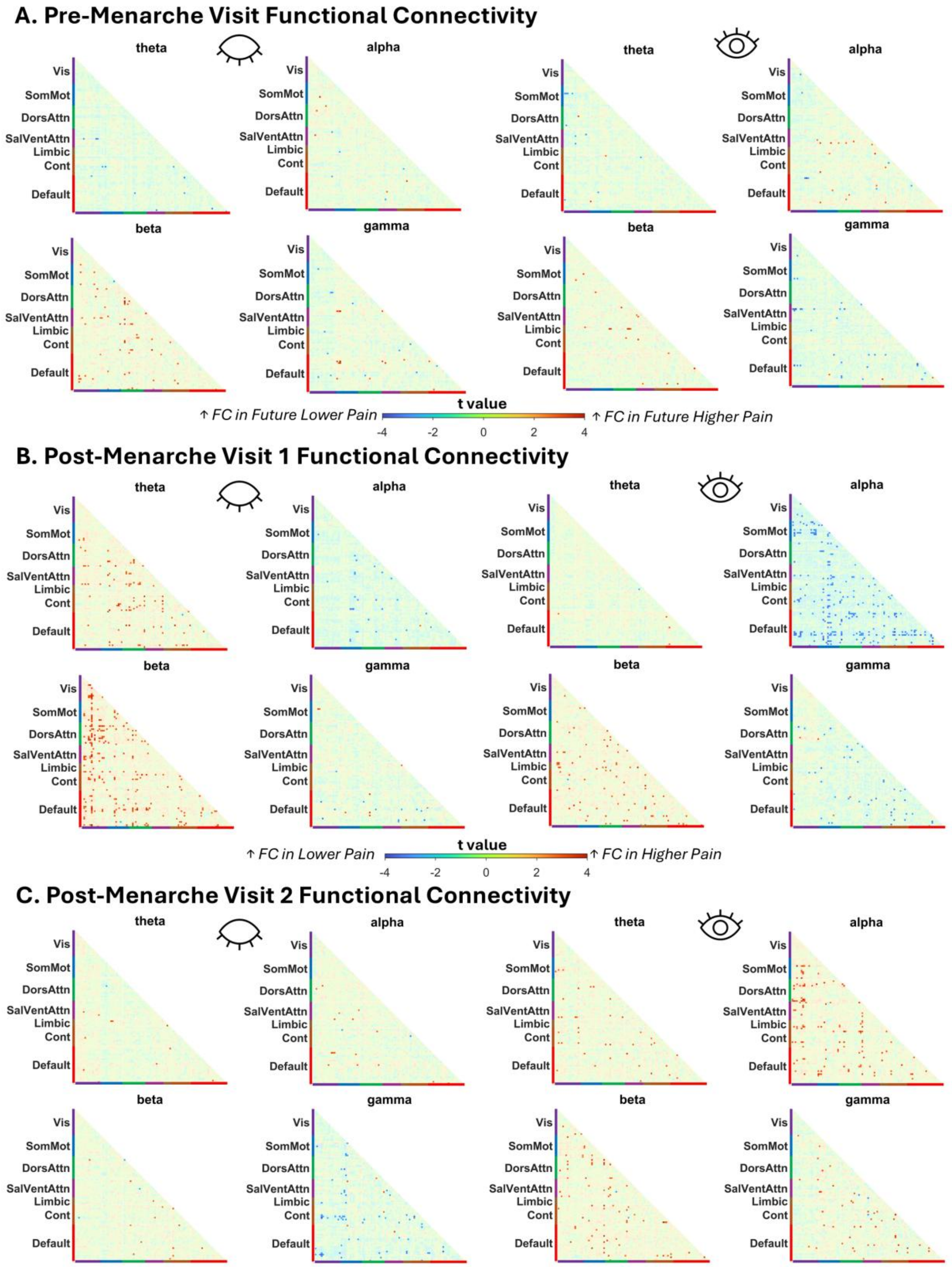

**Supplementary Figure 6. No significant differences in functional connectivity between higher and lower menstrual pain.** Cross-sectional resting functional connectivity differences between adolescents with future (pre-menarche) and current (post-menarche visits 1 & 2) higher vs. lower menstrual pain were investigated. Differences in functional connectivity at the pre-menarche visit (eyes-closed higher menstrual pain n= 91, lower menstrual pain n= 83; eyes-open higher menstrual pain n= 94, lower menstrual pain n= 82), post-menarche visit 1 (eyes-closed higher menstrual pain n= 86, lower menstrual pain n= 74; eyes-open higher menstrual pain n= 82, lower menstrual pain n= 73), and post-menarche visit 2 (eyes-closed higher menstrual pain n= 77, lower menstrual pain n= 57; eyes-open higher menstrual pain n= 76, lower menstrual pain n= 56) did not survive FDR correction for multiple comparisons. The connectivity matrices presented here are uncorrected, thresholded at $p \leq 0.01$ for illustrative purposes. FC = functional connectivity

### Supplementary Tables

Supplementary Tables 1 & 2 are found below. Supplementary Tables 3 – 9, which detail the significant ROI pairs for each frequency band in each functional connectivity analysis, are available as separate tabs in a combined excel spreadsheet due to their considerable size.

**Supplementary Table 1 Schaefer atlas ROI coordinates with Talairach atlas labels**

| ROI Label | ROI Name | R | A | S | Talairach |
|---|---|---|---|---|---|
| 1 | 7Networks_LH_Vis_1 | -26 | -34 | -17 | Left Fusiform Cortex |
| 2 | 7Networks_LH_Vis_2 | -26 | -77 | -14 | Left Visual Association Cortex |
| 3 | 7Networks_LH_Vis_3 | -17 | -60 | -7 | Left Visual Association Cortex |
| 4 | 7Networks_LH_Vis_4 | -27 | -95 | -4 | Left Secondary Visual Cortex |
| 5 | 7Networks_LH_Vis_5 | -5 | -92 | -2 | Left Secondary Visual Cortex |
| 6 | 7Networks_LH_Vis_6 | -12 | -67 | 7 | Left Primary Visual Cortex |
| 7 | 7Networks_LH_Vis_7 | -47 | -71 | 11 | Left Visual Association Cortex |
| 8 | 7Networks_LH_Vis_8 | -25 | -88 | 20 | Left Visual Association Cortex |
| 9 | 7Networks_LH_Vis_9 | -6 | -82 | 26 | Left Visual Association Cortex |
| 10 | 7Networks_LH_SomMot_1 | -53 | -23 | 8 | Left Supramarginal Gyrus |
| 11 | 7Networks_LH_SomMot_2 | -37 | -21 | 16 | Left Primary Sensory Cortex |
| 12 | 7Networks_LH_SomMot_3 | -54 | -12 | 13 | Left Primary Sensory Cortex |
| 13 | 7Networks_LH_SomMot_4 | -55 | -8 | 34 | Left PreMotor & Supplementary Motor Area |
| 14 | 7Networks_LH_SomMot_5 | -39 | -23 | 59 | Left Primary Motor Cortex |
| 15 | 7Networks_LH_SomMot_6 | -6 | -29 | 70 | Left Primary Motor Cortex |
| 16 | 7Networks_LH_DorsAttn_Post_1 | -47 | -58 | -13 | Left Fusiform Cortex |
| 17 | 7Networks_LH_DorsAttn_Post_2 | -57 | -25 | 39 | Left Supramarginal Gyrus |
| 18 | 7Networks_LH_DorsAttn_Post_3 | -24 | -68 | 49 | Left Visual Motor Cortex |
| 19 | 7Networks_LH_DorsAttn_Post_4 | -42 | -34 | 48 | Left Primary Sensory Cortex |
| 20 | 7Networks_LH_DorsAttn_Post_5 | -6 | -60 | 56 | Left Visual Motor Cortex |
| 21 | 7Networks_LH_DorsAttn_Post_6 | -22 | -51 | 66 | Left Visual Motor Cortex |
| 22 | 7Networks_LH_DorsAttn_PrCv_1 | -48 | 6 | 28 | Left PreMotor & Supplementary Motor Area |
| 23 | 7Networks_LH_DorsAttn_FEF_1 | -26 | -3 | 59 | Left PreMotor & Supplementary Motor Area |
| 24 | 7Networks_LH_SalVentAttn_ParOper_1 | -59 | -38 | 29 | Left Supramarginal Gyrus |
| 25 | 7Networks_LH_SalVentAttn_FrOperIns_1 | -41 | -1 | -7 | Left Insula |
| 26 | 7Networks_LH_SalVentAttn_FrOperIns_2 | -38 | 12 | 6 | Left Insula |
| 27 | 7Networks_LH_SalVentAttn_PFCl_1 | -30 | 44 | 30 | Left Anterior Prefrontal Cortex |
| 28 | 7Networks_LH_SalVentAttn_Med_1 | -5 | 20 | 34 | Left Dorsal Anterior Cingulate Cortex |
| 29 | 7Networks_LH_SalVentAttn_Med_2 | -11 | -34 | 45 | Left Dorsal Posterior Cingulate Cortex |
| 30 | 7Networks_LH_SalVentAttn_Med_3 | -6 | 4 | 62 | Left PreMotor & Supplementary Motor Area |
| 31 | 7Networks_LH_Limbic_OFC_1 | -14 | 32 | -20 | Left Orbitofrontal Cortex |
| 32 | 7Networks_LH_Limbic_TempPole_1 | -32 | 2 | -37 | Left Temporal Pole |
| 33 | 7Networks_LH_Limbic_TempPole_2 | -57 | -33 | -21 | Left Inferior Temporal Gyrus |
| 34 | 7Networks_LH_Cont_Par_1 | -37 | -53 | 46 | Left Angular Gyrus |
| 35 | 7Networks_LH_Cont_PFCl_1 | -43 | 33 | 21 | Left Dorsolateral Prefrontal Cortex |
| 36 | 7Networks_LH_Cont_pCun_1 | -9 | -73 | 38 | Left Visual Motor Cortex |
| 37 | 7Networks_LH_Cont_Cing_1 | -4 | -26 | 33 | Left Ventral Posterior Cingulate Cortex |
| 38 | 7Networks_LH_Default_Temp_1 | -55 | -4 | -20 | Left Medial Temporal Gyrus |
| 39 | 7Networks_LH_Default_Temp_2 | -58 | -32 | -1 | Left Medial Temporal Gyrus |
| 40 | 7Networks_LH_Default_Par_1 | -57 | -50 | 12 | Left Angular Gyrus |
| 41 | 7Networks_LH_Default_Par_2 | -48 | -63 | 35 | Left Angular Gyrus |
| 42 | 7Networks_LH_Default_PFC_1 | -35 | 21 | -11 | Left Pars Orbitalis |
| 43 | 7Networks_LH_Default_PFC_2 | -47 | 33 | -3 | Left Pars Orbitalis |
| 44 | 7Networks_LH_Default_PFC_3 | -6 | 47 | 0 | Left Dorsal Posterior Cingulate Cortex |
| 45 | 7Networks_LH_Default_PFC_4 | -24 | 61 | -1 | Left Anterior Prefrontal Cortex |
| 46 | 7Networks_LH_Default_PFC_5 | -9 | 48 | 41 | Left Dorsolateral Prefrontal Cortex |
| 47 | 7Networks_LH_Default_PFC_6 | -41 | 14 | 48 | Left PreMotor & Supplementary Motor Area |

| | | | | | |
|---|---|---|---|---|---|
| 48 | 7Networks_LH_Default_PFC_7 | -25 | 20 | 51 | Left Frontal Eye Fields |
| 49 | 7Networks_LH_Default_pCunPCC_1 | -11 | -56 | 13 | Left Ventral Posterior Cingulate Cortex |
| 50 | 7Networks_LH_Default_pCunPCC_2 | -6 | -53 | 33 | Left Ventral Posterior Cingulate Cortex |
| 51 | 7Networks_RH_Vis_1 | 32 | -31 | -22 | Right Fusiform Cortex |
| 52 | 7Networks_RH_Vis_2 | 27 | -66 | -12 | Right Fusiform Cortex |
| 53 | 7Networks_RH_Vis_3 | 49 | -60 | -11 | Right Fusiform Cortex |
| 54 | 7Networks_RH_Vis_4 | 22 | -96 | -5 | Right Secondary Visual Cortex |
| 55 | 7Networks_RH_Vis_5 | 8 | -76 | 5 | Right Primary Visual Cortex |
| 56 | 7Networks_RH_Vis_6 | 17 | -57 | 5 | Right Secondary Visual Cortex |
| 57 | 7Networks_RH_Vis_7 | 36 | -82 | 16 | Right Visual Association Cortex |
| 58 | 7Networks_RH_Vis_8 | 13 | -86 | 29 | Right Visual Association Cortex |
| 59 | 7Networks_RH_SomMot_1 | 53 | -16 | 7 | Right Primary Auditory Cortex |
| 60 | 7Networks_RH_SomMot_2 | 40 | -15 | 15 | Right Insula |
| 61 | 7Networks_RH_SomMot_3 | 57 | -4 | 11 | Right Primary Motor Cortex |
| 62 | 7Networks_RH_SomMot_4 | 58 | -5 | 31 | Right Primary Motor Cortex |
| 63 | 7Networks_RH_SomMot_5 | 47 | -11 | 48 | Right PreMotor & Supplementary Motor Area |
| 64 | 7Networks_RH_SomMot_6 | 41 | -22 | 60 | Right Primary Motor Cortex |
| 65 | 7Networks_RH_SomMot_7 | 30 | -37 | 64 | Right Primary Sensory Cortex |
| 66 | 7Networks_RH_SomMot_8 | 6 | -26 | 70 | Right Primary Motor Cortex |
| 67 | 7Networks_RH_DorsAttn_Post_1 | 50 | -62 | 16 | Right Visual Association Cortex |
| 68 | 7Networks_RH_DorsAttn_Post_2 | 50 | -24 | 42 | Right Primary Sensory Cortex |
| 69 | 7Networks_RH_DorsAttn_Post_3 | 38 | -45 | 49 | Right Visual Motor Cortex |
| 70 | 7Networks_RH_DorsAttn_Post_4 | 27 | -67 | 51 | Right Visual Motor Cortex |
| 71 | 7Networks_RH_DorsAttn_Post_5 | 14 | -52 | 66 | Right Visual Motor Cortex |
| 72 | 7Networks_RH_DorsAttn_PrCv_1 | 49 | 10 | 27 | Right Broca's Area/Operculum |
| 73 | 7Networks_RH_DorsAttn_FEF_1 | 28 | -3 | 59 | Right PreMotor & Supplementary Motor Area |
| 74 | 7Networks_RH_SalVentAttn_TempOccPar_1 | 58 | -42 | 13 | Right Superior Temporal Gyrus |
| 75 | 7Networks_RH_SalVentAttn_TempOccPar_2 | 61 | -26 | 27 | Right Supramarginal Gyrus |
| 76 | 7Networks_RH_SalVentAttn_FrOperIns_1 | 40 | 8 | 1 | Right Insula |
| 77 | 7Networks_RH_SalVentAttn_Med_1 | 11 | -31 | 45 | Right Dorsal Posterior Cingulate Cortex |
| 78 | 7Networks_RH_SalVentAttn_Med_2 | 7 | 6 | 52 | Right PreMotor & Supplementary Motor Area |
| 79 | 7Networks_RH_Limbic_OFC_1 | 12 | 35 | -20 | Right Orbitofrontal Cortex |
| 80 | 7Networks_RH_Limbic_TempPole_1 | 38 | 1 | -38 | Right Inferior Temporal Gyrus |
| 81 | 7Networks_RH_Cont_Par_1 | 57 | -39 | 44 | Right Supramarginal Gyrus |
| 82 | 7Networks_RH_Cont_Par_2 | 45 | -63 | 46 | Right Angular Gyrus |
| 83 | 7Networks_RH_Cont_PFCl_1 | 30 | 58 | -3 | Right Anterior Prefrontal Cortex |
| 84 | 7Networks_RH_Cont_PFCl_2 | 45 | 39 | 15 | Right Dorsolateral Prefrontal Cortex |
| 85 | 7Networks_RH_Cont_PFCl_3 | 32 | 46 | 29 | Right Dorsolateral Prefrontal Cortex |
| 86 | 7Networks_RH_Cont_PFCl_4 | 43 | 16 | 45 | Right Frontal Eye Fields |
| 87 | 7Networks_RH_Cont_Cing_1 | 5 | -27 | 33 | Right Ventral Posterior Cingulate Cortex |
| 88 | 7Networks_RH_Cont_PFCmp_1 | 6 | 28 | 30 | Right Frontal Eye Fields |
| 89 | 7Networks_RH_Cont_pCun_1 | 9 | -66 | 43 | Right Visual Motor Cortex |
| 90 | 7Networks_RH_Default_Par_1 | 55 | -51 | 31 | Right Angular Gyrus |
| 91 | 7Networks_RH_Default_Temp_1 | 62 | -23 | -19 | Right Medial Temporal Gyrus |
| 92 | 7Networks_RH_Default_Temp_2 | 51 | 7 | -18 | Right Temporal Pole |
| 93 | 7Networks_RH_Default_Temp_3 | 57 | -26 | -2 | Right Superior Temporal Gyrus |
| 94 | 7Networks_RH_Default_PFCv_1 | 35 | 26 | -15 | Right Pars Orbitalis |
| 95 | 7Networks_RH_Default_PFCv_2 | 51 | 28 | 0 | Right Broca's Area/ Pars Triangularis |
| 96 | 7Networks_RH_Default_PFCdPFCm_1 | 7 | 48 | 1 | Right Anterior Prefrontal Cortex |
| 97 | 7Networks_RH_Default_PFCdPFCm_2 | 11 | 50 | 39 | Right Dorsolateral Prefrontal Cortex |

| | | | | | |
|---|---|---|---|---|---|
| 98 | 7Networks_RH_Default_PFCdPFCm_3 | 26 | 24 | 50 | Right Frontal Eye Fields |
| 99 | 7Networks_RH_Default_pCunPCC_1 | 12 | -54 | 14 | Right Ventral Posterior Cingulate Cortex |
| 100 | 7Networks_RH_Default_pCunPCC_2 | 7 | -52 | 31 | Right Ventral Posterior Cingulate Cortex |

Centroids for the 100 7-network Schaefer atlas ROIs with corresponding anatomical labels derived from an MNI to Talairach converter.

**Supplementary Table 2 Results of LMMs evaluating menstrual group differences in global and occipital PAF**

| | | | | | | |
|---|---|---|---|---|---|---|
| **A. Full-cohort LMM (eyes-closed global PAF)** | | | | | | |
| all participants | Pre-menarche (n) | Post-menarche visit 1 (n) | Post-menarche visit 2 (n) | | | |
| | 174 | 160 | 134 | | | |
| **Fixed Effects** | | | | | | |
| term | **estimate (Hz)** | **SE** | **95% CI** | **t** | **df** | **p** |
| pre-menarche visit PAF (Hz) | 9.920 | 0.028 | [9.864, 9.976] | 349.77 | 194.6 | 2.74E-274 |
| PAF change per visit (Hz/visit) | 0.056 | 0.009 | [0.037, 0.074] | 5.95 | 152.5 | 1.74E-08 |
| **B. Full-cohort LMM (eyes-open global PAF)** | | | | | | |
| all participants | Pre-menarche (n) | Post-menarche visit 1 (n) | Post-menarche visit 2 (n) | | | |
| | 176 | 155 | 132 | | | |
| **Fixed Effects** | | | | | | |
| term | **estimate (Hz)** | **SE** | **95% CI** | **t** | **df** | **p** |
| pre-menarche visit PAF (Hz) | 10.022 | 0.023 | [9.977, 10.067] | 437.73 | 197.5 | 5.31E-297 |
| PAF change per visit (Hz/visit) | 0.028 | 0.008 | [0.012, 0.045] | 3.38 | 157.8 | 9.13E-04 |
| **C. Group-comparison LMM (eyes-closed global PAF)** | | | | | | |
| | Pre-menarche (n) | Post-menarche visit 1 (n) | Post-menarche visit 2 (n) | | | |
| Higher menstrual pain | 91 | 86 | 77 | | | |
| Lower menstrual pain | 83 | 74 | 57 | | | |
| **Fixed Effects** | | | | | | |
| term | **estimate (Hz)** | **SE** | **95% CI** | **t** | **df** | **p** |
| Lower menstrual pain (LMP) pre-menarche visit PAF (Hz) | 9.876 | 0.034 | [9.81, 9.943] | 293.68 | 250.8 | 0 |
| LMP PAF change per visit (Hz/visit) | 0.072 | 0.014 | [0.044, 0.101] | 5.02 | 184.4 | 0 |
| Higher menstrual pain (HMP) - LMP PAF at pre-menarche visit (Hz) | 0.081 | 0.035 | [0.013, 0.150] | 2.33 | 176.0 | 0.021 |
| HMP - LMP PAF change per visit (Hz/visit) | -0.030 | 0.020 | [-0.069, 0.009] | -1.53 | 201.9 | 0.127 |
| **Group Contrasts (By Visit)** | | | | | | |
| Visit | **Higher - Lower menstrual pain** | **95% CI** | **t** | **df** | **p** | |
| Pre-menarche | 0.081 | [-0.004, 0.167] | 2.31 | 172.9 | 0.066 | |
| Post-menarche Visit 1 | 0.051 | [-0.016, 0.118] | 1.84 | 299.7 | 0.133 | |
| Post-menarche Visit 2 | 0.021 | [-0.059, 0.101] | 0.63 | 293.3 | 0.529 | |
| **D. Group-comparison LMM (eyes-open global PAF)** | | | | | | |
| | Pre-menarche (n) | Post-menarche visit 1 (n) | Post-menarche visit 2 (n) | | | |
| Higher menstrual pain | 94 | 82 | 76 | | | |
| Lower menstrual pain | 82 | 73 | 56 | | | |

**Fixed Effects**

| term | estimate (Hz) | SE | 95% CI | t | df | p |
|---|---|---|---|---|---|---|
| Lower menstrual pain (LMP) pre-menarche visit PAF (Hz) | 9.989 | 0.028 | [9.933, 10.045] | 352.00 | 246.4 | 0 |
| LMP PAF change per visit (Hz/visit) | 0.042 | 0.013 | [0.016, 0.067] | 3.18 | 192.3 | 0.002 |
| Higher menstrual pain (HMP) - LMP PAF at pre-menarche visit (Hz) | 0.061 | 0.032 | [-0.002, 0.124] | 1.91 | 183.2 | 0.058 |
| HMP - LMP PAF change per visit (Hz/visit) | -0.024 | 0.018 | [-0.059, 0.011] | -1.33 | 203.8 | 0.185 |

**Group Contrasts (By Visit)**

| Visit | Higher - Lower menstrual pain | 95% CI | t | df | p |
|---|---|---|---|---|---|
| Pre-menarche | 0.061 | [-0.017, 0.139] | 1.89 | 186.8 | 0.18 |
| Post-menarche Visit 1 | 0.037 | [-0.023, 0.098] | 1.49 | 344.3 | 0.272 |
| Post-menarche Visit 2 | 0.014 | [-0.057, 0.084] | 0.47 | 303.6 | 0.639 |

**E. Group-comparison LMM (eyes-closed occipital PAF)**

| | Pre-menarche (n) | Post-menarche visit 1 (n) | Post-menarche visit 2 (n) |
|---|---|---|---|
| Higher menstrual pain | 91 | 86 | 77 |
| Lower menstrual pain | 83 | 74 | 57 |

**Fixed Effects**

| term | estimate (Hz) | SE | 95% CI | t | df | p |
|---|---|---|---|---|---|---|
| Lower menstrual pain (LMP) pre-menarche visit PAF (Hz) | 9.866 | 0.039 | [9.789, 9.943] | 253.28 | 248.4 | 0 |
| LMP PAF change per visit (Hz/visit) | 0.101 | 0.017 | [0.068, 0.134] | 5.97 | 181.9 | 0 |
| Higher menstrual pain (HMP) - LMP PAF at pre-menarche visit (Hz) | 0.110 | 0.041 | [0.030, 0.191] | 2.70 | 174.5 | 0.008 |
| HMP - LMP PAF change per visit (Hz/visit) | -0.031 | 0.023 | [-0.077, 0.015] | -1.34 | 199.7 | 0.182 |

**Group Contrasts (By Visit)**

| Visit | Higher - Lower menstrual pain | 95% CI | t | df | p |
|---|---|---|---|---|---|
| Pre-menarche | 0.11 | [0.011, 0.210] | 2.67 | 172.1 | 0.025 |
| Post-menarche Visit 1 | 0.079 | [0.0004, 0.158] | 2.42 | 298.6 | 0.032 |
| Post-menarche Visit 2 | 0.048 | [-0.047, 0.142] | 1.22 | 290.1 | 0.224 |

**F. Group-comparison LMM (eyes-open occipital PAF)**

| | Pre-menarche (n) | Post-menarche visit 1 (n) | Post-menarche visit 2 (n) |
|---|---|---|---|
| Higher menstrual pain | 94 | 82 | 76 |
| Lower menstrual pain | 82 | 73 | 56 |

**Fixed Effects**

| term | estimate (Hz) | SE | 95% CI | t | df | p |
|---|---|---|---|---|---|---|
| Lower menstrual pain (LMP) pre-menarche visit PAF (Hz) | 9.989 | 0.032 | [9.925, 10.052] | 310.32 | 241.3 | 0 |

| | | | | | | |
|---|---|---|---|---|---|---|
| LMP PAF change per visit (Hz/visit) | 0.066 | 0.015 | [0.037, 0.096] | 4.46 | 181.7 | 0 |
| Higher menstrual pain (HMP) - LMP PAF at pre-menarche visit (Hz) | 0.090 | 0.036 | [0.019, 0.161] | 2.48 | 173.9 | 0.014 |
| HMP - LMP PAF change per visit (Hz/visit) | -0.029 | 0.020 | [-0.069, 0.011] | -1.43 | 192.9 | 0.153 |
| **Group Contrasts (By Visit)** | | | | | | |
| Visit | **Higher - Lower menstrual pain** | **95% CI** | **t** | **df** | **p** | |
| Pre-menarche | 0.09 | [0.002, 0.178] | 2.46 | 184.4 | 0.044 | |
| Post-menarche Visit 1 | 0.061 | [-0.008, 0.130] | 2.13 | 335.6 | 0.069 | |
| Post-menarche Visit 2 | 0.032 | [-0.050, 0.113] | 0.94 | 295.4 | 0.348 | |

LMM = linear mixed model, PAF = peak alpha frequency

See supplementary excel file for Supplementary Tables 3 to 9.

**Supplementary Table 3 Functional connectivity results for all participants (n=147) from the pre-menarche to post-menarche visit 1**

**Eyes-Open Theta**

| **ROI1 Labels (Schaefer; Talairach)** | **X** | **Y** | **Z** | **ROI2 Labels (Schaefer; Talairach)** | **X** | **Y** | **Z** | **t** | **q** |
|---|---|---|---|---|---|---|---|---|---|
| LH_Limbic_TempPole_2; Left Inferior Temporal Gyrus | -57 | -33 | -21 | RH_Default_Par_1; Right Angular Gyrus | 55 | -51 | 31 | 5.66 | 0.0008 |
| LH_Vis_1; Left Fusiform Cortex | -26 | -34 | -17 | RH_DorsAttn_Post_3; Right Visual Motor Cortex | 38 | -45 | 49 | 5.43 | 0.001 |
| RH_DorsAttn_Post_4; Right Visual Motor Cortex | 27 | -67 | 51 | LH_Limbic_TempPole_2; Left Inferior Temporal Gyrus | -57 | -33 | -21 | 5.01 | 0.005 |
| LH_Limbic_TempPole_2; Left Inferior Temporal Gyrus | -57 | -33 | -21 | RH_Cont_Par_2; Right Angular Gyrus | 45 | -63 | 46 | 4.92 | 0.006 |
| LH_Vis_1; Left Fusiform Cortex | -26 | -34 | -17 | RH_DorsAttn_Post_4; Right Visual Motor Cortex | 27 | -67 | 51 | 4.75 | 0.010 |
| RH_DorsAttn_Post_3; Right Visual Motor Cortex | 38 | -45 | 49 | LH_Limbic_TempPole_2; Left Inferior Temporal Gyrus | -57 | -33 | -21 | 4.65 | 0.011 |
| RH_DorsAttn_Post_4; Right Visual Motor Cortex | 27 | -67 | 51 | LH_Default_Temp_1; Left Medial Temporal Gyrus | -55 | -4 | -20 | 4.65 | 0.011 |
| RH_Vis_2; Right Fusiform Cortex | 27 | -66 | -12 | LH_Cont_Par_1; Left Angular Gyrus | -37 | -53 | 46 | 4.36 | 0.022 |
| LH_DorsAttn_Post_1; Left Fusiform Cortex | -47 | -58 | -13 | LH_Default_Temp_1; Left Medial Temporal Gyrus | -55 | -4 | -20 | 4.36 | 0.022 |
| RH_SalVentAttn_TempOccPar_2; Right Supramarginal Gyrus | 61 | -26 | 27 | LH_Limbic_TempPole_2; Left Inferior Temporal Gyrus | -57 | -33 | -21 | 4.35 | 0.022 |
| LH_Vis_4; Left Secondary Visual Cortex | -27 | -95 | -4 | LH_Limbic_TempPole_2; Left Inferior Temporal Gyrus | -57 | -33 | -21 | 4.35 | 0.022 |
| RH_Vis_6; Right Secondary Visual Cortex | 17 | -57 | 5 | LH_Limbic_TempPole_2; Left Inferior Temporal Gyrus | -57 | -33 | -21 | 4.29 | 0.022 |
| LH_Vis_3; Left Visual Association Cortex | -17 | -60 | -7 | LH_Cont_PFCl_1; Left Dorsolateral Prefrontal Cortex | -43 | 33 | 21 | 4.28 | 0.022 |
| RH_SomMot_1; Right Primary Auditory Cortex | 53 | -16 | 7 | RH_Cont_Par_2; Right Angular Gyrus | 45 | -63 | 46 | 4.27 | 0.022 |
| LH_Vis_8; Left Visual Association Cortex | -25 | -88 | 20 | LH_Limbic_TempPole_2; Left Inferior Temporal Gyrus | -57 | -33 | -21 | 4.25 | 0.022 |
| LH_Limbic_TempPole_2; Left Inferior Temporal Gyrus | -57 | -33 | -21 | RH_Cont_Par_1; Right Supramarginal Gyrus | 57 | -39 | 44 | 4.25 | 0.022 |
| LH_SalVentAttn_ParOper_1; Left Supramarginal Gyrus | -59 | -38 | 29 | LH_Default_PFC_2; Left Pars Orbitalis | -47 | 33 | -3 | 4.24 | 0.022 |
| LH_DorsAttn_FEF_1; Left PreMotor & Supplementary Motor Area | -26 | -3 | 59 | LH_Cont_pCun_1; Left Visual Motor Cortex | -9 | -73 | 38 | 4.23 | 0.022 |
| LH_Limbic_TempPole_2; Left Inferior Temporal Gyrus | -57 | -33 | -21 | RH_Default_PFCdPFCm_2; Right Dorsolateral Prefrontal Cortex | 11 | 50 | 39 | 4.22 | 0.022 |
| RH_Vis_8; Right Visual Association Cortex | 13 | -86 | 29 | LH_Limbic_TempPole_2; Left Inferior Temporal Gyrus | -57 | -33 | -21 | 4.19 | 0.024 |
| LH_Vis_9; Left Visual Association Cortex | -6 | -82 | 26 | LH_SalVentAttn_Med_3; Left PreMotor & Supplementary Motor Area | -6 | 4 | 62 | 4.18 | 0.024 |
| RH_Vis_5; Right Primary Visual Cortex | 8 | -76 | 5 | LH_Limbic_TempPole_2; Left Inferior Temporal Gyrus | -57 | -33 | -21 | 4.14 | 0.026 |
| LH_Default_Temp_1; Left Medial Temporal Gyrus | -55 | -4 | -20 | RH_Default_Par_1; Right Angular Gyrus | 55 | -51 | 31 | 4.13 | 0.026 |
| RH_Vis_3; Right Fusiform Cortex | 49 | -60 | -11 | LH_DorsAttn_FEF_1; Left PreMotor & Supplementary Motor Area | -26 | -3 | 59 | 4.12 | 0.026 |
| LH_Limbic_TempPole_2; Left Inferior Temporal Gyrus | -57 | -33 | -21 | LH_Default_Temp_1; Left Medial Temporal Gyrus | -55 | -4 | -20 | 4.11 | 0.026 |
| RH_Vis_2; Right Fusiform Cortex | 27 | -66 | -12 | LH_DorsAttn_FEF_1; Left PreMotor & Supplementary Motor Area | -26 | -3 | 59 | 4.10 | 0.026 |
| LH_DorsAttn_Post_3; Left Visual Motor Cortex | -24 | -68 | 49 | LH_Default_PFC_2; Left Pars Orbitalis | -47 | 33 | -3 | 4.06 | 0.030 |
| LH_SalVentAttn_Med_3; Left PreMotor & Supplementary Motor Area | -6 | 4 | 62 | LH_Default_PFC_1; Left Pars Orbitalis | -35 | 21 | -11 | 4.05 | 0.030 |
| RH_Vis_2; Right Fusiform Cortex | 27 | -66 | -12 | LH_Limbic_TempPole_2; Left Inferior Temporal Gyrus | -57 | -33 | -21 | 4.03 | 0.031 |

| | | | | | | | | | |
|---|---|---|---|---|---|---|---|---|---|
| LH_SalVentAttn_Med_3; Left PreMotor & Supplementary Motor Area | -6 | 4 | 62 | LH_Limbic_TempPole_1; Left Temporal Pole | -32 | 2 | -37 | 3.99 | 0.034 |
| RH_Cont_Par_2; Right Angular Gyrus | 45 | -63 | 46 | LH_Default_Temp_1; Left Medial Temporal Gyrus | -55 | -4 | -20 | 3.98 | 0.034 |
| LH_Vis_2; Left Visual Association Cortex | -26 | -77 | -14 | RH_Default_Par_1; Right Angular Gyrus | 55 | -51 | 31 | 3.97 | 0.034 |
| LH_SalVentAttn_Med_3; Left PreMotor & Supplementary Motor Area | -6 | 4 | 62 | LH_Cont_PFCl_1; Left Dorsolateral Prefrontal Cortex | -43 | 33 | 21 | 3.97 | 0.034 |
| RH_Cont_PFCl_1; Right Anterior Prefrontal Cortex | 30 | 58 | -3 | RH_Default_pCunPCC_2; Right Ventral Posterior Cingulate Cortex | 7 | -52 | 31 | 3.96 | 0.035 |
| LH_Vis_3; Left Visual Association Cortex | -17 | -60 | -7 | LH_SomMot_5; Left Primary Motor Cortex | -39 | -23 | 59 | 3.94 | 0.036 |
| RH_SomMot_8; Right Primary Motor Cortex | 6 | -26 | 70 | LH_SalVentAttn_Med_3; Left PreMotor & Supplementary Motor Area | -6 | 4 | 62 | 3.91 | 0.038 |
| RH_Vis_2; Right Fusiform Cortex | 27 | -66 | -12 | RH_Default_Par_1; Right Angular Gyrus | 55 | -51 | 31 | 3.90 | 0.038 |
| LH_Vis_3; Left Visual Association Cortex | -17 | -60 | -7 | LH_Default_PFC_2; Left Pars Orbitalis | -47 | 33 | -3 | 3.90 | 0.038 |
| LH_DorsAttn_Post_1; Left Fusiform Cortex | -47 | -58 | -13 | RH_DorsAttn_Post_3; Right Visual Motor Cortex | 38 | -45 | 49 | 3.89 | 0.038 |
| RH_Vis_5; Right Primary Visual Cortex | 8 | -76 | 5 | LH_Default_PFC_1; Left Pars Orbitalis | -35 | 21 | -11 | 3.89 | 0.038 |
| RH_Vis_7; Right Visual Association Cortex | 36 | -82 | 16 | LH_Limbic_TempPole_2; Left Inferior Temporal Gyrus | -57 | -33 | -21 | 3.88 | 0.038 |
| RH_DorsAttn_Post_1; Right Visual Association Cortex | 50 | -62 | 16 | LH_Limbic_TempPole_2; Left Inferior Temporal Gyrus | -57 | -33 | -21 | 3.87 | 0.038 |
| LH_Limbic_TempPole_2; Left Inferior Temporal Gyrus | -57 | -33 | -21 | LH_Default_PFC_1; Left Pars Orbitalis | -35 | 21 | -11 | 3.87 | 0.038 |
| LH_SalVentAttn_PFCl_1; Left Anterior Prefrontal Cortex | -30 | 44 | 30 | LH_SalVentAttn_Med_3; Left PreMotor & Supplementary Motor Area | -6 | 4 | 62 | 3.86 | 0.038 |
| RH_Vis_6; Right Secondary Visual Cortex | 17 | -57 | 5 | LH_DorsAttn_FEF_1; Left PreMotor & Supplementary Motor Area | -26 | -3 | 59 | 3.84 | 0.041 |
| LH_DorsAttn_Post_1; Left Fusiform Cortex | -47 | -58 | -13 | LH_Limbic_TempPole_2; Left Inferior Temporal Gyrus | -57 | -33 | -21 | 3.82 | 0.041 |
| LH_Vis_5; Left Secondary Visual Cortex | -5 | -92 | -2 | LH_Default_PFC_6; Left PreMotor & Supplementary Motor Area | -41 | 14 | 48 | 3.81 | 0.041 |
| RH_SomMot_7; Right Primary Sensory Cortex | 30 | -37 | 64 | RH_DorsAttn_Post_4; Right Visual Motor Cortex | 27 | -67 | 51 | 3.81 | 0.041 |
| LH_Vis_9; Left Visual Association Cortex | -6 | -82 | 26 | LH_DorsAttn_FEF_1; Left PreMotor & Supplementary Motor Area | -26 | -3 | 59 | 3.81 | 0.041 |
| LH_Vis_9; Left Visual Association Cortex | -6 | -82 | 26 | LH_Limbic_TempPole_2; Left Inferior Temporal Gyrus | -57 | -33 | -21 | 3.81 | 0.041 |
| RH_Cont_Cing_1; Right Ventral Posterior Cingulate Cortex | 5 | -27 | 33 | RH_Default_PFCv_1; Right Pars Orbitalis | 35 | 26 | -15 | 3.80 | 0.041 |
| LH_Vis_5; Left Secondary Visual Cortex | -5 | -92 | -2 | LH_Limbic_TempPole_2; Left Inferior Temporal Gyrus | -57 | -33 | -21 | 3.80 | 0.041 |
| RH_DorsAttn_Post_1; Right Visual Association Cortex | 50 | -62 | 16 | LH_Default_Temp_1; Left Medial Temporal Gyrus | -55 | -4 | -20 | 3.78 | 0.041 |
| LH_Vis_2; Left Visual Association Cortex | -26 | -77 | -14 | LH_Vis_5; Left Secondary Visual Cortex | -5 | -92 | -2 | 3.78 | 0.041 |
| LH_Vis_8; Left Visual Association Cortex | -25 | -88 | 20 | LH_DorsAttn_Post_1; Left Fusiform Cortex | -47 | -58 | -13 | 3.78 | 0.041 |
| LH_DorsAttn_Post_1; Left Fusiform Cortex | -47 | -58 | -13 | LH_Default_PFC_1; Left Pars Orbitalis | -35 | 21 | -11 | 3.78 | 0.041 |
| RH_Vis_2; Right Fusiform Cortex | 27 | -66 | -12 | RH_Cont_Par_1; Right Supramarginal Gyrus | 57 | -39 | 44 | 3.76 | 0.042 |
| LH_Vis_3; Left Visual Association Cortex | -17 | -60 | -7 | LH_DorsAttn_Post_4; Left Primary Sensory Cortex | -42 | -34 | 48 | 3.75 | 0.043 |
| LH_DorsAttn_Post_3; Left Visual Motor Cortex | -24 | -68 | 49 | LH_Default_Temp_1; Left Medial Temporal Gyrus | -55 | -4 | -20 | 3.75 | 0.043 |
| LH_SalVentAttn_PFCl_1; Left Anterior Prefrontal Cortex | -30 | 44 | 30 | LH_Limbic_TempPole_2; Left Inferior Temporal Gyrus | -57 | -33 | -21 | 3.74 | 0.043 |
| LH_DorsAttn_Post_3; Left Visual Motor Cortex | -24 | -68 | 49 | LH_Default_PFC_4; Left Anterior Prefrontal Cortex | -24 | 61 | -1 | 3.74 | 0.043 |

| | | | | | | | | | |
|---|---|---|---|---|---|---|---|---|---|
| RH_Vis_5; Right Primary Visual Cortex | 8 | -76 | 5 | LH_Default_PFC_6; Left PreMotor & Supplementary Motor Area | -41 | 14 | 48 | 3.74 | 0.043 |
| LH_Vis_4; Left Secondary Visual Cortex | -27 | -95 | -4 | LH_DorsAttn_Post_4; Left Primary Sensory Cortex | -42 | -34 | 48 | 3.72 | 0.044 |
| RH_Vis_3; Right Fusiform Cortex | 49 | -60 | -11 | LH_Default_PFC_1; Left Pars Orbitalis | -35 | 21 | -11 | 3.72 | 0.044 |
| LH_DorsAttn_Post_5; Left Visual Motor Cortex | -6 | -60 | 56 | LH_Default_PFC_4; Left Anterior Prefrontal Cortex | -24 | 61 | -1 | 3.72 | 0.044 |
| RH_SalVentAttn_Med_2; Right PreMotor & Supplementary Motor Area | 7 | 6 | 52 | RH_Default_PFCdPFCm_1; Right Anterior Prefrontal Cortex | 7 | 48 | 1 | 3.71 | 0.044 |
| LH_SalVentAttn_ParOper_1; Left Supramarginal Gyrus | -59 | -38 | 29 | LH_Default_PFC_1; Left Pars Orbitalis | -35 | 21 | -11 | 3.71 | 0.044 |
| LH_Vis_3; Left Visual Association Cortex | -17 | -60 | -7 | LH_DorsAttn_Post_6; Left Visual Motor Cortex | -22 | -51 | 66 | 3.70 | 0.044 |
| LH_Vis_2; Left Visual Association Cortex | -26 | -77 | -14 | RH_Cont_PFCl_1; Right Anterior Prefrontal Cortex | 30 | 58 | -3 | 3.70 | 0.044 |
| LH_Cont_PFCl_1; Left Dorsolateral Prefrontal Cortex | -43 | 33 | 21 | RH_Cont_PFCmp_1; Right Frontal Eye Fields | 6 | 28 | 30 | 3.68 | 0.046 |
| RH_Vis_2; Right Fusiform Cortex | 27 | -66 | -12 | LH_SalVentAttn_Med_3; Left PreMotor & Supplementary Motor Area | -6 | 4 | 62 | 3.66 | 0.048 |
| RH_Vis_8; Right Visual Association Cortex | 13 | -86 | 29 | LH_DorsAttn_FEF_1; Left PreMotor & Supplementary Motor Area | -26 | -3 | 59 | 3.66 | 0.048 |
| RH_Vis_8; Right Visual Association Cortex | 13 | -86 | 29 | LH_Default_PFC_1; Left Pars Orbitalis | -35 | 21 | -11 | 3.65 | 0.048 |
| LH_SalVentAttn_Med_1; Left Dorsal Anterior Cingulate Cortex | -5 | 20 | 34 | LH_Cont_PFCl_1; Left Dorsolateral Prefrontal Cortex | -43 | 33 | 21 | 3.65 | 0.048 |
| LH_DorsAttn_Post_1; Left Fusiform Cortex | -47 | -58 | -13 | RH_Cont_Par_2; Right Angular Gyrus | 45 | -63 | 46 | 3.65 | 0.048 |
| LH_DorsAttn_PrCv_1; Left PreMotor & Supplementary Motor Area | -48 | 6 | 28 | LH_Cont_Par_1; Left Angular Gyrus | -37 | -53 | 46 | 3.63 | 0.050 |
| RH_Vis_6; Right Secondary Visual Cortex | 17 | -57 | 5 | LH_SomMot_5; Left Primary Motor Cortex | -39 | -23 | 59 | 3.63 | 0.050 |
| **Eyes-Closed Beta** | | | | | | | | | |
| **ROI1 Labels (Schaefer; Talairach)** | **X** | **Y** | **Z** | **ROI2 Labels (Schaefer; Talairach)** | **X** | **Y** | **Z** | **t** | **q** |
| RH_SomMot_7; Right Primary Sensory Cortex | 30 | -37 | 64 | RH_SalVentAttn_Med_2; Right PreMotor & Supplementary Motor Area | 7 | 6 | 52 | -4.79 | 0.034 |
| LH_SomMot_5; Left Primary Motor Cortex | -39 | -23 | 59 | RH_SalVentAttn_Med_1; Right Dorsal Posterior Cingulate Cortex | 11 | -31 | 45 | -4.64 | 0.034 |
| LH_DorsAttn_Post_6; Left Visual Motor Cortex | -22 | -51 | 66 | RH_DorsAttn_FEF_1; Right PreMotor & Supplementary Motor Area | 28 | -3 | 59 | -4.57 | 0.034 |
| LH_SalVentAttn_Med_1; Left Dorsal Anterior Cingulate Cortex | -5 | 20 | 34 | RH_SalVentAttn_Med_1; Right Dorsal Posterior Cingulate Cortex | 11 | -31 | 45 | -4.49 | 0.036 |
| **Eyes-Closed Gamma** | | | | | | | | | |
| **ROI1 Labels (Schaefer; Talairach)** | **X** | **Y** | **Z** | **ROI2 Labels (Schaefer; Talairach)** | **X** | **Y** | **Z** | **t** | **q** |
| RH_SomMot_3; Right Primary Motor Cortex | 57 | -4 | 11 | RH_Default_Par_1; Right Angular Gyrus | 55 | -51 | 31 | 5.13 | 0.009 |
| RH_SomMot_3; Right Primary Motor Cortex | 57 | -4 | 11 | RH_SalVentAttn_TempOccPar_2; Right Supramarginal Gyrus | 61 | -26 | 27 | 4.67 | 0.022 |
| RH_SomMot_4; Right Primary Motor Cortex | 58 | -5 | 31 | RH_Default_Par_1; Right Angular Gyrus | 55 | -51 | 31 | 4.67 | 0.022 |

q ≤ 0.05 (FDR-corrected)

**Supplementary Table 4 Functional connectivity results for lower menstrual pain participants (n=69) from the pre-menarche to post-menarche visit 1**

| **Eyes-Closed Theta** | | | | | | | | | |
|---|---|---|---|---|---|---|---|---|---|
| **ROI1 Labels (Schaefer; Talairach)** | **X** | **Y** | **Z** | **ROI2 Labels (Schaefer; Talairach)** | **X** | **Y** | **Z** | **t** | **q** |
| LH_DorsAttn_FEF_1; Left PreMotor & Supplementary Motor Area | -26 | -3 | 59 | LH_Default_pCunPCC_1; Left Ventral Posterior Cingulate Cortex | -11 | -56 | 13 | 5.97 | 0.001 |
| LH_DorsAttn_FEF_1; Left PreMotor & Supplementary Motor Area | -26 | -3 | 59 | RH_Default_pCunPCC_1; Right Ventral Posterior Cingulate Cortex | 12 | -54 | 14 | 5.89 | 0.001 |
| LH_Vis_6; Left Primary Visual Cortex | -12 | -67 | 7 | LH_DorsAttn_FEF_1; Left PreMotor & Supplementary Motor Area | -26 | -3 | 59 | 5.18 | 0.006 |
| LH_DorsAttn_FEF_1; Left PreMotor & Supplementary Motor Area | -26 | -3 | 59 | LH_Default_pCunPCC_2; Left Ventral Posterior Cingulate Cortex | -6 | -53 | 33 | 5.11 | 0.006 |
| RH_Limbic_TempPole_1; Right Inferior Temporal Gyrus | 38 | 1 | -38 | LH_Cont_PFCl_1; Left Dorsolateral Prefrontal Cortex | -43 | 33 | 21 | 5.09 | 0.006 |
| LH_SalVentAttn_FrOperIns_2; Left Insula | -38 | 12 | 6 | LH_Default_PFC_2; Left Pars Orbitalis | -47 | 33 | -3 | 4.84 | 0.013 |
| RH_Vis_6; Right Secondary Visual Cortex | 17 | -57 | 5 | LH_SalVentAttn_PFCl_1; Left Anterior Prefrontal Cortex | -30 | 44 | 30 | 4.69 | 0.020 |
| LH_DorsAttn_Post_1; Left Fusiform Cortex | -47 | -58 | -13 | LH_DorsAttn_PrCv_1; Left PreMotor & Supplementary Motor Area | -48 | 6 | 28 | 4.59 | 0.025 |
| RH_Vis_6; Right Secondary Visual Cortex | 17 | -57 | 5 | LH_DorsAttn_FEF_1; Left PreMotor & Supplementary Motor Area | -26 | -3 | 59 | 4.50 | 0.028 |
| LH_Vis_7; Left Visual Association Cortex | -47 | -71 | 11 | LH_DorsAttn_FEF_1; Left PreMotor & Supplementary Motor Area | -26 | -3 | 59 | 4.47 | 0.028 |
| LH_Cont_PFCl_1; Left Dorsolateral Prefrontal Cortex | -43 | 33 | 21 | LH_Cont_pCun_1; Left Visual Motor Cortex | -9 | -73 | 38 | 4.46 | 0.028 |
| LH_Vis_4; Left Secondary Visual Cortex | -27 | -95 | -4 | LH_DorsAttn_FEF_1; Left PreMotor & Supplementary Motor Area | -26 | -3 | 59 | 4.42 | 0.030 |
| LH_DorsAttn_FEF_1; Left PreMotor & Supplementary Motor Area | -26 | -3 | 59 | LH_Cont_Cing_1; Left Ventral Posterior Cingulate Cortex | -4 | -26 | 33 | 4.34 | 0.036 |
| RH_Vis_5; Right Primary Visual Cortex | 8 | -76 | 5 | LH_DorsAttn_FEF_1; Left PreMotor & Supplementary Motor Area | -26 | -3 | 59 | 4.28 | 0.036 |
| LH_Vis_9; Left Visual Association Cortex | -6 | -82 | 26 | LH_Cont_PFCl_1; Left Dorsolateral Prefrontal Cortex | -43 | 33 | 21 | 4.27 | 0.036 |
| RH_Vis_4; Right Secondary Visual Cortex | 22 | -96 | -5 | LH_Cont_PFCl_1; Left Dorsolateral Prefrontal Cortex | -43 | 33 | 21 | 4.26 | 0.036 |
| LH_DorsAttn_FEF_1; Left PreMotor & Supplementary Motor Area | -26 | -3 | 59 | LH_Cont_PFCl_1; Left Dorsolateral Prefrontal Cortex | -43 | 33 | 21 | 4.26 | 0.036 |
| LH_Cont_Par_1; Left Angular Gyrus | -37 | -53 | 46 | RH_Cont_PFCl_2; Right Dorsolateral Prefrontal Cortex | 45 | 39 | 15 | 4.26 | 0.036 |
| LH_DorsAttn_FEF_1; Left PreMotor & Supplementary Motor Area | -26 | -3 | 59 | RH_DorsAttn_Post_5; Right Visual Motor Cortex | 14 | -52 | 66 | 4.23 | 0.037 |
| RH_Vis_6; Right Secondary Visual Cortex | 17 | -57 | 5 | LH_Cont_PFCl_1; Left Dorsolateral Prefrontal Cortex | -43 | 33 | 21 | 4.16 | 0.046 |
| **Eyes-Closed Beta** | | | | | | | | | |
| **ROI1 Labels (Schaefer; Talairach)** | **X** | **Y** | **Z** | **ROI2 Labels (Schaefer; Talairach)** | **X** | **Y** | **Z** | **t** | **q** |
| LH_SalVentAttn_Med_2; Left Dorsal Posterior Cingulate Cortex | -11 | -34 | 45 | RH_Cont_Cing_1; Right Ventral Posterior Cingulate Cortex | 5 | -27 | 33 | -5.17 | 0.022 |

$q \leq 0.05$ (FDR-corrected)

**Supplementary Table 5 Functional connectivity results for all participants (n=111) from post-menarche visits 1 to 2**

**Eyes-Open Theta**

| ROI1 Labels (Schaefer; Talairach) | X | Y | Z | ROI2 Labels (Schaefer; Talairach) | X | Y | Z | t | q |
|---|---|---|---|---|---|---|---|---|---|
| RH_SalVentAttn_TempOccPar_2; Right Supramarginal Gyrus | 61 | -26 | 27 | RH_Cont_PFCl_4; Right Frontal Eye Fields | 43 | 16 | 45 | 5.32 | 0.004 |
| LH_SalVentAttn_FrOperIns_2; Left Insula | -38 | 12 | 6 | RH_Cont_PFCl_2; Right Dorsolateral Prefrontal Cortex | 45 | 39 | 15 | 5.22 | 0.004 |
| LH_Cont_pCun_1; Left Visual Motor Cortex | -9 | -73 | 38 | RH_Cont_PFCl_3; Right Dorsolateral Prefrontal Cortex | 32 | 46 | 29 | 4.80 | 0.013 |
| RH_DorsAttn_PrCv_1; Right Broca's Area/Operculum | 49 | 10 | 27 | RH_SalVentAttn_TempOccPar_2; Right Supramarginal Gyrus | 61 | -26 | 27 | 4.75 | 0.013 |
| LH_Vis_8; Left Visual Association Cortex | -25 | -88 | 20 | RH_Vis_3; Right Fusiform Cortex | 49 | -60 | -11 | 4.73 | 0.013 |
| LH_SomMot_2; Left Primary Sensory Cortex | -37 | -21 | 16 | RH_DorsAttn_PrCv_1; Right Broca's Area/Operculum | 49 | 10 | 27 | 4.64 | 0.016 |
| LH_SalVentAttn_FrOperIns_2; Left Insula | -38 | 12 | 6 | LH_Default_PFC_2; Left Pars Orbitalis | -47 | 33 | -3 | 4.57 | 0.016 |
| RH_SalVentAttn_TempOccPar_1; Right Superior Temporal Gyrus | 58 | -42 | 13 | LH_Default_PFC_4; Left Anterior Prefrontal Cortex | -24 | 61 | -1 | 4.55 | 0.016 |
| RH_SomMot_6; Right Primary Motor Cortex | 41 | -22 | 60 | RH_Cont_PFCl_2; Right Dorsolateral Prefrontal Cortex | 45 | 39 | 15 | 4.55 | 0.016 |
| RH_SomMot_7; Right Primary Sensory Cortex | 30 | -37 | 64 | LH_Default_PFC_2; Left Pars Orbitalis | -47 | 33 | -3 | 4.47 | 0.017 |
| LH_Vis_4; Left Secondary Visual Cortex | -27 | -95 | -4 | RH_Vis_3; Right Fusiform Cortex | 49 | -60 | -11 | 4.39 | 0.017 |
| RH_SalVentAttn_TempOccPar_2; Right Supramarginal Gyrus | 61 | -26 | 27 | RH_Default_PFCv_2; Right Broca's Area/ Pars Triangularis | 51 | 28 | 0 | 4.36 | 0.017 |
| RH_Cont_PFCl_3; Right Dorsolateral Prefrontal Cortex | 32 | 46 | 29 | LH_Default_pCunPCC_1; Left Ventral Posterior Cingulate Cortex | -11 | -56 | 13 | 4.36 | 0.017 |
| RH_Cont_PFCl_3; Right Dorsolateral Prefrontal Cortex | 32 | 46 | 29 | LH_Default_pCunPCC_2; Left Ventral Posterior Cingulate Cortex | -6 | -53 | 33 | 4.35 | 0.017 |
| RH_Cont_PFCl_2; Right Dorsolateral Prefrontal Cortex | 45 | 39 | 15 | LH_Default_pCunPCC_2; Left Ventral Posterior Cingulate Cortex | -6 | -53 | 33 | 4.34 | 0.017 |
| LH_SalVentAttn_FrOperIns_1; Left Insula | -41 | -1 | -7 | RH_Cont_PFCl_2; Right Dorsolateral Prefrontal Cortex | 45 | 39 | 15 | 4.34 | 0.017 |
| LH_Default_PFC_2; Left Pars Orbitalis | -47 | 33 | -3 | RH_Default_Temp_3; Right Superior Temporal Gyrus | 57 | -26 | -2 | 4.33 | 0.017 |
| LH_Vis_9; Left Visual Association Cortex | -6 | -82 | 26 | RH_Vis_3; Right Fusiform Cortex | 49 | -60 | -11 | 4.32 | 0.017 |
| LH_Default_Temp_1; Left Medial Temporal Gyrus | -55 | -4 | -20 | RH_Default_PFCv_2; Right Broca's Area/ Pars Triangularis | 51 | 28 | 0 | 4.32 | 0.017 |
| LH_SalVentAttn_FrOperIns_2; Left Insula | -38 | 12 | 6 | RH_Cont_PFCl_3; Right Dorsolateral Prefrontal Cortex | 32 | 46 | 29 | 4.31 | 0.017 |
| LH_Default_PFC_2; Left Pars Orbitalis | -47 | 33 | -3 | RH_Default_Temp_1; Right Medial Temporal Gyrus | 62 | -23 | -19 | 4.31 | 0.017 |
| LH_Vis_9; Left Visual Association Cortex | -6 | -82 | 26 | RH_Default_PFCv_2; Right Broca's Area/ Pars Triangularis | 51 | 28 | 0 | 4.27 | 0.019 |
| RH_DorsAttn_Post_2; Right Primary Sensory Cortex | 50 | -24 | 42 | RH_Default_PFCv_2; Right Broca's Area/ Pars Triangularis | 51 | 28 | 0 | 4.26 | 0.019 |
| RH_SomMot_5; Right PreMotor & Supplementary Motor Area | 47 | -11 | 48 | RH_SalVentAttn_FrOperIns_1; Right Insula | 40 | 8 | 1 | 4.23 | 0.020 |
| RH_SomMot_2; Right Insula | 40 | -15 | 15 | LH_Cont_Par_1; Left Angular Gyrus | -37 | -53 | 46 | 4.17 | 0.025 |
| LH_Default_Temp_1; Left Medial Temporal Gyrus | -55 | -4 | -20 | LH_Default_PFC_3; Left Dorsal Posterior Cingulate Cortex | -6 | 47 | 0 | 4.14 | 0.025 |
| RH_DorsAttn_Post_2; Right Primary Sensory Cortex | 50 | -24 | 42 | RH_SalVentAttn_FrOperIns_1; Right Insula | 40 | 8 | 1 | 4.14 | 0.025 |
| RH_Vis_8; Right Visual Association Cortex | 13 | -86 | 29 | RH_Cont_PFCl_2; Right Dorsolateral Prefrontal Cortex | 45 | 39 | 15 | 4.12 | 0.026 |
| RH_Cont_PFCl_2; Right Dorsolateral Prefrontal Cortex | 45 | 39 | 15 | RH_Default_PFCv_1; Right Pars Orbitalis | 35 | 26 | -15 | 4.10 | 0.026 |

| | | | | | | | | | |
|---|---|---|---|---|---|---|---|---|---|
| LH_SomMot_5; Left Primary Motor Cortex | -39 | -23 | 59 | RH_SalVentAttn_TempOccPar_2; Right Supramarginal Gyrus | 61 | -26 | 27 | 4.10 | 0.026 |
| LH_SomMot_2; Left Primary Sensory Cortex | -37 | -21 | 16 | RH_SomMot_4; Right Primary Motor Cortex | 58 | -5 | 31 | 4.09 | 0.026 |
| RH_SomMot_4; Right Primary Motor Cortex | 58 | -5 | 31 | RH_Cont_PFCl_2; Right Dorsolateral Prefrontal Cortex | 45 | 39 | 15 | 4.08 | 0.027 |
| LH_Vis_3; Left Visual Association Cortex | -17 | -60 | -7 | LH_DorsAttn_PrCv_1; Left PreMotor & Supplementary Motor Area | -48 | 6 | 28 | 4.07 | 0.027 |
| LH_Cont_pCun_1; Left Visual Motor Cortex | -9 | -73 | 38 | RH_Default_PFCv_2; Right Broca's Area/ Pars Triangularis | 51 | 28 | 0 | 4.06 | 0.027 |
| RH_Vis_5; Right Primary Visual Cortex | 8 | -76 | 5 | RH_Cont_PFCl_3; Right Dorsolateral Prefrontal Cortex | 32 | 46 | 29 | 4.05 | 0.027 |
| LH_DorsAttn_Post_1; Left Fusiform Cortex | -47 | -58 | -13 | RH_Cont_PFCl_3; Right Dorsolateral Prefrontal Cortex | 32 | 46 | 29 | 4.03 | 0.028 |
| RH_Vis_8; Right Visual Association Cortex | 13 | -86 | 29 | RH_Default_PFCv_2; Right Broca's Area/ Pars Triangularis | 51 | 28 | 0 | 4.02 | 0.028 |
| RH_DorsAttn_Post_2; Right Primary Sensory Cortex | 50 | -24 | 42 | RH_Cont_PFCl_2; Right Dorsolateral Prefrontal Cortex | 45 | 39 | 15 | 4.01 | 0.028 |
| RH_DorsAttn_Post_1; Right Visual Association Cortex | 50 | -62 | 16 | LH_Default_PFC_4; Left Anterior Prefrontal Cortex | -24 | 61 | -1 | 4.01 | 0.028 |
| LH_Vis_9; Left Visual Association Cortex | -6 | -82 | 26 | RH_Cont_PFCl_2; Right Dorsolateral Prefrontal Cortex | 45 | 39 | 15 | 4.00 | 0.028 |
| RH_Cont_PFCl_2; Right Dorsolateral Prefrontal Cortex | 45 | 39 | 15 | RH_Default_pCunPCC_2; Right Ventral Posterior Cingulate Cortex | 7 | -52 | 31 | 4.00 | 0.028 |
| RH_SomMot_6; Right Primary Motor Cortex | 41 | -22 | 60 | RH_Default_PFCv_2; Right Broca's Area/ Pars Triangularis | 51 | 28 | 0 | 3.98 | 0.029 |
| LH_Default_Temp_1; Left Medial Temporal Gyrus | -55 | -4 | -20 | RH_Default_PFCdPFCm_3; Right Frontal Eye Fields | 26 | 24 | 50 | 3.97 | 0.030 |
| RH_SomMot_5; Right PreMotor & Supplementary Motor Area | 47 | -11 | 48 | LH_Cont_pCun_1; Left Visual Motor Cortex | -9 | -73 | 38 | 3.96 | 0.030 |
| RH_Cont_PFCl_1; Right Anterior Prefrontal Cortex | 30 | 58 | -3 | RH_Cont_PFCl_2; Right Dorsolateral Prefrontal Cortex | 45 | 39 | 15 | 3.96 | 0.030 |
| RH_Cont_PFCl_3; Right Dorsolateral Prefrontal Cortex | 32 | 46 | 29 | RH_Cont_pCun_1; Right Visual Motor Cortex | 9 | -66 | 43 | 3.95 | 0.030 |
| LH_DorsAttn_Post_4; Left Primary Sensory Cortex | -42 | -34 | 48 | RH_SalVentAttn_TempOccPar_2; Right Supramarginal Gyrus | 61 | -26 | 27 | 3.93 | 0.031 |
| RH_Cont_Par_2; Right Angular Gyrus | 45 | -63 | 46 | RH_Cont_PFCmp_1; Right Frontal Eye Fields | 6 | 28 | 30 | 3.93 | 0.031 |
| RH_Cont_PFCl_2; Right Dorsolateral Prefrontal Cortex | 45 | 39 | 15 | LH_Default_PFC_1; Left Pars Orbitalis | -35 | 21 | -11 | 3.91 | 0.032 |
| LH_SomMot_2; Left Primary Sensory Cortex | -37 | -21 | 16 | RH_SomMot_3; Right Primary Motor Cortex | 57 | -4 | 11 | 3.90 | 0.032 |
| RH_Vis_2; Right Fusiform Cortex | 27 | -66 | -12 | RH_Vis_3; Right Fusiform Cortex | 49 | -60 | -11 | 3.90 | 0.032 |
| LH_Vis_9; Left Visual Association Cortex | -6 | -82 | 26 | RH_Cont_Cing_1; Right Ventral Posterior Cingulate Cortex | 5 | -27 | 33 | 3.90 | 0.032 |
| LH_Vis_6; Left Primary Visual Cortex | -12 | -67 | 7 | RH_Cont_PFCl_3; Right Dorsolateral Prefrontal Cortex | 32 | 46 | 29 | 3.89 | 0.032 |
| RH_SomMot_5; Right PreMotor & Supplementary Motor Area | 47 | -11 | 48 | LH_Cont_Par_1; Left Angular Gyrus | -37 | -53 | 46 | 3.88 | 0.032 |
| RH_DorsAttn_Post_1; Right Visual Association Cortex | 50 | -62 | 16 | RH_Cont_PFCl_2; Right Dorsolateral Prefrontal Cortex | 45 | 39 | 15 | 3.87 | 0.032 |
| LH_Cont_pCun_1; Left Visual Motor Cortex | -9 | -73 | 38 | RH_Cont_PFCl_2; Right Dorsolateral Prefrontal Cortex | 45 | 39 | 15 | 3.86 | 0.032 |
| RH_DorsAttn_Post_1; Right Visual Association Cortex | 50 | -62 | 16 | RH_DorsAttn_PrCv_1; Right Broca's Area/Operculum | 49 | 10 | 27 | 3.85 | 0.032 |
| RH_Cont_PFCl_3; Right Dorsolateral Prefrontal Cortex | 32 | 46 | 29 | RH_Default_pCunPCC_2; Right Ventral Posterior Cingulate Cortex | 7 | -52 | 31 | 3.85 | 0.032 |
| RH_SomMot_5; Right PreMotor & Supplementary Motor Area | 47 | -11 | 48 | LH_Default_pCunPCC_2; Left Ventral Posterior Cingulate Cortex | -6 | -53 | 33 | 3.85 | 0.032 |
| RH_Cont_Par_2; Right Angular Gyrus | 45 | -63 | 46 | LH_Default_PFC_1; Left Pars Orbitalis | -35 | 21 | -11 | 3.85 | 0.032 |
| RH_SalVentAttn_TempOccPar_2; Right Supramarginal Gyrus | 61 | -26 | 27 | RH_Cont_PFCl_2; Right Dorsolateral Prefrontal Cortex | 45 | 39 | 15 | 3.85 | 0.032 |

| | | | | | | | | | |
|---|---|---|---|---|---|---|---|---|---|
| RH_Vis_2; Right Fusiform Cortex | 27 | -66 | -12 | LH_Cont_Cing_1; Left Ventral Posterior Cingulate Cortex | -4 | -26 | 33 | 3.85 | 0.032 |
| LH_Cont_Cing_1; Left Ventral Posterior Cingulate Cortex | -4 | -26 | 33 | RH_Cont_PFCl_2; Right Dorsolateral Prefrontal Cortex | 45 | 39 | 15 | 3.84 | 0.032 |
| RH_Cont_PFCl_2; Right Dorsolateral Prefrontal Cortex | 45 | 39 | 15 | LH_Default_pCunPCC_1; Left Ventral Posterior Cingulate Cortex | -11 | -56 | 13 | 3.84 | 0.032 |
| LH_Vis_8; Left Visual Association Cortex | -25 | -88 | 20 | RH_Default_PFCdPFCm_1; Right Anterior Prefrontal Cortex | 7 | 48 | 1 | 3.83 | 0.033 |
| RH_DorsAttn_PrCv_1; Right Broca's Area/Operculum | 49 | 10 | 27 | RH_Default_Par_1; Right Angular Gyrus | 55 | -51 | 31 | 3.83 | 0.033 |
| RH_Default_Temp_1; Right Medial Temporal Gyrus | 62 | -23 | -19 | RH_Default_Temp_2; Right Temporal Pole | 51 | 7 | -18 | 3.81 | 0.033 |
| RH_DorsAttn_Post_2; Right Primary Sensory Cortex | 50 | -24 | 42 | LH_Cont_pCun_1; Left Visual Motor Cortex | -9 | -73 | 38 | 3.81 | 0.033 |
| RH_Cont_PFCl_2; Right Dorsolateral Prefrontal Cortex | 45 | 39 | 15 | RH_Default_pCunPCC_1; Right Ventral Posterior Cingulate Cortex | 12 | -54 | 14 | 3.81 | 0.033 |
| RH_Vis_4; Right Secondary Visual Cortex | 22 | -96 | -5 | RH_Cont_PFCl_3; Right Dorsolateral Prefrontal Cortex | 32 | 46 | 29 | 3.80 | 0.034 |
| RH_SomMot_4; Right Primary Motor Cortex | 58 | -5 | 31 | RH_DorsAttn_Post_5; Right Visual Motor Cortex | 14 | -52 | 66 | 3.79 | 0.034 |
| LH_SomMot_2; Left Primary Sensory Cortex | -37 | -21 | 16 | LH_Default_Temp_1; Left Medial Temporal Gyrus | -55 | -4 | -20 | 3.79 | 0.034 |
| RH_Cont_PFCl_3; Right Dorsolateral Prefrontal Cortex | 32 | 46 | 29 | RH_Default_pCunPCC_1; Right Ventral Posterior Cingulate Cortex | 12 | -54 | 14 | 3.78 | 0.034 |
| RH_SalVentAttn_Med_1; Right Dorsal Posterior Cingulate Cortex | 11 | -31 | 45 | LH_Cont_Cing_1; Left Ventral Posterior Cingulate Cortex | -4 | -26 | 33 | 3.77 | 0.034 |
| RH_Vis_4; Right Secondary Visual Cortex | 22 | -96 | -5 | LH_SalVentAttn_FrOperIns_2; Left Insula | -38 | 12 | 6 | 3.77 | 0.034 |
| LH_Default_pCunPCC_1; Left Ventral Posterior Cingulate Cortex | -11 | -56 | 13 | RH_Default_PFCv_2; Right Broca's Area/ Pars Triangularis | 51 | 28 | 0 | 3.77 | 0.034 |
| LH_SalVentAttn_Med_1; Left Dorsal Anterior Cingulate Cortex | -5 | 20 | 34 | RH_Cont_Par_2; Right Angular Gyrus | 45 | -63 | 46 | 3.77 | 0.034 |
| RH_Cont_Par_1; Right Supramarginal Gyrus | 57 | -39 | 44 | RH_Default_PFCdPFCm_3; Right Frontal Eye Fields | 26 | 24 | 50 | 3.77 | 0.034 |
| RH_Default_PFCv_1; Right Pars Orbitalis | 35 | 26 | -15 | RH_Default_PFCv_2; Right Broca's Area/ Pars Triangularis | 51 | 28 | 0 | 3.77 | 0.034 |
| RH_SomMot_8; Right Primary Motor Cortex | 6 | -26 | 70 | RH_Cont_PFCl_4; Right Frontal Eye Fields | 43 | 16 | 45 | 3.76 | 0.034 |
| LH_SomMot_2; Left Primary Sensory Cortex | -37 | -21 | 16 | LH_SalVentAttn_FrOperIns_2; Left Insula | -38 | 12 | 6 | 3.75 | 0.035 |
| RH_DorsAttn_Post_1; Right Visual Association Cortex | 50 | -62 | 16 | RH_Cont_PFCl_3; Right Dorsolateral Prefrontal Cortex | 32 | 46 | 29 | 3.74 | 0.035 |
| RH_DorsAttn_Post_1; Right Visual Association Cortex | 50 | -62 | 16 | RH_Default_Par_1; Right Angular Gyrus | 55 | -51 | 31 | 3.74 | 0.035 |
| LH_SomMot_2; Left Primary Sensory Cortex | -37 | -21 | 16 | LH_Default_PFC_5; Left Dorsolateral Prefrontal Cortex | -9 | 48 | 41 | 3.74 | 0.035 |
| RH_DorsAttn_Post_1; Right Visual Association Cortex | 50 | -62 | 16 | RH_Default_PFCv_2; Right Broca's Area/ Pars Triangularis | 51 | 28 | 0 | 3.74 | 0.035 |
| RH_Vis_3; Right Fusiform Cortex | 49 | -60 | -11 | RH_Vis_6; Right Secondary Visual Cortex | 17 | -57 | 5 | 3.74 | 0.035 |
| RH_SalVentAttn_TempOccPar_2; Right Supramarginal Gyrus | 61 | -26 | 27 | RH_SalVentAttn_FrOperIns_1; Right Insula | 40 | 8 | 1 | 3.73 | 0.035 |
| RH_Vis_3; Right Fusiform Cortex | 49 | -60 | -11 | LH_Cont_pCun_1; Left Visual Motor Cortex | -9 | -73 | 38 | 3.72 | 0.036 |
| RH_SomMot_3; Right Primary Motor Cortex | 57 | -4 | 11 | RH_DorsAttn_Post_2; Right Primary Sensory Cortex | 50 | -24 | 42 | 3.72 | 0.036 |
| LH_Vis_3; Left Visual Association Cortex | -17 | -60 | -7 | LH_Default_PFC_6; Left PreMotor & Supplementary Motor Area | -41 | 14 | 48 | 3.71 | 0.036 |
| RH_SomMot_4; Right Primary Motor Cortex | 58 | -5 | 31 | RH_Default_Par_1; Right Angular Gyrus | 55 | -51 | 31 | 3.71 | 0.036 |
| LH_Vis_6; Left Primary Visual Cortex | -12 | -67 | 7 | RH_SomMot_5; Right PreMotor & Supplementary Motor Area | 47 | -11 | 48 | 3.70 | 0.037 |
| RH_Vis_2; Right Fusiform Cortex | 27 | -66 | -12 | RH_Vis_7; Right Visual Association Cortex | 36 | -82 | 16 | 3.69 | 0.037 |

| | | | | | | | | | |
|---|---|---|---|---|---|---|---|---|---|
| LH_SomMot_2; Left Primary Sensory Cortex | -37 | -21 | 16 | RH_SalVentAttn_FrOperIns_1; Right Insula | 40 | 8 | 1 | 3.68 | 0.037 |
| RH_SomMot_6; Right Primary Motor Cortex | 41 | -22 | 60 | RH_Cont_PFCmp_1; Right Frontal Eye Fields | 6 | 28 | 30 | 3.68 | 0.037 |
| RH_Vis_2; Right Fusiform Cortex | 27 | -66 | -12 | RH_Cont_pCun_1; Right Visual Motor Cortex | 9 | -66 | 43 | 3.68 | 0.037 |
| RH_DorsAttn_Post_4; Right Visual Motor Cortex | 27 | -67 | 51 | RH_Cont_PFCl_3; Right Dorsolateral Prefrontal Cortex | 32 | 46 | 29 | 3.68 | 0.037 |
| LH_SalVentAttn_FrOperIns_1; Left Insula | -41 | -1 | -7 | RH_Cont_PFCmp_1; Right Frontal Eye Fields | 6 | 28 | 30 | 3.68 | 0.037 |
| RH_SomMot_4; Right Primary Motor Cortex | 58 | -5 | 31 | LH_Default_pCunPCC_1; Left Ventral Posterior Cingulate Cortex | -11 | -56 | 13 | 3.68 | 0.037 |
| RH_SalVentAttn_Med_1; Right Dorsal Posterior Cingulate Cortex | 11 | -31 | 45 | LH_Cont_Par_1; Left Angular Gyrus | -37 | -53 | 46 | 3.67 | 0.037 |
| LH_SomMot_4; Left PreMotor & Supplementary Motor Area | -55 | -8 | 34 | RH_Cont_PFCmp_1; Right Frontal Eye Fields | 6 | 28 | 30 | 3.67 | 0.037 |
| LH_Cont_Par_1; Left Angular Gyrus | -37 | -53 | 46 | RH_Cont_PFCl_4; Right Frontal Eye Fields | 43 | 16 | 45 | 3.67 | 0.037 |
| RH_SomMot_4; Right Primary Motor Cortex | 58 | -5 | 31 | RH_SalVentAttn_TempOccPar_2; Right Supramarginal Gyrus | 61 | -26 | 27 | 3.66 | 0.038 |
| RH_SomMot_4; Right Primary Motor Cortex | 58 | -5 | 31 | LH_Cont_Par_1; Left Angular Gyrus | -37 | -53 | 46 | 3.66 | 0.038 |
| LH_Default_PFC_2; Left Pars Orbitalis | -47 | 33 | -3 | RH_Default_PFCdPFCm_3; Right Frontal Eye Fields | 26 | 24 | 50 | 3.65 | 0.038 |
| RH_SomMot_3; Right Primary Motor Cortex | 57 | -4 | 11 | RH_SalVentAttn_TempOccPar_2; Right Supramarginal Gyrus | 61 | -26 | 27 | 3.65 | 0.038 |
| RH_DorsAttn_Post_4; Right Visual Motor Cortex | 27 | -67 | 51 | RH_Cont_PFCl_4; Right Frontal Eye Fields | 43 | 16 | 45 | 3.65 | 0.038 |
| RH_DorsAttn_Post_2; Right Primary Sensory Cortex | 50 | -24 | 42 | LH_Cont_Par_1; Left Angular Gyrus | -37 | -53 | 46 | 3.64 | 0.039 |
| RH_SomMot_4; Right Primary Motor Cortex | 58 | -5 | 31 | RH_Cont_PFCl_4; Right Frontal Eye Fields | 43 | 16 | 45 | 3.63 | 0.039 |
| RH_Cont_Par_2; Right Angular Gyrus | 45 | -63 | 46 | LH_Default_PFC_4; Left Anterior Prefrontal Cortex | -24 | 61 | -1 | 3.63 | 0.039 |
| RH_SomMot_1; Right Primary Auditory Cortex | 53 | -16 | 7 | RH_DorsAttn_PrCv_1; Right Broca's Area/Operculum | 49 | 10 | 27 | 3.63 | 0.039 |
| RH_SomMot_5; Right PreMotor & Supplementary Motor Area | 47 | -11 | 48 | LH_Default_pCunPCC_1; Left Ventral Posterior Cingulate Cortex | -11 | -56 | 13 | 3.63 | 0.039 |
| LH_Vis_4; Left Secondary Visual Cortex | -27 | -95 | -4 | RH_Cont_PFCmp_1; Right Frontal Eye Fields | 6 | 28 | 30 | 3.62 | 0.039 |
| RH_Vis_7; Right Visual Association Cortex | 36 | -82 | 16 | RH_Cont_Cing_1; Right Ventral Posterior Cingulate Cortex | 5 | -27 | 33 | 3.62 | 0.040 |
| LH_SomMot_1; Left Supramarginal Gyrus | -53 | -23 | 8 | RH_Default_PFCv_2; Right Broca's Area/ Pars Triangularis | 51 | 28 | 0 | 3.61 | 0.040 |
| LH_SomMot_2; Left Primary Sensory Cortex | -37 | -21 | 16 | RH_SomMot_5; Right PreMotor & Supplementary Motor Area | 47 | -11 | 48 | 3.60 | 0.041 |
| RH_DorsAttn_FEF_1; Right PreMotor & Supplementary Motor Area | 28 | -3 | 59 | RH_Cont_PFCl_2; Right Dorsolateral Prefrontal Cortex | 45 | 39 | 15 | 3.60 | 0.041 |
| LH_Vis_4; Left Secondary Visual Cortex | -27 | -95 | -4 | RH_DorsAttn_Post_3; Right Visual Motor Cortex | 38 | -45 | 49 | 3.60 | 0.041 |
| RH_Vis_4; Right Secondary Visual Cortex | 22 | -96 | -5 | RH_Default_PFCv_2; Right Broca's Area/ Pars Triangularis | 51 | 28 | 0 | 3.60 | 0.041 |
| RH_Default_PFCv_2; Right Broca's Area/ Pars Triangularis | 51 | 28 | 0 | RH_Default_pCunPCC_1; Right Ventral Posterior Cingulate Cortex | 12 | -54 | 14 | 3.59 | 0.041 |
| RH_SalVentAttn_TempOccPar_2; Right Supramarginal Gyrus | 61 | -26 | 27 | RH_Cont_PFCl_3; Right Dorsolateral Prefrontal Cortex | 32 | 46 | 29 | 3.59 | 0.041 |
| RH_Vis_7; Right Visual Association Cortex | 36 | -82 | 16 | LH_Default_PFC_6; Left PreMotor & Supplementary Motor Area | -41 | 14 | 48 | 3.58 | 0.041 |
| LH_Vis_6; Left Primary Visual Cortex | -12 | -67 | 7 | RH_SomMot_4; Right Primary Motor Cortex | 58 | -5 | 31 | 3.58 | 0.041 |
| LH_Vis_3; Left Visual Association Cortex | -17 | -60 | -7 | RH_SomMot_6; Right Primary Motor Cortex | 41 | -22 | 60 | 3.58 | 0.041 |
| RH_Cont_PFCmp_1; Right Frontal Eye Fields | 6 | 28 | 30 | LH_Default_PFC_1; Left Pars Orbitalis | -35 | 21 | -11 | 3.58 | 0.041 |

| | | | | | | | | | |
|---|---|---|---|---|---|---|---|---|---|
| LH_DorsAttn_Post_2; Left Supramarginal Gyrus | -57 | -25 | 39 | RH_Cont_PFCl_4; Right Frontal Eye Fields | 43 | 16 | 45 | 3.58 | 0.041 |
| RH_SomMot_5; Right PreMotor & Supplementary Motor Area | 47 | -11 | 48 | RH_DorsAttn_Post_5; Right Visual Motor Cortex | 14 | -52 | 66 | 3.57 | 0.041 |
| LH_Vis_4; Left Secondary Visual Cortex | -27 | -95 | -4 | LH_Default_PFC_4; Left Anterior Prefrontal Cortex | -24 | 61 | -1 | 3.57 | 0.041 |
| LH_Vis_4; Left Secondary Visual Cortex | -27 | -95 | -4 | LH_Default_PFC_3; Left Dorsal Posterior Cingulate Cortex | -6 | 47 | 0 | 3.57 | 0.041 |
| RH_DorsAttn_Post_2; Right Primary Sensory Cortex | 50 | -24 | 42 | RH_Default_Temp_3; Right Superior Temporal Gyrus | 57 | -26 | -2 | 3.57 | 0.041 |
| RH_SomMot_4; Right Primary Motor Cortex | 58 | -5 | 31 | RH_DorsAttn_Post_2; Right Primary Sensory Cortex | 50 | -24 | 42 | 3.57 | 0.041 |
| LH_Limbic_TempPole_1; Left Temporal Pole | -32 | 2 | -37 | LH_Default_Temp_1; Left Medial Temporal Gyrus | -55 | -4 | -20 | 3.56 | 0.042 |
| LH_SalVentAttn_FrOperIns_1; Left Insula | -41 | -1 | -7 | RH_Default_PFCv_2; Right Broca's Area/ Pars Triangularis | 51 | 28 | 0 | 3.55 | 0.043 |
| LH_Vis_2; Left Visual Association Cortex | -26 | -77 | -14 | LH_Limbic_OFC_1; Left Orbitofrontal Cortex | -14 | 32 | -20 | 3.55 | 0.043 |
| RH_Cont_PFCl_2; Right Dorsolateral Prefrontal Cortex | 45 | 39 | 15 | RH_Default_PFCdPFCm_1; Right Anterior Prefrontal Cortex | 7 | 48 | 1 | 3.55 | 0.043 |
| RH_Limbic_TempPole_1; Right Inferior Temporal Gyrus | 38 | 1 | -38 | RH_Default_PFCdPFCm_2; Right Dorsolateral Prefrontal Cortex | 11 | 50 | 39 | 3.54 | 0.043 |
| RH_Vis_7; Right Visual Association Cortex | 36 | -82 | 16 | LH_Cont_Cing_1; Left Ventral Posterior Cingulate Cortex | -4 | -26 | 33 | 3.54 | 0.043 |
| RH_SomMot_4; Right Primary Motor Cortex | 58 | -5 | 31 | LH_Default_Temp_2; Left Medial Temporal Gyrus | -58 | -32 | -1 | 3.53 | 0.043 |
| RH_Cont_PFCl_4; Right Frontal Eye Fields | 43 | 16 | 45 | RH_Default_Par_1; Right Angular Gyrus | 55 | -51 | 31 | 3.53 | 0.043 |
| LH_SomMot_3; Left Primary Sensory Cortex | -54 | -12 | 13 | RH_Default_PFCv_2; Right Broca's Area/ Pars Triangularis | 51 | 28 | 0 | 3.53 | 0.043 |
| RH_Cont_PFCl_2; Right Dorsolateral Prefrontal Cortex | 45 | 39 | 15 | LH_Default_PFC_2; Left Pars Orbitalis | -47 | 33 | -3 | 3.53 | 0.043 |
| LH_Default_pCunPCC_2; Left Ventral Posterior Cingulate Cortex | -6 | -53 | 33 | RH_Default_PFCv_2; Right Broca's Area/ Pars Triangularis | 51 | 28 | 0 | 3.53 | 0.043 |
| RH_SomMot_4; Right Primary Motor Cortex | 58 | -5 | 31 | RH_Cont_pCun_1; Right Visual Motor Cortex | 9 | -66 | 43 | 3.52 | 0.044 |
| RH_SomMot_1; Right Primary Auditory Cortex | 53 | -16 | 7 | LH_Default_PFC_2; Left Pars Orbitalis | -47 | 33 | -3 | 3.51 | 0.044 |
| RH_SomMot_5; Right PreMotor & Supplementary Motor Area | 47 | -11 | 48 | RH_Default_PFCdPFCm_2; Right Dorsolateral Prefrontal Cortex | 11 | 50 | 39 | 3.51 | 0.044 |
| RH_Vis_6; Right Secondary Visual Cortex | 17 | -57 | 5 | LH_Cont_pCun_1; Left Visual Motor Cortex | -9 | -73 | 38 | 3.51 | 0.044 |
| LH_Cont_Par_1; Left Angular Gyrus | -37 | -53 | 46 | LH_Default_Temp_1; Left Medial Temporal Gyrus | -55 | -4 | -20 | 3.51 | 0.045 |
| RH_Cont_PFCl_3; Right Dorsolateral Prefrontal Cortex | 32 | 46 | 29 | LH_Default_PFC_1; Left Pars Orbitalis | -35 | 21 | -11 | 3.50 | 0.045 |
| LH_SomMot_2; Left Primary Sensory Cortex | -37 | -21 | 16 | RH_SomMot_2; Right Insula | 40 | -15 | 15 | 3.50 | 0.045 |
| LH_Default_PFC_2; Left Pars Orbitalis | -47 | 33 | -3 | LH_Default_PFC_5; Left Dorsolateral Prefrontal Cortex | -9 | 48 | 41 | 3.50 | 0.045 |
| RH_DorsAttn_PrCv_1; Right Broca's Area/Operculum | 49 | 10 | 27 | RH_Cont_pCun_1; Right Visual Motor Cortex | 9 | -66 | 43 | 3.50 | 0.045 |
| RH_DorsAttn_Post_4; Right Visual Motor Cortex | 27 | -67 | 51 | RH_DorsAttn_PrCv_1; Right Broca's Area/Operculum | 49 | 10 | 27 | 3.50 | 0.045 |
| RH_Cont_Par_2; Right Angular Gyrus | 45 | -63 | 46 | RH_Cont_PFCl_3; Right Dorsolateral Prefrontal Cortex | 32 | 46 | 29 | 3.50 | 0.045 |
| RH_Vis_7; Right Visual Association Cortex | 36 | -82 | 16 | RH_Cont_PFCl_1; Right Anterior Prefrontal Cortex | 30 | 58 | -3 | 3.49 | 0.045 |
| RH_SomMot_4; Right Primary Motor Cortex | 58 | -5 | 31 | RH_Default_Temp_3; Right Superior Temporal Gyrus | 57 | -26 | -2 | 3.49 | 0.045 |
| RH_Cont_PFCl_2; Right Dorsolateral Prefrontal Cortex | 45 | 39 | 15 | RH_Default_PFCv_2; Right Broca's Area/ Pars Triangularis | 51 | 28 | 0 | 3.49 | 0.045 |
| RH_Vis_7; Right Visual Association Cortex | 36 | -82 | 16 | LH_Default_pCunPCC_2; Left Ventral Posterior Cingulate Cortex | -6 | -53 | 33 | 3.49 | 0.045 |

| | | | | | | | | | |
|---|---|---|---|---|---|---|---|---|---|
| RH_Vis_2; Right Fusiform Cortex | 27 | -66 | -12 | LH_Default_PFC_2; Left Pars Orbitalis | -47 | 33 | -3 | 3.49 | 0.045 |
| RH_SomMot_5; Right PreMotor & Supplementary Motor Area | 47 | -11 | 48 | RH_Default_PFCdPFCm_3; Right Frontal Eye Fields | 26 | 24 | 50 | 3.48 | 0.045 |
| RH_Vis_2; Right Fusiform Cortex | 27 | -66 | -12 | RH_Cont_Cing_1; Right Ventral Posterior Cingulate Cortex | 5 | -27 | 33 | 3.48 | 0.045 |
| RH_SomMot_5; Right PreMotor & Supplementary Motor Area | 47 | -11 | 48 | RH_Default_Temp_1; Right Medial Temporal Gyrus | 62 | -23 | -19 | 3.47 | 0.045 |
| RH_Vis_4; Right Secondary Visual Cortex | 22 | -96 | -5 | RH_Cont_PFCl_2; Right Dorsolateral Prefrontal Cortex | 45 | 39 | 15 | 3.47 | 0.045 |
| RH_Vis_7; Right Visual Association Cortex | 36 | -82 | 16 | RH_Default_PFCv_2; Right Broca's Area/ Pars Triangularis | 51 | 28 | 0 | 3.47 | 0.045 |
| RH_Cont_PFCl_2; Right Dorsolateral Prefrontal Cortex | 45 | 39 | 15 | LH_Default_PFC_4; Left Anterior Prefrontal Cortex | -24 | 61 | -1 | 3.47 | 0.046 |
| RH_DorsAttn_PrCv_1; Right Broca's Area/Operculum | 49 | 10 | 27 | RH_Cont_Par_1; Right Supramarginal Gyrus | 57 | -39 | 44 | 3.46 | 0.047 |
| LH_SomMot_2; Left Primary Sensory Cortex | -37 | -21 | 16 | RH_Cont_PFCl_4; Right Frontal Eye Fields | 43 | 16 | 45 | 3.46 | 0.047 |
| RH_Cont_PFCmp_1; Right Frontal Eye Fields | 6 | 28 | 30 | LH_Default_Temp_1; Left Medial Temporal Gyrus | -55 | -4 | -20 | 3.46 | 0.047 |
| RH_Limbic_TempPole_1; Right Inferior Temporal Gyrus | 38 | 1 | -38 | RH_Cont_PFCl_3; Right Dorsolateral Prefrontal Cortex | 32 | 46 | 29 | 3.45 | 0.047 |
| RH_Vis_6; Right Secondary Visual Cortex | 17 | -57 | 5 | LH_Default_PFC_2; Left Pars Orbitalis | -47 | 33 | -3 | 3.45 | 0.047 |
| RH_Vis_2; Right Fusiform Cortex | 27 | -66 | -12 | LH_Cont_pCun_1; Left Visual Motor Cortex | -9 | -73 | 38 | 3.45 | 0.047 |
| RH_SomMot_1; Right Primary Auditory Cortex | 53 | -16 | 7 | RH_SomMot_5; Right PreMotor & Supplementary Motor Area | 47 | -11 | 48 | 3.45 | 0.047 |
| RH_SomMot_1; Right Primary Auditory Cortex | 53 | -16 | 7 | RH_DorsAttn_Post_2; Right Primary Sensory Cortex | 50 | -24 | 42 | 3.44 | 0.047 |
| RH_Cont_PFCl_2; Right Dorsolateral Prefrontal Cortex | 45 | 39 | 15 | RH_Cont_PFCmp_1; Right Frontal Eye Fields | 6 | 28 | 30 | 3.44 | 0.047 |
| LH_SomMot_5; Left Primary Motor Cortex | -39 | -23 | 59 | LH_DorsAttn_Post_2; Left Supramarginal Gyrus | -57 | -25 | 39 | 3.44 | 0.047 |
| RH_SomMot_5; Right PreMotor & Supplementary Motor Area | 47 | -11 | 48 | LH_Cont_Cing_1; Left Ventral Posterior Cingulate Cortex | -4 | -26 | 33 | 3.44 | 0.047 |
| RH_SalVentAttn_TempOccPar_2; Right Supramarginal Gyrus | 61 | -26 | 27 | LH_Cont_Par_1; Left Angular Gyrus | -37 | -53 | 46 | 3.43 | 0.047 |
| RH_Vis_5; Right Primary Visual Cortex | 8 | -76 | 5 | RH_Cont_PFCl_2; Right Dorsolateral Prefrontal Cortex | 45 | 39 | 15 | 3.43 | 0.047 |
| RH_Cont_PFCl_2; Right Dorsolateral Prefrontal Cortex | 45 | 39 | 15 | RH_Default_Temp_1; Right Medial Temporal Gyrus | 62 | -23 | -19 | 3.43 | 0.047 |
| RH_SomMot_6; Right Primary Motor Cortex | 41 | -22 | 60 | LH_Cont_pCun_1; Left Visual Motor Cortex | -9 | -73 | 38 | 3.43 | 0.047 |
| LH_Default_PFC_4; Left Anterior Prefrontal Cortex | -24 | 61 | -1 | RH_Default_Par_1; Right Angular Gyrus | 55 | -51 | 31 | 3.43 | 0.047 |
| RH_Vis_7; Right Visual Association Cortex | 36 | -82 | 16 | LH_SomMot_2; Left Primary Sensory Cortex | -37 | -21 | 16 | 3.42 | 0.047 |
| RH_DorsAttn_Post_3; Right Visual Motor Cortex | 38 | -45 | 49 | RH_Cont_PFCl_3; Right Dorsolateral Prefrontal Cortex | 32 | 46 | 29 | 3.42 | 0.047 |
| RH_SomMot_7; Right Primary Sensory Cortex | 30 | -37 | 64 | RH_Cont_PFCmp_1; Right Frontal Eye Fields | 6 | 28 | 30 | 3.42 | 0.047 |
| LH_SalVentAttn_Med_2; Left Dorsal Posterior Cingulate Cortex | -11 | -34 | 45 | RH_Cont_Par_2; Right Angular Gyrus | 45 | -63 | 46 | 3.42 | 0.047 |
| RH_SomMot_6; Right Primary Motor Cortex | 41 | -22 | 60 | RH_Default_PFCdPFCm_1; Right Anterior Prefrontal Cortex | 7 | 48 | 1 | 3.42 | 0.047 |
| RH_DorsAttn_Post_3; Right Visual Motor Cortex | 38 | -45 | 49 | RH_SalVentAttn_Med_2; Right PreMotor & Supplementary Motor Area | 7 | 6 | 52 | 3.42 | 0.048 |
| RH_Vis_3; Right Fusiform Cortex | 49 | -60 | -11 | RH_DorsAttn_Post_2; Right Primary Sensory Cortex | 50 | -24 | 42 | 3.41 | 0.049 |
| LH_DorsAttn_Post_2; Left Supramarginal Gyrus | -57 | -25 | 39 | LH_Default_PFC_2; Left Pars Orbitalis | -47 | 33 | -3 | 3.41 | 0.049 |

| | | | | | | | | | |
|---|---|---|---|---|---|---|---|---|---|
| RH_SomMot_5; Right PreMotor & Supplementary Motor Area | 47 | -11 | 48 | RH_Default_Temp_3; Right Superior Temporal Gyrus | 57 | -26 | -2 | 3.40 | 0.049 |
| RH_SomMot_6; Right Primary Motor Cortex | 41 | -22 | 60 | LH_Default_PFC_2; Left Pars Orbitalis | -47 | 33 | -3 | 3.40 | 0.049 |
| LH_Vis_8; Left Visual Association Cortex | -25 | -88 | 20 | RH_Cont_PFCmp_1; Right Frontal Eye Fields | 6 | 28 | 30 | 3.39 | 0.050 |
| LH_Vis_9; Left Visual Association Cortex | -6 | -82 | 26 | RH_Vis_5; Right Primary Visual Cortex | 8 | -76 | 5 | 3.39 | 0.050 |
| RH_DorsAttn_Post_2; Right Primary Sensory Cortex | 50 | -24 | 42 | RH_Cont_PFCl_3; Right Dorsolateral Prefrontal Cortex | 32 | 46 | 29 | 3.39 | 0.050 |
| LH_Default_Temp_1; Left Medial Temporal Gyrus | -55 | -4 | -20 | LH_Default_PFC_4; Left Anterior Prefrontal Cortex | -24 | 61 | -1 | 3.39 | 0.050 |
| LH_Vis_1; Left Fusiform Cortex | -26 | -34 | -17 | RH_SalVentAttn_TempOccPar_2; Right Supramarginal Gyrus | 61 | -26 | 27 | 3.39 | 0.050 |
| LH_DorsAttn_Post_2; Left Supramarginal Gyrus | -57 | -25 | 39 | RH_Default_PFCdPFCm_2; Right Dorsolateral Prefrontal Cortex | 11 | 50 | 39 | 3.39 | 0.050 |
| RH_SomMot_5; Right PreMotor & Supplementary Motor Area | 47 | -11 | 48 | LH_DorsAttn_Post_3; Left Visual Motor Cortex | -24 | -68 | 49 | 3.39 | 0.050 |
| LH_SomMot_2; Left Primary Sensory Cortex | -37 | -21 | 16 | LH_Default_PFC_2; Left Pars Orbitalis | -47 | 33 | -3 | 3.38 | 0.050 |
| RH_Vis_1; Right Fusiform Cortex | 32 | -31 | -22 | RH_Vis_2; Right Fusiform Cortex | 27 | -66 | -12 | 3.38 | 0.050 |
| LH_SomMot_2; Left Primary Sensory Cortex | -37 | -21 | 16 | RH_DorsAttn_Post_2; Right Primary Sensory Cortex | 50 | -24 | 42 | 3.38 | 0.050 |
| RH_SalVentAttn_Med_1; Right Dorsal Posterior Cingulate Cortex | 11 | -31 | 45 | RH_Default_pCunPCC_1; Right Ventral Posterior Cingulate Cortex | 12 | -54 | 14 | 3.38 | 0.050 |
| LH_Cont_Cing_1; Left Ventral Posterior Cingulate Cortex | -4 | -26 | 33 | RH_Cont_Par_2; Right Angular Gyrus | 45 | -63 | 46 | 3.38 | 0.050 |
| LH_SomMot_3; Left Primary Sensory Cortex | -54 | -12 | 13 | RH_Default_Temp_3; Right Superior Temporal Gyrus | 57 | -26 | -2 | 3.37 | 0.050 |
| RH_DorsAttn_Post_5; Right Visual Motor Cortex | 14 | -52 | 66 | RH_SalVentAttn_Med_2; Right PreMotor & Supplementary Motor Area | 7 | 6 | 52 | 3.37 | 0.050 |
| RH_Vis_7; Right Visual Association Cortex | 36 | -82 | 16 | RH_Cont_PFCl_2; Right Dorsolateral Prefrontal Cortex | 45 | 39 | 15 | 3.37 | 0.050 |

**Eyes-Closed Theta**

| ROI1 Labels (Schaefer; Talairach) | X | Y | Z | ROI2 Labels (Schaefer; Talairach) | X | Y | Z | t | q |
|---|---|---|---|---|---|---|---|---|---|
| LH_Vis_4; Left Secondary Visual Cortex | -27 | -95 | -4 | RH_Vis_3; Right Fusiform Cortex | 49 | -60 | -11 | 5.35 | 0.003 |
| RH_DorsAttn_Post_1; Right Visual Association Cortex | 50 | -62 | 16 | RH_Limbic_OFC_1; Right Orbitofrontal Cortex | 12 | 35 | -20 | 5.24 | 0.003 |
| LH_Vis_8; Left Visual Association Cortex | -25 | -88 | 20 | LH_Vis_9; Left Visual Association Cortex | -6 | -82 | 26 | 5.18 | 0.003 |
| LH_Vis_6; Left Primary Visual Cortex | -12 | -67 | 7 | RH_Cont_Cing_1; Right Ventral Posterior Cingulate Cortex | 5 | -27 | 33 | 4.94 | 0.006 |
| LH_Vis_4; Left Secondary Visual Cortex | -27 | -95 | -4 | RH_SomMot_2; Right Insula | 40 | -15 | 15 | 4.92 | 0.006 |
| RH_DorsAttn_Post_1; Right Visual Association Cortex | 50 | -62 | 16 | LH_Default_PFC_3; Left Dorsal Posterior Cingulate Cortex | -6 | 47 | 0 | 4.81 | 0.008 |
| LH_Cont_PFCl_1; Left Dorsolateral Prefrontal Cortex | -43 | 33 | 21 | LH_Default_PFC_4; Left Anterior Prefrontal Cortex | -24 | 61 | -1 | 4.68 | 0.012 |
| LH_Vis_8; Left Visual Association Cortex | -25 | -88 | 20 | RH_Limbic_TempPole_1; Right Inferior Temporal Gyrus | 38 | 1 | -38 | 4.65 | 0.012 |
| LH_DorsAttn_Post_1; Left Fusiform Cortex | -47 | -58 | -13 | RH_DorsAttn_PrCv_1; Right Broca's Area/Operculum | 49 | 10 | 27 | 4.62 | 0.012 |
| LH_Vis_6; Left Primary Visual Cortex | -12 | -67 | 7 | RH_SalVentAttn_Med_1; Right Dorsal Posterior Cingulate Cortex | 11 | -31 | 45 | 4.46 | 0.020 |
| LH_Vis_4; Left Secondary Visual Cortex | -27 | -95 | -4 | RH_Default_pCunPCC_2; Right Ventral Posterior Cingulate Cortex | 7 | -52 | 31 | 4.32 | 0.028 |
| LH_DorsAttn_Post_1; Left Fusiform Cortex | -47 | -58 | -13 | RH_Default_pCunPCC_1; Right Ventral Posterior Cingulate Cortex | 12 | -54 | 14 | 4.27 | 0.028 |

| | | | | | | | | | |
|---|---|---|---|---|---|---|---|---|---|
| RH_DorsAttn_Post_1; Right Visual Association Cortex | 50 | -62 | 16 | RH_Default_PFCdPFCm_1; Right Anterior Prefrontal Cortex | 7 | 48 | 1 | 4.26 | 0.028 |
| RH_Default_Par_1; Right Angular Gyrus | 55 | -51 | 31 | RH_Default_PFCdPFCm_1; Right Anterior Prefrontal Cortex | 7 | 48 | 1 | 4.22 | 0.028 |
| RH_SomMot_4; Right Primary Motor Cortex | 58 | -5 | 31 | RH_Default_PFCdPFCm_1; Right Anterior Prefrontal Cortex | 7 | 48 | 1 | 4.21 | 0.028 |
| RH_Cont_PFCl_1; Right Anterior Prefrontal Cortex | 30 | 58 | -3 | RH_Default_Par_1; Right Angular Gyrus | 55 | -51 | 31 | 4.21 | 0.028 |
| RH_Cont_Cing_1; Right Ventral Posterior Cingulate Cortex | 5 | -27 | 33 | LH_Default_pCunPCC_1; Left Ventral Posterior Cingulate Cortex | -11 | -56 | 13 | 4.20 | 0.028 |
| LH_Vis_4; Left Secondary Visual Cortex | -27 | -95 | -4 | LH_Vis_6; Left Primary Visual Cortex | -12 | -67 | 7 | 4.20 | 0.028 |
| LH_DorsAttn_Post_1; Left Fusiform Cortex | -47 | -58 | -13 | RH_Cont_Cing_1; Right Ventral Posterior Cingulate Cortex | 5 | -27 | 33 | 4.18 | 0.028 |
| LH_Vis_8; Left Visual Association Cortex | -25 | -88 | 20 | RH_DorsAttn_Post_1; Right Visual Association Cortex | 50 | -62 | 16 | 4.15 | 0.028 |
| RH_SalVentAttn_TempOccPar_1; Right Superior Temporal Gyrus | 58 | -42 | 13 | RH_Limbic_OFC_1; Right Orbitofrontal Cortex | 12 | 35 | -20 | 4.15 | 0.028 |
| LH_Vis_8; Left Visual Association Cortex | -25 | -88 | 20 | LH_Default_pCunPCC_1; Left Ventral Posterior Cingulate Cortex | -11 | -56 | 13 | 4.12 | 0.028 |
| LH_Vis_8; Left Visual Association Cortex | -25 | -88 | 20 | LH_Default_pCunPCC_2; Left Ventral Posterior Cingulate Cortex | -6 | -53 | 33 | 4.12 | 0.028 |
| RH_DorsAttn_Post_5; Right Visual Motor Cortex | 14 | -52 | 66 | LH_Cont_PFCl_1; Left Dorsolateral Prefrontal Cortex | -43 | 33 | 21 | 4.12 | 0.028 |
| LH_Vis_4; Left Secondary Visual Cortex | -27 | -95 | -4 | RH_Cont_PFCl_4; Right Frontal Eye Fields | 43 | 16 | 45 | 4.11 | 0.028 |
| LH_Vis_4; Left Secondary Visual Cortex | -27 | -95 | -4 | RH_Limbic_TempPole_1; Right Inferior Temporal Gyrus | 38 | 1 | -38 | 4.10 | 0.028 |
| LH_Vis_5; Left Secondary Visual Cortex | -5 | -92 | -2 | RH_Vis_3; Right Fusiform Cortex | 49 | -60 | -11 | 4.09 | 0.028 |
| LH_Vis_9; Left Visual Association Cortex | -6 | -82 | 26 | LH_DorsAttn_Post_1; Left Fusiform Cortex | -47 | -58 | -13 | 4.09 | 0.028 |
| LH_SalVentAttn_Med_3; Left PreMotor & Supplementary Motor Area | -6 | 4 | 62 | RH_Cont_Cing_1; Right Ventral Posterior Cingulate Cortex | 5 | -27 | 33 | 4.08 | 0.028 |
| LH_SalVentAttn_Med_3; Left PreMotor & Supplementary Motor Area | -6 | 4 | 62 | LH_Cont_PFCl_1; Left Dorsolateral Prefrontal Cortex | -43 | 33 | 21 | 4.07 | 0.028 |
| RH_SomMot_2; Right Insula | 40 | -15 | 15 | LH_DorsAttn_Post_4; Left Primary Sensory Cortex | -42 | -34 | 48 | 4.07 | 0.028 |
| LH_Vis_1; Left Fusiform Cortex | -26 | -34 | -17 | LH_Vis_9; Left Visual Association Cortex | -6 | -82 | 26 | 4.06 | 0.028 |
| LH_Vis_4; Left Secondary Visual Cortex | -27 | -95 | -4 | LH_Cont_PFCl_1; Left Dorsolateral Prefrontal Cortex | -43 | 33 | 21 | 4.05 | 0.028 |
| RH_Cont_PFCl_4; Right Frontal Eye Fields | 43 | 16 | 45 | LH_Default_PFC_3; Left Dorsal Posterior Cingulate Cortex | -6 | 47 | 0 | 4.05 | 0.028 |
| LH_Vis_8; Left Visual Association Cortex | -25 | -88 | 20 | RH_Default_pCunPCC_2; Right Ventral Posterior Cingulate Cortex | 7 | -52 | 31 | 4.04 | 0.028 |
| LH_Vis_6; Left Primary Visual Cortex | -12 | -67 | 7 | LH_Vis_8; Left Visual Association Cortex | -25 | -88 | 20 | 4.03 | 0.028 |
| LH_Vis_5; Left Secondary Visual Cortex | -5 | -92 | -2 | RH_DorsAttn_Post_1; Right Visual Association Cortex | 50 | -62 | 16 | 4.02 | 0.029 |
| LH_Vis_9; Left Visual Association Cortex | -6 | -82 | 26 | RH_Vis_2; Right Fusiform Cortex | 27 | -66 | -12 | 4.01 | 0.029 |
| LH_Vis_4; Left Secondary Visual Cortex | -27 | -95 | -4 | RH_DorsAttn_Post_1; Right Visual Association Cortex | 50 | -62 | 16 | 4.00 | 0.029 |
| LH_Vis_8; Left Visual Association Cortex | -25 | -88 | 20 | LH_Default_Temp_2; Left Medial Temporal Gyrus | -58 | -32 | -1 | 4.00 | 0.029 |
| LH_DorsAttn_FEF_1; Left PreMotor & Supplementary Motor Area | -26 | -3 | 59 | LH_Default_Temp_1; Left Medial Temporal Gyrus | -55 | -4 | -20 | 3.99 | 0.029 |
| LH_Vis_4; Left Secondary Visual Cortex | -27 | -95 | -4 | RH_SalVentAttn_TempOccPar_2; Right Supramarginal Gyrus | 61 | -26 | 27 | 3.99 | 0.029 |
| LH_SomMot_2; Left Primary Sensory Cortex | -37 | -21 | 16 | RH_SomMot_2; Right Insula | 40 | -15 | 15 | 3.98 | 0.029 |
| LH_Vis_3; Left Visual Association Cortex | -17 | -60 | -7 | LH_Default_Par_1; Left Angular Gyrus | -57 | -50 | 12 | 3.96 | 0.030 |

| | | | | | | | | | |
|---|---|---|---|---|---|---|---|---|---|
| LH_Vis_4; Left Secondary Visual Cortex | -27 | -95 | -4 | RH_Vis_7; Right Visual Association Cortex | 36 | -82 | 16 | 3.96 | 0.030 |
| LH_Vis_6; Left Primary Visual Cortex | -12 | -67 | 7 | LH_Cont_Cing_1; Left Ventral Posterior Cingulate Cortex | -4 | -26 | 33 | 3.94 | 0.030 |
| LH_Vis_2; Left Visual Association Cortex | -26 | -77 | -14 | LH_Cont_Cing_1; Left Ventral Posterior Cingulate Cortex | -4 | -26 | 33 | 3.94 | 0.030 |
| RH_SomMot_8; Right Primary Motor Cortex | 6 | -26 | 70 | LH_SalVentAttn_Med_3; Left PreMotor & Supplementary Motor Area | -6 | 4 | 62 | 3.94 | 0.030 |
| RH_SomMot_4; Right Primary Motor Cortex | 58 | -5 | 31 | LH_Default_PFC_3; Left Dorsal Posterior Cingulate Cortex | -6 | 47 | 0 | 3.93 | 0.030 |
| LH_Vis_6; Left Primary Visual Cortex | -12 | -67 | 7 | LH_Default_Par_1; Left Angular Gyrus | -57 | -50 | 12 | 3.92 | 0.030 |
| RH_SomMot_3; Right Primary Motor Cortex | 57 | -4 | 11 | RH_Cont_PFCmp_1; Right Frontal Eye Fields | 6 | 28 | 30 | 3.92 | 0.030 |
| LH_Vis_3; Left Visual Association Cortex | -17 | -60 | -7 | LH_Cont_Cing_1; Left Ventral Posterior Cingulate Cortex | -4 | -26 | 33 | 3.90 | 0.031 |
| RH_Vis_5; Right Primary Visual Cortex | 8 | -76 | 5 | RH_DorsAttn_Post_1; Right Visual Association Cortex | 50 | -62 | 16 | 3.90 | 0.031 |
| LH_DorsAttn_Post_4; Left Primary Sensory Cortex | -42 | -34 | 48 | RH_Default_Temp_3; Right Superior Temporal Gyrus | 57 | -26 | -2 | 3.90 | 0.031 |
| RH_Vis_8; Right Visual Association Cortex | 13 | -86 | 29 | LH_DorsAttn_Post_1; Left Fusiform Cortex | -47 | -58 | -13 | 3.89 | 0.031 |
| LH_Cont_PFCl_1; Left Dorsolateral Prefrontal Cortex | -43 | 33 | 21 | RH_Default_Temp_2; Right Temporal Pole | 51 | 7 | -18 | 3.89 | 0.031 |
| LH_Vis_5; Left Secondary Visual Cortex | -5 | -92 | -2 | LH_Vis_6; Left Primary Visual Cortex | -12 | -67 | 7 | 3.87 | 0.032 |
| LH_Vis_4; Left Secondary Visual Cortex | -27 | -95 | -4 | RH_Vis_2; Right Fusiform Cortex | 27 | -66 | -12 | 3.87 | 0.032 |
| LH_Vis_4; Left Secondary Visual Cortex | -27 | -95 | -4 | LH_Vis_9; Left Visual Association Cortex | -6 | -82 | 26 | 3.86 | 0.032 |
| LH_Vis_2; Left Visual Association Cortex | -26 | -77 | -14 | RH_Cont_Cing_1; Right Ventral Posterior Cingulate Cortex | 5 | -27 | 33 | 3.86 | 0.032 |
| RH_DorsAttn_Post_1; Right Visual Association Cortex | 50 | -62 | 16 | RH_Default_PFCv_1; Right Pars Orbitalis | 35 | 26 | -15 | 3.86 | 0.032 |
| RH_Limbic_OFC_1; Right Orbitofrontal Cortex | 12 | 35 | -20 | RH_Default_Par_1; Right Angular Gyrus | 55 | -51 | 31 | 3.85 | 0.032 |
| RH_Vis_3; Right Fusiform Cortex | 49 | -60 | -11 | LH_Default_pCunPCC_1; Left Ventral Posterior Cingulate Cortex | -11 | -56 | 13 | 3.85 | 0.032 |
| LH_DorsAttn_Post_1; Left Fusiform Cortex | -47 | -58 | -13 | LH_Cont_Cing_1; Left Ventral Posterior Cingulate Cortex | -4 | -26 | 33 | 3.84 | 0.032 |
| LH_SalVentAttn_Med_3; Left PreMotor & Supplementary Motor Area | -6 | 4 | 62 | LH_Cont_Cing_1; Left Ventral Posterior Cingulate Cortex | -4 | -26 | 33 | 3.84 | 0.032 |
| LH_Vis_4; Left Secondary Visual Cortex | -27 | -95 | -4 | LH_Default_pCunPCC_1; Left Ventral Posterior Cingulate Cortex | -11 | -56 | 13 | 3.83 | 0.032 |
| RH_Vis_5; Right Primary Visual Cortex | 8 | -76 | 5 | LH_DorsAttn_Post_1; Left Fusiform Cortex | -47 | -58 | -13 | 3.83 | 0.032 |
| LH_Vis_2; Left Visual Association Cortex | -26 | -77 | -14 | LH_Vis_4; Left Secondary Visual Cortex | -27 | -95 | -4 | 3.81 | 0.034 |
| LH_Cont_PFCl_1; Left Dorsolateral Prefrontal Cortex | -43 | 33 | 21 | RH_Default_Temp_3; Right Superior Temporal Gyrus | 57 | -26 | -2 | 3.81 | 0.034 |
| LH_Vis_4; Left Secondary Visual Cortex | -27 | -95 | -4 | RH_Default_Temp_3; Right Superior Temporal Gyrus | 57 | -26 | -2 | 3.80 | 0.034 |
| LH_Vis_2; Left Visual Association Cortex | -26 | -77 | -14 | LH_Default_PFC_2; Left Pars Orbitalis | -47 | 33 | -3 | 3.79 | 0.034 |
| RH_Limbic_OFC_1; Right Orbitofrontal Cortex | 12 | 35 | -20 | RH_Cont_Par_2; Right Angular Gyrus | 45 | -63 | 46 | 3.79 | 0.034 |
| LH_Vis_9; Left Visual Association Cortex | -6 | -82 | 26 | RH_Vis_6; Right Secondary Visual Cortex | 17 | -57 | 5 | 3.78 | 0.034 |
| LH_DorsAttn_Post_1; Left Fusiform Cortex | -47 | -58 | -13 | RH_DorsAttn_Post_1; Right Visual Association Cortex | 50 | -62 | 16 | 3.78 | 0.034 |
| RH_SalVentAttn_TempOccPar_2; Right Supramarginal Gyrus | 61 | -26 | 27 | RH_Cont_PFCl_1; Right Anterior Prefrontal Cortex | 30 | 58 | -3 | 3.78 | 0.034 |

| | | | | | | | | | |
|---|---|---|---|---|---|---|---|---|---|
| LH_Vis_4; Left Secondary Visual Cortex | -27 | -95 | -4 | RH_DorsAttn_PrCv_1; Right Broca's Area/Operculum | 49 | 10 | 27 | 3.77 | 0.035 |
| LH_DorsAttn_Post_1; Left Fusiform Cortex | -47 | -58 | -13 | RH_DorsAttn_Post_4; Right Visual Motor Cortex | 27 | -67 | 51 | 3.77 | 0.035 |
| RH_Vis_3; Right Fusiform Cortex | 49 | -60 | -11 | LH_Limbic_OFC_1; Left Orbitofrontal Cortex | -14 | 32 | -20 | 3.74 | 0.036 |
| LH_DorsAttn_Post_4; Left Primary Sensory Cortex | -42 | -34 | 48 | RH_Default_Temp_1; Right Medial Temporal Gyrus | 62 | -23 | -19 | 3.74 | 0.036 |
| RH_Vis_6; Right Secondary Visual Cortex | 17 | -57 | 5 | LH_Default_PFC_3; Left Dorsal Posterior Cingulate Cortex | -6 | 47 | 0 | 3.74 | 0.036 |
| RH_DorsAttn_Post_4; Right Visual Motor Cortex | 27 | -67 | 51 | RH_Limbic_OFC_1; Right Orbitofrontal Cortex | 12 | 35 | -20 | 3.73 | 0.036 |
| LH_Vis_2; Left Visual Association Cortex | -26 | -77 | -14 | LH_Cont_PFCl_1; Left Dorsolateral Prefrontal Cortex | -43 | 33 | 21 | 3.73 | 0.036 |
| LH_Vis_4; Left Secondary Visual Cortex | -27 | -95 | -4 | RH_Default_Par_1; Right Angular Gyrus | 55 | -51 | 31 | 3.72 | 0.036 |
| LH_Vis_4; Left Secondary Visual Cortex | -27 | -95 | -4 | RH_Vis_5; Right Primary Visual Cortex | 8 | -76 | 5 | 3.72 | 0.036 |
| RH_Vis_2; Right Fusiform Cortex | 27 | -66 | -12 | RH_Vis_8; Right Visual Association Cortex | 13 | -86 | 29 | 3.72 | 0.036 |
| LH_Vis_3; Left Visual Association Cortex | -17 | -60 | -7 | RH_Cont_Cing_1; Right Ventral Posterior Cingulate Cortex | 5 | -27 | 33 | 3.72 | 0.036 |
| RH_Vis_5; Right Primary Visual Cortex | 8 | -76 | 5 | RH_SomMot_2; Right Insula | 40 | -15 | 15 | 3.71 | 0.036 |
| LH_Vis_8; Left Visual Association Cortex | -25 | -88 | 20 | RH_SomMot_1; Right Primary Auditory Cortex | 53 | -16 | 7 | 3.71 | 0.036 |
| LH_Vis_2; Left Visual Association Cortex | -26 | -77 | -14 | LH_Vis_5; Left Secondary Visual Cortex | -5 | -92 | -2 | 3.71 | 0.036 |
| LH_Vis_5; Left Secondary Visual Cortex | -5 | -92 | -2 | RH_Vis_2; Right Fusiform Cortex | 27 | -66 | -12 | 3.71 | 0.036 |
| LH_Vis_4; Left Secondary Visual Cortex | -27 | -95 | -4 | RH_Default_Temp_1; Right Medial Temporal Gyrus | 62 | -23 | -19 | 3.71 | 0.036 |
| RH_Default_Par_1; Right Angular Gyrus | 55 | -51 | 31 | RH_Default_PFCv_1; Right Pars Orbitalis | 35 | 26 | -15 | 3.70 | 0.037 |
| LH_Default_Par_1; Left Angular Gyrus | -57 | -50 | 12 | LH_Default_pCunPCC_1; Left Ventral Posterior Cingulate Cortex | -11 | -56 | 13 | 3.70 | 0.037 |
| LH_Vis_6; Left Primary Visual Cortex | -12 | -67 | 7 | RH_Vis_3; Right Fusiform Cortex | 49 | -60 | -11 | 3.69 | 0.037 |
| LH_Vis_2; Left Visual Association Cortex | -26 | -77 | -14 | LH_Default_Temp_2; Left Medial Temporal Gyrus | -58 | -32 | -1 | 3.69 | 0.037 |
| RH_Cont_PFCl_4; Right Frontal Eye Fields | 43 | 16 | 45 | RH_Cont_PFCmp_1; Right Frontal Eye Fields | 6 | 28 | 30 | 3.69 | 0.037 |
| RH_SalVentAttn_TempOccPar_2; Right Supramarginal Gyrus | 61 | -26 | 27 | RH_Limbic_OFC_1; Right Orbitofrontal Cortex | 12 | 35 | -20 | 3.68 | 0.037 |
| LH_SomMot_6; Left Primary Motor Cortex | -6 | -29 | 70 | RH_Default_pCunPCC_1; Right Ventral Posterior Cingulate Cortex | 12 | -54 | 14 | 3.68 | 0.037 |
| LH_DorsAttn_Post_5; Left Visual Motor Cortex | -6 | -60 | 56 | RH_Limbic_OFC_1; Right Orbitofrontal Cortex | 12 | 35 | -20 | 3.67 | 0.037 |
| LH_Vis_7; Left Visual Association Cortex | -47 | -71 | 11 | LH_Default_Temp_2; Left Medial Temporal Gyrus | -58 | -32 | -1 | 3.67 | 0.037 |
| RH_SalVentAttn_TempOccPar_2; Right Supramarginal Gyrus | 61 | -26 | 27 | RH_Default_PFCdPFCm_1; Right Anterior Prefrontal Cortex | 7 | 48 | 1 | 3.67 | 0.037 |
| LH_Vis_5; Left Secondary Visual Cortex | -5 | -92 | -2 | RH_SomMot_2; Right Insula | 40 | -15 | 15 | 3.66 | 0.038 |
| RH_Cont_PFCl_4; Right Frontal Eye Fields | 43 | 16 | 45 | RH_Default_PFCdPFCm_1; Right Anterior Prefrontal Cortex | 7 | 48 | 1 | 3.66 | 0.038 |
| LH_SomMot_2; Left Primary Sensory Cortex | -37 | -21 | 16 | RH_Default_Temp_1; Right Medial Temporal Gyrus | 62 | -23 | -19 | 3.66 | 0.038 |
| LH_Vis_6; Left Primary Visual Cortex | -12 | -67 | 7 | RH_Default_pCunPCC_2; Right Ventral Posterior Cingulate Cortex | 7 | -52 | 31 | 3.64 | 0.040 |
| RH_SomMot_5; Right PreMotor & Supplementary Motor Area | 47 | -11 | 48 | LH_Default_PFC_3; Left Dorsal Posterior Cingulate Cortex | -6 | 47 | 0 | 3.63 | 0.040 |
| LH_SomMot_2; Left Primary Sensory Cortex | -37 | -21 | 16 | RH_Cont_PFCl_4; Right Frontal Eye Fields | 43 | 16 | 45 | 3.63 | 0.041 |

| | | | | | | | | | |
|---|---|---|---|---|---|---|---|---|---|
| RH_SomMot_2; Right Insula | 40 | -15 | 15 | RH_Default_pCunPCC_1; Right Ventral Posterior Cingulate Cortex | 12 | -54 | 14 | 3.62 | 0.041 |
| LH_DorsAttn_Post_1; Left Fusiform Cortex | -47 | -58 | -13 | LH_DorsAttn_FEF_1; Left PreMotor & Supplementary Motor Area | -26 | -3 | 59 | 3.62 | 0.041 |
| LH_SomMot_2; Left Primary Sensory Cortex | -37 | -21 | 16 | RH_Default_Temp_2; Right Temporal Pole | 51 | 7 | -18 | 3.62 | 0.041 |
| LH_SomMot_2; Left Primary Sensory Cortex | -37 | -21 | 16 | RH_SomMot_4; Right Primary Motor Cortex | 58 | -5 | 31 | 3.62 | 0.041 |
| LH_Cont_Par_1; Left Angular Gyrus | -37 | -53 | 46 | RH_Default_Temp_3; Right Superior Temporal Gyrus | 57 | -26 | -2 | 3.61 | 0.041 |
| LH_Vis_1; Left Fusiform Cortex | -26 | -34 | -17 | LH_Default_pCunPCC_1; Left Ventral Posterior Cingulate Cortex | -11 | -56 | 13 | 3.60 | 0.043 |
| RH_DorsAttn_Post_1; Right Visual Association Cortex | 50 | -62 | 16 | RH_Cont_PFCl_1; Right Anterior Prefrontal Cortex | 30 | 58 | -3 | 3.59 | 0.043 |
| LH_Vis_1; Left Fusiform Cortex | -26 | -34 | -17 | RH_Vis_6; Right Secondary Visual Cortex | 17 | -57 | 5 | 3.59 | 0.043 |
| RH_Cont_Par_1; Right Supramarginal Gyrus | 57 | -39 | 44 | RH_Default_PFCdPFCm_1; Right Anterior Prefrontal Cortex | 7 | 48 | 1 | 3.58 | 0.043 |
| RH_SalVentAttn_TempOccPar_1; Right Superior Temporal Gyrus | 58 | -42 | 13 | RH_Default_PFCdPFCm_1; Right Anterior Prefrontal Cortex | 7 | 48 | 1 | 3.58 | 0.043 |
| LH_Vis_8; Left Visual Association Cortex | -25 | -88 | 20 | RH_Default_Temp_3; Right Superior Temporal Gyrus | 57 | -26 | -2 | 3.58 | 0.043 |
| RH_SomMot_4; Right Primary Motor Cortex | 58 | -5 | 31 | LH_DorsAttn_Post_1; Left Fusiform Cortex | -47 | -58 | -13 | 3.58 | 0.043 |
| RH_DorsAttn_Post_4; Right Visual Motor Cortex | 27 | -67 | 51 | LH_Default_Par_1; Left Angular Gyrus | -57 | -50 | 12 | 3.58 | 0.043 |
| LH_Vis_5; Left Secondary Visual Cortex | -5 | -92 | -2 | RH_SalVentAttn_TempOccPar_1; Right Superior Temporal Gyrus | 58 | -42 | 13 | 3.58 | 0.043 |
| LH_Vis_9; Left Visual Association Cortex | -6 | -82 | 26 | RH_SomMot_2; Right Insula | 40 | -15 | 15 | 3.57 | 0.044 |
| LH_Vis_6; Left Primary Visual Cortex | -12 | -67 | 7 | RH_SomMot_2; Right Insula | 40 | -15 | 15 | 3.57 | 0.044 |
| LH_Vis_9; Left Visual Association Cortex | -6 | -82 | 26 | RH_SomMot_1; Right Primary Auditory Cortex | 53 | -16 | 7 | 3.56 | 0.044 |
| LH_Vis_4; Left Secondary Visual Cortex | -27 | -95 | -4 | RH_SomMot_4; Right Primary Motor Cortex | 58 | -5 | 31 | 3.56 | 0.044 |
| LH_Vis_4; Left Secondary Visual Cortex | -27 | -95 | -4 | RH_Default_PFCdPFCm_3; Right Frontal Eye Fields | 26 | 24 | 50 | 3.55 | 0.045 |
| LH_Vis_8; Left Visual Association Cortex | -25 | -88 | 20 | RH_Cont_Cing_1; Right Ventral Posterior Cingulate Cortex | 5 | -27 | 33 | 3.55 | 0.045 |
| RH_SalVentAttn_TempOccPar_1; Right Superior Temporal Gyrus | 58 | -42 | 13 | RH_Default_PFCv_1; Right Pars Orbitalis | 35 | 26 | -15 | 3.55 | 0.045 |
| LH_Vis_4; Left Secondary Visual Cortex | -27 | -95 | -4 | RH_SomMot_3; Right Primary Motor Cortex | 57 | -4 | 11 | 3.54 | 0.045 |
| LH_Vis_7; Left Visual Association Cortex | -47 | -71 | 11 | RH_DorsAttn_Post_1; Right Visual Association Cortex | 50 | -62 | 16 | 3.53 | 0.045 |
| LH_Cont_PFCl_1; Left Dorsolateral Prefrontal Cortex | -43 | 33 | 21 | RH_Default_Temp_1; Right Medial Temporal Gyrus | 62 | -23 | -19 | 3.53 | 0.045 |
| LH_SomMot_6; Left Primary Motor Cortex | -6 | -29 | 70 | RH_Default_pCunPCC_2; Right Ventral Posterior Cingulate Cortex | 7 | -52 | 31 | 3.53 | 0.045 |
| RH_SomMot_3; Right Primary Motor Cortex | 57 | -4 | 11 | LH_SalVentAttn_Med_1; Left Dorsal Anterior Cingulate Cortex | -5 | 20 | 34 | 3.53 | 0.045 |
| RH_Vis_6; Right Secondary Visual Cortex | 17 | -57 | 5 | LH_DorsAttn_Post_1; Left Fusiform Cortex | -47 | -58 | -13 | 3.53 | 0.045 |
| LH_SomMot_6; Left Primary Motor Cortex | -6 | -29 | 70 | RH_Cont_Cing_1; Right Ventral Posterior Cingulate Cortex | 5 | -27 | 33 | 3.53 | 0.045 |
| RH_Vis_5; Right Primary Visual Cortex | 8 | -76 | 5 | RH_SalVentAttn_TempOccPar_2; Right Supramarginal Gyrus | 61 | -26 | 27 | 3.52 | 0.046 |
| RH_Vis_2; Right Fusiform Cortex | 27 | -66 | -12 | RH_Vis_5; Right Primary Visual Cortex | 8 | -76 | 5 | 3.52 | 0.047 |
| RH_SomMot_6; Right Primary Motor Cortex | 41 | -22 | 60 | RH_SomMot_7; Right Primary Sensory Cortex | 30 | -37 | 64 | 3.51 | 0.048 |
| RH_SomMot_4; Right Primary Motor Cortex | 58 | -5 | 31 | RH_Limbic_OFC_1; Right Orbitofrontal Cortex | 12 | 35 | -20 | 3.50 | 0.048 |

| | | | | | | | | | |
|---|---|---|---|---|---|---|---|---|---|
| RH_SomMot_2; Right Insula | 40 | -15 | 15 | LH_Cont_pCun_1; Left Visual Motor Cortex | -9 | -73 | 38 | 3.50 | 0.048 |
| LH_Vis_8; Left Visual Association Cortex | -25 | -88 | 20 | LH_Cont_Cing_1; Left Ventral Posterior Cingulate Cortex | -4 | -26 | 33 | 3.50 | 0.048 |
| RH_DorsAttn_Post_2; Right Primary Sensory Cortex | 50 | -24 | 42 | RH_Cont_PFCl_1; Right Anterior Prefrontal Cortex | 30 | 58 | -3 | 3.50 | 0.048 |
| RH_SomMot_1; Right Primary Auditory Cortex | 53 | -16 | 7 | LH_DorsAttn_Post_4; Left Primary Sensory Cortex | -42 | -34 | 48 | 3.49 | 0.048 |
| RH_Vis_2; Right Fusiform Cortex | 27 | -66 | -12 | RH_Vis_6; Right Secondary Visual Cortex | 17 | -57 | 5 | 3.49 | 0.048 |
| LH_Vis_2; Left Visual Association Cortex | -26 | -77 | -14 | LH_Vis_8; Left Visual Association Cortex | -25 | -88 | 20 | 3.49 | 0.048 |
| LH_Vis_5; Left Secondary Visual Cortex | -5 | -92 | -2 | RH_Vis_7; Right Visual Association Cortex | 36 | -82 | 16 | 3.49 | 0.048 |
| LH_SalVentAttn_Med_2; Left Dorsal Posterior Cingulate Cortex | -11 | -34 | 45 | LH_Cont_PFCl_1; Left Dorsolateral Prefrontal Cortex | -43 | 33 | 21 | 3.49 | 0.048 |
| LH_Vis_7; Left Visual Association Cortex | -47 | -71 | 11 | RH_Cont_Cing_1; Right Ventral Posterior Cingulate Cortex | 5 | -27 | 33 | 3.48 | 0.049 |
| RH_DorsAttn_Post_1; Right Visual Association Cortex | 50 | -62 | 16 | LH_Cont_PFCl_1; Left Dorsolateral Prefrontal Cortex | -43 | 33 | 21 | 3.48 | 0.049 |
| LH_Vis_1; Left Fusiform Cortex | -26 | -34 | -17 | RH_DorsAttn_PrCv_1; Right Broca's Area/Operculum | 49 | 10 | 27 | 3.47 | 0.049 |
| RH_SalVentAttn_FrOperIns_1; Right Insula | 40 | 8 | 1 | LH_Default_PFC_4; Left Anterior Prefrontal Cortex | -24 | 61 | -1 | 3.47 | 0.049 |
| LH_Vis_9; Left Visual Association Cortex | -6 | -82 | 26 | RH_DorsAttn_Post_1; Right Visual Association Cortex | 50 | -62 | 16 | 3.47 | 0.050 |

q ≤ 0.05 (FDR-corrected)

**Supplementary Table 6 Functional connectivity results for higher menstrual pain participants (n=62) from post-menarche visits 1 to 2**

**Eyes-Closed Theta**

| ROI1 Labels (Schaefer; Talairach) | X | Y | Z | ROI2 Labels (Schaefer; Talairach) | X | Y | Z | t | q |
|---|---|---|---|---|---|---|---|---|---|
| RH_DorsAttn_Post_5; Right Visual Motor Cortex | 14 | -52 | 66 | LH_Cont_PFCl_1; Left Dorsolateral Prefrontal Cortex | -43 | 33 | 21 | 4.78 | 0.047 |
| LH_Vis_4; Left Secondary Visual Cortex | -27 | -95 | -4 | RH_SalVentAttn_TempOccPar_2; Right Supramarginal Gyrus | 61 | -26 | 27 | 4.66 | 0.047 |
| LH_Vis_8; Left Visual Association Cortex | -25 | -88 | 20 | RH_Limbic_TempPole_1; Right Inferior Temporal Gyrus | 38 | 1 | -38 | 4.64 | 0.047 |
| LH_Vis_4; Left Secondary Visual Cortex | -27 | -95 | -4 | RH_SomMot_2; Right Insula | 40 | -15 | 15 | 4.61 | 0.047 |
| LH_Vis_4; Left Secondary Visual Cortex | -27 | -95 | -4 | RH_DorsAttn_Post_5; Right Visual Motor Cortex | 14 | -52 | 66 | 4.54 | 0.047 |
| LH_Vis_4; Left Secondary Visual Cortex | -27 | -95 | -4 | LH_Vis_6; Left Primary Visual Cortex | -12 | -67 | 7 | 4.51 | 0.047 |
| RH_SomMot_2; Right Insula | 40 | -15 | 15 | LH_Cont_pCun_1; Left Visual Motor Cortex | -9 | -73 | 38 | 4.47 | 0.047 |
| LH_Vis_4; Left Secondary Visual Cortex | -27 | -95 | -4 | RH_Default_pCunPCC_2; Right Ventral Posterior Cingulate Cortex | 7 | -52 | 31 | 4.44 | 0.047 |
| LH_Vis_8; Left Visual Association Cortex | -25 | -88 | 20 | RH_Default_Temp_1; Right Medial Temporal Gyrus | 62 | -23 | -19 | 4.38 | 0.049 |
| LH_Vis_8; Left Visual Association Cortex | -25 | -88 | 20 | LH_Vis_9; Left Visual Association Cortex | -6 | -82 | 26 | 4.36 | 0.049 |
| LH_Vis_4; Left Secondary Visual Cortex | -27 | -95 | -4 | LH_DorsAttn_Post_2; Left Supramarginal Gyrus | -57 | -25 | 39 | 4.24 | 0.049 |
| LH_Vis_4; Left Secondary Visual Cortex | -27 | -95 | -4 | LH_Vis_9; Left Visual Association Cortex | -6 | -82 | 26 | 4.24 | 0.049 |
| LH_Vis_4; Left Secondary Visual Cortex | -27 | -95 | -4 | LH_Default_pCunPCC_1; Left Ventral Posterior Cingulate Cortex | -11 | -56 | 13 | 4.24 | 0.049 |
| LH_Cont_PFCl_1; Left Dorsolateral Prefrontal Cortex | -43 | 33 | 21 | LH_Default_PFC_2; Left Pars Orbitalis | -47 | 33 | -3 | 4.23 | 0.049 |
| RH_DorsAttn_FEF_1; Right PreMotor & Supplementary Motor Area | 28 | -3 | 59 | RH_Cont_PFCl_1; Right Anterior Prefrontal Cortex | 30 | 58 | -3 | 4.23 | 0.049 |
| RH_Vis_3; Right Fusiform Cortex | 49 | -60 | -11 | RH_Default_Par_1; Right Angular Gyrus | 55 | -51 | 31 | 4.22 | 0.049 |
| LH_Vis_8; Left Visual Association Cortex | -25 | -88 | 20 | RH_Default_Temp_3; Right Superior Temporal Gyrus | 57 | -26 | -2 | 4.21 | 0.049 |
| LH_Vis_4; Left Secondary Visual Cortex | -27 | -95 | -4 | RH_Vis_3; Right Fusiform Cortex | 49 | -60 | -11 | 4.20 | 0.049 |

q ≤ 0.05 (FDR-corrected)

**Supplementary Table 7 Functional connectivity results for future higher vs. lower menstrual pain groups at the pre-menarche visit**

**Eyes-Open Theta**

| ROI1 Labels (Schaefer; Talairach) | X | Y | Z | ROI2 Labels (Schaefer; Talairach) | X | Y | Z | t | p (unc) |
|---|---|---|---|---|---|---|---|---|---|
| LH_SomMot_1; Left Supramarginal Gyrus | -53 | -23 | 8 | RH_Default_PFCv_1; Right Pars Orbitalis | 35 | 26 | -15 | 3.28 | 0.001 |
| LH_Vis_8; Left Visual Association Cortex | -25 | -88 | 20 | RH_SalVentAttn_FrOperIns_1; Right Insula | 40 | 8 | 1 | -3.04 | 0.003 |
| LH_Vis_1; Left Fusiform Cortex | -26 | -34 | -17 | LH_SomMot_6; Left Primary Motor Cortex | -6 | -29 | 70 | -3.01 | 0.003 |
| RH_Vis_1; Right Fusiform Cortex | 32 | -31 | -22 | LH_DorsAttn_PrCv_1; Left PreMotor & Supplementary Motor Area | -48 | 6 | 28 | 2.95 | 0.004 |
| LH_Vis_2; Left Visual Association Cortex | -26 | -77 | -14 | LH_SomMot_6; Left Primary Motor Cortex | -6 | -29 | 70 | -2.93 | 0.004 |
| RH_Vis_6; Right Secondary Visual Cortex | 17 | -57 | 5 | LH_SalVentAttn_PFCl_1; Left Anterior Prefrontal Cortex | -30 | 44 | 30 | -2.85 | 0.005 |
| LH_Vis_2; Left Visual Association Cortex | -26 | -77 | -14 | RH_DorsAttn_Post_5; Right Visual Motor Cortex | 14 | -52 | 66 | -2.82 | 0.005 |
| LH_SomMot_6; Left Primary Motor Cortex | -6 | -29 | 70 | LH_Default_pCunPCC_1; Left Ventral Posterior Cingulate Cortex | -11 | -56 | 13 | -2.77 | 0.006 |
| LH_Vis_3; Left Visual Association Cortex | -17 | -60 | -7 | LH_SomMot_6; Left Primary Motor Cortex | -6 | -29 | 70 | -2.76 | 0.006 |
| LH_SalVentAttn_Med_1; Left Dorsal Anterior Cingulate Cortex | -5 | 20 | 34 | LH_Cont_PFCl_1; Left Dorsolateral Prefrontal Cortex | -43 | 33 | 21 | -2.71 | 0.007 |
| LH_Vis_8; Left Visual Association Cortex | -25 | -88 | 20 | LH_SalVentAttn_Med_1; Left Dorsal Anterior Cingulate Cortex | -5 | 20 | 34 | -2.70 | 0.008 |
| LH_Vis_1; Left Fusiform Cortex | -26 | -34 | -17 | RH_Default_Temp_3; Right Superior Temporal Gyrus | 57 | -26 | -2 | -2.68 | 0.008 |
| RH_SomMot_5; Right PreMotor & Supplementary Motor Area | 47 | -11 | 48 | LH_Cont_Par_1; Left Angular Gyrus | -37 | -53 | 46 | 2.64 | 0.009 |
| LH_Vis_6; Left Primary Visual Cortex | -12 | -67 | 7 | LH_SomMot_6; Left Primary Motor Cortex | -6 | -29 | 70 | -2.61 | 0.010 |

**Eyes-Open Alpha**

| ROI1 Labels (Schaefer; Talairach) | X | Y | Z | ROI2 Labels (Schaefer; Talairach) | X | Y | Z | t | p (unc) |
|---|---|---|---|---|---|---|---|---|---|
| RH_Vis_2; Right Fusiform Cortex | 27 | -66 | -12 | LH_Default_Par_1; Left Angular Gyrus | -57 | -50 | 12 | -3.89 | 0.0001 |
| RH_SalVentAttn_FrOperIns_1; Right Insula | 40 | 8 | 1 | RH_Limbic_TempPole_1; Right Inferior Temporal Gyrus | 38 | 1 | -38 | 3.57 | 0.0005 |
| RH_Vis_1; Right Fusiform Cortex | 32 | -31 | -22 | LH_Default_Par_1; Left Angular Gyrus | -57 | -50 | 12 | -3.42 | 0.001 |
| RH_SomMot_4; Right Primary Motor Cortex | 58 | -5 | 31 | RH_SalVentAttn_FrOperIns_1; Right Insula | 40 | 8 | 1 | 3.33 | 0.001 |
| LH_SomMot_3; Left Primary Sensory Cortex | -54 | -12 | 13 | RH_SalVentAttn_FrOperIns_1; Right Insula | 40 | 8 | 1 | 3.13 | 0.002 |
| LH_SomMot_3; Left Primary Sensory Cortex | -54 | -12 | 13 | RH_Cont_PFCmp_1; Right Frontal Eye Fields | 6 | 28 | 30 | 3.04 | 0.003 |
| RH_Limbic_TempPole_1; Right Inferior Temporal Gyrus | 38 | 1 | -38 | RH_Default_PFCv_2; Right Broca's Area/ Pars Triangularis | 51 | 28 | 0 | 3.01 | 0.003 |
| LH_Cont_PFCl_1; Left Dorsolateral Prefrontal Cortex | -43 | 33 | 21 | RH_Cont_Par_2; Right Angular Gyrus | 45 | -63 | 46 | -2.89 | 0.004 |
| RH_DorsAttn_Post_3; Right Visual Motor Cortex | 38 | -45 | 49 | RH_SalVentAttn_TempOccPar_2; Right Supramarginal Gyrus | 61 | -26 | 27 | 2.89 | 0.004 |
| LH_SomMot_3; Left Primary Sensory Cortex | -54 | -12 | 13 | RH_Default_PFCv_2; Right Broca's Area/ Pars Triangularis | 51 | 28 | 0 | 2.88 | 0.004 |

| | | | | | | | | | |
|---|---|---|---|---|---|---|---|---|---|
| RH_Vis_3; Right Fusiform Cortex | 49 | -60 | -11 | LH_Default_Par_1; Left Angular Gyrus | -57 | -50 | 12 | -2.82 | 0.005 |
| LH_Vis_7; Left Visual Association Cortex | -47 | -71 | 11 | RH_Vis_2; Right Fusiform Cortex | 27 | -66 | -12 | -2.79 | 0.006 |
| LH_DorsAttn_FEF_1; Left PreMotor & Supplementary Motor Area | -26 | -3 | 59 | RH_Default_Par_1; Right Angular Gyrus | 55 | -51 | 31 | 2.78 | 0.006 |
| LH_Default_Par_2; Left Angular Gyrus | -48 | -63 | 35 | LH_Default_PFC_4; Left Anterior Prefrontal Cortex | -24 | 61 | -1 | -2.77 | 0.006 |
| RH_Limbic_TempPole_1; Right Inferior Temporal Gyrus | 38 | 1 | -38 | RH_Cont_PFCl_3; Right Dorsolateral Prefrontal Cortex | 32 | 46 | 29 | 2.76 | 0.006 |
| LH_SomMot_1; Left Supramarginal Gyrus | -53 | -23 | 8 | RH_SalVentAttn_TempOccPar_1; Right Superior Temporal Gyrus | 58 | -42 | 13 | 2.74 | 0.007 |
| LH_Vis_8; Left Visual Association Cortex | -25 | -88 | 20 | LH_SomMot_6; Left Primary Motor Cortex | -6 | -29 | 70 | -2.73 | 0.007 |
| LH_DorsAttn_Post_6; Left Visual Motor Cortex | -22 | -51 | 66 | RH_SalVentAttn_FrOperIns_1; Right Insula | 40 | 8 | 1 | 2.73 | 0.007 |
| RH_SalVentAttn_TempOccPar_1; Right Superior Temporal Gyrus | 58 | -42 | 13 | RH_SalVentAttn_Med_1; Right Dorsal Posterior Cingulate Cortex | 11 | -31 | 45 | 2.73 | 0.007 |
| LH_DorsAttn_Post_6; Left Visual Motor Cortex | -22 | -51 | 66 | RH_Default_Temp_3; Right Superior Temporal Gyrus | 57 | -26 | -2 | 2.70 | 0.008 |
| RH_SalVentAttn_Med_1; Right Dorsal Posterior Cingulate Cortex | 11 | -31 | 45 | LH_Default_PFC_6; Left PreMotor & Supplementary Motor Area | -41 | 14 | 48 | -2.66 | 0.009 |
| LH_DorsAttn_Post_1; Left Fusiform Cortex | -47 | -58 | -13 | RH_SalVentAttn_FrOperIns_1; Right Insula | 40 | 8 | 1 | 2.65 | 0.009 |
| RH_SomMot_3; Right Primary Motor Cortex | 57 | -4 | 11 | RH_Limbic_TempPole_1; Right Inferior Temporal Gyrus | 38 | 1 | -38 | 2.62 | 0.009 |
| RH_SomMot_5; Right PreMotor & Supplementary Motor Area | 47 | -11 | 48 | LH_Default_PFC_7; Left Frontal Eye Fields | -25 | 20 | 51 | 2.61 | 0.010 |
| RH_SalVentAttn_Med_1; Right Dorsal Posterior Cingulate Cortex | 11 | -31 | 45 | LH_Default_PFC_4; Left Anterior Prefrontal Cortex | -24 | 61 | -1 | 2.61 | 0.010 |
| RH_Limbic_TempPole_1; Right Inferior Temporal Gyrus | 38 | 1 | -38 | RH_Default_PFCv_1; Right Pars Orbitalis | 35 | 26 | -15 | 2.60 | 0.010 |
| RH_SomMot_4; Right Primary Motor Cortex | 58 | -5 | 31 | RH_Default_PFCv_1; Right Pars Orbitalis | 35 | 26 | -15 | 2.59 | 0.010 |

**Eyes-Open Beta**

| ROI1 Labels (Schaefer; Talairach) | X | Y | Z | ROI2 Labels (Schaefer; Talairach) | X | Y | Z | t | p (unc) |
|---|---|---|---|---|---|---|---|---|---|
| LH_DorsAttn_Post_6; Left Visual Motor Cortex | -22 | -51 | 66 | LH_SalVentAttn_ParOper_1; Left Supramarginal Gyrus | -59 | -38 | 29 | 3.66 | 0.0003 |
| LH_Default_PFC_5; Left Dorsolateral Prefrontal Cortex | -9 | 48 | 41 | LH_Default_PFC_7; Left Frontal Eye Fields | -25 | 20 | 51 | 3.60 | 0.0004 |
| RH_DorsAttn_Post_3; Right Visual Motor Cortex | 38 | -45 | 49 | LH_Limbic_TempPole_2; Left Inferior Temporal Gyrus | -57 | -33 | -21 | 3.47 | 0.0007 |
| RH_DorsAttn_Post_2; Right Primary Sensory Cortex | 50 | -24 | 42 | LH_Limbic_TempPole_2; Left Inferior Temporal Gyrus | -57 | -33 | -21 | 3.26 | 0.001 |
| RH_Cont_Cing_1; Right Ventral Posterior Cingulate Cortex | 5 | -27 | 33 | RH_Default_PFCdPFCm_2; Right Dorsolateral Prefrontal Cortex | 11 | 50 | 39 | 3.20 | 0.002 |
| LH_Cont_Cing_1; Left Ventral Posterior Cingulate Cortex | -4 | -26 | 33 | RH_Default_PFCdPFCm_2; Right Dorsolateral Prefrontal Cortex | 11 | 50 | 39 | 3.19 | 0.002 |
| RH_SomMot_6; Right Primary Motor Cortex | 41 | -22 | 60 | LH_Limbic_TempPole_2; Left Inferior Temporal Gyrus | -57 | -33 | -21 | 3.19 | 0.002 |
| RH_SomMot_7; Right Primary Sensory Cortex | 30 | -37 | 64 | LH_Limbic_TempPole_2; Left Inferior Temporal Gyrus | -57 | -33 | -21 | 3.17 | 0.002 |
| RH_Vis_3; Right Fusiform Cortex | 49 | -60 | -11 | RH_SomMot_2; Right Insula | 40 | -15 | 15 | 3.12 | 0.002 |

| | | | | | | | | | |
|---|---|---|---|---|---|---|---|---|---|
| LH_SalVentAttn_FrOperIns_1; Left Insula | -41 | -1 | -7 | RH_Cont_Par_1; Right Supramarginal Gyrus | 57 | -39 | 44 | 2.97 | 0.003 |
| LH_SomMot_3; Left Primary Sensory Cortex | -54 | -12 | 13 | LH_DorsAttn_Post_6; Left Visual Motor Cortex | -22 | -51 | 66 | 2.88 | 0.004 |
| LH_SalVentAttn_FrOperIns_1; Left Insula | -41 | -1 | -7 | LH_SalVentAttn_Med_3; Left PreMotor & Supplementary Motor Area | -6 | 4 | 62 | 2.83 | 0.005 |
| RH_Vis_4; Right Secondary Visual Cortex | 22 | -96 | -5 | LH_Limbic_TempPole_1; Left Temporal Pole | -32 | 2 | -37 | 2.81 | 0.005 |
| RH_DorsAttn_Post_1; Right Visual Association Cortex | 50 | -62 | 16 | LH_Cont_Cing_1; Left Ventral Posterior Cingulate Cortex | -4 | -26 | 33 | 2.78 | 0.006 |
| RH_Cont_Par_1; Right Supramarginal Gyrus | 57 | -39 | 44 | LH_Default_Temp_1; Left Medial Temporal Gyrus | -55 | -4 | -20 | 2.76 | 0.006 |
| LH_SalVentAttn_Med_1; Left Dorsal Anterior Cingulate Cortex | -5 | 20 | 34 | RH_Default_PFCdPFCm_2; Right Dorsolateral Prefrontal Cortex | 11 | 50 | 39 | 2.71 | 0.007 |
| LH_Vis_7; Left Visual Association Cortex | -47 | -71 | 11 | RH_SomMot_6; Right Primary Motor Cortex | 41 | -22 | 60 | 2.71 | 0.007 |
| RH_DorsAttn_Post_4; Right Visual Motor Cortex | 27 | -67 | 51 | LH_Limbic_TempPole_2; Left Inferior Temporal Gyrus | -57 | -33 | -21 | 2.62 | 0.010 |
| LH_SalVentAttn_ParOper_1; Left Supramarginal Gyrus | -59 | -38 | 29 | LH_SalVentAttn_Med_2; Left Dorsal Posterior Cingulate Cortex | -11 | -34 | 45 | 2.62 | 0.010 |
| RH_SalVentAttn_TempOccPar_1; Right Superior Temporal Gyrus | 58 | -42 | 13 | LH_Limbic_TempPole_1; Left Temporal Pole | -32 | 2 | -37 | 2.60 | 0.010 |
| RH_Vis_7; Right Visual Association Cortex | 36 | -82 | 16 | LH_Limbic_TempPole_1; Left Temporal Pole | -32 | 2 | -37 | 2.60 | 0.010 |
| LH_SomMot_1; Left Supramarginal Gyrus | -53 | -23 | 8 | RH_DorsAttn_FEF_1; Right PreMotor & Supplementary Motor Area | 28 | -3 | 59 | 2.60 | 0.010 |
| RH_SomMot_7; Right Primary Sensory Cortex | 30 | -37 | 64 | LH_Default_Temp_1; Left Medial Temporal Gyrus | -55 | -4 | -20 | 2.59 | 0.010 |

**Eyes-Open Gamma**

| ROI1 Labels (Schaefer; Talairach) | X | Y | Z | ROI2 Labels (Schaefer; Talairach) | X | Y | Z | t | p (unc) |
|---|---|---|---|---|---|---|---|---|---|
| LH_Vis_3; Left Visual Association Cortex | -17 | -60 | -7 | LH_SalVentAttn_PFCl_1; Left Anterior Prefrontal Cortex | -30 | 44 | 30 | -4.66 | 6.27E-06 |
| LH_Default_PFC_3; Left Dorsal Posterior Cingulate Cortex | -6 | 47 | 0 | LH_Default_PFC_6; Left PreMotor & Supplementary Motor Area | -41 | 14 | 48 | -4.43 | 1.68E-05 |
| LH_Vis_2; Left Visual Association Cortex | -26 | -77 | -14 | LH_Default_PFC_7; Left Frontal Eye Fields | -25 | 20 | 51 | -3.87 | 0.0002 |
| LH_Vis_1; Left Fusiform Cortex | -26 | -34 | -17 | LH_SalVentAttn_PFCl_1; Left Anterior Prefrontal Cortex | -30 | 44 | 30 | -3.57 | 0.0005 |
| LH_Vis_6; Left Primary Visual Cortex | -12 | -67 | 7 | LH_SalVentAttn_PFCl_1; Left Anterior Prefrontal Cortex | -30 | 44 | 30 | -3.44 | 0.0007 |
| LH_Vis_2; Left Visual Association Cortex | -26 | -77 | -14 | LH_SalVentAttn_Med_1; Left Dorsal Anterior Cingulate Cortex | -5 | 20 | 34 | -3.36 | 0.0009 |
| LH_DorsAttn_Post_4; Left Primary Sensory Cortex | -42 | -34 | 48 | LH_SalVentAttn_PFCl_1; Left Anterior Prefrontal Cortex | -30 | 44 | 30 | -3.35 | 0.0010 |
| LH_Vis_2; Left Visual Association Cortex | -26 | -77 | -14 | LH_SalVentAttn_PFCl_1; Left Anterior Prefrontal Cortex | -30 | 44 | 30 | -3.30 | 0.001 |
| LH_Default_PFC_3; Left Dorsal Posterior Cingulate Cortex | -6 | 47 | 0 | LH_Default_PFC_7; Left Frontal Eye Fields | -25 | 20 | 51 | -3.25 | 0.001 |
| RH_DorsAttn_FEF_1; Right PreMotor & Supplementary Motor Area | 28 | -3 | 59 | LH_SalVentAttn_ParOper_1; Left Supramarginal Gyrus | -59 | -38 | 29 | -3.25 | 0.001 |
| LH_SalVentAttn_PFCl_1; Left Anterior Prefrontal Cortex | -30 | 44 | 30 | LH_Default_pCunPCC_1; Left Ventral Posterior Cingulate Cortex | -11 | -56 | 13 | -3.20 | 0.002 |
| RH_SomMot_5; Right PreMotor & Supplementary Motor Area | 47 | -11 | 48 | LH_Default_Par_2; Left Angular Gyrus | -48 | -63 | 35 | -3.18 | 0.002 |

| | | | | | | | | | |
|---|---|---|---|---|---|---|---|---|---|
| LH_DorsAttn_Post_2; Left Supramarginal Gyrus | -57 | -25 | 39 | RH_Default_PFCv_1; Right Pars Orbitalis | 35 | 26 | -15 | -3.06 | 0.003 |
| RH_SomMot_7; Right Primary Sensory Cortex | 30 | -37 | 64 | RH_SalVentAttn_FrOperIns_1; Right Insula | 40 | 8 | 1 | -3.05 | 0.003 |
| LH_DorsAttn_Post_2; Left Supramarginal Gyrus | -57 | -25 | 39 | LH_Default_PFC_5; Left Dorsolateral Prefrontal Cortex | -9 | 48 | 41 | -3.03 | 0.003 |
| LH_Limbic_TempPole_1; Left Temporal Pole | -32 | 2 | -37 | RH_Default_PFCdPFCm_1; Right Anterior Prefrontal Cortex | 7 | 48 | 1 | 3.00 | 0.003 |
| LH_Vis_6; Left Primary Visual Cortex | -12 | -67 | 7 | LH_Default_PFC_7; Left Frontal Eye Fields | -25 | 20 | 51 | -2.90 | 0.004 |
| LH_DorsAttn_Post_2; Left Supramarginal Gyrus | -57 | -25 | 39 | RH_DorsAttn_FEF_1; Right PreMotor & Supplementary Motor Area | 28 | -3 | 59 | -2.90 | 0.004 |
| LH_DorsAttn_Post_2; Left Supramarginal Gyrus | -57 | -25 | 39 | RH_Default_PFCv_2; Right Broca's Area/ Pars Triangularis | 51 | 28 | 0 | -2.87 | 0.005 |
| LH_Default_PFC_6; Left PreMotor & Supplementary Motor Area | -41 | 14 | 48 | RH_Default_PFCdPFCm_1; Right Anterior Prefrontal Cortex | 7 | 48 | 1 | -2.87 | 0.005 |
| RH_SomMot_2; Right Insula | 40 | -15 | 15 | RH_Default_Temp_3; Right Superior Temporal Gyrus | 57 | -26 | -2 | 2.86 | 0.005 |
| LH_DorsAttn_Post_1; Left Fusiform Cortex | -47 | -58 | -13 | LH_SalVentAttn_PFCl_1; Left Anterior Prefrontal Cortex | -30 | 44 | 30 | -2.82 | 0.005 |
| RH_Vis_6; Right Secondary Visual Cortex | 17 | -57 | 5 | LH_SalVentAttn_PFCl_1; Left Anterior Prefrontal Cortex | -30 | 44 | 30 | -2.81 | 0.005 |
| LH_Vis_4; Left Secondary Visual Cortex | -27 | -95 | -4 | LH_Default_PFC_7; Left Frontal Eye Fields | -25 | 20 | 51 | -2.81 | 0.006 |
| LH_Vis_4; Left Secondary Visual Cortex | -27 | -95 | -4 | RH_Vis_6; Right Secondary Visual Cortex | 17 | -57 | 5 | -2.78 | 0.006 |
| LH_SalVentAttn_Med_1; Left Dorsal Anterior Cingulate Cortex | -5 | 20 | 34 | LH_Default_Par_2; Left Angular Gyrus | -48 | -63 | 35 | -2.75 | 0.007 |
| RH_SalVentAttn_TempOccPar_2; Right Supramarginal Gyrus | 61 | -26 | 27 | LH_Default_Par_2; Left Angular Gyrus | -48 | -63 | 35 | -2.75 | 0.007 |
| LH_Vis_7; Left Visual Association Cortex | -47 | -71 | 11 | LH_SalVentAttn_PFCl_1; Left Anterior Prefrontal Cortex | -30 | 44 | 30 | -2.73 | 0.007 |
| RH_Limbic_OFC_1; Right Orbitofrontal Cortex | 12 | 35 | -20 | LH_Default_Par_2; Left Angular Gyrus | -48 | -63 | 35 | -2.70 | 0.008 |
| RH_Default_Temp_3; Right Superior Temporal Gyrus | 57 | -26 | -2 | RH_Default_PFCv_2; Right Broca's Area/ Pars Triangularis | 51 | 28 | 0 | 2.69 | 0.008 |
| LH_SalVentAttn_ParOper_1; Left Supramarginal Gyrus | -59 | -38 | 29 | RH_Default_PFCdPFCm_1; Right Anterior Prefrontal Cortex | 7 | 48 | 1 | -2.69 | 0.008 |
| LH_Vis_4; Left Secondary Visual Cortex | -27 | -95 | -4 | RH_SomMot_7; Right Primary Sensory Cortex | 30 | -37 | 64 | -2.66 | 0.009 |
| LH_Default_pCunPCC_1; Left Ventral Posterior Cingulate Cortex | -11 | -56 | 13 | RH_Default_Temp_2; Right Temporal Pole | 51 | 7 | -18 | -2.66 | 0.009 |
| RH_Vis_4; Right Secondary Visual Cortex | 22 | -96 | -5 | LH_Cont_Par_1; Left Angular Gyrus | -37 | -53 | 46 | -2.65 | 0.009 |
| LH_Vis_2; Left Visual Association Cortex | -26 | -77 | -14 | RH_SomMot_7; Right Primary Sensory Cortex | 30 | -37 | 64 | -2.64 | 0.009 |

**Eyes-Closed Theta**

| **ROI1 Labels (Schaefer; Talairach)** | **X** | **Y** | **Z** | **ROI2 Labels (Schaefer; Talairach)** | **X** | **Y** | **Z** | **t** | **p (unc)** |
|---|---|---|---|---|---|---|---|---|---|
| LH_Default_pCunPCC_2; Left Ventral Posterior Cingulate Cortex | -6 | -53 | 33 | RH_Default_PFCdPFCm_3; Right Frontal Eye Fields | 26 | 24 | 50 | -3.34 | 0.001 |
| RH_Vis_5; Right Primary Visual Cortex | 8 | -76 | 5 | LH_SalVentAttn_Med_3; Left PreMotor & Supplementary Motor Area | -6 | 4 | 62 | -3.30 | 0.001 |
| RH_Cont_PFCl_2; Right Dorsolateral Prefrontal Cortex | 45 | 39 | 15 | LH_Default_Temp_2; Left Medial Temporal Gyrus | -58 | -32 | -1 | -3.26 | 0.001 |

| | | | | | | | | | |
|---|---|---|---|---|---|---|---|---|---|
| LH_SalVentAttn_Med_3; Left PreMotor & Supplementary Motor Area | -6 | 4 | 62 | RH_Default_pCunPCC_1; Right Ventral Posterior Cingulate Cortex | 12 | -54 | 14 | -3.19 | 0.002 |
| RH_Vis_6; Right Secondary Visual Cortex | 17 | -57 | 5 | LH_SalVentAttn_Med_3; Left PreMotor & Supplementary Motor Area | -6 | 4 | 62 | -3.05 | 0.003 |
| LH_Vis_6; Left Primary Visual Cortex | -12 | -67 | 7 | LH_SalVentAttn_Med_3; Left PreMotor & Supplementary Motor Area | -6 | 4 | 62 | -3.03 | 0.003 |
| LH_SomMot_3; Left Primary Sensory Cortex | -54 | -12 | 13 | RH_Cont_PFCl_2; Right Dorsolateral Prefrontal Cortex | 45 | 39 | 15 | -2.87 | 0.005 |
| RH_Cont_PFCl_2; Right Dorsolateral Prefrontal Cortex | 45 | 39 | 15 | LH_Default_pCunPCC_1; Left Ventral Posterior Cingulate Cortex | -11 | -56 | 13 | -2.85 | 0.005 |
| RH_Vis_2; Right Fusiform Cortex | 27 | -66 | -12 | LH_Default_Temp_1; Left Medial Temporal Gyrus | -55 | -4 | -20 | -2.74 | 0.007 |
| LH_DorsAttn_FEF_1; Left PreMotor & Supplementary Motor Area | -26 | -3 | 59 | LH_Default_pCunPCC_1; Left Ventral Posterior Cingulate Cortex | -11 | -56 | 13 | -2.71 | 0.007 |
| LH_SomMot_1; Left Supramarginal Gyrus | -53 | -23 | 8 | RH_Cont_PFCl_2; Right Dorsolateral Prefrontal Cortex | 45 | 39 | 15 | -2.70 | 0.008 |
| RH_Default_PFCdPFCm_3; Right Frontal Eye Fields | 26 | 24 | 50 | RH_Default_pCunPCC_2; Right Ventral Posterior Cingulate Cortex | 7 | -52 | 31 | -2.67 | 0.008 |
| LH_Cont_Par_1; Left Angular Gyrus | -37 | -53 | 46 | RH_Cont_PFCl_2; Right Dorsolateral Prefrontal Cortex | 45 | 39 | 15 | -2.66 | 0.009 |

**Eyes-Closed Alpha**

| ROI1 Labels (Schaefer; Talairach) | X | Y | Z | ROI2 Labels (Schaefer; Talairach) | X | Y | Z | t | p (unc) |
|---|---|---|---|---|---|---|---|---|---|
| LH_Vis_8; Left Visual Association Cortex | -25 | -88 | 20 | RH_SomMot_4; Right Primary Motor Cortex | 58 | -5 | 31 | 3.53 | 0.0005 |
| LH_Cont_PFCl_1; Left Dorsolateral Prefrontal Cortex | -43 | 33 | 21 | LH_Default_PFC_2; Left Pars Orbitalis | -47 | 33 | -3 | 2.93 | 0.004 |
| LH_Vis_5; Left Secondary Visual Cortex | -5 | -92 | -2 | LH_SalVentAttn_Med_2; Left Dorsal Posterior Cingulate Cortex | -11 | -34 | 45 | 2.89 | 0.004 |
| LH_Vis_6; Left Primary Visual Cortex | -12 | -67 | 7 | LH_DorsAttn_Post_5; Left Visual Motor Cortex | -6 | -60 | 56 | 2.79 | 0.006 |
| RH_Vis_3; Right Fusiform Cortex | 49 | -60 | -11 | LH_DorsAttn_Post_2; Left Supramarginal Gyrus | -57 | -25 | 39 | 2.75 | 0.007 |
| LH_SomMot_4; Left PreMotor & Supplementary Motor Area | -55 | -8 | 34 | RH_Limbic_TempPole_1; Right Inferior Temporal Gyrus | 38 | 1 | -38 | -2.75 | 0.007 |
| LH_SalVentAttn_Med_1; Left Dorsal Anterior Cingulate Cortex | -5 | 20 | 34 | RH_Default_PFCv_1; Right Pars Orbitalis | 35 | 26 | -15 | -2.72 | 0.007 |
| RH_Vis_7; Right Visual Association Cortex | 36 | -82 | 16 | LH_SalVentAttn_PFCl_1; Left Anterior Prefrontal Cortex | -30 | 44 | 30 | -2.71 | 0.007 |
| LH_SalVentAttn_Med_2; Left Dorsal Posterior Cingulate Cortex | -11 | -34 | 45 | LH_Default_pCunPCC_2; Left Ventral Posterior Cingulate Cortex | -6 | -53 | 33 | 2.71 | 0.007 |
| RH_Vis_7; Right Visual Association Cortex | 36 | -82 | 16 | LH_Default_Par_1; Left Angular Gyrus | -57 | -50 | 12 | -2.64 | 0.009 |
| RH_Limbic_TempPole_1; Right Inferior Temporal Gyrus | 38 | 1 | -38 | RH_Cont_Cing_1; Right Ventral Posterior Cingulate Cortex | 5 | -27 | 33 | -2.61 | 0.010 |
| LH_SomMot_3; Left Primary Sensory Cortex | -54 | -12 | 13 | LH_SalVentAttn_ParOper_1; Left Supramarginal Gyrus | -59 | -38 | 29 | 2.59 | 0.010 |

**Eyes-Closed Beta**

| ROI1 Labels (Schaefer; Talairach) | X | Y | Z | ROI2 Labels (Schaefer; Talairach) | X | Y | Z | t | p (unc) |
|---|---|---|---|---|---|---|---|---|---|
| LH_DorsAttn_Post_3; Left Visual Motor Cortex | -24 | -68 | 49 | RH_DorsAttn_Post_3; Right Visual Motor Cortex | 38 | -45 | 49 | 4.11 | 6.03E-05 |
| LH_DorsAttn_Post_3; Left Visual Motor Cortex | -24 | -68 | 49 | RH_DorsAttn_Post_4; Right Visual Motor Cortex | 27 | -67 | 51 | 4.01 | 8.95E- |

| | | | | | | | | | | |
|---|---|---|---|---|---|---|---|---|---|---|
| | | | | | | | | | | 05 |
| LH_SalVentAttn_Med_2; Left Dorsal Posterior Cingulate Cortex | -11 | -34 | 45 | RH_Cont_Cing_1; Right Ventral Posterior Cingulate Cortex | 5 | -27 | 33 | 3.71 | 0.0003 |
| LH_DorsAttn_Post_3; Left Visual Motor Cortex | -24 | -68 | 49 | RH_Cont_Par_2; Right Angular Gyrus | 45 | -63 | 46 | 3.58 | 0.0004 |
| LH_DorsAttn_Post_3; Left Visual Motor Cortex | -24 | -68 | 49 | RH_SalVentAttn_TempOccPar_1; Right Superior Temporal Gyrus | 58 | -42 | 13 | 3.42 | 0.0008 |
| RH_Cont_Par_2; Right Angular Gyrus | 45 | -63 | 46 | RH_Default_PFCdPFCm_1; Right Anterior Prefrontal Cortex | 7 | 48 | 1 | 3.32 | 0.001 |
| LH_DorsAttn_Post_3; Left Visual Motor Cortex | -24 | -68 | 49 | RH_DorsAttn_Post_1; Right Visual Association Cortex | 50 | -62 | 16 | 3.29 | 0.001 |
| RH_SomMot_8; Right Primary Motor Cortex | 6 | -26 | 70 | RH_SalVentAttn_Med_1; Right Dorsal Posterior Cingulate Cortex | 11 | -31 | 45 | 3.26 | 0.001 |
| LH_DorsAttn_Post_6; Left Visual Motor Cortex | -22 | -51 | 66 | RH_DorsAttn_Post_3; Right Visual Motor Cortex | 38 | -45 | 49 | 3.13 | 0.002 |
| LH_DorsAttn_FEF_1; Left PreMotor & Supplementary Motor Area | -26 | -3 | 59 | RH_Cont_Cing_1; Right Ventral Posterior Cingulate Cortex | 5 | -27 | 33 | 3.12 | 0.002 |
| LH_SomMot_6; Left Primary Motor Cortex | -6 | -29 | 70 | RH_Default_pCunPCC_2; Right Ventral Posterior Cingulate Cortex | 7 | -52 | 31 | 3.12 | 0.002 |
| LH_Vis_4; Left Secondary Visual Cortex | -27 | -95 | -4 | RH_Default_Temp_1; Right Medial Temporal Gyrus | 62 | -23 | -19 | 3.10 | 0.002 |
| LH_Vis_4; Left Secondary Visual Cortex | -27 | -95 | -4 | RH_Default_PFCv_2; Right Broca's Area/ Pars Triangularis | 51 | 28 | 0 | 3.08 | 0.002 |
| LH_DorsAttn_Post_3; Left Visual Motor Cortex | -24 | -68 | 49 | RH_Default_Par_1; Right Angular Gyrus | 55 | -51 | 31 | 3.08 | 0.002 |
| LH_DorsAttn_Post_3; Left Visual Motor Cortex | -24 | -68 | 49 | RH_Cont_Par_1; Right Supramarginal Gyrus | 57 | -39 | 44 | 3.06 | 0.003 |
| RH_Vis_7; Right Visual Association Cortex | 36 | -82 | 16 | LH_DorsAttn_Post_3; Left Visual Motor Cortex | -24 | -68 | 49 | 3.05 | 0.003 |
| LH_DorsAttn_Post_6; Left Visual Motor Cortex | -22 | -51 | 66 | LH_Default_pCunPCC_2; Left Ventral Posterior Cingulate Cortex | -6 | -53 | 33 | 3.04 | 0.003 |
| RH_SomMot_8; Right Primary Motor Cortex | 6 | -26 | 70 | RH_Cont_Cing_1; Right Ventral Posterior Cingulate Cortex | 5 | -27 | 33 | 3.03 | 0.003 |
| LH_DorsAttn_Post_3; Left Visual Motor Cortex | -24 | -68 | 49 | RH_SalVentAttn_Med_1; Right Dorsal Posterior Cingulate Cortex | 11 | -31 | 45 | 3.02 | 0.003 |
| RH_DorsAttn_Post_3; Right Visual Motor Cortex | 38 | -45 | 49 | LH_Cont_Par_1; Left Angular Gyrus | -37 | -53 | 46 | 3.02 | 0.003 |
| LH_DorsAttn_Post_3; Left Visual Motor Cortex | -24 | -68 | 49 | RH_DorsAttn_Post_5; Right Visual Motor Cortex | 14 | -52 | 66 | 3.01 | 0.003 |
| LH_DorsAttn_FEF_1; Left PreMotor & Supplementary Motor Area | -26 | -3 | 59 | RH_SalVentAttn_Med_1; Right Dorsal Posterior Cingulate Cortex | 11 | -31 | 45 | 3.01 | 0.003 |
| LH_SalVentAttn_FrOperIns_1; Left Insula | -41 | -1 | -7 | LH_SalVentAttn_PFCl_1; Left Anterior Prefrontal Cortex | -30 | 44 | 30 | 2.97 | 0.003 |
| RH_Vis_6; Right Secondary Visual Cortex | 17 | -57 | 5 | LH_DorsAttn_FEF_1; Left PreMotor & Supplementary Motor Area | -26 | -3 | 59 | 2.93 | 0.004 |
| LH_Vis_4; Left Secondary Visual Cortex | -27 | -95 | -4 | RH_Default_Temp_3; Right Superior Temporal Gyrus | 57 | -26 | -2 | 2.92 | 0.004 |
| LH_Cont_Par_1; Left Angular Gyrus | -37 | -53 | 46 | RH_Cont_Par_2; Right Angular Gyrus | 45 | -63 | 46 | 2.92 | 0.004 |
| LH_DorsAttn_FEF_1; Left PreMotor & Supplementary Motor Area | -26 | -3 | 59 | RH_Default_pCunPCC_1; Right Ventral Posterior Cingulate Cortex | 12 | -54 | 14 | 2.92 | 0.004 |
| RH_Vis_5; Right Primary Visual Cortex | 8 | -76 | 5 | RH_SomMot_8; Right Primary Motor Cortex | 6 | -26 | 70 | 2.89 | 0.004 |
| RH_Cont_Par_2; Right Angular Gyrus | 45 | -63 | 46 | LH_Default_PFC_3; Left Dorsal Posterior Cingulate Cortex | -6 | 47 | 0 | 2.89 | 0.004 |

| | | | | | | | | | |
|---|---|---|---|---|---|---|---|---|---|
| LH_Vis_5; Left Secondary Visual Cortex | -5 | -92 | -2 | RH_Default_Temp_3; Right Superior Temporal Gyrus | 57 | -26 | -2 | 2.89 | 0.004 |
| LH_SomMot_6; Left Primary Motor Cortex | -6 | -29 | 70 | LH_SalVentAttn_Med_2; Left Dorsal Posterior Cingulate Cortex | -11 | -34 | 45 | 2.87 | 0.005 |
| LH_Vis_5; Left Secondary Visual Cortex | -5 | -92 | -2 | RH_SomMot_8; Right Primary Motor Cortex | 6 | -26 | 70 | 2.87 | 0.005 |
| RH_Vis_5; Right Primary Visual Cortex | 8 | -76 | 5 | LH_SomMot_6; Left Primary Motor Cortex | -6 | -29 | 70 | 2.87 | 0.005 |
| LH_Vis_5; Left Secondary Visual Cortex | -5 | -92 | -2 | LH_SomMot_6; Left Primary Motor Cortex | -6 | -29 | 70 | 2.86 | 0.005 |
| LH_DorsAttn_Post_6; Left Visual Motor Cortex | -22 | -51 | 66 | RH_Default_pCunPCC_2; Right Ventral Posterior Cingulate Cortex | 7 | -52 | 31 | 2.85 | 0.005 |
| LH_DorsAttn_Post_5; Left Visual Motor Cortex | -6 | -60 | 56 | RH_DorsAttn_Post_5; Right Visual Motor Cortex | 14 | -52 | 66 | 2.84 | 0.005 |
| RH_DorsAttn_Post_4; Right Visual Motor Cortex | 27 | -67 | 51 | LH_Cont_pCun_1; Left Visual Motor Cortex | -9 | -73 | 38 | 2.84 | 0.005 |
| RH_SomMot_4; Right Primary Motor Cortex | 58 | -5 | 31 | RH_SomMot_6; Right Primary Motor Cortex | 41 | -22 | 60 | -2.79 | 0.006 |
| LH_DorsAttn_Post_5; Left Visual Motor Cortex | -6 | -60 | 56 | RH_Default_pCunPCC_2; Right Ventral Posterior Cingulate Cortex | 7 | -52 | 31 | 2.76 | 0.006 |
| LH_Vis_8; Left Visual Association Cortex | -25 | -88 | 20 | RH_Default_Temp_1; Right Medial Temporal Gyrus | 62 | -23 | -19 | 2.76 | 0.006 |
| LH_SomMot_4; Left PreMotor & Supplementary Motor Area | -55 | -8 | 34 | RH_DorsAttn_FEF_1; Right PreMotor & Supplementary Motor Area | 28 | -3 | 59 | 2.75 | 0.007 |
| LH_Cont_Cing_1; Left Ventral Posterior Cingulate Cortex | -4 | -26 | 33 | RH_Default_PFCv_1; Right Pars Orbitalis | 35 | 26 | -15 | 2.74 | 0.007 |
| RH_SalVentAttn_Med_2; Right PreMotor & Supplementary Motor Area | 7 | 6 | 52 | LH_Default_Temp_1; Left Medial Temporal Gyrus | -55 | -4 | -20 | 2.73 | 0.007 |
| LH_DorsAttn_PrCv_1; Left PreMotor & Supplementary Motor Area | -48 | 6 | 28 | RH_Cont_Cing_1; Right Ventral Posterior Cingulate Cortex | 5 | -27 | 33 | 2.73 | 0.007 |
| RH_Default_PFCv_1; Right Pars Orbitalis | 35 | 26 | -15 | RH_Default_pCunPCC_2; Right Ventral Posterior Cingulate Cortex | 7 | -52 | 31 | 2.71 | 0.007 |
| LH_SalVentAttn_Med_2; Left Dorsal Posterior Cingulate Cortex | -11 | -34 | 45 | LH_Cont_Cing_1; Left Ventral Posterior Cingulate Cortex | -4 | -26 | 33 | 2.70 | 0.008 |
| LH_Vis_5; Left Secondary Visual Cortex | -5 | -92 | -2 | LH_SomMot_1; Left Supramarginal Gyrus | -53 | -23 | 8 | 2.69 | 0.008 |
| LH_Vis_6; Left Primary Visual Cortex | -12 | -67 | 7 | LH_SalVentAttn_Med_3; Left PreMotor & Supplementary Motor Area | -6 | 4 | 62 | 2.69 | 0.008 |
| LH_DorsAttn_Post_6; Left Visual Motor Cortex | -22 | -51 | 66 | RH_Cont_pCun_1; Right Visual Motor Cortex | 9 | -66 | 43 | 2.69 | 0.008 |
| LH_SomMot_1; Left Supramarginal Gyrus | -53 | -23 | 8 | LH_SalVentAttn_Med_3; Left PreMotor & Supplementary Motor Area | -6 | 4 | 62 | 2.68 | 0.008 |
| RH_Vis_4; Right Secondary Visual Cortex | 22 | -96 | -5 | RH_Default_Temp_3; Right Superior Temporal Gyrus | 57 | -26 | -2 | 2.68 | 0.008 |
| RH_SomMot_8; Right Primary Motor Cortex | 6 | -26 | 70 | RH_Default_pCunPCC_2; Right Ventral Posterior Cingulate Cortex | 7 | -52 | 31 | 2.67 | 0.008 |
| LH_SomMot_2; Left Primary Sensory Cortex | -37 | -21 | 16 | LH_SalVentAttn_Med_3; Left PreMotor & Supplementary Motor Area | -6 | 4 | 62 | 2.67 | 0.008 |
| LH_DorsAttn_PrCv_1; Left PreMotor & Supplementary Motor Area | -48 | 6 | 28 | RH_SalVentAttn_Med_1; Right Dorsal Posterior Cingulate Cortex | 11 | -31 | 45 | 2.66 | 0.008 |
| LH_SomMot_1; Left Supramarginal Gyrus | -53 | -23 | 8 | LH_Default_Temp_1; Left Medial Temporal Gyrus | -55 | -4 | -20 | 2.66 | 0.008 |
| RH_DorsAttn_Post_3; Right Visual Motor Cortex | 38 | -45 | 49 | LH_Cont_pCun_1; Left Visual Motor Cortex | -9 | -73 | 38 | 2.66 | 0.009 |
| LH_DorsAttn_FEF_1; Left PreMotor & Supplementary Motor Area | -26 | -3 | 59 | RH_Default_pCunPCC_2; Right Ventral Posterior Cingulate Cortex | 7 | -52 | 31 | 2.66 | 0.009 |

| | | | | | | | | | |
|---|---|---|---|---|---|---|---|---|---|
| LH_SalVentAttn_Med_3; Left PreMotor & Supplementary Motor Area | -6 | 4 | 62 | RH_Default_PFCv_1; Right Pars Orbitalis | 35 | 26 | -15 | 2.65 | 0.009 |
| LH_Vis_8; Left Visual Association Cortex | -25 | -88 | 20 | RH_SalVentAttn_Med_1; Right Dorsal Posterior Cingulate Cortex | 11 | -31 | 45 | 2.64 | 0.009 |
| RH_Vis_5; Right Primary Visual Cortex | 8 | -76 | 5 | LH_SalVentAttn_Med_3; Left PreMotor & Supplementary Motor Area | -6 | 4 | 62 | 2.64 | 0.009 |
| RH_DorsAttn_Post_3; Right Visual Motor Cortex | 38 | -45 | 49 | RH_Default_pCunPCC_2; Right Ventral Posterior Cingulate Cortex | 7 | -52 | 31 | 2.63 | 0.009 |
| LH_SomMot_6; Left Primary Motor Cortex | -6 | -29 | 70 | LH_Default_pCunPCC_1; Left Ventral Posterior Cingulate Cortex | -11 | -56 | 13 | 2.63 | 0.009 |
| LH_SomMot_5; Left Primary Motor Cortex | -39 | -23 | 59 | LH_DorsAttn_Post_3; Left Visual Motor Cortex | -24 | -68 | 49 | 2.62 | 0.010 |
| LH_SalVentAttn_Med_3; Left PreMotor & Supplementary Motor Area | -6 | 4 | 62 | RH_Default_pCunPCC_2; Right Ventral Posterior Cingulate Cortex | 7 | -52 | 31 | 2.61 | 0.010 |
| LH_SalVentAttn_Med_3; Left PreMotor & Supplementary Motor Area | -6 | 4 | 62 | LH_Default_pCunPCC_2; Left Ventral Posterior Cingulate Cortex | -6 | -53 | 33 | 2.60 | 0.010 |
| LH_Vis_9; Left Visual Association Cortex | -6 | -82 | 26 | RH_DorsAttn_Post_4; Right Visual Motor Cortex | 27 | -67 | 51 | 2.60 | 0.010 |
| LH_DorsAttn_Post_5; Left Visual Motor Cortex | -6 | -60 | 56 | RH_SalVentAttn_Med_1; Right Dorsal Posterior Cingulate Cortex | 11 | -31 | 45 | 2.60 | 0.010 |
| LH_SomMot_6; Left Primary Motor Cortex | -6 | -29 | 70 | LH_Default_pCunPCC_2; Left Ventral Posterior Cingulate Cortex | -6 | -53 | 33 | 2.59 | 0.010 |
| LH_DorsAttn_Post_6; Left Visual Motor Cortex | -22 | -51 | 66 | RH_Cont_Par_1; Right Supramarginal Gyrus | 57 | -39 | 44 | 2.59 | 0.010 |

**Eyes-Closed Gamma**

| ROI1 Labels (Schaefer; Talairach) | X | Y | Z | ROI2 Labels (Schaefer; Talairach) | X | Y | Z | t | p (unc) |
|---|---|---|---|---|---|---|---|---|---|
| LH_SomMot_3; Left Primary Sensory Cortex | -54 | -12 | 13 | LH_SalVentAttn_FrOperIns_2; Left Insula | -38 | 12 | 6 | 3.90 | 0.0001 |
| LH_SomMot_3; Left Primary Sensory Cortex | -54 | -12 | 13 | LH_Default_PFC_1; Left Pars Orbitalis | -35 | 21 | -11 | 3.77 | 0.0002 |
| LH_SomMot_3; Left Primary Sensory Cortex | -54 | -12 | 13 | LH_Default_PFC_2; Left Pars Orbitalis | -47 | 33 | -3 | 3.47 | 0.0007 |
| LH_SomMot_1; Left Supramarginal Gyrus | -53 | -23 | 8 | LH_Default_PFC_1; Left Pars Orbitalis | -35 | 21 | -11 | 3.32 | 0.001 |
| LH_SalVentAttn_ParOper_1; Left Supramarginal Gyrus | -59 | -38 | 29 | LH_Default_PFC_1; Left Pars Orbitalis | -35 | 21 | -11 | 3.31 | 0.001 |
| LH_SomMot_1; Left Supramarginal Gyrus | -53 | -23 | 8 | RH_Default_Temp_2; Right Temporal Pole | 51 | 7 | -18 | 3.16 | 0.002 |
| LH_SomMot_1; Left Supramarginal Gyrus | -53 | -23 | 8 | LH_SalVentAttn_FrOperIns_2; Left Insula | -38 | 12 | 6 | 3.15 | 0.002 |
| LH_SalVentAttn_FrOperIns_2; Left Insula | -38 | 12 | 6 | LH_Default_PFC_2; Left Pars Orbitalis | -47 | 33 | -3 | 3.10 | 0.002 |
| LH_Cont_PFCl_1; Left Dorsolateral Prefrontal Cortex | -43 | 33 | 21 | RH_Cont_PFCl_1; Right Anterior Prefrontal Cortex | 30 | 58 | -3 | -3.08 | 0.002 |
| RH_Cont_Par_2; Right Angular Gyrus | 45 | -63 | 46 | LH_Default_pCunPCC_1; Left Ventral Posterior Cingulate Cortex | -11 | -56 | 13 | -3.05 | 0.003 |
| LH_SalVentAttn_ParOper_1; Left Supramarginal Gyrus | -59 | -38 | 29 | LH_SalVentAttn_FrOperIns_2; Left Insula | -38 | 12 | 6 | 3.04 | 0.003 |
| RH_SomMot_5; Right PreMotor & Supplementary Motor Area | 47 | -11 | 48 | RH_DorsAttn_Post_5; Right Visual Motor Cortex | 14 | -52 | 66 | -2.96 | 0.004 |
| RH_Vis_6; Right Secondary Visual Cortex | 17 | -57 | 5 | LH_DorsAttn_Post_6; Left Visual Motor Cortex | -22 | -51 | 66 | -2.86 | 0.005 |
| LH_Vis_6; Left Primary Visual Cortex | -12 | -67 | 7 | RH_Default_Temp_1; Right Medial Temporal Gyrus | 62 | -23 | -19 | -2.85 | 0.005 |

| | | | | | | | | | |
|---|---|---|---|---|---|---|---|---|---|
| LH_Vis_6; Left Primary Visual Cortex | -12 | -67 | 7 | LH_SomMot_5; Left Primary Motor Cortex | -39 | -23 | 59 | -2.82 | 0.005 |
| LH_Default_PFC_2; Left Pars Orbitalis | -47 | 33 | -3 | LH_Default_PFC_4; Left Anterior Prefrontal Cortex | -24 | 61 | -1 | -2.77 | 0.006 |
| LH_Default_Temp_1; Left Medial Temporal Gyrus | -55 | -4 | -20 | LH_Default_PFC_1; Left Pars Orbitalis | -35 | 21 | -11 | 2.76 | 0.006 |
| RH_SalVentAttn_Med_2; Right PreMotor & Supplementary Motor Area | 7 | 6 | 52 | RH_Cont_Cing_1; Right Ventral Posterior Cingulate Cortex | 5 | -27 | 33 | -2.75 | 0.007 |
| RH_Vis_3; Right Fusiform Cortex | 49 | -60 | -11 | RH_Cont_pCun_1; Right Visual Motor Cortex | 9 | -66 | 43 | -2.75 | 0.007 |
| RH_Vis_1; Right Fusiform Cortex | 32 | -31 | -22 | RH_Limbic_OFC_1; Right Orbitofrontal Cortex | 12 | 35 | -20 | -2.72 | 0.007 |
| LH_DorsAttn_Post_5; Left Visual Motor Cortex | -6 | -60 | 56 | RH_Default_Par_1; Right Angular Gyrus | 55 | -51 | 31 | -2.69 | 0.008 |
| LH_Default_Par_2; Left Angular Gyrus | -48 | -63 | 35 | RH_Default_pCunPCC_1; Right Ventral Posterior Cingulate Cortex | 12 | -54 | 14 | 2.68 | 0.008 |
| LH_DorsAttn_FEF_1; Left PreMotor & Supplementary Motor Area | -26 | -3 | 59 | RH_Default_Temp_2; Right Temporal Pole | 51 | 7 | -18 | 2.68 | 0.008 |
| RH_Vis_5; Right Primary Visual Cortex | 8 | -76 | 5 | LH_DorsAttn_Post_6; Left Visual Motor Cortex | -22 | -51 | 66 | -2.66 | 0.009 |
| LH_SomMot_2; Left Primary Sensory Cortex | -37 | -21 | 16 | LH_Default_PFC_2; Left Pars Orbitalis | -47 | 33 | -3 | 2.65 | 0.009 |
| LH_Limbic_TempPole_2; Left Inferior Temporal Gyrus | -57 | -33 | -21 | LH_Default_Par_2; Left Angular Gyrus | -48 | -63 | 35 | 2.62 | 0.010 |
| RH_Vis_5; Right Primary Visual Cortex | 8 | -76 | 5 | RH_Default_Par_1; Right Angular Gyrus | 55 | -51 | 31 | -2.62 | 0.010 |
| LH_SomMot_2; Left Primary Sensory Cortex | -37 | -21 | 16 | LH_SalVentAttn_FrOperIns_2; Left Insula | -38 | 12 | 6 | 2.61 | 0.010 |
| LH_SomMot_3; Left Primary Sensory Cortex | -54 | -12 | 13 | LH_Cont_PFCl_1; Left Dorsolateral Prefrontal Cortex | -43 | 33 | 21 | 2.60 | 0.010 |

$p \leq 0.01$ uncorrected

**Supplementary Table 8 Functional connectivity results for higher vs. lower menstrual pain groups at post-menarche visit 1**

**Eyes-Open Theta**

| ROI1 Labels (Schaefer; Talairach) | X | Y | Z | ROI2 Labels (Schaefer; Talairach) | X | Y | Z | t | p (unc) |
|---|---|---|---|---|---|---|---|---|---|
| LH_Default_Temp_1; Left Medial Temporal Gyrus | -55 | -4 | -20 | LH_Default_PFC_1; Left Pars Orbitalis | -35 | 21 | -11 | 3.10 | 0.002 |
| LH_Default_Par_1; Left Angular Gyrus | -57 | -50 | 12 | LH_Default_pCunPCC_1; Left Ventral Posterior Cingulate Cortex | -11 | -56 | 13 | 2.93 | 0.004 |
| LH_Limbic_OFC_1; Left Orbitofrontal Cortex | -14 | 32 | -20 | RH_Limbic_TempPole_1; Right Inferior Temporal Gyrus | 38 | 1 | -38 | -2.64 | 0.009 |
| LH_Default_Par_1; Left Angular Gyrus | -57 | -50 | 12 | RH_Default_pCunPCC_1; Right Ventral Posterior Cingulate Cortex | 12 | -54 | 14 | 2.63 | 0.009 |
| LH_Vis_6; Left Primary Visual Cortex | -12 | -67 | 7 | LH_Default_Par_1; Left Angular Gyrus | -57 | -50 | 12 | 2.61 | 0.010 |
| RH_DorsAttn_Post_4; Right Visual Motor Cortex | 27 | -67 | 51 | RH_Cont_PFCl_4; Right Frontal Eye Fields | 43 | 16 | 45 | 2.60 | 0.010 |

**Eyes-Open Alpha**

| ROI1 Labels (Schaefer; Talairach) | X | Y | Z | ROI2 Labels (Schaefer; Talairach) | X | Y | Z | t | p (unc) |
|---|---|---|---|---|---|---|---|---|---|
| RH_Default_Temp_3; Right Superior Temporal Gyrus | 57 | -26 | -2 | RH_Default_PFCdPFCm_2; Right Dorsolateral Prefrontal Cortex | 11 | 50 | 39 | -4.40 | 2.00E-05 |
| LH_SomMot_4; Left PreMotor & Supplementary Motor Area | -55 | -8 | 34 | RH_Default_PFCdPFCm_1; Right Anterior Prefrontal Cortex | 7 | 48 | 1 | -4.02 | 9.01E-05 |
| LH_DorsAttn_Post_2; Left Supramarginal Gyrus | -57 | -25 | 39 | RH_Default_PFCdPFCm_1; Right Anterior Prefrontal Cortex | 7 | 48 | 1 | -4.02 | 9.08E-05 |
| LH_DorsAttn_Post_2; Left Supramarginal Gyrus | -57 | -25 | 39 | LH_Default_PFC_3; Left Dorsal Posterior Cingulate Cortex | -6 | 47 | 0 | -3.98 | 0.0001 |
| RH_Cont_Par_1; Right Supramarginal Gyrus | 57 | -39 | 44 | RH_Default_Temp_1; Right Medial Temporal Gyrus | 62 | -23 | -19 | -3.87 | 0.0002 |
| RH_Default_Temp_2; Right Temporal Pole | 51 | 7 | -18 | RH_Default_Temp_3; Right Superior Temporal Gyrus | 57 | -26 | -2 | -3.82 | 0.0002 |
| RH_Cont_Par_2; Right Angular Gyrus | 45 | -63 | 46 | LH_Default_PFC_1; Left Pars Orbitalis | -35 | 21 | -11 | -3.80 | 0.0002 |
| LH_Default_PFC_5; Left Dorsolateral Prefrontal Cortex | -9 | 48 | 41 | RH_Default_Temp_3; Right Superior Temporal Gyrus | 57 | -26 | -2 | -3.77 | 0.0002 |
| LH_Vis_6; Left Primary Visual Cortex | -12 | -67 | 7 | RH_Default_Temp_2; Right Temporal Pole | 51 | 7 | -18 | -3.70 | 0.0003 |
| LH_SomMot_4; Left PreMotor & Supplementary Motor Area | -55 | -8 | 34 | LH_Default_PFC_3; Left Dorsal Posterior Cingulate Cortex | -6 | 47 | 0 | -3.68 | 0.0003 |
| LH_SomMot_4; Left PreMotor & Supplementary Motor Area | -55 | -8 | 34 | RH_Limbic_TempPole_1; Right Inferior Temporal Gyrus | 38 | 1 | -38 | -3.64 | 0.0004 |
| RH_Vis_7; Right Visual Association Cortex | 36 | -82 | 16 | RH_SomMot_4; Right Primary Motor Cortex | 58 | -5 | 31 | -3.60 | 0.0004 |
| LH_Default_PFC_1; Left Pars Orbitalis | -35 | 21 | -11 | RH_Default_Temp_3; Right Superior Temporal Gyrus | 57 | -26 | -2 | -3.57 | 0.0005 |
| RH_SalVentAttn_FrOperIns_1; Right Insula | 40 | 8 | 1 | RH_Default_Temp_3; Right Superior Temporal Gyrus | 57 | -26 | -2 | -3.57 | 0.0005 |
| RH_SomMot_1; Right Primary Auditory Cortex | 53 | -16 | 7 | RH_Default_PFCdPFCm_2; Right Dorsolateral Prefrontal Cortex | 11 | 50 | 39 | -3.56 | 0.0005 |
| RH_Cont_Par_1; Right Supramarginal Gyrus | 57 | -39 | 44 | LH_Default_Par_1; Left Angular Gyrus | -57 | -50 | 12 | -3.55 | 0.0005 |
| RH_Cont_Par_1; Right Supramarginal Gyrus | 57 | -39 | 44 | RH_Default_Temp_3; Right Superior Temporal Gyrus | 57 | -26 | -2 | -3.54 | 0.0005 |
| LH_Cont_Cing_1; Left Ventral Posterior Cingulate Cortex | -4 | -26 | 33 | RH_Default_Temp_3; Right Superior Temporal Gyrus | 57 | -26 | -2 | -3.53 | 0.0005 |
| RH_SomMot_6; Right Primary Motor Cortex | 41 | -22 | 60 | LH_Default_Par_2; Left Angular Gyrus | -48 | -63 | 35 | -3.52 | 0.0006 |
| LH_DorsAttn_Post_1; Left Fusiform Cortex | -47 | -58 | -13 | RH_Cont_PFCl_2; Right Dorsolateral Prefrontal Cortex | 45 | 39 | 15 | -3.51 | 0.0006 |

| | | | | | | | | | |
|---|---|---|---|---|---|---|---|---|---|
| LH_DorsAttn_Post_2; Left Supramarginal Gyrus | -57 | -25 | 39 | RH_Default_Temp_3; Right Superior Temporal Gyrus | 57 | -26 | -2 | -3.49 | 0.0006 |
| LH_Vis_2; Left Visual Association Cortex | -26 | -77 | -14 | RH_SomMot_3; Right Primary Motor Cortex | 57 | -4 | 11 | -3.49 | 0.0006 |
| LH_Vis_6; Left Primary Visual Cortex | -12 | -67 | 7 | RH_Default_Temp_3; Right Superior Temporal Gyrus | 57 | -26 | -2 | -3.46 | 0.0007 |
| RH_Vis_5; Right Primary Visual Cortex | 8 | -76 | 5 | LH_SomMot_2; Left Primary Sensory Cortex | -37 | -21 | 16 | -3.44 | 0.0007 |
| RH_SomMot_2; Right Insula | 40 | -15 | 15 | RH_Cont_Cing_1; Right Ventral Posterior Cingulate Cortex | 5 | -27 | 33 | -3.42 | 0.0008 |
| LH_Vis_3; Left Visual Association Cortex | -17 | -60 | -7 | RH_Default_Temp_2; Right Temporal Pole | 51 | 7 | -18 | -3.41 | 0.0008 |
| LH_Default_PFC_1; Left Pars Orbitalis | -35 | 21 | -11 | RH_Default_Temp_1; Right Medial Temporal Gyrus | 62 | -23 | -19 | -3.40 | 0.0008 |
| RH_SomMot_2; Right Insula | 40 | -15 | 15 | LH_SalVentAttn_ParOper_1; Left Supramarginal Gyrus | -59 | -38 | 29 | -3.40 | 0.0009 |
| LH_Vis_2; Left Visual Association Cortex | -26 | -77 | -14 | RH_Default_Temp_3; Right Superior Temporal Gyrus | 57 | -26 | -2 | -3.38 | 0.0009 |
| RH_SomMot_1; Right Primary Auditory Cortex | 53 | -16 | 7 | RH_Default_Temp_2; Right Temporal Pole | 51 | 7 | -18 | -3.36 | 0.0010 |
| RH_Cont_PFCmp_1; Right Frontal Eye Fields | 6 | 28 | 30 | RH_Default_Temp_3; Right Superior Temporal Gyrus | 57 | -26 | -2 | -3.36 | 0.0010 |
| LH_Limbic_TempPole_2; Left Inferior Temporal Gyrus | -57 | -33 | -21 | LH_Default_PFC_7; Left Frontal Eye Fields | -25 | 20 | 51 | -3.35 | 0.001 |
| LH_SomMot_4; Left PreMotor & Supplementary Motor Area | -55 | -8 | 34 | LH_Default_Temp_2; Left Medial Temporal Gyrus | -58 | -32 | -1 | -3.33 | 0.001 |
| RH_SalVentAttn_TempOccPar_1; Right Superior Temporal Gyrus | 58 | -42 | 13 | LH_Default_Par_1; Left Angular Gyrus | -57 | -50 | 12 | -3.32 | 0.001 |
| LH_Default_Par_1; Left Angular Gyrus | -57 | -50 | 12 | RH_Default_Temp_3; Right Superior Temporal Gyrus | 57 | -26 | -2 | -3.31 | 0.001 |
| LH_Vis_2; Left Visual Association Cortex | -26 | -77 | -14 | RH_Default_Temp_1; Right Medial Temporal Gyrus | 62 | -23 | -19 | -3.30 | 0.001 |
| RH_Cont_Par_2; Right Angular Gyrus | 45 | -63 | 46 | LH_Default_Par_1; Left Angular Gyrus | -57 | -50 | 12 | -3.30 | 0.001 |
| RH_SomMot_2; Right Insula | 40 | -15 | 15 | LH_Default_Par_1; Left Angular Gyrus | -57 | -50 | 12 | -3.28 | 0.001 |
| LH_Vis_6; Left Primary Visual Cortex | -12 | -67 | 7 | RH_Default_Temp_1; Right Medial Temporal Gyrus | 62 | -23 | -19 | -3.27 | 0.001 |
| RH_DorsAttn_Post_2; Right Primary Sensory Cortex | 50 | -24 | 42 | LH_Default_Par_2; Left Angular Gyrus | -48 | -63 | 35 | -3.27 | 0.001 |
| RH_SomMot_7; Right Primary Sensory Cortex | 30 | -37 | 64 | LH_DorsAttn_Post_4; Left Primary Sensory Cortex | -42 | -34 | 48 | -3.25 | 0.001 |
| RH_SomMot_1; Right Primary Auditory Cortex | 53 | -16 | 7 | RH_SalVentAttn_FrOperIns_1; Right Insula | 40 | 8 | 1 | -3.25 | 0.001 |
| LH_Vis_6; Left Primary Visual Cortex | -12 | -67 | 7 | RH_Cont_Par_2; Right Angular Gyrus | 45 | -63 | 46 | -3.25 | 0.001 |
| LH_DorsAttn_Post_2; Left Supramarginal Gyrus | -57 | -25 | 39 | RH_Default_Temp_2; Right Temporal Pole | 51 | 7 | -18 | -3.24 | 0.001 |
| RH_SomMot_1; Right Primary Auditory Cortex | 53 | -16 | 7 | LH_Default_PFC_5; Left Dorsolateral Prefrontal Cortex | -9 | 48 | 41 | -3.23 | 0.002 |
| RH_SomMot_1; Right Primary Auditory Cortex | 53 | -16 | 7 | RH_Cont_PFCmp_1; Right Frontal Eye Fields | 6 | 28 | 30 | -3.22 | 0.002 |
| RH_SomMot_1; Right Primary Auditory Cortex | 53 | -16 | 7 | RH_Default_pCunPCC_1; Right Ventral Posterior Cingulate Cortex | 12 | -54 | 14 | -3.22 | 0.002 |
| LH_Vis_1; Left Fusiform Cortex | -26 | -34 | -17 | LH_SomMot_2; Left Primary Sensory Cortex | -37 | -21 | 16 | -3.22 | 0.002 |
| LH_Vis_2; Left Visual Association Cortex | -26 | -77 | -14 | RH_Default_Temp_2; Right Temporal Pole | 51 | 7 | -18 | -3.22 | 0.002 |
| RH_Default_Temp_3; Right Superior Temporal Gyrus | 57 | -26 | -2 | RH_Default_PFCv_2; Right Broca's Area/ Pars Triangularis | 51 | 28 | 0 | -3.21 | 0.002 |
| LH_SalVentAttn_FrOperIns_1; Left Insula | -41 | -1 | -7 | RH_Default_PFCv_2; Right Broca's Area/ Pars Triangularis | 51 | 28 | 0 | -3.19 | 0.002 |
| RH_DorsAttn_Post_1; Right Visual Association Cortex | 50 | -62 | 16 | LH_Default_Par_1; Left Angular Gyrus | -57 | -50 | 12 | -3.19 | 0.002 |

| | | | | | | | | | |
|---|---|---|---|---|---|---|---|---|---|
| LH_Vis_2; Left Visual Association Cortex | -26 | -77 | -14 | RH_DorsAttn_PrCv_1; Right Broca's Area/Operculum | 49 | 10 | 27 | -3.18 | 0.002 |
| LH_SomMot_1; Left Supramarginal Gyrus | -53 | -23 | 8 | LH_SomMot_4; Left PreMotor & Supplementary Motor Area | -55 | -8 | 34 | -3.17 | 0.002 |
| RH_Default_Temp_3; Right Superior Temporal Gyrus | 57 | -26 | -2 | RH_Default_pCunPCC_1; Right Ventral Posterior Cingulate Cortex | 12 | -54 | 14 | -3.17 | 0.002 |
| RH_SomMot_3; Right Primary Motor Cortex | 57 | -4 | 11 | RH_Default_PFCdPFCm_2; Right Dorsolateral Prefrontal Cortex | 11 | 50 | 39 | -3.17 | 0.002 |
| LH_SalVentAttn_ParOper_1; Left Supramarginal Gyrus | -59 | -38 | 29 | RH_Default_Temp_2; Right Temporal Pole | 51 | 7 | -18 | -3.17 | 0.002 |
| LH_SomMot_6; Left Primary Motor Cortex | -6 | -29 | 70 | RH_Default_PFCdPFCm_3; Right Frontal Eye Fields | 26 | 24 | 50 | -3.17 | 0.002 |
| LH_DorsAttn_Post_2; Left Supramarginal Gyrus | -57 | -25 | 39 | LH_Default_PFC_4; Left Anterior Prefrontal Cortex | -24 | 61 | -1 | -3.16 | 0.002 |
| RH_SomMot_3; Right Primary Motor Cortex | 57 | -4 | 11 | RH_Default_Temp_2; Right Temporal Pole | 51 | 7 | -18 | -3.16 | 0.002 |
| LH_Vis_5; Left Secondary Visual Cortex | -5 | -92 | -2 | RH_Cont_PFCl_1; Right Anterior Prefrontal Cortex | 30 | 58 | -3 | -3.16 | 0.002 |
| LH_Limbic_TempPole_2; Left Inferior Temporal Gyrus | -57 | -33 | -21 | LH_Default_Par_1; Left Angular Gyrus | -57 | -50 | 12 | -3.16 | 0.002 |
| RH_SomMot_1; Right Primary Auditory Cortex | 53 | -16 | 7 | LH_Cont_Cing_1; Left Ventral Posterior Cingulate Cortex | -4 | -26 | 33 | -3.15 | 0.002 |
| RH_SomMot_4; Right Primary Motor Cortex | 58 | -5 | 31 | RH_SalVentAttn_TempOccPar_2; Right Supramarginal Gyrus | 61 | -26 | 27 | -3.15 | 0.002 |
| LH_SalVentAttn_Med_1; Left Dorsal Anterior Cingulate Cortex | -5 | 20 | 34 | LH_Limbic_TempPole_2; Left Inferior Temporal Gyrus | -57 | -33 | -21 | -3.15 | 0.002 |
| LH_Vis_5; Left Secondary Visual Cortex | -5 | -92 | -2 | RH_Default_PFCv_1; Right Pars Orbitalis | 35 | 26 | -15 | -3.14 | 0.002 |
| LH_SomMot_3; Left Primary Sensory Cortex | -54 | -12 | 13 | LH_DorsAttn_PrCv_1; Left PreMotor & Supplementary Motor Area | -48 | 6 | 28 | -3.13 | 0.002 |
| RH_SalVentAttn_TempOccPar_1; Right Superior Temporal Gyrus | 58 | -42 | 13 | RH_Default_PFCdPFCm_2; Right Dorsolateral Prefrontal Cortex | 11 | 50 | 39 | -3.13 | 0.002 |
| RH_Vis_4; Right Secondary Visual Cortex | 22 | -96 | -5 | LH_SomMot_2; Left Primary Sensory Cortex | -37 | -21 | 16 | -3.12 | 0.002 |
| RH_DorsAttn_FEF_1; Right PreMotor & Supplementary Motor Area | 28 | -3 | 59 | RH_Default_Temp_3; Right Superior Temporal Gyrus | 57 | -26 | -2 | -3.12 | 0.002 |
| RH_Cont_Par_1; Right Supramarginal Gyrus | 57 | -39 | 44 | RH_Default_pCunPCC_2; Right Ventral Posterior Cingulate Cortex | 7 | -52 | 31 | -3.11 | 0.002 |
| RH_SomMot_2; Right Insula | 40 | -15 | 15 | RH_DorsAttn_PrCv_1; Right Broca's Area/Operculum | 49 | 10 | 27 | -3.11 | 0.002 |
| RH_SomMot_6; Right Primary Motor Cortex | 41 | -22 | 60 | RH_Default_Temp_3; Right Superior Temporal Gyrus | 57 | -26 | -2 | -3.11 | 0.002 |
| LH_DorsAttn_PrCv_1; Left PreMotor & Supplementary Motor Area | -48 | 6 | 28 | LH_Default_PFC_2; Left Pars Orbitalis | -47 | 33 | -3 | -3.10 | 0.002 |
| LH_Default_Par_1; Left Angular Gyrus | -57 | -50 | 12 | RH_Default_PFCdPFCm_3; Right Frontal Eye Fields | 26 | 24 | 50 | -3.10 | 0.002 |
| RH_DorsAttn_Post_4; Right Visual Motor Cortex | 27 | -67 | 51 | LH_Default_PFC_1; Left Pars Orbitalis | -35 | 21 | -11 | -3.10 | 0.002 |
| LH_SomMot_4; Left PreMotor & Supplementary Motor Area | -55 | -8 | 34 | RH_SalVentAttn_FrOperIns_1; Right Insula | 40 | 8 | 1 | -3.09 | 0.002 |
| RH_SomMot_1; Right Primary Auditory Cortex | 53 | -16 | 7 | LH_DorsAttn_Post_2; Left Supramarginal Gyrus | -57 | -25 | 39 | -3.09 | 0.002 |
| RH_Cont_Par_1; Right Supramarginal Gyrus | 57 | -39 | 44 | LH_Default_pCunPCC_2; Left Ventral Posterior Cingulate Cortex | -6 | -53 | 33 | -3.09 | 0.002 |
| LH_Vis_6; Left Primary Visual Cortex | -12 | -67 | 7 | RH_SomMot_3; Right Primary Motor Cortex | 57 | -4 | 11 | -3.09 | 0.002 |
| RH_Cont_PFCl_4; Right Frontal Eye Fields | 43 | 16 | 45 | RH_Default_Temp_3; Right Superior Temporal Gyrus | 57 | -26 | -2 | -3.08 | 0.002 |
| RH_SomMot_4; Right Primary Motor Cortex | 58 | -5 | 31 | LH_Limbic_TempPole_1; Left Temporal Pole | -32 | 2 | -37 | -3.08 | 0.002 |
| RH_Vis_4; Right Secondary Visual Cortex | 22 | -96 | -5 | RH_SomMot_3; Right Primary Motor Cortex | 57 | -4 | 11 | -3.07 | 0.003 |
| RH_DorsAttn_FEF_1; Right PreMotor & Supplementary Motor Area | 28 | -3 | 59 | RH_Default_Temp_2; Right Temporal Pole | 51 | 7 | -18 | -3.06 | 0.003 |

| | | | | | | | | | |
|---|---|---|---|---|---|---|---|---|---|
| LH_Vis_5; Left Secondary Visual Cortex | -5 | -92 | -2 | RH_Cont_PFCl_2; Right Dorsolateral Prefrontal Cortex | 45 | 39 | 15 | -3.06 | 0.003 |
| RH_SomMot_1; Right Primary Auditory Cortex | 53 | -16 | 7 | RH_Cont_Cing_1; Right Ventral Posterior Cingulate Cortex | 5 | -27 | 33 | -3.06 | 0.003 |
| RH_SomMot_2; Right Insula | 40 | -15 | 15 | RH_Cont_PFCl_1; Right Anterior Prefrontal Cortex | 30 | 58 | -3 | -3.05 | 0.003 |
| LH_SomMot_3; Left Primary Sensory Cortex | -54 | -12 | 13 | LH_DorsAttn_Post_1; Left Fusiform Cortex | -47 | -58 | -13 | -3.05 | 0.003 |
| LH_SomMot_4; Left PreMotor & Supplementary Motor Area | -55 | -8 | 34 | LH_DorsAttn_PrCv_1; Left PreMotor & Supplementary Motor Area | -48 | 6 | 28 | -3.05 | 0.003 |
| RH_SomMot_3; Right Primary Motor Cortex | 57 | -4 | 11 | RH_SalVentAttn_TempOccPar_1; Right Superior Temporal Gyrus | 58 | -42 | 13 | -3.05 | 0.003 |
| RH_Vis_4; Right Secondary Visual Cortex | 22 | -96 | -5 | RH_Default_Temp_2; Right Temporal Pole | 51 | 7 | -18 | -3.04 | 0.003 |
| RH_Vis_6; Right Secondary Visual Cortex | 17 | -57 | 5 | RH_SomMot_1; Right Primary Auditory Cortex | 53 | -16 | 7 | -3.04 | 0.003 |
| RH_DorsAttn_Post_1; Right Visual Association Cortex | 50 | -62 | 16 | LH_Default_PFC_1; Left Pars Orbitalis | -35 | 21 | -11 | -3.04 | 0.003 |
| LH_SalVentAttn_Med_1; Left Dorsal Anterior Cingulate Cortex | -5 | 20 | 34 | RH_Default_Temp_1; Right Medial Temporal Gyrus | 62 | -23 | -19 | -3.03 | 0.003 |
| LH_Vis_4; Left Secondary Visual Cortex | -27 | -95 | -4 | RH_SomMot_3; Right Primary Motor Cortex | 57 | -4 | 11 | -3.03 | 0.003 |
| LH_SalVentAttn_Med_1; Left Dorsal Anterior Cingulate Cortex | -5 | 20 | 34 | RH_Default_Temp_3; Right Superior Temporal Gyrus | 57 | -26 | -2 | -3.03 | 0.003 |
| LH_SomMot_4; Left PreMotor & Supplementary Motor Area | -55 | -8 | 34 | RH_Default_PFCv_1; Right Pars Orbitalis | 35 | 26 | -15 | -3.02 | 0.003 |
| LH_DorsAttn_Post_3; Left Visual Motor Cortex | -24 | -68 | 49 | RH_Cont_Par_1; Right Supramarginal Gyrus | 57 | -39 | 44 | -3.01 | 0.003 |
| RH_Vis_5; Right Primary Visual Cortex | 8 | -76 | 5 | RH_SomMot_3; Right Primary Motor Cortex | 57 | -4 | 11 | -3.01 | 0.003 |
| RH_SomMot_5; Right PreMotor & Supplementary Motor Area | 47 | -11 | 48 | RH_SalVentAttn_TempOccPar_2; Right Supramarginal Gyrus | 61 | -26 | 27 | -3.01 | 0.003 |
| RH_SalVentAttn_TempOccPar_2; Right Supramarginal Gyrus | 61 | -26 | 27 | LH_Default_Par_1; Left Angular Gyrus | -57 | -50 | 12 | -3.00 | 0.003 |
| RH_SomMot_2; Right Insula | 40 | -15 | 15 | RH_Default_Temp_3; Right Superior Temporal Gyrus | 57 | -26 | -2 | -3.00 | 0.003 |
| LH_SomMot_3; Left Primary Sensory Cortex | -54 | -12 | 13 | LH_SomMot_4; Left PreMotor & Supplementary Motor Area | -55 | -8 | 34 | -3.00 | 0.003 |
| LH_Vis_6; Left Primary Visual Cortex | -12 | -67 | 7 | LH_SomMot_2; Left Primary Sensory Cortex | -37 | -21 | 16 | -3.00 | 0.003 |
| RH_Cont_Cing_1; Right Ventral Posterior Cingulate Cortex | 5 | -27 | 33 | RH_Default_Temp_3; Right Superior Temporal Gyrus | 57 | -26 | -2 | -3.00 | 0.003 |
| RH_Vis_6; Right Secondary Visual Cortex | 17 | -57 | 5 | RH_Default_Temp_3; Right Superior Temporal Gyrus | 57 | -26 | -2 | -2.99 | 0.003 |
| LH_Vis_7; Left Visual Association Cortex | -47 | -71 | 11 | RH_Vis_6; Right Secondary Visual Cortex | 17 | -57 | 5 | -2.99 | 0.003 |
| RH_Vis_6; Right Secondary Visual Cortex | 17 | -57 | 5 | RH_SomMot_4; Right Primary Motor Cortex | 58 | -5 | 31 | -2.98 | 0.003 |
| RH_SomMot_2; Right Insula | 40 | -15 | 15 | RH_Default_PFCdPFCm_2; Right Dorsolateral Prefrontal Cortex | 11 | 50 | 39 | -2.98 | 0.003 |
| LH_SomMot_4; Left PreMotor & Supplementary Motor Area | -55 | -8 | 34 | RH_Default_PFCv_2; Right Broca's Area/ Pars Triangularis | 51 | 28 | 0 | -2.98 | 0.003 |
| RH_Vis_6; Right Secondary Visual Cortex | 17 | -57 | 5 | RH_Default_PFCdPFCm_2; Right Dorsolateral Prefrontal Cortex | 11 | 50 | 39 | -2.97 | 0.003 |
| RH_SalVentAttn_TempOccPar_1; Right Superior Temporal Gyrus | 58 | -42 | 13 | RH_Default_pCunPCC_2; Right Ventral Posterior Cingulate Cortex | 7 | -52 | 31 | -2.96 | 0.004 |
| RH_SomMot_1; Right Primary Auditory Cortex | 53 | -16 | 7 | RH_DorsAttn_PrCv_1; Right Broca's Area/Operculum | 49 | 10 | 27 | -2.96 | 0.004 |
| RH_DorsAttn_FEF_1; Right PreMotor & Supplementary Motor Area | 28 | -3 | 59 | RH_Default_PFCdPFCm_3; Right Frontal Eye Fields | 26 | 24 | 50 | -2.96 | 0.004 |
| RH_SalVentAttn_TempOccPar_1; Right Superior Temporal Gyrus | 58 | -42 | 13 | RH_Cont_PFCl_4; Right Frontal Eye Fields | 43 | 16 | 45 | -2.96 | 0.004 |
| RH_SomMot_3; Right Primary Motor Cortex | 57 | -4 | 11 | LH_Default_PFC_5; Left Dorsolateral Prefrontal Cortex | -9 | 48 | 41 | -2.95 | 0.004 |

| | | | | | | | | | |
|---|---|---|---|---|---|---|---|---|---|
| RH_SomMot_1; Right Primary Auditory Cortex | 53 | -16 | 7 | LH_Default_PFC_1; Left Pars Orbitalis | -35 | 21 | -11 | -2.95 | 0.004 |
| LH_Vis_1; Left Fusiform Cortex | -26 | -34 | -17 | LH_SomMot_4; Left PreMotor & Supplementary Motor Area | -55 | -8 | 34 | -2.95 | 0.004 |
| LH_DorsAttn_Post_1; Left Fusiform Cortex | -47 | -58 | -13 | LH_Default_PFC_6; Left PreMotor & Supplementary Motor Area | -41 | 14 | 48 | -2.95 | 0.004 |
| RH_Vis_6; Right Secondary Visual Cortex | 17 | -57 | 5 | RH_Default_Temp_1; Right Medial Temporal Gyrus | 62 | -23 | -19 | -2.95 | 0.004 |
| RH_SomMot_1; Right Primary Auditory Cortex | 53 | -16 | 7 | LH_Default_pCunPCC_1; Left Ventral Posterior Cingulate Cortex | -11 | -56 | 13 | -2.94 | 0.004 |
| RH_SomMot_2; Right Insula | 40 | -15 | 15 | LH_Cont_Cing_1; Left Ventral Posterior Cingulate Cortex | -4 | -26 | 33 | -2.94 | 0.004 |
| RH_SomMot_1; Right Primary Auditory Cortex | 53 | -16 | 7 | RH_Default_PFCv_2; Right Broca's Area/ Pars Triangularis | 51 | 28 | 0 | -2.93 | 0.004 |
| LH_Vis_6; Left Primary Visual Cortex | -12 | -67 | 7 | RH_SalVentAttn_TempOccPar_1; Right Superior Temporal Gyrus | 58 | -42 | 13 | -2.93 | 0.004 |
| LH_SomMot_3; Left Primary Sensory Cortex | -54 | -12 | 13 | RH_Default_Temp_2; Right Temporal Pole | 51 | 7 | -18 | -2.93 | 0.004 |
| RH_Default_Temp_1; Right Medial Temporal Gyrus | 62 | -23 | -19 | RH_Default_PFCdPFCm_2; Right Dorsolateral Prefrontal Cortex | 11 | 50 | 39 | -2.92 | 0.004 |
| LH_Default_PFC_1; Left Pars Orbitalis | -35 | 21 | -11 | RH_Default_Par_1; Right Angular Gyrus | 55 | -51 | 31 | -2.92 | 0.004 |
| RH_SalVentAttn_TempOccPar_2; Right Supramarginal Gyrus | 61 | -26 | 27 | LH_Limbic_TempPole_1; Left Temporal Pole | -32 | 2 | -37 | -2.92 | 0.004 |
| RH_Cont_PFCl_2; Right Dorsolateral Prefrontal Cortex | 45 | 39 | 15 | RH_Default_Temp_3; Right Superior Temporal Gyrus | 57 | -26 | -2 | -2.92 | 0.004 |
| LH_Vis_4; Left Secondary Visual Cortex | -27 | -95 | -4 | RH_Cont_PFCl_1; Right Anterior Prefrontal Cortex | 30 | 58 | -3 | -2.91 | 0.004 |
| LH_DorsAttn_FEF_1; Left PreMotor & Supplementary Motor Area | -26 | -3 | 59 | LH_Default_Par_1; Left Angular Gyrus | -57 | -50 | 12 | -2.91 | 0.004 |
| RH_SalVentAttn_Med_1; Right Dorsal Posterior Cingulate Cortex | 11 | -31 | 45 | RH_Cont_Cing_1; Right Ventral Posterior Cingulate Cortex | 5 | -27 | 33 | -2.90 | 0.004 |
| RH_DorsAttn_PrCv_1; Right Broca's Area/Operculum | 49 | 10 | 27 | RH_Default_Temp_1; Right Medial Temporal Gyrus | 62 | -23 | -19 | -2.90 | 0.004 |
| RH_Vis_4; Right Secondary Visual Cortex | 22 | -96 | -5 | RH_SomMot_1; Right Primary Auditory Cortex | 53 | -16 | 7 | -2.90 | 0.004 |
| RH_Vis_7; Right Visual Association Cortex | 36 | -82 | 16 | RH_SomMot_3; Right Primary Motor Cortex | 57 | -4 | 11 | -2.90 | 0.004 |
| LH_Default_pCunPCC_1; Left Ventral Posterior Cingulate Cortex | -11 | -56 | 13 | RH_Default_Temp_3; Right Superior Temporal Gyrus | 57 | -26 | -2 | -2.90 | 0.004 |
| RH_SomMot_3; Right Primary Motor Cortex | 57 | -4 | 11 | RH_SalVentAttn_FrOperIns_1; Right Insula | 40 | 8 | 1 | -2.90 | 0.004 |
| LH_DorsAttn_Post_1; Left Fusiform Cortex | -47 | -58 | -13 | RH_Cont_PFCl_3; Right Dorsolateral Prefrontal Cortex | 32 | 46 | 29 | -2.89 | 0.004 |
| RH_DorsAttn_Post_4; Right Visual Motor Cortex | 27 | -67 | 51 | RH_Default_PFCdPFCm_1; Right Anterior Prefrontal Cortex | 7 | 48 | 1 | -2.89 | 0.004 |
| LH_Vis_6; Left Primary Visual Cortex | -12 | -67 | 7 | RH_SomMot_1; Right Primary Auditory Cortex | 53 | -16 | 7 | -2.89 | 0.004 |
| LH_Vis_6; Left Primary Visual Cortex | -12 | -67 | 7 | LH_Vis_7; Left Visual Association Cortex | -47 | -71 | 11 | -2.88 | 0.005 |
| RH_SomMot_3; Right Primary Motor Cortex | 57 | -4 | 11 | LH_DorsAttn_Post_2; Left Supramarginal Gyrus | -57 | -25 | 39 | -2.87 | 0.005 |
| LH_Limbic_TempPole_1; Left Temporal Pole | -32 | 2 | -37 | RH_Default_PFCv_2; Right Broca's Area/ Pars Triangularis | 51 | 28 | 0 | -2.87 | 0.005 |
| LH_Cont_Par_1; Left Angular Gyrus | -37 | -53 | 46 | LH_Default_PFC_1; Left Pars Orbitalis | -35 | 21 | -11 | -2.87 | 0.005 |
| LH_Vis_5; Left Secondary Visual Cortex | -5 | -92 | -2 | LH_DorsAttn_Post_2; Left Supramarginal Gyrus | -57 | -25 | 39 | -2.87 | 0.005 |
| LH_DorsAttn_Post_5; Left Visual Motor Cortex | -6 | -60 | 56 | LH_Default_PFC_1; Left Pars Orbitalis | -35 | 21 | -11 | -2.85 | 0.005 |
| RH_SalVentAttn_TempOccPar_1; Right Superior Temporal Gyrus | 58 | -42 | 13 | RH_SalVentAttn_FrOperIns_1; Right Insula | 40 | 8 | 1 | -2.85 | 0.005 |
| RH_SomMot_1; Right Primary Auditory Cortex | 53 | -16 | 7 | LH_Limbic_TempPole_1; Left Temporal Pole | -32 | 2 | -37 | -2.85 | 0.005 |

| | | | | | | | | | |
|---|---|---|---|---|---|---|---|---|---|
| LH_SomMot_3; Left Primary Sensory Cortex | -54 | -12 | 13 | RH_Cont_PFCl_1; Right Anterior Prefrontal Cortex | 30 | 58 | -3 | -2.85 | 0.005 |
| RH_DorsAttn_Post_5; Right Visual Motor Cortex | 14 | -52 | 66 | RH_Cont_PFCl_4; Right Frontal Eye Fields | 43 | 16 | 45 | -2.85 | 0.005 |
| RH_Vis_1; Right Fusiform Cortex | 32 | -31 | -22 | LH_SomMot_4; Left PreMotor & Supplementary Motor Area | -55 | -8 | 34 | -2.85 | 0.005 |
| LH_Vis_2; Left Visual Association Cortex | -26 | -77 | -14 | RH_SomMot_5; Right PreMotor & Supplementary Motor Area | 47 | -11 | 48 | -2.85 | 0.005 |
| RH_DorsAttn_Post_3; Right Visual Motor Cortex | 38 | -45 | 49 | LH_Default_Par_1; Left Angular Gyrus | -57 | -50 | 12 | -2.84 | 0.005 |
| LH_DorsAttn_Post_1; Left Fusiform Cortex | -47 | -58 | -13 | LH_Cont_PFCl_1; Left Dorsolateral Prefrontal Cortex | -43 | 33 | 21 | -2.84 | 0.005 |
| RH_SomMot_1; Right Primary Auditory Cortex | 53 | -16 | 7 | RH_Default_PFCv_1; Right Pars Orbitalis | 35 | 26 | -15 | -2.84 | 0.005 |
| LH_Vis_2; Left Visual Association Cortex | -26 | -77 | -14 | RH_SomMot_4; Right Primary Motor Cortex | 58 | -5 | 31 | -2.84 | 0.005 |
| RH_SomMot_6; Right Primary Motor Cortex | 41 | -22 | 60 | LH_Cont_Par_1; Left Angular Gyrus | -37 | -53 | 46 | -2.84 | 0.005 |
| RH_SomMot_2; Right Insula | 40 | -15 | 15 | LH_Default_PFC_5; Left Dorsolateral Prefrontal Cortex | -9 | 48 | 41 | -2.83 | 0.005 |
| RH_SalVentAttn_TempOccPar_1; Right Superior Temporal Gyrus | 58 | -42 | 13 | LH_Cont_Cing_1; Left Ventral Posterior Cingulate Cortex | -4 | -26 | 33 | -2.83 | 0.005 |
| LH_Default_Par_1; Left Angular Gyrus | -57 | -50 | 12 | RH_Default_Par_1; Right Angular Gyrus | 55 | -51 | 31 | -2.83 | 0.005 |
| LH_Vis_2; Left Visual Association Cortex | -26 | -77 | -14 | RH_SomMot_1; Right Primary Auditory Cortex | 53 | -16 | 7 | -2.83 | 0.005 |
| LH_Vis_2; Left Visual Association Cortex | -26 | -77 | -14 | RH_Cont_PFCl_2; Right Dorsolateral Prefrontal Cortex | 45 | 39 | 15 | -2.83 | 0.005 |
| RH_SomMot_3; Right Primary Motor Cortex | 57 | -4 | 11 | RH_Default_Temp_3; Right Superior Temporal Gyrus | 57 | -26 | -2 | -2.83 | 0.005 |
| RH_Vis_7; Right Visual Association Cortex | 36 | -82 | 16 | RH_SomMot_1; Right Primary Auditory Cortex | 53 | -16 | 7 | -2.82 | 0.005 |
| LH_Vis_9; Left Visual Association Cortex | -6 | -82 | 26 | LH_Cont_Cing_1; Left Ventral Posterior Cingulate Cortex | -4 | -26 | 33 | -2.82 | 0.005 |
| LH_SomMot_4; Left PreMotor & Supplementary Motor Area | -55 | -8 | 34 | RH_Cont_PFCmp_1; Right Frontal Eye Fields | 6 | 28 | 30 | -2.81 | 0.006 |
| RH_Cont_PFCl_4; Right Frontal Eye Fields | 43 | 16 | 45 | RH_Default_Temp_1; Right Medial Temporal Gyrus | 62 | -23 | -19 | -2.81 | 0.006 |
| LH_Vis_5; Left Secondary Visual Cortex | -5 | -92 | -2 | RH_SomMot_3; Right Primary Motor Cortex | 57 | -4 | 11 | -2.81 | 0.006 |
| RH_SalVentAttn_Med_2; Right PreMotor & Supplementary Motor Area | 7 | 6 | 52 | LH_Cont_Par_1; Left Angular Gyrus | -37 | -53 | 46 | -2.81 | 0.006 |
| RH_SomMot_6; Right Primary Motor Cortex | 41 | -22 | 60 | RH_Default_Temp_1; Right Medial Temporal Gyrus | 62 | -23 | -19 | -2.81 | 0.006 |
| RH_DorsAttn_Post_2; Right Primary Sensory Cortex | 50 | -24 | 42 | RH_Default_Temp_3; Right Superior Temporal Gyrus | 57 | -26 | -2 | -2.80 | 0.006 |
| LH_DorsAttn_PrCv_1; Left PreMotor & Supplementary Motor Area | -48 | 6 | 28 | LH_Default_Temp_2; Left Medial Temporal Gyrus | -58 | -32 | -1 | -2.80 | 0.006 |
| RH_SalVentAttn_TempOccPar_1; Right Superior Temporal Gyrus | 58 | -42 | 13 | LH_Default_PFC_1; Left Pars Orbitalis | -35 | 21 | -11 | -2.79 | 0.006 |
| RH_SomMot_5; Right PreMotor & Supplementary Motor Area | 47 | -11 | 48 | RH_Default_Temp_3; Right Superior Temporal Gyrus | 57 | -26 | -2 | -2.79 | 0.006 |
| RH_Vis_1; Right Fusiform Cortex | 32 | -31 | -22 | LH_Limbic_TempPole_1; Left Temporal Pole | -32 | 2 | -37 | -2.79 | 0.006 |
| RH_SomMot_1; Right Primary Auditory Cortex | 53 | -16 | 7 | RH_Cont_PFCl_1; Right Anterior Prefrontal Cortex | 30 | 58 | -3 | -2.78 | 0.006 |
| RH_SomMot_1; Right Primary Auditory Cortex | 53 | -16 | 7 | RH_SomMot_5; Right PreMotor & Supplementary Motor Area | 47 | -11 | 48 | -2.78 | 0.006 |
| RH_SalVentAttn_Med_1; Right Dorsal Posterior Cingulate Cortex | 11 | -31 | 45 | RH_Default_PFCdPFCm_3; Right Frontal Eye Fields | 26 | 24 | 50 | -2.78 | 0.006 |
| RH_Cont_Par_2; Right Angular Gyrus | 45 | -63 | 46 | RH_Default_PFCdPFCm_1; Right Anterior Prefrontal Cortex | 7 | 48 | 1 | -2.78 | 0.006 |
| RH_DorsAttn_Post_1; Right Visual Association Cortex | 50 | -62 | 16 | RH_SalVentAttn_TempOccPar_2; Right Supramarginal Gyrus | 61 | -26 | 27 | -2.78 | 0.006 |

| | | | | | | | | | |
|---|---|---|---|---|---|---|---|---|---|
| LH_SalVentAttn_Med_1; Left Dorsal Anterior Cingulate Cortex | -5 | 20 | 34 | LH_Default_Temp_2; Left Medial Temporal Gyrus | -58 | -32 | -1 | -2.78 | 0.006 |
| LH_Vis_6; Left Primary Visual Cortex | -12 | -67 | 7 | RH_Limbic_TempPole_1; Right Inferior Temporal Gyrus | 38 | 1 | -38 | -2.78 | 0.006 |
| LH_Vis_8; Left Visual Association Cortex | -25 | -88 | 20 | LH_SomMot_3; Left Primary Sensory Cortex | -54 | -12 | 13 | -2.77 | 0.006 |
| RH_Vis_7; Right Visual Association Cortex | 36 | -82 | 16 | RH_Default_Temp_3; Right Superior Temporal Gyrus | 57 | -26 | -2 | -2.77 | 0.006 |
| LH_Cont_PFCl_1; Left Dorsolateral Prefrontal Cortex | -43 | 33 | 21 | RH_Cont_Par_2; Right Angular Gyrus | 45 | -63 | 46 | -2.76 | 0.006 |
| LH_SomMot_5; Left Primary Motor Cortex | -39 | -23 | 59 | LH_Default_PFC_4; Left Anterior Prefrontal Cortex | -24 | 61 | -1 | -2.76 | 0.006 |
| RH_Default_Temp_2; Right Temporal Pole | 51 | 7 | -18 | RH_Default_PFCv_1; Right Pars Orbitalis | 35 | 26 | -15 | -2.76 | 0.006 |
| RH_DorsAttn_Post_2; Right Primary Sensory Cortex | 50 | -24 | 42 | RH_SalVentAttn_TempOccPar_2; Right Supramarginal Gyrus | 61 | -26 | 27 | -2.76 | 0.006 |
| LH_Limbic_TempPole_2; Left Inferior Temporal Gyrus | -57 | -33 | -21 | LH_Default_Temp_2; Left Medial Temporal Gyrus | -58 | -32 | -1 | -2.76 | 0.007 |
| LH_Vis_6; Left Primary Visual Cortex | -12 | -67 | 7 | LH_Default_Par_1; Left Angular Gyrus | -57 | -50 | 12 | -2.76 | 0.007 |
| RH_Vis_7; Right Visual Association Cortex | 36 | -82 | 16 | RH_SomMot_2; Right Insula | 40 | -15 | 15 | -2.75 | 0.007 |
| RH_SomMot_4; Right Primary Motor Cortex | 58 | -5 | 31 | RH_DorsAttn_Post_1; Right Visual Association Cortex | 50 | -62 | 16 | -2.75 | 0.007 |
| RH_SomMot_4; Right Primary Motor Cortex | 58 | -5 | 31 | LH_Limbic_OFC_1; Left Orbitofrontal Cortex | -14 | 32 | -20 | -2.75 | 0.007 |
| LH_Vis_6; Left Primary Visual Cortex | -12 | -67 | 7 | RH_Default_PFCv_1; Right Pars Orbitalis | 35 | 26 | -15 | -2.75 | 0.007 |
| RH_Cont_PFCmp_1; Right Frontal Eye Fields | 6 | 28 | 30 | RH_Default_Temp_1; Right Medial Temporal Gyrus | 62 | -23 | -19 | -2.74 | 0.007 |
| LH_Cont_Par_1; Left Angular Gyrus | -37 | -53 | 46 | RH_Cont_Par_1; Right Supramarginal Gyrus | 57 | -39 | 44 | -2.74 | 0.007 |
| LH_SalVentAttn_ParOper_1; Left Supramarginal Gyrus | -59 | -38 | 29 | RH_Default_Temp_3; Right Superior Temporal Gyrus | 57 | -26 | -2 | -2.74 | 0.007 |
| RH_Cont_PFCl_3; Right Dorsolateral Prefrontal Cortex | 32 | 46 | 29 | LH_Default_Par_1; Left Angular Gyrus | -57 | -50 | 12 | -2.74 | 0.007 |
| LH_Limbic_TempPole_1; Left Temporal Pole | -32 | 2 | -37 | RH_Cont_PFCl_4; Right Frontal Eye Fields | 43 | 16 | 45 | -2.74 | 0.007 |
| RH_SomMot_1; Right Primary Auditory Cortex | 53 | -16 | 7 | RH_SomMot_4; Right Primary Motor Cortex | 58 | -5 | 31 | -2.73 | 0.007 |
| LH_SalVentAttn_ParOper_1; Left Supramarginal Gyrus | -59 | -38 | 29 | RH_Default_PFCdPFCm_1; Right Anterior Prefrontal Cortex | 7 | 48 | 1 | -2.73 | 0.007 |
| RH_Vis_5; Right Primary Visual Cortex | 8 | -76 | 5 | LH_Default_Par_1; Left Angular Gyrus | -57 | -50 | 12 | -2.73 | 0.007 |
| RH_SomMot_4; Right Primary Motor Cortex | 58 | -5 | 31 | RH_Default_Temp_1; Right Medial Temporal Gyrus | 62 | -23 | -19 | -2.73 | 0.007 |
| RH_SalVentAttn_TempOccPar_1; Right Superior Temporal Gyrus | 58 | -42 | 13 | LH_Default_pCunPCC_2; Left Ventral Posterior Cingulate Cortex | -6 | -53 | 33 | -2.73 | 0.007 |
| LH_DorsAttn_Post_5; Left Visual Motor Cortex | -6 | -60 | 56 | RH_SalVentAttn_Med_2; Right PreMotor & Supplementary Motor Area | 7 | 6 | 52 | -2.72 | 0.007 |
| RH_SomMot_3; Right Primary Motor Cortex | 57 | -4 | 11 | RH_Default_pCunPCC_1; Right Ventral Posterior Cingulate Cortex | 12 | -54 | 14 | -2.72 | 0.007 |
| LH_Default_PFC_2; Left Pars Orbitalis | -47 | 33 | -3 | RH_Default_Par_1; Right Angular Gyrus | 55 | -51 | 31 | -2.72 | 0.007 |
| RH_SomMot_2; Right Insula | 40 | -15 | 15 | RH_SomMot_3; Right Primary Motor Cortex | 57 | -4 | 11 | -2.71 | 0.007 |
| RH_DorsAttn_Post_2; Right Primary Sensory Cortex | 50 | -24 | 42 | LH_Default_Par_1; Left Angular Gyrus | -57 | -50 | 12 | -2.71 | 0.008 |
| RH_DorsAttn_Post_2; Right Primary Sensory Cortex | 50 | -24 | 42 | RH_Cont_Par_1; Right Supramarginal Gyrus | 57 | -39 | 44 | -2.70 | 0.008 |
| RH_Vis_4; Right Secondary Visual Cortex | 22 | -96 | -5 | RH_Default_Temp_3; Right Superior Temporal Gyrus | 57 | -26 | -2 | -2.70 | 0.008 |

| | | | | | | | | | |
|---|---|---|---|---|---|---|---|---|---|
| RH_DorsAttn_PrCv_1; Right Broca's Area/Operculum | 49 | 10 | 27 | RH_Default_Temp_3; Right Superior Temporal Gyrus | 57 | -26 | -2 | -2.70 | 0.008 |
| RH_SomMot_3; Right Primary Motor Cortex | 57 | -4 | 11 | LH_Default_pCunPCC_1; Left Ventral Posterior Cingulate Cortex | -11 | -56 | 13 | -2.70 | 0.008 |
| LH_Cont_Cing_1; Left Ventral Posterior Cingulate Cortex | -4 | -26 | 33 | RH_Default_Temp_1; Right Medial Temporal Gyrus | 62 | -23 | -19 | -2.70 | 0.008 |
| RH_DorsAttn_Post_4; Right Visual Motor Cortex | 27 | -67 | 51 | RH_Default_Temp_2; Right Temporal Pole | 51 | 7 | -18 | -2.69 | 0.008 |
| RH_SomMot_2; Right Insula | 40 | -15 | 15 | LH_Default_PFC_1; Left Pars Orbitalis | -35 | 21 | -11 | -2.69 | 0.008 |
| RH_Vis_6; Right Secondary Visual Cortex | 17 | -57 | 5 | RH_Cont_PFCl_2; Right Dorsolateral Prefrontal Cortex | 45 | 39 | 15 | -2.69 | 0.008 |
| RH_Vis_6; Right Secondary Visual Cortex | 17 | -57 | 5 | LH_SomMot_3; Left Primary Sensory Cortex | -54 | -12 | 13 | -2.68 | 0.008 |
| LH_DorsAttn_PrCv_1; Left PreMotor & Supplementary Motor Area | -48 | 6 | 28 | LH_Default_Par_1; Left Angular Gyrus | -57 | -50 | 12 | -2.68 | 0.008 |
| LH_Limbic_TempPole_2; Left Inferior Temporal Gyrus | -57 | -33 | -21 | RH_Default_PFCv_1; Right Pars Orbitalis | 35 | 26 | -15 | -2.68 | 0.008 |
| RH_SomMot_5; Right PreMotor & Supplementary Motor Area | 47 | -11 | 48 | RH_Default_Temp_1; Right Medial Temporal Gyrus | 62 | -23 | -19 | -2.68 | 0.008 |
| LH_Vis_7; Left Visual Association Cortex | -47 | -71 | 11 | RH_Default_Temp_1; Right Medial Temporal Gyrus | 62 | -23 | -19 | -2.67 | 0.008 |
| LH_Default_pCunPCC_1; Left Ventral Posterior Cingulate Cortex | -11 | -56 | 13 | RH_Default_Temp_1; Right Medial Temporal Gyrus | 62 | -23 | -19 | -2.67 | 0.008 |
| RH_Vis_2; Right Fusiform Cortex | 27 | -66 | -12 | RH_SalVentAttn_TempOccPar_1; Right Superior Temporal Gyrus | 58 | -42 | 13 | -2.66 | 0.009 |
| LH_SalVentAttn_ParOper_1; Left Supramarginal Gyrus | -59 | -38 | 29 | RH_Cont_PFCl_3; Right Dorsolateral Prefrontal Cortex | 32 | 46 | 29 | -2.66 | 0.009 |
| LH_SomMot_2; Left Primary Sensory Cortex | -37 | -21 | 16 | LH_DorsAttn_PrCv_1; Left PreMotor & Supplementary Motor Area | -48 | 6 | 28 | -2.66 | 0.009 |
| LH_Vis_4; Left Secondary Visual Cortex | -27 | -95 | -4 | RH_Vis_5; Right Primary Visual Cortex | 8 | -76 | 5 | -2.66 | 0.009 |
| RH_DorsAttn_Post_2; Right Primary Sensory Cortex | 50 | -24 | 42 | LH_Cont_Par_1; Left Angular Gyrus | -37 | -53 | 46 | -2.66 | 0.009 |
| RH_SomMot_3; Right Primary Motor Cortex | 57 | -4 | 11 | RH_DorsAttn_Post_1; Right Visual Association Cortex | 50 | -62 | 16 | -2.66 | 0.009 |
| RH_Vis_3; Right Fusiform Cortex | 49 | -60 | -11 | RH_Default_pCunPCC_2; Right Ventral Posterior Cingulate Cortex | 7 | -52 | 31 | -2.65 | 0.009 |
| LH_Default_PFC_2; Left Pars Orbitalis | -47 | 33 | -3 | RH_Default_Temp_3; Right Superior Temporal Gyrus | 57 | -26 | -2 | -2.65 | 0.009 |
| RH_Cont_pCun_1; Right Visual Motor Cortex | 9 | -66 | 43 | RH_Default_Temp_2; Right Temporal Pole | 51 | 7 | -18 | -2.65 | 0.009 |
| LH_DorsAttn_Post_1; Left Fusiform Cortex | -47 | -58 | -13 | RH_Default_PFCdPFCm_3; Right Frontal Eye Fields | 26 | 24 | 50 | -2.64 | 0.009 |
| LH_DorsAttn_Post_2; Left Supramarginal Gyrus | -57 | -25 | 39 | RH_Cont_PFCl_1; Right Anterior Prefrontal Cortex | 30 | 58 | -3 | -2.64 | 0.009 |
| RH_Cont_Par_1; Right Supramarginal Gyrus | 57 | -39 | 44 | LH_Default_Par_2; Left Angular Gyrus | -48 | -63 | 35 | -2.63 | 0.009 |
| LH_SalVentAttn_ParOper_1; Left Supramarginal Gyrus | -59 | -38 | 29 | RH_SalVentAttn_FrOperIns_1; Right Insula | 40 | 8 | 1 | -2.63 | 0.009 |
| LH_DorsAttn_Post_2; Left Supramarginal Gyrus | -57 | -25 | 39 | RH_Default_Temp_1; Right Medial Temporal Gyrus | 62 | -23 | -19 | -2.63 | 0.009 |
| LH_Cont_pCun_1; Left Visual Motor Cortex | -9 | -73 | 38 | LH_Cont_Cing_1; Left Ventral Posterior Cingulate Cortex | -4 | -26 | 33 | -2.63 | 0.009 |
| LH_DorsAttn_Post_6; Left Visual Motor Cortex | -22 | -51 | 66 | RH_Cont_Par_1; Right Supramarginal Gyrus | 57 | -39 | 44 | -2.63 | 0.009 |
| RH_DorsAttn_Post_3; Right Visual Motor Cortex | 38 | -45 | 49 | LH_SalVentAttn_ParOper_1; Left Supramarginal Gyrus | -59 | -38 | 29 | -2.63 | 0.009 |
| LH_SomMot_4; Left PreMotor & Supplementary Motor Area | -55 | -8 | 34 | LH_Cont_PFCl_1; Left Dorsolateral Prefrontal Cortex | -43 | 33 | 21 | -2.63 | 0.009 |
| RH_SomMot_3; Right Primary Motor Cortex | 57 | -4 | 11 | LH_Limbic_TempPole_1; Left Temporal Pole | -32 | 2 | -37 | -2.63 | 0.009 |
| RH_SomMot_5; Right PreMotor & Supplementary Motor Area | 47 | -11 | 48 | RH_Cont_PFCl_4; Right Frontal Eye Fields | 43 | 16 | 45 | -2.62 | 0.010 |

| | | | | | | | | | |
|---|---|---|---|---|---|---|---|---|---|
| RH_Vis_6; Right Secondary Visual Cortex | 17 | -57 | 5 | RH_SomMot_3; Right Primary Motor Cortex | 57 | -4 | 11 | -2.62 | 0.010 |
| LH_Vis_2; Left Visual Association Cortex | -26 | -77 | -14 | RH_SalVentAttn_TempOccPar_1; Right Superior Temporal Gyrus | 58 | -42 | 13 | -2.62 | 0.010 |
| LH_Vis_3; Left Visual Association Cortex | -17 | -60 | -7 | RH_Default_Temp_3; Right Superior Temporal Gyrus | 57 | -26 | -2 | -2.62 | 0.010 |
| LH_SalVentAttn_Med_2; Left Dorsal Posterior Cingulate Cortex | -11 | -34 | 45 | RH_Default_PFCdPFCm_3; Right Frontal Eye Fields | 26 | 24 | 50 | -2.62 | 0.010 |
| RH_SomMot_1; Right Primary Auditory Cortex | 53 | -16 | 7 | LH_SalVentAttn_ParOper_1; Left Supramarginal Gyrus | -59 | -38 | 29 | -2.62 | 0.010 |
| LH_Default_Temp_1; Left Medial Temporal Gyrus | -55 | -4 | -20 | RH_Default_Temp_3; Right Superior Temporal Gyrus | 57 | -26 | -2 | -2.61 | 0.010 |
| LH_SomMot_2; Left Primary Sensory Cortex | -37 | -21 | 16 | RH_Cont_Par_2; Right Angular Gyrus | 45 | -63 | 46 | -2.61 | 0.010 |
| LH_Vis_3; Left Visual Association Cortex | -17 | -60 | -7 | RH_SomMot_3; Right Primary Motor Cortex | 57 | -4 | 11 | -2.61 | 0.010 |
| RH_Vis_6; Right Secondary Visual Cortex | 17 | -57 | 5 | LH_SomMot_2; Left Primary Sensory Cortex | -37 | -21 | 16 | -2.61 | 0.010 |
| LH_Vis_7; Left Visual Association Cortex | -47 | -71 | 11 | RH_Default_Temp_3; Right Superior Temporal Gyrus | 57 | -26 | -2 | -2.61 | 0.010 |
| RH_Vis_4; Right Secondary Visual Cortex | 22 | -96 | -5 | RH_SomMot_2; Right Insula | 40 | -15 | 15 | -2.61 | 0.010 |
| RH_SalVentAttn_FrOperIns_1; Right Insula | 40 | 8 | 1 | LH_Default_PFC_5; Left Dorsolateral Prefrontal Cortex | -9 | 48 | 41 | -2.61 | 0.010 |
| LH_DorsAttn_Post_4; Left Primary Sensory Cortex | -42 | -34 | 48 | RH_Cont_Par_1; Right Supramarginal Gyrus | 57 | -39 | 44 | -2.60 | 0.010 |
| RH_Vis_2; Right Fusiform Cortex | 27 | -66 | -12 | RH_Default_Temp_3; Right Superior Temporal Gyrus | 57 | -26 | -2 | -2.60 | 0.010 |
| LH_Default_PFC_5; Left Dorsolateral Prefrontal Cortex | -9 | 48 | 41 | RH_Default_Temp_1; Right Medial Temporal Gyrus | 62 | -23 | -19 | -2.60 | 0.010 |
| RH_Default_Temp_3; Right Superior Temporal Gyrus | 57 | -26 | -2 | RH_Default_pCunPCC_2; Right Ventral Posterior Cingulate Cortex | 7 | -52 | 31 | -2.60 | 0.010 |
| LH_SomMot_3; Left Primary Sensory Cortex | -54 | -12 | 13 | RH_Default_PFCv_1; Right Pars Orbitalis | 35 | 26 | -15 | -2.59 | 0.010 |
| RH_Vis_7; Right Visual Association Cortex | 36 | -82 | 16 | RH_Cont_PFCl_4; Right Frontal Eye Fields | 43 | 16 | 45 | -2.59 | 0.010 |
| **Eyes-Open Beta** | | | | | | | | | |
| **ROI1 Labels (Schaefer; Talairach)** | **X** | **Y** | **Z** | **ROI2 Labels (Schaefer; Talairach)** | **X** | **Y** | **Z** | **t** | **p (unc)** |
| RH_SalVentAttn_Med_1; Right Dorsal Posterior Cingulate Cortex | 11 | -31 | 45 | RH_Limbic_OFC_1; Right Orbitofrontal Cortex | 12 | 35 | -20 | 4.22 | 4.25E-05 |
| RH_Limbic_OFC_1; Right Orbitofrontal Cortex | 12 | 35 | -20 | RH_Cont_Cing_1; Right Ventral Posterior Cingulate Cortex | 5 | -27 | 33 | 3.71 | 0.0003 |
| RH_Limbic_OFC_1; Right Orbitofrontal Cortex | 12 | 35 | -20 | LH_Default_pCunPCC_2; Left Ventral Posterior Cingulate Cortex | -6 | -53 | 33 | 3.70 | 0.0003 |
| RH_Limbic_OFC_1; Right Orbitofrontal Cortex | 12 | 35 | -20 | RH_Default_pCunPCC_2; Right Ventral Posterior Cingulate Cortex | 7 | -52 | 31 | 3.51 | 0.0006 |
| LH_DorsAttn_Post_6; Left Visual Motor Cortex | -22 | -51 | 66 | LH_SalVentAttn_FrOperIns_1; Left Insula | -41 | -1 | -7 | 3.41 | 0.0008 |
| LH_Vis_4; Left Secondary Visual Cortex | -27 | -95 | -4 | LH_Default_Temp_1; Left Medial Temporal Gyrus | -55 | -4 | -20 | 3.38 | 0.0009 |
| RH_Vis_6; Right Secondary Visual Cortex | 17 | -57 | 5 | LH_Default_PFC_7; Left Frontal Eye Fields | -25 | 20 | 51 | 3.35 | 0.001 |
| RH_Cont_pCun_1; Right Visual Motor Cortex | 9 | -66 | 43 | RH_Default_PFCv_1; Right Pars Orbitalis | 35 | 26 | -15 | 3.34 | 0.001 |
| LH_Cont_Cing_1; Left Ventral Posterior Cingulate Cortex | -4 | -26 | 33 | LH_Default_PFC_1; Left Pars Orbitalis | -35 | 21 | -11 | 3.27 | 0.001 |
| LH_Default_Temp_1; Left Medial Temporal Gyrus | -55 | -4 | -20 | RH_Default_PFCdPFCm_2; Right Dorsolateral Prefrontal Cortex | 11 | 50 | 39 | 3.26 | 0.001 |
| LH_DorsAttn_Post_6; Left Visual Motor Cortex | -22 | -51 | 66 | LH_DorsAttn_PrCv_1; Left PreMotor & Supplementary Motor Area | -48 | 6 | 28 | 3.21 | 0.002 |

| | | | | | | | | | |
|---|---|---|---|---|---|---|---|---|---|
| LH_Vis_5; Left Secondary Visual Cortex | -5 | -92 | -2 | LH_Limbic_OFC_1; Left Orbitofrontal Cortex | -14 | 32 | -20 | 3.20 | 0.002 |
| LH_Cont_pCun_1; Left Visual Motor Cortex | -9 | -73 | 38 | RH_Default_PFCv_1; Right Pars Orbitalis | 35 | 26 | -15 | 3.19 | 0.002 |
| LH_DorsAttn_Post_5; Left Visual Motor Cortex | -6 | -60 | 56 | RH_Limbic_OFC_1; Right Orbitofrontal Cortex | 12 | 35 | -20 | 3.15 | 0.002 |
| LH_DorsAttn_Post_5; Left Visual Motor Cortex | -6 | -60 | 56 | RH_Default_PFCv_1; Right Pars Orbitalis | 35 | 26 | -15 | 3.14 | 0.002 |
| RH_Vis_5; Right Primary Visual Cortex | 8 | -76 | 5 | LH_Limbic_OFC_1; Left Orbitofrontal Cortex | -14 | 32 | -20 | 3.12 | 0.002 |
| LH_DorsAttn_FEF_1; Left PreMotor & Supplementary Motor Area | -26 | -3 | 59 | RH_DorsAttn_Post_5; Right Visual Motor Cortex | 14 | -52 | 66 | 3.10 | 0.002 |
| LH_SalVentAttn_FrOperIns_1; Left Insula | -41 | -1 | -7 | RH_Default_PFCdPFCm_2; Right Dorsolateral Prefrontal Cortex | 11 | 50 | 39 | 3.05 | 0.003 |
| RH_Cont_Cing_1; Right Ventral Posterior Cingulate Cortex | 5 | -27 | 33 | LH_Default_PFC_1; Left Pars Orbitalis | -35 | 21 | -11 | 3.04 | 0.003 |
| RH_Vis_3; Right Fusiform Cortex | 49 | -60 | -11 | LH_Default_pCunPCC_2; Left Ventral Posterior Cingulate Cortex | -6 | -53 | 33 | 3.02 | 0.003 |
| LH_Vis_5; Left Secondary Visual Cortex | -5 | -92 | -2 | RH_Limbic_OFC_1; Right Orbitofrontal Cortex | 12 | 35 | -20 | 2.97 | 0.003 |
| RH_SomMot_7; Right Primary Sensory Cortex | 30 | -37 | 64 | RH_Limbic_TempPole_1; Right Inferior Temporal Gyrus | 38 | 1 | -38 | 2.97 | 0.003 |
| LH_SomMot_2; Left Primary Sensory Cortex | -37 | -21 | 16 | LH_Default_PFC_5; Left Dorsolateral Prefrontal Cortex | -9 | 48 | 41 | 2.95 | 0.004 |
| LH_SalVentAttn_FrOperIns_1; Left Insula | -41 | -1 | -7 | LH_Default_Par_2; Left Angular Gyrus | -48 | -63 | 35 | 2.95 | 0.004 |
| RH_Limbic_OFC_1; Right Orbitofrontal Cortex | 12 | 35 | -20 | LH_Cont_Cing_1; Left Ventral Posterior Cingulate Cortex | -4 | -26 | 33 | 2.93 | 0.004 |
| RH_SomMot_1; Right Primary Auditory Cortex | 53 | -16 | 7 | RH_Default_PFCv_1; Right Pars Orbitalis | 35 | 26 | -15 | 2.92 | 0.004 |
| LH_DorsAttn_FEF_1; Left PreMotor & Supplementary Motor Area | -26 | -3 | 59 | LH_Cont_Par_1; Left Angular Gyrus | -37 | -53 | 46 | 2.91 | 0.004 |
| RH_Limbic_OFC_1; Right Orbitofrontal Cortex | 12 | 35 | -20 | LH_Default_pCunPCC_1; Left Ventral Posterior Cingulate Cortex | -11 | -56 | 13 | 2.90 | 0.004 |
| LH_Default_PFC_4; Left Anterior Prefrontal Cortex | -24 | 61 | -1 | RH_Default_pCunPCC_1; Right Ventral Posterior Cingulate Cortex | 12 | -54 | 14 | 2.89 | 0.004 |
| LH_Vis_6; Left Primary Visual Cortex | -12 | -67 | 7 | RH_Limbic_OFC_1; Right Orbitofrontal Cortex | 12 | 35 | -20 | 2.88 | 0.004 |
| RH_Limbic_OFC_1; Right Orbitofrontal Cortex | 12 | 35 | -20 | RH_Cont_pCun_1; Right Visual Motor Cortex | 9 | -66 | 43 | 2.88 | 0.005 |
| LH_SalVentAttn_PFCl_1; Left Anterior Prefrontal Cortex | -30 | 44 | 30 | RH_Default_PFCdPFCm_2; Right Dorsolateral Prefrontal Cortex | 11 | 50 | 39 | 2.88 | 0.005 |
| LH_Vis_4; Left Secondary Visual Cortex | -27 | -95 | -4 | LH_Limbic_OFC_1; Left Orbitofrontal Cortex | -14 | 32 | -20 | 2.88 | 0.005 |
| LH_SomMot_5; Left Primary Motor Cortex | -39 | -23 | 59 | LH_SomMot_6; Left Primary Motor Cortex | -6 | -29 | 70 | 2.86 | 0.005 |
| LH_Default_PFC_7; Left Frontal Eye Fields | -25 | 20 | 51 | RH_Default_pCunPCC_2; Right Ventral Posterior Cingulate Cortex | 7 | -52 | 31 | 2.86 | 0.005 |
| LH_SomMot_6; Left Primary Motor Cortex | -6 | -29 | 70 | LH_SalVentAttn_ParOper_1; Left Supramarginal Gyrus | -59 | -38 | 29 | 2.83 | 0.005 |
| RH_SalVentAttn_TempOccPar_2; Right Supramarginal Gyrus | 61 | -26 | 27 | LH_Default_PFC_6; Left PreMotor & Supplementary Motor Area | -41 | 14 | 48 | 2.82 | 0.005 |
| LH_SomMot_5; Left Primary Motor Cortex | -39 | -23 | 59 | RH_Cont_PFCl_1; Right Anterior Prefrontal Cortex | 30 | 58 | -3 | 2.81 | 0.006 |
| LH_DorsAttn_Post_3; Left Visual Motor Cortex | -24 | -68 | 49 | RH_Limbic_OFC_1; Right Orbitofrontal Cortex | 12 | 35 | -20 | 2.81 | 0.006 |
| LH_DorsAttn_Post_4; Left Primary Sensory Cortex | -42 | -34 | 48 | RH_Cont_pCun_1; Right Visual Motor Cortex | 9 | -66 | 43 | 2.80 | 0.006 |
| RH_DorsAttn_Post_4; Right Visual Motor Cortex | 27 | -67 | 51 | LH_Limbic_TempPole_1; Left Temporal Pole | -32 | 2 | -37 | 2.79 | 0.006 |
| LH_Default_pCunPCC_2; Left Ventral Posterior Cingulate Cortex | -6 | -53 | 33 | RH_Default_PFCv_1; Right Pars Orbitalis | 35 | 26 | -15 | 2.79 | 0.006 |
| RH_Vis_3; Right Fusiform Cortex | 49 | -60 | -11 | RH_Cont_pCun_1; Right Visual Motor Cortex | 9 | -66 | 43 | 2.79 | 0.006 |

| | | | | | | | | | |
|---|---|---|---|---|---|---|---|---|---|
| LH_Vis_4; Left Secondary Visual Cortex | -27 | -95 | -4 | LH_DorsAttn_Post_1; Left Fusiform Cortex | -47 | -58 | -13 | 2.78 | 0.006 |
| RH_Vis_6; Right Secondary Visual Cortex | 17 | -57 | 5 | LH_DorsAttn_Post_5; Left Visual Motor Cortex | -6 | -60 | 56 | 2.77 | 0.006 |
| LH_Vis_9; Left Visual Association Cortex | -6 | -82 | 26 | LH_SalVentAttn_Med_2; Left Dorsal Posterior Cingulate Cortex | -11 | -34 | 45 | 2.77 | 0.006 |
| LH_Default_PFC_7; Left Frontal Eye Fields | -25 | 20 | 51 | RH_Default_pCunPCC_1; Right Ventral Posterior Cingulate Cortex | 12 | -54 | 14 | 2.76 | 0.006 |
| LH_DorsAttn_FEF_1; Left PreMotor & Supplementary Motor Area | -26 | -3 | 59 | RH_DorsAttn_Post_3; Right Visual Motor Cortex | 38 | -45 | 49 | 2.76 | 0.006 |
| LH_DorsAttn_Post_4; Left Primary Sensory Cortex | -42 | -34 | 48 | RH_SalVentAttn_Med_2; Right PreMotor & Supplementary Motor Area | 7 | 6 | 52 | 2.76 | 0.006 |
| LH_SalVentAttn_FrOperIns_1; Left Insula | -41 | -1 | -7 | RH_Cont_PFCl_3; Right Dorsolateral Prefrontal Cortex | 32 | 46 | 29 | 2.75 | 0.007 |
| RH_Vis_3; Right Fusiform Cortex | 49 | -60 | -11 | LH_Cont_Cing_1; Left Ventral Posterior Cingulate Cortex | -4 | -26 | 33 | 2.75 | 0.007 |
| RH_Cont_Par_1; Right Supramarginal Gyrus | 57 | -39 | 44 | LH_Default_PFC_6; Left PreMotor & Supplementary Motor Area | -41 | 14 | 48 | 2.74 | 0.007 |
| LH_Vis_5; Left Secondary Visual Cortex | -5 | -92 | -2 | LH_Limbic_TempPole_1; Left Temporal Pole | -32 | 2 | -37 | 2.73 | 0.007 |
| LH_Default_pCunPCC_2; Left Ventral Posterior Cingulate Cortex | -6 | -53 | 33 | RH_Default_pCunPCC_2; Right Ventral Posterior Cingulate Cortex | 7 | -52 | 31 | 2.73 | 0.007 |
| LH_SalVentAttn_Med_1; Left Dorsal Anterior Cingulate Cortex | -5 | 20 | 34 | RH_Default_Par_1; Right Angular Gyrus | 55 | -51 | 31 | 2.73 | 0.007 |
| RH_Vis_3; Right Fusiform Cortex | 49 | -60 | -11 | LH_DorsAttn_FEF_1; Left PreMotor & Supplementary Motor Area | -26 | -3 | 59 | 2.73 | 0.007 |
| LH_Cont_Cing_1; Left Ventral Posterior Cingulate Cortex | -4 | -26 | 33 | RH_Default_PFCv_1; Right Pars Orbitalis | 35 | 26 | -15 | 2.73 | 0.007 |
| LH_SalVentAttn_PFCl_1; Left Anterior Prefrontal Cortex | -30 | 44 | 30 | LH_SalVentAttn_Med_1; Left Dorsal Anterior Cingulate Cortex | -5 | 20 | 34 | 2.71 | 0.008 |
| LH_SalVentAttn_Med_2; Left Dorsal Posterior Cingulate Cortex | -11 | -34 | 45 | RH_Limbic_OFC_1; Right Orbitofrontal Cortex | 12 | 35 | -20 | 2.71 | 0.008 |
| LH_Vis_6; Left Primary Visual Cortex | -12 | -67 | 7 | LH_Cont_PFCl_1; Left Dorsolateral Prefrontal Cortex | -43 | 33 | 21 | 2.68 | 0.008 |
| RH_Vis_2; Right Fusiform Cortex | 27 | -66 | -12 | RH_Cont_PFCl_1; Right Anterior Prefrontal Cortex | 30 | 58 | -3 | -2.68 | 0.008 |
| RH_Cont_Cing_1; Right Ventral Posterior Cingulate Cortex | 5 | -27 | 33 | RH_Default_PFCv_1; Right Pars Orbitalis | 35 | 26 | -15 | 2.68 | 0.008 |
| LH_DorsAttn_Post_4; Left Primary Sensory Cortex | -42 | -34 | 48 | RH_DorsAttn_Post_5; Right Visual Motor Cortex | 14 | -52 | 66 | 2.68 | 0.008 |
| LH_SomMot_2; Left Primary Sensory Cortex | -37 | -21 | 16 | RH_SalVentAttn_TempOccPar_2; Right Supramarginal Gyrus | 61 | -26 | 27 | 2.67 | 0.008 |
| LH_Vis_8; Left Visual Association Cortex | -25 | -88 | 20 | LH_SalVentAttn_Med_2; Left Dorsal Posterior Cingulate Cortex | -11 | -34 | 45 | 2.67 | 0.008 |
| RH_SomMot_7; Right Primary Sensory Cortex | 30 | -37 | 64 | LH_Default_pCunPCC_2; Left Ventral Posterior Cingulate Cortex | -6 | -53 | 33 | 2.67 | 0.008 |
| LH_Limbic_OFC_1; Left Orbitofrontal Cortex | -14 | 32 | -20 | RH_Default_pCunPCC_2; Right Ventral Posterior Cingulate Cortex | 7 | -52 | 31 | 2.67 | 0.008 |
| LH_DorsAttn_Post_5; Left Visual Motor Cortex | -6 | -60 | 56 | RH_Default_pCunPCC_1; Right Ventral Posterior Cingulate Cortex | 12 | -54 | 14 | 2.66 | 0.009 |
| RH_DorsAttn_Post_4; Right Visual Motor Cortex | 27 | -67 | 51 | RH_Cont_Par_1; Right Supramarginal Gyrus | 57 | -39 | 44 | 2.65 | 0.009 |
| LH_SomMot_2; Left Primary Sensory Cortex | -37 | -21 | 16 | RH_Default_Par_1; Right Angular Gyrus | 55 | -51 | 31 | 2.65 | 0.009 |
| RH_DorsAttn_Post_5; Right Visual Motor Cortex | 14 | -52 | 66 | LH_SalVentAttn_Med_2; Left Dorsal Posterior Cingulate Cortex | -11 | -34 | 45 | 2.65 | 0.009 |
| LH_DorsAttn_Post_5; Left Visual Motor Cortex | -6 | -60 | 56 | LH_Cont_Par_1; Left Angular Gyrus | -37 | -53 | 46 | 2.64 | 0.009 |
| RH_DorsAttn_Post_5; Right Visual Motor Cortex | 14 | -52 | 66 | LH_SalVentAttn_ParOper_1; Left Supramarginal Gyrus | -59 | -38 | 29 | 2.64 | 0.009 |
| LH_DorsAttn_FEF_1; Left PreMotor & Supplementary Motor Area | -26 | -3 | 59 | RH_Default_pCunPCC_1; Right Ventral Posterior Cingulate Cortex | 12 | -54 | 14 | 2.64 | 0.009 |

| | | | | | | | | | |
|---|---|---|---|---|---|---|---|---|---|
| LH_SomMot_2; Left Primary Sensory Cortex | -37 | -21 | 16 | LH_SomMot_6; Left Primary Motor Cortex | -6 | -29 | 70 | 2.64 | 0.009 |
| LH_Vis_4; Left Secondary Visual Cortex | -27 | -95 | -4 | LH_Limbic_TempPole_1; Left Temporal Pole | -32 | 2 | -37 | 2.64 | 0.009 |
| RH_Default_pCunPCC_1; Right Ventral Posterior Cingulate Cortex | 12 | -54 | 14 | RH_Default_pCunPCC_2; Right Ventral Posterior Cingulate Cortex | 7 | -52 | 31 | 2.63 | 0.009 |
| RH_DorsAttn_Post_5; Right Visual Motor Cortex | 14 | -52 | 66 | LH_Default_PFC_6; Left PreMotor & Supplementary Motor Area | -41 | 14 | 48 | 2.63 | 0.009 |
| LH_Vis_9; Left Visual Association Cortex | -6 | -82 | 26 | RH_Vis_6; Right Secondary Visual Cortex | 17 | -57 | 5 | 2.63 | 0.009 |
| RH_DorsAttn_Post_1; Right Visual Association Cortex | 50 | -62 | 16 | LH_Default_PFC_1; Left Pars Orbitalis | -35 | 21 | -11 | 2.62 | 0.010 |
| LH_SomMot_6; Left Primary Motor Cortex | -6 | -29 | 70 | RH_Default_PFCdPFCm_2; Right Dorsolateral Prefrontal Cortex | 11 | 50 | 39 | 2.61 | 0.010 |
| LH_Vis_4; Left Secondary Visual Cortex | -27 | -95 | -4 | LH_SalVentAttn_Med_2; Left Dorsal Posterior Cingulate Cortex | -11 | -34 | 45 | 2.61 | 0.010 |
| RH_DorsAttn_Post_4; Right Visual Motor Cortex | 27 | -67 | 51 | LH_Limbic_OFC_1; Left Orbitofrontal Cortex | -14 | 32 | -20 | 2.61 | 0.010 |
| RH_SalVentAttn_TempOccPar_2; Right Supramarginal Gyrus | 61 | -26 | 27 | LH_Limbic_OFC_1; Left Orbitofrontal Cortex | -14 | 32 | -20 | 2.60 | 0.010 |
| LH_Vis_7; Left Visual Association Cortex | -47 | -71 | 11 | LH_Cont_Cing_1; Left Ventral Posterior Cingulate Cortex | -4 | -26 | 33 | 2.60 | 0.010 |
| LH_DorsAttn_Post_2; Left Supramarginal Gyrus | -57 | -25 | 39 | RH_DorsAttn_Post_5; Right Visual Motor Cortex | 14 | -52 | 66 | 2.60 | 0.010 |
| RH_SalVentAttn_Med_1; Right Dorsal Posterior Cingulate Cortex | 11 | -31 | 45 | RH_Default_PFCv_1; Right Pars Orbitalis | 35 | 26 | -15 | 2.59 | 0.010 |

**Eyes-Open Gamma**

| **ROI1 Labels (Schaefer; Talairach)** | **X** | **Y** | **Z** | **ROI2 Labels (Schaefer; Talairach)** | **X** | **Y** | **Z** | **t** | **p (unc)** |
|---|---|---|---|---|---|---|---|---|---|
| LH_SomMot_2; Left Primary Sensory Cortex | -37 | -21 | 16 | RH_Limbic_OFC_1; Right Orbitofrontal Cortex | 12 | 35 | -20 | -3.72 | 0.0003 |
| LH_Vis_4; Left Secondary Visual Cortex | -27 | -95 | -4 | LH_DorsAttn_Post_4; Left Primary Sensory Cortex | -42 | -34 | 48 | -3.64 | 0.0004 |
| LH_Cont_PFCl_1; Left Dorsolateral Prefrontal Cortex | -43 | 33 | 21 | RH_Default_pCunPCC_1; Right Ventral Posterior Cingulate Cortex | 12 | -54 | 14 | -3.53 | 0.0006 |
| RH_Cont_Cing_1; Right Ventral Posterior Cingulate Cortex | 5 | -27 | 33 | LH_Default_PFC_3; Left Dorsal Posterior Cingulate Cortex | -6 | 47 | 0 | -3.52 | 0.0006 |
| RH_Default_Temp_2; Right Temporal Pole | 51 | 7 | -18 | RH_Default_PFCdPFCm_2; Right Dorsolateral Prefrontal Cortex | 11 | 50 | 39 | -3.41 | 0.0008 |
| LH_Cont_PFCl_1; Left Dorsolateral Prefrontal Cortex | -43 | 33 | 21 | RH_Default_Temp_1; Right Medial Temporal Gyrus | 62 | -23 | -19 | -3.31 | 0.001 |
| RH_SomMot_2; Right Insula | 40 | -15 | 15 | LH_DorsAttn_Post_5; Left Visual Motor Cortex | -6 | -60 | 56 | -3.25 | 0.001 |
| LH_Cont_Cing_1; Left Ventral Posterior Cingulate Cortex | -4 | -26 | 33 | RH_Default_PFCdPFCm_1; Right Anterior Prefrontal Cortex | 7 | 48 | 1 | -3.16 | 0.002 |
| LH_DorsAttn_FEF_1; Left PreMotor & Supplementary Motor Area | -26 | -3 | 59 | LH_SalVentAttn_PFCl_1; Left Anterior Prefrontal Cortex | -30 | 44 | 30 | -3.08 | 0.002 |
| RH_Cont_Cing_1; Right Ventral Posterior Cingulate Cortex | 5 | -27 | 33 | RH_Cont_pCun_1; Right Visual Motor Cortex | 9 | -66 | 43 | -3.08 | 0.002 |
| RH_Vis_8; Right Visual Association Cortex | 13 | -86 | 29 | RH_DorsAttn_Post_4; Right Visual Motor Cortex | 27 | -67 | 51 | 3.05 | 0.003 |
| LH_Cont_Cing_1; Left Ventral Posterior Cingulate Cortex | -4 | -26 | 33 | LH_Default_pCunPCC_2; Left Ventral Posterior Cingulate Cortex | -6 | -53 | 33 | -3.04 | 0.003 |
| RH_SomMot_3; Right Primary Motor Cortex | 57 | -4 | 11 | RH_Default_PFCdPFCm_3; Right Frontal Eye Fields | 26 | 24 | 50 | -2.99 | 0.003 |
| RH_Cont_Cing_1; Right Ventral Posterior Cingulate Cortex | 5 | -27 | 33 | RH_Default_PFCdPFCm_1; Right Anterior Prefrontal Cortex | 7 | 48 | 1 | -2.98 | 0.003 |
| RH_DorsAttn_Post_4; Right Visual Motor Cortex | 27 | -67 | 51 | RH_SalVentAttn_Med_2; Right PreMotor & Supplementary Motor Area | 7 | 6 | 52 | -2.97 | 0.003 |
| RH_SalVentAttn_TempOccPar_2; Right Supramarginal Gyrus | 61 | -26 | 27 | LH_Default_PFC_2; Left Pars Orbitalis | -47 | 33 | -3 | -2.96 | 0.004 |

| | | | | | | | | | |
|---|---|---|---|---|---|---|---|---|---|
| LH_SalVentAttn_PFCl_1; Left Anterior Prefrontal Cortex | -30 | 44 | 30 | LH_Default_PFC_6; Left PreMotor & Supplementary Motor Area | -41 | 14 | 48 | -2.95 | 0.004 |
| LH_Cont_PFCl_1; Left Dorsolateral Prefrontal Cortex | -43 | 33 | 21 | RH_Default_Temp_3; Right Superior Temporal Gyrus | 57 | -26 | -2 | -2.94 | 0.004 |
| RH_Default_Temp_2; Right Temporal Pole | 51 | 7 | -18 | RH_Default_PFCdPFCm_3; Right Frontal Eye Fields | 26 | 24 | 50 | -2.94 | 0.004 |
| LH_Cont_Cing_1; Left Ventral Posterior Cingulate Cortex | -4 | -26 | 33 | RH_Cont_pCun_1; Right Visual Motor Cortex | 9 | -66 | 43 | -2.91 | 0.004 |
| LH_DorsAttn_Post_3; Left Visual Motor Cortex | -24 | -68 | 49 | RH_SalVentAttn_TempOccPar_2; Right Supramarginal Gyrus | 61 | -26 | 27 | -2.89 | 0.004 |
| LH_Default_Par_1; Left Angular Gyrus | -57 | -50 | 12 | RH_Default_Temp_2; Right Temporal Pole | 51 | 7 | -18 | -2.89 | 0.004 |
| LH_Vis_1; Left Fusiform Cortex | -26 | -34 | -17 | LH_Default_Par_1; Left Angular Gyrus | -57 | -50 | 12 | -2.83 | 0.005 |
| RH_Vis_2; Right Fusiform Cortex | 27 | -66 | -12 | LH_Cont_PFCl_1; Left Dorsolateral Prefrontal Cortex | -43 | 33 | 21 | -2.81 | 0.006 |
| RH_Cont_PFCmp_1; Right Frontal Eye Fields | 6 | 28 | 30 | RH_Default_Temp_2; Right Temporal Pole | 51 | 7 | -18 | -2.80 | 0.006 |
| LH_Limbic_TempPole_1; Left Temporal Pole | -32 | 2 | -37 | RH_Cont_PFCl_1; Right Anterior Prefrontal Cortex | 30 | 58 | -3 | -2.80 | 0.006 |
| RH_SomMot_1; Right Primary Auditory Cortex | 53 | -16 | 7 | RH_Default_PFCdPFCm_3; Right Frontal Eye Fields | 26 | 24 | 50 | -2.80 | 0.006 |
| RH_Vis_6; Right Secondary Visual Cortex | 17 | -57 | 5 | LH_Cont_PFCl_1; Left Dorsolateral Prefrontal Cortex | -43 | 33 | 21 | -2.80 | 0.006 |
| LH_SalVentAttn_Med_2; Left Dorsal Posterior Cingulate Cortex | -11 | -34 | 45 | LH_Default_PFC_3; Left Dorsal Posterior Cingulate Cortex | -6 | 47 | 0 | -2.79 | 0.006 |
| LH_SomMot_2; Left Primary Sensory Cortex | -37 | -21 | 16 | RH_Cont_PFCl_2; Right Dorsolateral Prefrontal Cortex | 45 | 39 | 15 | -2.78 | 0.006 |
| RH_Cont_PFCl_2; Right Dorsolateral Prefrontal Cortex | 45 | 39 | 15 | LH_Default_Temp_1; Left Medial Temporal Gyrus | -55 | -4 | -20 | -2.77 | 0.006 |
| RH_DorsAttn_Post_4; Right Visual Motor Cortex | 27 | -67 | 51 | RH_DorsAttn_FEF_1; Right PreMotor & Supplementary Motor Area | 28 | -3 | 59 | -2.76 | 0.007 |
| LH_SalVentAttn_FrOperIns_2; Left Insula | -38 | 12 | 6 | RH_Default_Temp_3; Right Superior Temporal Gyrus | 57 | -26 | -2 | -2.75 | 0.007 |
| LH_DorsAttn_Post_6; Left Visual Motor Cortex | -22 | -51 | 66 | LH_Default_PFC_2; Left Pars Orbitalis | -47 | 33 | -3 | -2.75 | 0.007 |
| LH_DorsAttn_PrCv_1; Left PreMotor & Supplementary Motor Area | -48 | 6 | 28 | LH_Default_PFC_1; Left Pars Orbitalis | -35 | 21 | -11 | 2.74 | 0.007 |
| LH_Cont_Cing_1; Left Ventral Posterior Cingulate Cortex | -4 | -26 | 33 | LH_Default_PFC_3; Left Dorsal Posterior Cingulate Cortex | -6 | 47 | 0 | -2.74 | 0.007 |
| RH_Limbic_OFC_1; Right Orbitofrontal Cortex | 12 | 35 | -20 | LH_Default_PFC_6; Left PreMotor & Supplementary Motor Area | -41 | 14 | 48 | -2.73 | 0.007 |
| RH_SomMot_5; Right PreMotor & Supplementary Motor Area | 47 | -11 | 48 | LH_Default_PFC_1; Left Pars Orbitalis | -35 | 21 | -11 | -2.70 | 0.008 |
| LH_Vis_8; Left Visual Association Cortex | -25 | -88 | 20 | LH_DorsAttn_Post_5; Left Visual Motor Cortex | -6 | -60 | 56 | -2.69 | 0.008 |
| LH_Limbic_TempPole_1; Left Temporal Pole | -32 | 2 | -37 | RH_Cont_PFCl_2; Right Dorsolateral Prefrontal Cortex | 45 | 39 | 15 | -2.68 | 0.008 |
| LH_Vis_1; Left Fusiform Cortex | -26 | -34 | -17 | LH_Default_pCunPCC_2; Left Ventral Posterior Cingulate Cortex | -6 | -53 | 33 | -2.67 | 0.008 |
| RH_SomMot_4; Right Primary Motor Cortex | 58 | -5 | 31 | RH_Default_Temp_3; Right Superior Temporal Gyrus | 57 | -26 | -2 | 2.66 | 0.009 |
| LH_SomMot_4; Left PreMotor & Supplementary Motor Area | -55 | -8 | 34 | RH_DorsAttn_Post_1; Right Visual Association Cortex | 50 | -62 | 16 | -2.65 | 0.009 |
| LH_Vis_5; Left Secondary Visual Cortex | -5 | -92 | -2 | RH_DorsAttn_Post_3; Right Visual Motor Cortex | 38 | -45 | 49 | 2.65 | 0.009 |
| RH_DorsAttn_FEF_1; Right PreMotor & Supplementary Motor Area | 28 | -3 | 59 | LH_SalVentAttn_FrOperIns_1; Left Insula | -41 | -1 | -7 | -2.64 | 0.009 |
| LH_SomMot_2; Left Primary Sensory Cortex | -37 | -21 | 16 | RH_Default_PFCv_2; Right Broca's Area/ Pars Triangularis | 51 | 28 | 0 | -2.64 | 0.009 |
| RH_Default_Par_1; Right Angular Gyrus | 55 | -51 | 31 | RH_Default_pCunPCC_2; Right Ventral Posterior Cingulate Cortex | 7 | -52 | 31 | -2.64 | 0.009 |
| RH_DorsAttn_Post_5; Right Visual Motor Cortex | 14 | -52 | 66 | LH_Default_PFC_4; Left Anterior Prefrontal Cortex | -24 | 61 | -1 | -2.63 | 0.010 |

| | | | | | | | | | |
|---|---|---|---|---|---|---|---|---|---|
| LH_DorsAttn_Post_5; Left Visual Motor Cortex | -6 | -60 | 56 | LH_SalVentAttn_FrOperIns_1; Left Insula | -41 | -1 | -7 | -2.62 | 0.010 |
| LH_DorsAttn_PrCv_1; Left PreMotor & Supplementary Motor Area | -48 | 6 | 28 | RH_SalVentAttn_TempOccPar_2; Right Supramarginal Gyrus | 61 | -26 | 27 | -2.62 | 0.010 |
| LH_Limbic_TempPole_1; Left Temporal Pole | -32 | 2 | -37 | LH_Cont_Par_1; Left Angular Gyrus | -37 | -53 | 46 | -2.61 | 0.010 |
| LH_Default_Temp_1; Left Medial Temporal Gyrus | -55 | -4 | -20 | RH_Default_PFCdPFCm_3; Right Frontal Eye Fields | 26 | 24 | 50 | -2.60 | 0.010 |
| LH_Limbic_OFC_1; Left Orbitofrontal Cortex | -14 | 32 | -20 | LH_Cont_Cing_1; Left Ventral Posterior Cingulate Cortex | -4 | -26 | 33 | -2.60 | 0.010 |
| **Eyes-Closed Theta** | | | | | | | | | |
| **ROI1 Labels (Schaefer; Talairach)** | **X** | **Y** | **Z** | **ROI2 Labels (Schaefer; Talairach)** | **X** | **Y** | **Z** | **t** | **p (unc)** |
| RH_SalVentAttn_Med_2; Right PreMotor & Supplementary Motor Area | 7 | 6 | 52 | RH_Cont_Par_2; Right Angular Gyrus | 45 | -63 | 46 | 3.88 | 0.0002 |
| LH_Cont_pCun_1; Left Visual Motor Cortex | -9 | -73 | 38 | LH_Default_PFC_5; Left Dorsolateral Prefrontal Cortex | -9 | 48 | 41 | 3.81 | 0.0002 |
| LH_DorsAttn_Post_5; Left Visual Motor Cortex | -6 | -60 | 56 | LH_Default_PFC_5; Left Dorsolateral Prefrontal Cortex | -9 | 48 | 41 | 3.74 | 0.0003 |
| LH_SomMot_6; Left Primary Motor Cortex | -6 | -29 | 70 | RH_Cont_PFCmp_1; Right Frontal Eye Fields | 6 | 28 | 30 | 3.63 | 0.0004 |
| RH_DorsAttn_FEF_1; Right PreMotor & Supplementary Motor Area | 28 | -3 | 59 | LH_Cont_pCun_1; Left Visual Motor Cortex | -9 | -73 | 38 | 3.61 | 0.0004 |
| RH_SomMot_6; Right Primary Motor Cortex | 41 | -22 | 60 | RH_Cont_PFCl_3; Right Dorsolateral Prefrontal Cortex | 32 | 46 | 29 | 3.55 | 0.0005 |
| LH_DorsAttn_Post_3; Left Visual Motor Cortex | -24 | -68 | 49 | LH_Default_PFC_5; Left Dorsolateral Prefrontal Cortex | -9 | 48 | 41 | 3.55 | 0.0005 |
| LH_DorsAttn_FEF_1; Left PreMotor & Supplementary Motor Area | -26 | -3 | 59 | RH_DorsAttn_Post_3; Right Visual Motor Cortex | 38 | -45 | 49 | 3.54 | 0.0005 |
| LH_SomMot_5; Left Primary Motor Cortex | -39 | -23 | 59 | RH_DorsAttn_Post_4; Right Visual Motor Cortex | 27 | -67 | 51 | 3.53 | 0.0005 |
| RH_SomMot_4; Right Primary Motor Cortex | 58 | -5 | 31 | RH_DorsAttn_FEF_1; Right PreMotor & Supplementary Motor Area | 28 | -3 | 59 | 3.50 | 0.0006 |
| RH_SomMot_6; Right Primary Motor Cortex | 41 | -22 | 60 | RH_Cont_PFCl_2; Right Dorsolateral Prefrontal Cortex | 45 | 39 | 15 | 3.45 | 0.0007 |
| LH_SomMot_5; Left Primary Motor Cortex | -39 | -23 | 59 | LH_SalVentAttn_Med_2; Left Dorsal Posterior Cingulate Cortex | -11 | -34 | 45 | 3.34 | 0.001 |
| LH_SomMot_6; Left Primary Motor Cortex | -6 | -29 | 70 | RH_Default_PFCdPFCm_1; Right Anterior Prefrontal Cortex | 7 | 48 | 1 | 3.30 | 0.001 |
| RH_Cont_pCun_1; Right Visual Motor Cortex | 9 | -66 | 43 | LH_Default_PFC_5; Left Dorsolateral Prefrontal Cortex | -9 | 48 | 41 | 3.21 | 0.002 |
| RH_DorsAttn_Post_4; Right Visual Motor Cortex | 27 | -67 | 51 | RH_SalVentAttn_Med_2; Right PreMotor & Supplementary Motor Area | 7 | 6 | 52 | 3.20 | 0.002 |
| RH_Cont_Cing_1; Right Ventral Posterior Cingulate Cortex | 5 | -27 | 33 | RH_Default_PFCdPFCm_3; Right Frontal Eye Fields | 26 | 24 | 50 | 3.17 | 0.002 |
| LH_SomMot_5; Left Primary Motor Cortex | -39 | -23 | 59 | RH_Cont_pCun_1; Right Visual Motor Cortex | 9 | -66 | 43 | 3.14 | 0.002 |
| LH_DorsAttn_Post_3; Left Visual Motor Cortex | -24 | -68 | 49 | RH_Cont_PFCmp_1; Right Frontal Eye Fields | 6 | 28 | 30 | 3.07 | 0.003 |
| LH_Vis_9; Left Visual Association Cortex | -6 | -82 | 26 | RH_SalVentAttn_Med_2; Right PreMotor & Supplementary Motor Area | 7 | 6 | 52 | 3.06 | 0.003 |
| RH_Cont_PFCl_3; Right Dorsolateral Prefrontal Cortex | 32 | 46 | 29 | RH_Default_PFCdPFCm_3; Right Frontal Eye Fields | 26 | 24 | 50 | 3.03 | 0.003 |
| RH_SalVentAttn_Med_2; Right PreMotor & Supplementary Motor Area | 7 | 6 | 52 | LH_Cont_pCun_1; Left Visual Motor Cortex | -9 | -73 | 38 | 2.99 | 0.003 |
| LH_Vis_3; Left Visual Association Cortex | -17 | -60 | -7 | RH_SomMot_6; Right Primary Motor Cortex | 41 | -22 | 60 | 2.99 | 0.003 |
| RH_SalVentAttn_Med_2; Right PreMotor & Supplementary Motor Area | 7 | 6 | 52 | RH_Cont_pCun_1; Right Visual Motor Cortex | 9 | -66 | 43 | 2.98 | 0.003 |
| RH_DorsAttn_Post_4; Right Visual Motor Cortex | 27 | -67 | 51 | RH_Default_PFCdPFCm_3; Right Frontal Eye Fields | 26 | 24 | 50 | 2.97 | 0.003 |

| | | | | | | | | | |
|---|---|---|---|---|---|---|---|---|---|
| LH_Vis_6; Left Primary Visual Cortex | -12 | -67 | 7 | RH_SomMot_5; Right PreMotor & Supplementary Motor Area | 47 | -11 | 48 | 2.95 | 0.004 |
| LH_Vis_6; Left Primary Visual Cortex | -12 | -67 | 7 | RH_DorsAttn_FEF_1; Right PreMotor & Supplementary Motor Area | 28 | -3 | 59 | 2.95 | 0.004 |
| LH_Vis_7; Left Visual Association Cortex | -47 | -71 | 11 | RH_SalVentAttn_Med_2; Right PreMotor & Supplementary Motor Area | 7 | 6 | 52 | 2.95 | 0.004 |
| RH_SomMot_5; Right PreMotor & Supplementary Motor Area | 47 | -11 | 48 | RH_Cont_PFCl_3; Right Dorsolateral Prefrontal Cortex | 32 | 46 | 29 | 2.94 | 0.004 |
| LH_DorsAttn_Post_5; Left Visual Motor Cortex | -6 | -60 | 56 | RH_Cont_PFCmp_1; Right Frontal Eye Fields | 6 | 28 | 30 | 2.94 | 0.004 |
| LH_DorsAttn_Post_5; Left Visual Motor Cortex | -6 | -60 | 56 | RH_Cont_PFCl_4; Right Frontal Eye Fields | 43 | 16 | 45 | 2.93 | 0.004 |
| LH_DorsAttn_Post_3; Left Visual Motor Cortex | -24 | -68 | 49 | LH_SalVentAttn_PFCl_1; Left Anterior Prefrontal Cortex | -30 | 44 | 30 | 2.92 | 0.004 |
| LH_SomMot_6; Left Primary Motor Cortex | -6 | -29 | 70 | RH_DorsAttn_Post_4; Right Visual Motor Cortex | 27 | -67 | 51 | 2.92 | 0.004 |
| RH_SomMot_2; Right Insula | 40 | -15 | 15 | RH_Default_PFCdPFCm_3; Right Frontal Eye Fields | 26 | 24 | 50 | 2.92 | 0.004 |
| LH_DorsAttn_FEF_1; Left PreMotor & Supplementary Motor Area | -26 | -3 | 59 | RH_Cont_Cing_1; Right Ventral Posterior Cingulate Cortex | 5 | -27 | 33 | 2.92 | 0.004 |
| LH_DorsAttn_FEF_1; Left PreMotor & Supplementary Motor Area | -26 | -3 | 59 | RH_SalVentAttn_Med_1; Right Dorsal Posterior Cingulate Cortex | 11 | -31 | 45 | 2.91 | 0.004 |
| LH_SalVentAttn_Med_3; Left PreMotor & Supplementary Motor Area | -6 | 4 | 62 | RH_Cont_Par_2; Right Angular Gyrus | 45 | -63 | 46 | 2.91 | 0.004 |
| LH_DorsAttn_Post_3; Left Visual Motor Cortex | -24 | -68 | 49 | LH_SalVentAttn_Med_1; Left Dorsal Anterior Cingulate Cortex | -5 | 20 | 34 | 2.90 | 0.004 |
| RH_DorsAttn_Post_3; Right Visual Motor Cortex | 38 | -45 | 49 | RH_SalVentAttn_Med_2; Right PreMotor & Supplementary Motor Area | 7 | 6 | 52 | 2.89 | 0.004 |
| RH_Vis_8; Right Visual Association Cortex | 13 | -86 | 29 | RH_SalVentAttn_Med_2; Right PreMotor & Supplementary Motor Area | 7 | 6 | 52 | 2.88 | 0.005 |
| LH_DorsAttn_FEF_1; Left PreMotor & Supplementary Motor Area | -26 | -3 | 59 | RH_DorsAttn_Post_5; Right Visual Motor Cortex | 14 | -52 | 66 | 2.88 | 0.005 |
| LH_SomMot_6; Left Primary Motor Cortex | -6 | -29 | 70 | LH_Limbic_OFC_1; Left Orbitofrontal Cortex | -14 | 32 | -20 | 2.88 | 0.005 |
| LH_DorsAttn_FEF_1; Left PreMotor & Supplementary Motor Area | -26 | -3 | 59 | RH_DorsAttn_Post_4; Right Visual Motor Cortex | 27 | -67 | 51 | 2.88 | 0.005 |
| RH_DorsAttn_FEF_1; Right PreMotor & Supplementary Motor Area | 28 | -3 | 59 | LH_Default_pCunPCC_2; Left Ventral Posterior Cingulate Cortex | -6 | -53 | 33 | 2.87 | 0.005 |
| RH_DorsAttn_PrCv_1; Right Broca's Area/Operculum | 49 | 10 | 27 | RH_DorsAttn_FEF_1; Right PreMotor & Supplementary Motor Area | 28 | -3 | 59 | 2.87 | 0.005 |
| LH_DorsAttn_Post_5; Left Visual Motor Cortex | -6 | -60 | 56 | LH_SalVentAttn_Med_1; Left Dorsal Anterior Cingulate Cortex | -5 | 20 | 34 | 2.87 | 0.005 |
| RH_SomMot_8; Right Primary Motor Cortex | 6 | -26 | 70 | RH_Default_PFCdPFCm_1; Right Anterior Prefrontal Cortex | 7 | 48 | 1 | 2.85 | 0.005 |
| RH_SalVentAttn_Med_2; Right PreMotor & Supplementary Motor Area | 7 | 6 | 52 | RH_Default_Par_1; Right Angular Gyrus | 55 | -51 | 31 | 2.85 | 0.005 |
| RH_SalVentAttn_Med_2; Right PreMotor & Supplementary Motor Area | 7 | 6 | 52 | RH_Cont_Cing_1; Right Ventral Posterior Cingulate Cortex | 5 | -27 | 33 | 2.84 | 0.005 |
| LH_Vis_9; Left Visual Association Cortex | -6 | -82 | 26 | LH_Default_PFC_5; Left Dorsolateral Prefrontal Cortex | -9 | 48 | 41 | 2.84 | 0.005 |
| LH_DorsAttn_Post_6; Left Visual Motor Cortex | -22 | -51 | 66 | RH_SalVentAttn_Med_2; Right PreMotor & Supplementary Motor Area | 7 | 6 | 52 | 2.82 | 0.005 |
| RH_DorsAttn_Post_3; Right Visual Motor Cortex | 38 | -45 | 49 | RH_Default_PFCdPFCm_3; Right Frontal Eye Fields | 26 | 24 | 50 | 2.80 | 0.006 |
| LH_Vis_7; Left Visual Association Cortex | -47 | -71 | 11 | RH_DorsAttn_FEF_1; Right PreMotor & Supplementary Motor Area | 28 | -3 | 59 | 2.80 | 0.006 |
| RH_SalVentAttn_Med_1; Right Dorsal Posterior Cingulate Cortex | 11 | -31 | 45 | LH_Default_PFC_6; Left PreMotor & Supplementary Motor Area | -41 | 14 | 48 | 2.80 | 0.006 |
| LH_SalVentAttn_Med_2; Left Dorsal Posterior Cingulate Cortex | -11 | -34 | 45 | LH_Default_PFC_6; Left PreMotor & Supplementary Motor Area | -41 | 14 | 48 | 2.79 | 0.006 |

| | | | | | | | | | |
|---|---|---|---|---|---|---|---|---|---|
| RH_DorsAttn_FEF_1; Right PreMotor & Supplementary Motor Area | 28 | -3 | 59 | RH_Cont_PFCl_3; Right Dorsolateral Prefrontal Cortex | 32 | 46 | 29 | 2.79 | 0.006 |
| LH_Vis_7; Left Visual Association Cortex | -47 | -71 | 11 | LH_DorsAttn_Post_5; Left Visual Motor Cortex | -6 | -60 | 56 | 2.78 | 0.006 |
| LH_DorsAttn_Post_5; Left Visual Motor Cortex | -6 | -60 | 56 | LH_Limbic_OFC_1; Left Orbitofrontal Cortex | -14 | 32 | -20 | 2.77 | 0.006 |
| LH_Vis_5; Left Secondary Visual Cortex | -5 | -92 | -2 | RH_SalVentAttn_Med_2; Right PreMotor & Supplementary Motor Area | 7 | 6 | 52 | 2.76 | 0.006 |
| LH_DorsAttn_Post_3; Left Visual Motor Cortex | -24 | -68 | 49 | RH_SalVentAttn_Med_2; Right PreMotor & Supplementary Motor Area | 7 | 6 | 52 | 2.76 | 0.006 |
| RH_DorsAttn_FEF_1; Right PreMotor & Supplementary Motor Area | 28 | -3 | 59 | LH_Default_pCunPCC_1; Left Ventral Posterior Cingulate Cortex | -11 | -56 | 13 | 2.76 | 0.006 |
| LH_Default_PFC_5; Left Dorsolateral Prefrontal Cortex | -9 | 48 | 41 | LH_Default_pCunPCC_2; Left Ventral Posterior Cingulate Cortex | -6 | -53 | 33 | 2.76 | 0.006 |
| LH_Vis_7; Left Visual Association Cortex | -47 | -71 | 11 | RH_Default_PFCdPFCm_3; Right Frontal Eye Fields | 26 | 24 | 50 | 2.74 | 0.007 |
| RH_DorsAttn_FEF_1; Right PreMotor & Supplementary Motor Area | 28 | -3 | 59 | RH_Default_pCunPCC_1; Right Ventral Posterior Cingulate Cortex | 12 | -54 | 14 | 2.73 | 0.007 |
| RH_DorsAttn_FEF_1; Right PreMotor & Supplementary Motor Area | 28 | -3 | 59 | RH_Default_pCunPCC_2; Right Ventral Posterior Cingulate Cortex | 7 | -52 | 31 | 2.72 | 0.007 |
| LH_DorsAttn_Post_6; Left Visual Motor Cortex | -22 | -51 | 66 | LH_SalVentAttn_Med_1; Left Dorsal Anterior Cingulate Cortex | -5 | 20 | 34 | 2.72 | 0.007 |
| RH_SomMot_8; Right Primary Motor Cortex | 6 | -26 | 70 | RH_Cont_PFCmp_1; Right Frontal Eye Fields | 6 | 28 | 30 | 2.71 | 0.008 |
| RH_Vis_1; Right Fusiform Cortex | 32 | -31 | -22 | LH_Cont_pCun_1; Left Visual Motor Cortex | -9 | -73 | 38 | 2.70 | 0.008 |
| LH_SomMot_6; Left Primary Motor Cortex | -6 | -29 | 70 | LH_SalVentAttn_Med_1; Left Dorsal Anterior Cingulate Cortex | -5 | 20 | 34 | 2.70 | 0.008 |
| LH_Cont_Cing_1; Left Ventral Posterior Cingulate Cortex | -4 | -26 | 33 | RH_Default_PFCdPFCm_3; Right Frontal Eye Fields | 26 | 24 | 50 | 2.69 | 0.008 |
| RH_SomMot_6; Right Primary Motor Cortex | 41 | -22 | 60 | LH_Default_pCunPCC_1; Left Ventral Posterior Cingulate Cortex | -11 | -56 | 13 | 2.69 | 0.008 |
| RH_DorsAttn_FEF_1; Right PreMotor & Supplementary Motor Area | 28 | -3 | 59 | RH_Cont_Par_2; Right Angular Gyrus | 45 | -63 | 46 | 2.69 | 0.008 |
| RH_SalVentAttn_FrOperIns_1; Right Insula | 40 | 8 | 1 | RH_Default_PFCdPFCm_3; Right Frontal Eye Fields | 26 | 24 | 50 | 2.68 | 0.008 |
| RH_Cont_PFCl_2; Right Dorsolateral Prefrontal Cortex | 45 | 39 | 15 | RH_Default_PFCdPFCm_3; Right Frontal Eye Fields | 26 | 24 | 50 | 2.68 | 0.008 |
| LH_DorsAttn_FEF_1; Left PreMotor & Supplementary Motor Area | -26 | -3 | 59 | RH_Cont_Par_2; Right Angular Gyrus | 45 | -63 | 46 | 2.68 | 0.008 |
| RH_Cont_PFCmp_1; Right Frontal Eye Fields | 6 | 28 | 30 | RH_Cont_pCun_1; Right Visual Motor Cortex | 9 | -66 | 43 | 2.67 | 0.008 |
| RH_Cont_PFCl_2; Right Dorsolateral Prefrontal Cortex | 45 | 39 | 15 | LH_Default_Temp_2; Left Medial Temporal Gyrus | -58 | -32 | -1 | -2.66 | 0.009 |
| RH_DorsAttn_Post_2; Right Primary Sensory Cortex | 50 | -24 | 42 | RH_Cont_PFCl_4; Right Frontal Eye Fields | 43 | 16 | 45 | 2.66 | 0.009 |
| RH_DorsAttn_Post_4; Right Visual Motor Cortex | 27 | -67 | 51 | RH_SalVentAttn_Med_1; Right Dorsal Posterior Cingulate Cortex | 11 | -31 | 45 | 2.66 | 0.009 |
| RH_SalVentAttn_Med_2; Right PreMotor & Supplementary Motor Area | 7 | 6 | 52 | RH_Cont_PFCl_2; Right Dorsolateral Prefrontal Cortex | 45 | 39 | 15 | 2.66 | 0.009 |
| LH_DorsAttn_Post_5; Left Visual Motor Cortex | -6 | -60 | 56 | RH_SalVentAttn_Med_2; Right PreMotor & Supplementary Motor Area | 7 | 6 | 52 | 2.66 | 0.009 |
| RH_Vis_8; Right Visual Association Cortex | 13 | -86 | 29 | RH_DorsAttn_FEF_1; Right PreMotor & Supplementary Motor Area | 28 | -3 | 59 | 2.66 | 0.009 |
| LH_Cont_Par_1; Left Angular Gyrus | -37 | -53 | 46 | LH_Default_Par_2; Left Angular Gyrus | -48 | -63 | 35 | 2.65 | 0.009 |
| RH_Cont_PFCl_3; Right Dorsolateral Prefrontal Cortex | 32 | 46 | 29 | RH_Default_Par_1; Right Angular Gyrus | 55 | -51 | 31 | 2.64 | 0.009 |
| LH_SalVentAttn_Med_2; Left Dorsal Posterior Cingulate Cortex | -11 | -34 | 45 | LH_Limbic_OFC_1; Left Orbitofrontal Cortex | -14 | 32 | -20 | 2.64 | 0.009 |
| RH_DorsAttn_Post_4; Right Visual Motor Cortex | 27 | -67 | 51 | RH_Cont_PFCl_3; Right Dorsolateral Prefrontal Cortex | 32 | 46 | 29 | 2.64 | 0.009 |

| | | | | | | | | | |
|---|---|---|---|---|---|---|---|---|---|
| LH_DorsAttn_Post_6; Left Visual Motor Cortex | -22 | -51 | 66 | RH_Cont_PFCmp_1; Right Frontal Eye Fields | 6 | 28 | 30 | 2.64 | 0.009 |
| LH_SomMot_5; Left Primary Motor Cortex | -39 | -23 | 59 | LH_DorsAttn_Post_5; Left Visual Motor Cortex | -6 | -60 | 56 | 2.63 | 0.009 |
| LH_DorsAttn_Post_6; Left Visual Motor Cortex | -22 | -51 | 66 | RH_DorsAttn_Post_5; Right Visual Motor Cortex | 14 | -52 | 66 | 2.63 | 0.010 |
| RH_SomMot_3; Right Primary Motor Cortex | 57 | -4 | 11 | RH_SalVentAttn_TempOccPar_2; Right Supramarginal Gyrus | 61 | -26 | 27 | 2.62 | 0.010 |
| LH_Vis_9; Left Visual Association Cortex | -6 | -82 | 26 | RH_DorsAttn_FEF_1; Right PreMotor & Supplementary Motor Area | 28 | -3 | 59 | 2.62 | 0.010 |
| LH_Vis_4; Left Secondary Visual Cortex | -27 | -95 | -4 | LH_DorsAttn_Post_3; Left Visual Motor Cortex | -24 | -68 | 49 | 2.62 | 0.010 |
| RH_SomMot_7; Right Primary Sensory Cortex | 30 | -37 | 64 | RH_Cont_PFCl_2; Right Dorsolateral Prefrontal Cortex | 45 | 39 | 15 | 2.62 | 0.010 |
| LH_SomMot_6; Left Primary Motor Cortex | -6 | -29 | 70 | RH_Default_PFCdPFCm_3; Right Frontal Eye Fields | 26 | 24 | 50 | 2.62 | 0.010 |
| LH_DorsAttn_FEF_1; Left PreMotor & Supplementary Motor Area | -26 | -3 | 59 | LH_Cont_Cing_1; Left Ventral Posterior Cingulate Cortex | -4 | -26 | 33 | 2.61 | 0.010 |
| LH_Vis_6; Left Primary Visual Cortex | -12 | -67 | 7 | RH_SomMot_6; Right Primary Motor Cortex | 41 | -22 | 60 | 2.61 | 0.010 |
| LH_DorsAttn_Post_5; Left Visual Motor Cortex | -6 | -60 | 56 | RH_DorsAttn_FEF_1; Right PreMotor & Supplementary Motor Area | 28 | -3 | 59 | 2.60 | 0.010 |
| LH_DorsAttn_Post_5; Left Visual Motor Cortex | -6 | -60 | 56 | LH_Cont_pCun_1; Left Visual Motor Cortex | -9 | -73 | 38 | 2.60 | 0.010 |
| RH_SalVentAttn_Med_1; Right Dorsal Posterior Cingulate Cortex | 11 | -31 | 45 | LH_Default_Par_1; Left Angular Gyrus | -57 | -50 | 12 | 2.60 | 0.010 |
| LH_DorsAttn_Post_6; Left Visual Motor Cortex | -22 | -51 | 66 | RH_Cont_PFCl_4; Right Frontal Eye Fields | 43 | 16 | 45 | 2.59 | 0.010 |

**Eyes-Closed Alpha**

| ROI1 Labels (Schaefer; Talairach) | X | Y | Z | ROI2 Labels (Schaefer; Talairach) | X | Y | Z | t | p (unc) |
|---|---|---|---|---|---|---|---|---|---|
| RH_SomMot_3; Right Primary Motor Cortex | 57 | -4 | 11 | RH_Cont_PFCmp_1; Right Frontal Eye Fields | 6 | 28 | 30 | -3.60 | 0.0004 |
| RH_DorsAttn_Post_2; Right Primary Sensory Cortex | 50 | -24 | 42 | RH_Cont_Par_2; Right Angular Gyrus | 45 | -63 | 46 | -3.32 | 0.001 |
| RH_SomMot_1; Right Primary Auditory Cortex | 53 | -16 | 7 | RH_Cont_PFCmp_1; Right Frontal Eye Fields | 6 | 28 | 30 | -3.22 | 0.002 |
| LH_DorsAttn_Post_5; Left Visual Motor Cortex | -6 | -60 | 56 | LH_Default_PFC_2; Left Pars Orbitalis | -47 | 33 | -3 | -3.12 | 0.002 |
| RH_SomMot_5; Right PreMotor & Supplementary Motor Area | 47 | -11 | 48 | RH_DorsAttn_Post_4; Right Visual Motor Cortex | 27 | -67 | 51 | -3.10 | 0.002 |
| RH_SomMot_6; Right Primary Motor Cortex | 41 | -22 | 60 | RH_Cont_Par_2; Right Angular Gyrus | 45 | -63 | 46 | -3.05 | 0.003 |
| RH_Cont_Cing_1; Right Ventral Posterior Cingulate Cortex | 5 | -27 | 33 | LH_Default_Temp_2; Left Medial Temporal Gyrus | -58 | -32 | -1 | -3.00 | 0.003 |
| RH_DorsAttn_PrCv_1; Right Broca's Area/Operculum | 49 | 10 | 27 | RH_Cont_PFCmp_1; Right Frontal Eye Fields | 6 | 28 | 30 | -2.96 | 0.004 |
| RH_SomMot_2; Right Insula | 40 | -15 | 15 | RH_Cont_PFCmp_1; Right Frontal Eye Fields | 6 | 28 | 30 | -2.86 | 0.005 |
| RH_SomMot_1; Right Primary Auditory Cortex | 53 | -16 | 7 | RH_Default_PFCdPFCm_1; Right Anterior Prefrontal Cortex | 7 | 48 | 1 | -2.84 | 0.005 |
| LH_Limbic_TempPole_2; Left Inferior Temporal Gyrus | -57 | -33 | -21 | LH_Default_pCunPCC_2; Left Ventral Posterior Cingulate Cortex | -6 | -53 | 33 | -2.83 | 0.005 |
| RH_DorsAttn_Post_2; Right Primary Sensory Cortex | 50 | -24 | 42 | RH_Default_Par_1; Right Angular Gyrus | 55 | -51 | 31 | -2.81 | 0.006 |
| RH_SomMot_5; Right PreMotor & Supplementary Motor Area | 47 | -11 | 48 | RH_Default_Par_1; Right Angular Gyrus | 55 | -51 | 31 | -2.80 | 0.006 |
| RH_SalVentAttn_TempOccPar_2; Right Supramarginal Gyrus | 61 | -26 | 27 | RH_Cont_Par_2; Right Angular Gyrus | 45 | -63 | 46 | -2.77 | 0.006 |
| LH_Vis_9; Left Visual Association Cortex | -6 | -82 | 26 | RH_SalVentAttn_Med_1; Right Dorsal Posterior Cingulate Cortex | 11 | -31 | 45 | -2.76 | 0.006 |
| RH_SomMot_4; Right Primary Motor Cortex | 58 | -5 | 31 | RH_DorsAttn_Post_1; Right Visual Association Cortex | 50 | -62 | 16 | -2.74 | 0.007 |

| | | | | | | | | | |
|---|---|---|---|---|---|---|---|---|---|
| RH_DorsAttn_Post_1; Right Visual Association Cortex | 50 | -62 | 16 | LH_SalVentAttn_ParOper_1; Left Supramarginal Gyrus | -59 | -38 | 29 | -2.72 | 0.007 |
| RH_SomMot_4; Right Primary Motor Cortex | 58 | -5 | 31 | LH_Cont_Cing_1; Left Ventral Posterior Cingulate Cortex | -4 | -26 | 33 | -2.72 | 0.007 |
| RH_SomMot_4; Right Primary Motor Cortex | 58 | -5 | 31 | LH_SalVentAttn_Med_2; Left Dorsal Posterior Cingulate Cortex | -11 | -34 | 45 | -2.71 | 0.007 |
| RH_Cont_PFCl_1; Right Anterior Prefrontal Cortex | 30 | 58 | -3 | LH_Default_PFC_6; Left PreMotor & Supplementary Motor Area | -41 | 14 | 48 | 2.70 | 0.008 |
| LH_Vis_3; Left Visual Association Cortex | -17 | -60 | -7 | RH_SomMot_3; Right Primary Motor Cortex | 57 | -4 | 11 | -2.68 | 0.008 |
| RH_SomMot_6; Right Primary Motor Cortex | 41 | -22 | 60 | RH_Default_Par_1; Right Angular Gyrus | 55 | -51 | 31 | -2.67 | 0.008 |
| LH_Default_Temp_1; Left Medial Temporal Gyrus | -55 | -4 | -20 | LH_Default_PFC_7; Left Frontal Eye Fields | -25 | 20 | 51 | -2.66 | 0.009 |
| RH_SalVentAttn_TempOccPar_1; Right Superior Temporal Gyrus | 58 | -42 | 13 | RH_Default_PFCv_2; Right Broca's Area/ Pars Triangularis | 51 | 28 | 0 | -2.66 | 0.009 |
| RH_SomMot_4; Right Primary Motor Cortex | 58 | -5 | 31 | LH_SalVentAttn_Med_1; Left Dorsal Anterior Cingulate Cortex | -5 | 20 | 34 | -2.65 | 0.009 |
| RH_DorsAttn_PrCv_1; Right Broca's Area/Operculum | 49 | 10 | 27 | LH_SalVentAttn_Med_1; Left Dorsal Anterior Cingulate Cortex | -5 | 20 | 34 | -2.63 | 0.009 |
| LH_Default_pCunPCC_2; Left Ventral Posterior Cingulate Cortex | -6 | -53 | 33 | RH_Default_Par_1; Right Angular Gyrus | 55 | -51 | 31 | -2.62 | 0.010 |
| RH_SomMot_1; Right Primary Auditory Cortex | 53 | -16 | 7 | RH_Default_PFCv_2; Right Broca's Area/ Pars Triangularis | 51 | 28 | 0 | -2.62 | 0.010 |
| LH_SalVentAttn_Med_1; Left Dorsal Anterior Cingulate Cortex | -5 | 20 | 34 | RH_SalVentAttn_TempOccPar_2; Right Supramarginal Gyrus | 61 | -26 | 27 | -2.62 | 0.010 |
| RH_DorsAttn_Post_2; Right Primary Sensory Cortex | 50 | -24 | 42 | RH_SalVentAttn_TempOccPar_2; Right Supramarginal Gyrus | 61 | -26 | 27 | -2.60 | 0.010 |
| RH_Cont_Par_2; Right Angular Gyrus | 45 | -63 | 46 | LH_Default_PFC_2; Left Pars Orbitalis | -47 | 33 | -3 | -2.60 | 0.010 |
| LH_DorsAttn_Post_1; Left Fusiform Cortex | -47 | -58 | -13 | RH_Default_PFCv_2; Right Broca's Area/ Pars Triangularis | 51 | 28 | 0 | -2.59 | 0.010 |

**Eyes-Closed Beta**

| ROI1 Labels (Schaefer; Talairach) | X | Y | Z | ROI2 Labels (Schaefer; Talairach) | X | Y | Z | t | p (unc) |
|---|---|---|---|---|---|---|---|---|---|
| LH_Vis_7; Left Visual Association Cortex | -47 | -71 | 11 | LH_Default_pCunPCC_1; Left Ventral Posterior Cingulate Cortex | -11 | -56 | 13 | 4.38 | 2.13E-05 |
| LH_Vis_7; Left Visual Association Cortex | -47 | -71 | 11 | RH_Default_pCunPCC_1; Right Ventral Posterior Cingulate Cortex | 12 | -54 | 14 | 4.24 | 3.75E-05 |
| LH_Vis_7; Left Visual Association Cortex | -47 | -71 | 11 | LH_SalVentAttn_Med_3; Left PreMotor & Supplementary Motor Area | -6 | 4 | 62 | 4.23 | 3.99E-05 |
| LH_DorsAttn_Post_3; Left Visual Motor Cortex | -24 | -68 | 49 | RH_DorsAttn_Post_1; Right Visual Association Cortex | 50 | -62 | 16 | 3.95 | 0.0001 |
| LH_Vis_6; Left Primary Visual Cortex | -12 | -67 | 7 | LH_Vis_7; Left Visual Association Cortex | -47 | -71 | 11 | 3.86 | 0.0002 |
| LH_DorsAttn_Post_6; Left Visual Motor Cortex | -22 | -51 | 66 | LH_Default_pCunPCC_2; Left Ventral Posterior Cingulate Cortex | -6 | -53 | 33 | 3.82 | 0.0002 |
| LH_Vis_9; Left Visual Association Cortex | -6 | -82 | 26 | RH_DorsAttn_Post_4; Right Visual Motor Cortex | 27 | -67 | 51 | 3.80 | 0.0002 |
| LH_Vis_7; Left Visual Association Cortex | -47 | -71 | 11 | RH_Vis_5; Right Primary Visual Cortex | 8 | -76 | 5 | 3.77 | 0.0002 |
| LH_Vis_7; Left Visual Association Cortex | -47 | -71 | 11 | RH_Vis_6; Right Secondary Visual Cortex | 17 | -57 | 5 | 3.77 | 0.0002 |
| LH_Vis_7; Left Visual Association Cortex | -47 | -71 | 11 | LH_Default_pCunPCC_2; Left Ventral Posterior Cingulate Cortex | -6 | -53 | 33 | 3.69 | 0.0003 |
| LH_Vis_7; Left Visual Association Cortex | -47 | -71 | 11 | RH_SalVentAttn_TempOccPar_1; Right Superior Temporal Gyrus | 58 | -42 | 13 | 3.67 | 0.0003 |
| LH_Vis_7; Left Visual Association Cortex | -47 | -71 | 11 | RH_DorsAttn_FEF_1; Right PreMotor & Supplementary Motor Area | 28 | -3 | 59 | 3.67 | 0.0003 |
| RH_Vis_3; Right Fusiform Cortex | 49 | -60 | -11 | RH_SomMot_7; Right Primary Sensory Cortex | 30 | -37 | 64 | 3.66 | 0.0003 |

| | | | | | | | | | |
|---|---|---|---|---|---|---|---|---|---|
| RH_DorsAttn_Post_4; Right Visual Motor Cortex | 27 | -67 | 51 | RH_Default_pCunPCC_2; Right Ventral Posterior Cingulate Cortex | 7 | -52 | 31 | 3.66 | 0.0003 |
| RH_DorsAttn_Post_1; Right Visual Association Cortex | 50 | -62 | 16 | LH_Cont_pCun_1; Left Visual Motor Cortex | -9 | -73 | 38 | 3.64 | 0.0004 |
| RH_SomMot_5; Right PreMotor & Supplementary Motor Area | 47 | -11 | 48 | LH_DorsAttn_Post_3; Left Visual Motor Cortex | -24 | -68 | 49 | 3.63 | 0.0004 |
| RH_Vis_3; Right Fusiform Cortex | 49 | -60 | -11 | LH_DorsAttn_Post_3; Left Visual Motor Cortex | -24 | -68 | 49 | 3.62 | 0.0004 |
| LH_Vis_9; Left Visual Association Cortex | -6 | -82 | 26 | RH_DorsAttn_Post_1; Right Visual Association Cortex | 50 | -62 | 16 | 3.52 | 0.0006 |
| LH_Vis_5; Left Secondary Visual Cortex | -5 | -92 | -2 | RH_DorsAttn_Post_4; Right Visual Motor Cortex | 27 | -67 | 51 | 3.52 | 0.0006 |
| LH_SomMot_5; Left Primary Motor Cortex | -39 | -23 | 59 | RH_SalVentAttn_TempOccPar_1; Right Superior Temporal Gyrus | 58 | -42 | 13 | 3.51 | 0.0006 |
| LH_Vis_9; Left Visual Association Cortex | -6 | -82 | 26 | RH_Cont_Par_2; Right Angular Gyrus | 45 | -63 | 46 | 3.49 | 0.0006 |
| LH_Vis_7; Left Visual Association Cortex | -47 | -71 | 11 | RH_DorsAttn_Post_2; Right Primary Sensory Cortex | 50 | -24 | 42 | 3.49 | 0.0006 |
| LH_SomMot_5; Left Primary Motor Cortex | -39 | -23 | 59 | RH_Default_Par_1; Right Angular Gyrus | 55 | -51 | 31 | 3.48 | 0.0007 |
| LH_DorsAttn_Post_3; Left Visual Motor Cortex | -24 | -68 | 49 | RH_SalVentAttn_TempOccPar_1; Right Superior Temporal Gyrus | 58 | -42 | 13 | 3.47 | 0.0007 |
| RH_Vis_3; Right Fusiform Cortex | 49 | -60 | -11 | RH_DorsAttn_Post_3; Right Visual Motor Cortex | 38 | -45 | 49 | 3.45 | 0.0007 |
| LH_Vis_5; Left Secondary Visual Cortex | -5 | -92 | -2 | RH_DorsAttn_Post_3; Right Visual Motor Cortex | 38 | -45 | 49 | 3.45 | 0.0007 |
| LH_Vis_6; Left Primary Visual Cortex | -12 | -67 | 7 | RH_DorsAttn_Post_4; Right Visual Motor Cortex | 27 | -67 | 51 | 3.45 | 0.0007 |
| LH_Vis_3; Left Visual Association Cortex | -17 | -60 | -7 | RH_DorsAttn_PrCv_1; Right Broca's Area/Operculum | 49 | 10 | 27 | 3.44 | 0.0007 |
| LH_Vis_7; Left Visual Association Cortex | -47 | -71 | 11 | RH_Vis_7; Right Visual Association Cortex | 36 | -82 | 16 | 3.44 | 0.0008 |
| RH_DorsAttn_Post_4; Right Visual Motor Cortex | 27 | -67 | 51 | LH_Default_pCunPCC_2; Left Ventral Posterior Cingulate Cortex | -6 | -53 | 33 | 3.41 | 0.0008 |
| LH_Vis_7; Left Visual Association Cortex | -47 | -71 | 11 | LH_SomMot_2; Left Primary Sensory Cortex | -37 | -21 | 16 | 3.38 | 0.0009 |
| LH_Vis_7; Left Visual Association Cortex | -47 | -71 | 11 | RH_SomMot_6; Right Primary Motor Cortex | 41 | -22 | 60 | 3.38 | 0.0009 |
| RH_Vis_3; Right Fusiform Cortex | 49 | -60 | -11 | RH_Cont_Par_1; Right Supramarginal Gyrus | 57 | -39 | 44 | 3.36 | 0.0010 |
| LH_Vis_2; Left Visual Association Cortex | -26 | -77 | -14 | RH_SalVentAttn_TempOccPar_1; Right Superior Temporal Gyrus | 58 | -42 | 13 | 3.35 | 0.001 |
| LH_DorsAttn_Post_3; Left Visual Motor Cortex | -24 | -68 | 49 | RH_Default_pCunPCC_1; Right Ventral Posterior Cingulate Cortex | 12 | -54 | 14 | 3.35 | 0.001 |
| LH_Vis_7; Left Visual Association Cortex | -47 | -71 | 11 | RH_Vis_4; Right Secondary Visual Cortex | 22 | -96 | -5 | 3.32 | 0.001 |
| LH_Vis_7; Left Visual Association Cortex | -47 | -71 | 11 | RH_SomMot_5; Right PreMotor & Supplementary Motor Area | 47 | -11 | 48 | 3.32 | 0.001 |
| RH_SalVentAttn_Med_1; Right Dorsal Posterior Cingulate Cortex | 11 | -31 | 45 | RH_Cont_Cing_1; Right Ventral Posterior Cingulate Cortex | 5 | -27 | 33 | 3.31 | 0.001 |
| LH_DorsAttn_Post_6; Left Visual Motor Cortex | -22 | -51 | 66 | RH_Default_pCunPCC_2; Right Ventral Posterior Cingulate Cortex | 7 | -52 | 31 | 3.30 | 0.001 |
| LH_SomMot_2; Left Primary Sensory Cortex | -37 | -21 | 16 | LH_Default_Par_2; Left Angular Gyrus | -48 | -63 | 35 | 3.30 | 0.001 |
| LH_Cont_pCun_1; Left Visual Motor Cortex | -9 | -73 | 38 | RH_Cont_pCun_1; Right Visual Motor Cortex | 9 | -66 | 43 | 3.29 | 0.001 |
| RH_Vis_7; Right Visual Association Cortex | 36 | -82 | 16 | RH_DorsAttn_Post_5; Right Visual Motor Cortex | 14 | -52 | 66 | 3.26 | 0.001 |
| LH_Vis_7; Left Visual Association Cortex | -47 | -71 | 11 | LH_DorsAttn_Post_3; Left Visual Motor Cortex | -24 | -68 | 49 | 3.25 | 0.001 |
| LH_Vis_5; Left Secondary Visual Cortex | -5 | -92 | -2 | RH_Vis_6; Right Secondary Visual Cortex | 17 | -57 | 5 | 3.25 | 0.001 |
| LH_SomMot_5; Left Primary Motor Cortex | -39 | -23 | 59 | RH_Cont_Par_1; Right Supramarginal Gyrus | 57 | -39 | 44 | 3.25 | 0.001 |

| | | | | | | | | | |
|---|---|---|---|---|---|---|---|---|---|
| LH_Vis_7; Left Visual Association Cortex | -47 | -71 | 11 | RH_SomMot_4; Right Primary Motor Cortex | 58 | -5 | 31 | 3.24 | 0.001 |
| LH_DorsAttn_Post_3; Left Visual Motor Cortex | -24 | -68 | 49 | LH_Default_pCunPCC_1; Left Ventral Posterior Cingulate Cortex | -11 | -56 | 13 | 3.24 | 0.001 |
| LH_Vis_6; Left Primary Visual Cortex | -12 | -67 | 7 | LH_DorsAttn_Post_6; Left Visual Motor Cortex | -22 | -51 | 66 | 3.24 | 0.001 |
| LH_Vis_7; Left Visual Association Cortex | -47 | -71 | 11 | RH_SalVentAttn_Med_2; Right PreMotor & Supplementary Motor Area | 7 | 6 | 52 | 3.23 | 0.001 |
| RH_DorsAttn_FEF_1; Right PreMotor & Supplementary Motor Area | 28 | -3 | 59 | LH_Default_PFC_5; Left Dorsolateral Prefrontal Cortex | -9 | 48 | 41 | 3.23 | 0.002 |
| RH_Vis_5; Right Primary Visual Cortex | 8 | -76 | 5 | LH_DorsAttn_Post_3; Left Visual Motor Cortex | -24 | -68 | 49 | 3.22 | 0.002 |
| LH_Vis_7; Left Visual Association Cortex | -47 | -71 | 11 | RH_Default_pCunPCC_2; Right Ventral Posterior Cingulate Cortex | 7 | -52 | 31 | 3.21 | 0.002 |
| RH_DorsAttn_PrCv_1; Right Broca's Area/Operculum | 49 | 10 | 27 | LH_Default_pCunPCC_1; Left Ventral Posterior Cingulate Cortex | -11 | -56 | 13 | 3.21 | 0.002 |
| LH_Vis_2; Left Visual Association Cortex | -26 | -77 | -14 | RH_SomMot_4; Right Primary Motor Cortex | 58 | -5 | 31 | 3.21 | 0.002 |
| LH_DorsAttn_Post_3; Left Visual Motor Cortex | -24 | -68 | 49 | RH_Default_pCunPCC_2; Right Ventral Posterior Cingulate Cortex | 7 | -52 | 31 | 3.20 | 0.002 |
| RH_Vis_5; Right Primary Visual Cortex | 8 | -76 | 5 | RH_SalVentAttn_TempOccPar_2; Right Supramarginal Gyrus | 61 | -26 | 27 | 3.20 | 0.002 |
| RH_DorsAttn_Post_4; Right Visual Motor Cortex | 27 | -67 | 51 | LH_Cont_pCun_1; Left Visual Motor Cortex | -9 | -73 | 38 | 3.19 | 0.002 |
| RH_SomMot_7; Right Primary Sensory Cortex | 30 | -37 | 64 | LH_DorsAttn_Post_1; Left Fusiform Cortex | -47 | -58 | -13 | 3.18 | 0.002 |
| LH_Vis_6; Left Primary Visual Cortex | -12 | -67 | 7 | RH_Vis_5; Right Primary Visual Cortex | 8 | -76 | 5 | 3.18 | 0.002 |
| LH_Cont_Par_1; Left Angular Gyrus | -37 | -53 | 46 | LH_Default_pCunPCC_1; Left Ventral Posterior Cingulate Cortex | -11 | -56 | 13 | 3.16 | 0.002 |
| RH_SomMot_5; Right PreMotor & Supplementary Motor Area | 47 | -11 | 48 | LH_SalVentAttn_Med_2; Left Dorsal Posterior Cingulate Cortex | -11 | -34 | 45 | 3.16 | 0.002 |
| LH_Vis_7; Left Visual Association Cortex | -47 | -71 | 11 | RH_DorsAttn_Post_1; Right Visual Association Cortex | 50 | -62 | 16 | 3.15 | 0.002 |
| RH_DorsAttn_Post_4; Right Visual Motor Cortex | 27 | -67 | 51 | LH_Default_pCunPCC_1; Left Ventral Posterior Cingulate Cortex | -11 | -56 | 13 | 3.15 | 0.002 |
| LH_Vis_6; Left Primary Visual Cortex | -12 | -67 | 7 | RH_Cont_Par_2; Right Angular Gyrus | 45 | -63 | 46 | 3.14 | 0.002 |
| LH_Vis_6; Left Primary Visual Cortex | -12 | -67 | 7 | LH_DorsAttn_Post_3; Left Visual Motor Cortex | -24 | -68 | 49 | 3.14 | 0.002 |
| LH_Vis_6; Left Primary Visual Cortex | -12 | -67 | 7 | RH_DorsAttn_PrCv_1; Right Broca's Area/Operculum | 49 | 10 | 27 | 3.13 | 0.002 |
| RH_Vis_5; Right Primary Visual Cortex | 8 | -76 | 5 | RH_DorsAttn_Post_3; Right Visual Motor Cortex | 38 | -45 | 49 | 3.13 | 0.002 |
| LH_Vis_2; Left Visual Association Cortex | -26 | -77 | -14 | RH_SalVentAttn_TempOccPar_2; Right Supramarginal Gyrus | 61 | -26 | 27 | 3.12 | 0.002 |
| LH_Vis_7; Left Visual Association Cortex | -47 | -71 | 11 | LH_Cont_pCun_1; Left Visual Motor Cortex | -9 | -73 | 38 | 3.12 | 0.002 |
| LH_DorsAttn_Post_3; Left Visual Motor Cortex | -24 | -68 | 49 | RH_Default_PFCdPFCm_3; Right Frontal Eye Fields | 26 | 24 | 50 | 3.11 | 0.002 |
| LH_Vis_8; Left Visual Association Cortex | -25 | -88 | 20 | LH_DorsAttn_Post_5; Left Visual Motor Cortex | -6 | -60 | 56 | 3.11 | 0.002 |
| LH_SomMot_6; Left Primary Motor Cortex | -6 | -29 | 70 | LH_DorsAttn_Post_5; Left Visual Motor Cortex | -6 | -60 | 56 | 3.10 | 0.002 |
| LH_SomMot_5; Left Primary Motor Cortex | -39 | -23 | 59 | RH_SomMot_5; Right PreMotor & Supplementary Motor Area | 47 | -11 | 48 | 3.09 | 0.002 |
| LH_SomMot_5; Left Primary Motor Cortex | -39 | -23 | 59 | RH_SomMot_2; Right Insula | 40 | -15 | 15 | 3.08 | 0.002 |
| LH_Vis_7; Left Visual Association Cortex | -47 | -71 | 11 | LH_Default_PFC_7; Left Frontal Eye Fields | -25 | 20 | 51 | 3.08 | 0.002 |
| LH_Cont_PFCl_1; Left Dorsolateral Prefrontal Cortex | -43 | 33 | 21 | RH_Cont_Cing_1; Right Ventral Posterior Cingulate Cortex | 5 | -27 | 33 | 3.07 | 0.003 |

| | | | | | | | | | |
|---|---|---|---|---|---|---|---|---|---|
| LH_Vis_7; Left Visual Association Cortex | -47 | -71 | 11 | LH_Default_Par_2; Left Angular Gyrus | -48 | -63 | 35 | 3.07 | 0.003 |
| LH_Vis_8; Left Visual Association Cortex | -25 | -88 | 20 | RH_Cont_pCun_1; Right Visual Motor Cortex | 9 | -66 | 43 | 3.06 | 0.003 |
| LH_Default_Par_1; Left Angular Gyrus | -57 | -50 | 12 | RH_Default_Temp_1; Right Medial Temporal Gyrus | 62 | -23 | -19 | 3.06 | 0.003 |
| RH_Vis_5; Right Primary Visual Cortex | 8 | -76 | 5 | LH_DorsAttn_Post_6; Left Visual Motor Cortex | -22 | -51 | 66 | 3.06 | 0.003 |
| LH_DorsAttn_Post_3; Left Visual Motor Cortex | -24 | -68 | 49 | RH_Cont_Par_2; Right Angular Gyrus | 45 | -63 | 46 | 3.06 | 0.003 |
| LH_Vis_7; Left Visual Association Cortex | -47 | -71 | 11 | RH_SomMot_8; Right Primary Motor Cortex | 6 | -26 | 70 | 3.05 | 0.003 |
| LH_DorsAttn_Post_3; Left Visual Motor Cortex | -24 | -68 | 49 | LH_Default_pCunPCC_2; Left Ventral Posterior Cingulate Cortex | -6 | -53 | 33 | 3.05 | 0.003 |
| RH_Cont_Par_2; Right Angular Gyrus | 45 | -63 | 46 | RH_Default_PFCdPFCm_3; Right Frontal Eye Fields | 26 | 24 | 50 | 3.05 | 0.003 |
| RH_Vis_4; Right Secondary Visual Cortex | 22 | -96 | -5 | RH_SomMot_7; Right Primary Sensory Cortex | 30 | -37 | 64 | 3.04 | 0.003 |
| LH_SomMot_6; Left Primary Motor Cortex | -6 | -29 | 70 | LH_Default_PFC_2; Left Pars Orbitalis | -47 | 33 | -3 | 3.04 | 0.003 |
| LH_Vis_7; Left Visual Association Cortex | -47 | -71 | 11 | LH_Vis_9; Left Visual Association Cortex | -6 | -82 | 26 | 3.04 | 0.003 |
| RH_Vis_2; Right Fusiform Cortex | 27 | -66 | -12 | LH_DorsAttn_Post_6; Left Visual Motor Cortex | -22 | -51 | 66 | 3.03 | 0.003 |
| LH_Vis_7; Left Visual Association Cortex | -47 | -71 | 11 | LH_DorsAttn_FEF_1; Left PreMotor & Supplementary Motor Area | -26 | -3 | 59 | 3.03 | 0.003 |
| LH_Vis_7; Left Visual Association Cortex | -47 | -71 | 11 | LH_SalVentAttn_ParOper_1; Left Supramarginal Gyrus | -59 | -38 | 29 | 3.03 | 0.003 |
| LH_Vis_7; Left Visual Association Cortex | -47 | -71 | 11 | LH_SomMot_5; Left Primary Motor Cortex | -39 | -23 | 59 | 3.02 | 0.003 |
| LH_Default_pCunPCC_2; Left Ventral Posterior Cingulate Cortex | -6 | -53 | 33 | RH_Default_pCunPCC_2; Right Ventral Posterior Cingulate Cortex | 7 | -52 | 31 | 3.02 | 0.003 |
| LH_Vis_5; Left Secondary Visual Cortex | -5 | -92 | -2 | RH_Cont_Par_2; Right Angular Gyrus | 45 | -63 | 46 | 3.01 | 0.003 |
| LH_SomMot_2; Left Primary Sensory Cortex | -37 | -21 | 16 | LH_Cont_Par_1; Left Angular Gyrus | -37 | -53 | 46 | 3.00 | 0.003 |
| RH_SomMot_1; Right Primary Auditory Cortex | 53 | -16 | 7 | RH_SomMot_3; Right Primary Motor Cortex | 57 | -4 | 11 | 3.00 | 0.003 |
| LH_SomMot_5; Left Primary Motor Cortex | -39 | -23 | 59 | RH_Cont_Par_2; Right Angular Gyrus | 45 | -63 | 46 | 3.00 | 0.003 |
| LH_Vis_3; Left Visual Association Cortex | -17 | -60 | -7 | RH_SalVentAttn_TempOccPar_1; Right Superior Temporal Gyrus | 58 | -42 | 13 | 3.00 | 0.003 |
| RH_Vis_5; Right Primary Visual Cortex | 8 | -76 | 5 | RH_Default_Par_1; Right Angular Gyrus | 55 | -51 | 31 | 3.00 | 0.003 |
| LH_Default_pCunPCC_2; Left Ventral Posterior Cingulate Cortex | -6 | -53 | 33 | RH_Default_Par_1; Right Angular Gyrus | 55 | -51 | 31 | 2.99 | 0.003 |
| LH_Vis_5; Left Secondary Visual Cortex | -5 | -92 | -2 | LH_Vis_7; Left Visual Association Cortex | -47 | -71 | 11 | 2.99 | 0.003 |
| LH_Vis_2; Left Visual Association Cortex | -26 | -77 | -14 | RH_SomMot_3; Right Primary Motor Cortex | 57 | -4 | 11 | 2.99 | 0.003 |
| LH_Vis_2; Left Visual Association Cortex | -26 | -77 | -14 | RH_Default_PFCv_2; Right Broca's Area/ Pars Triangularis | 51 | 28 | 0 | 2.98 | 0.003 |
| LH_DorsAttn_Post_6; Left Visual Motor Cortex | -22 | -51 | 66 | LH_Default_pCunPCC_1; Left Ventral Posterior Cingulate Cortex | -11 | -56 | 13 | 2.98 | 0.003 |
| LH_Vis_7; Left Visual Association Cortex | -47 | -71 | 11 | LH_SomMot_6; Left Primary Motor Cortex | -6 | -29 | 70 | 2.97 | 0.003 |
| LH_DorsAttn_Post_6; Left Visual Motor Cortex | -22 | -51 | 66 | RH_Cont_pCun_1; Right Visual Motor Cortex | 9 | -66 | 43 | 2.97 | 0.003 |
| RH_SomMot_5; Right PreMotor & Supplementary Motor Area | 47 | -11 | 48 | LH_Default_pCunPCC_2; Left Ventral Posterior Cingulate Cortex | -6 | -53 | 33 | 2.97 | 0.003 |
| LH_Vis_3; Left Visual Association Cortex | -17 | -60 | -7 | RH_Default_Par_1; Right Angular Gyrus | 55 | -51 | 31 | 2.97 | 0.003 |
| RH_SomMot_8; Right Primary Motor Cortex | 6 | -26 | 70 | RH_Cont_pCun_1; Right Visual Motor Cortex | 9 | -66 | 43 | 2.96 | 0.004 |

| | | | | | | | | | |
|---|---|---|---|---|---|---|---|---|---|
| LH_SomMot_1; Left Supramarginal Gyrus | -53 | -23 | 8 | RH_DorsAttn_PrCv_1; Right Broca's Area/Operculum | 49 | 10 | 27 | 2.96 | 0.004 |
| RH_Vis_5; Right Primary Visual Cortex | 8 | -76 | 5 | RH_DorsAttn_Post_2; Right Primary Sensory Cortex | 50 | -24 | 42 | 2.95 | 0.004 |
| RH_Vis_6; Right Secondary Visual Cortex | 17 | -57 | 5 | LH_DorsAttn_Post_3; Left Visual Motor Cortex | -24 | -68 | 49 | 2.95 | 0.004 |
| LH_Vis_7; Left Visual Association Cortex | -47 | -71 | 11 | LH_DorsAttn_Post_2; Left Supramarginal Gyrus | -57 | -25 | 39 | 2.95 | 0.004 |
| LH_DorsAttn_Post_3; Left Visual Motor Cortex | -24 | -68 | 49 | RH_Default_Par_1; Right Angular Gyrus | 55 | -51 | 31 | 2.95 | 0.004 |
| RH_Default_Par_1; Right Angular Gyrus | 55 | -51 | 31 | RH_Default_pCunPCC_1; Right Ventral Posterior Cingulate Cortex | 12 | -54 | 14 | 2.95 | 0.004 |
| LH_Vis_3; Left Visual Association Cortex | -17 | -60 | -7 | LH_Default_PFC_7; Left Frontal Eye Fields | -25 | 20 | 51 | 2.94 | 0.004 |
| RH_Vis_5; Right Primary Visual Cortex | 8 | -76 | 5 | LH_Default_pCunPCC_2; Left Ventral Posterior Cingulate Cortex | -6 | -53 | 33 | 2.93 | 0.004 |
| RH_Vis_4; Right Secondary Visual Cortex | 22 | -96 | -5 | LH_DorsAttn_Post_6; Left Visual Motor Cortex | -22 | -51 | 66 | 2.92 | 0.004 |
| LH_Vis_5; Left Secondary Visual Cortex | -5 | -92 | -2 | RH_SomMot_7; Right Primary Sensory Cortex | 30 | -37 | 64 | 2.92 | 0.004 |
| RH_Vis_4; Right Secondary Visual Cortex | 22 | -96 | -5 | RH_DorsAttn_Post_3; Right Visual Motor Cortex | 38 | -45 | 49 | 2.92 | 0.004 |
| LH_Vis_9; Left Visual Association Cortex | -6 | -82 | 26 | RH_DorsAttn_Post_3; Right Visual Motor Cortex | 38 | -45 | 49 | 2.92 | 0.004 |
| LH_Cont_PFCl_1; Left Dorsolateral Prefrontal Cortex | -43 | 33 | 21 | RH_Default_PFCdPFCm_2; Right Dorsolateral Prefrontal Cortex | 11 | 50 | 39 | 2.92 | 0.004 |
| RH_DorsAttn_Post_3; Right Visual Motor Cortex | 38 | -45 | 49 | RH_DorsAttn_Post_5; Right Visual Motor Cortex | 14 | -52 | 66 | 2.92 | 0.004 |
| RH_SomMot_5; Right PreMotor & Supplementary Motor Area | 47 | -11 | 48 | LH_Cont_Par_1; Left Angular Gyrus | -37 | -53 | 46 | 2.91 | 0.004 |
| LH_SomMot_6; Left Primary Motor Cortex | -6 | -29 | 70 | RH_DorsAttn_FEF_1; Right PreMotor & Supplementary Motor Area | 28 | -3 | 59 | 2.91 | 0.004 |
| LH_Vis_7; Left Visual Association Cortex | -47 | -71 | 11 | LH_DorsAttn_Post_5; Left Visual Motor Cortex | -6 | -60 | 56 | 2.90 | 0.004 |
| RH_Vis_3; Right Fusiform Cortex | 49 | -60 | -11 | LH_SalVentAttn_Med_2; Left Dorsal Posterior Cingulate Cortex | -11 | -34 | 45 | 2.90 | 0.004 |
| RH_Vis_6; Right Secondary Visual Cortex | 17 | -57 | 5 | LH_DorsAttn_Post_6; Left Visual Motor Cortex | -22 | -51 | 66 | 2.90 | 0.004 |
| RH_Cont_Par_1; Right Supramarginal Gyrus | 57 | -39 | 44 | RH_Default_pCunPCC_1; Right Ventral Posterior Cingulate Cortex | 12 | -54 | 14 | 2.89 | 0.004 |
| LH_Cont_PFCl_1; Left Dorsolateral Prefrontal Cortex | -43 | 33 | 21 | LH_Cont_Cing_1; Left Ventral Posterior Cingulate Cortex | -4 | -26 | 33 | 2.89 | 0.004 |
| LH_Default_Temp_2; Left Medial Temporal Gyrus | -58 | -32 | -1 | LH_Default_Par_2; Left Angular Gyrus | -48 | -63 | 35 | 2.89 | 0.004 |
| LH_Vis_6; Left Primary Visual Cortex | -12 | -67 | 7 | LH_Cont_Par_1; Left Angular Gyrus | -37 | -53 | 46 | 2.89 | 0.004 |
| RH_DorsAttn_Post_5; Right Visual Motor Cortex | 14 | -52 | 66 | RH_Default_pCunPCC_2; Right Ventral Posterior Cingulate Cortex | 7 | -52 | 31 | 2.89 | 0.004 |
| RH_SomMot_6; Right Primary Motor Cortex | 41 | -22 | 60 | LH_Default_pCunPCC_2; Left Ventral Posterior Cingulate Cortex | -6 | -53 | 33 | 2.88 | 0.005 |
| RH_Vis_5; Right Primary Visual Cortex | 8 | -76 | 5 | LH_Cont_Par_1; Left Angular Gyrus | -37 | -53 | 46 | 2.88 | 0.005 |
| LH_Vis_8; Left Visual Association Cortex | -25 | -88 | 20 | RH_Limbic_OFC_1; Right Orbitofrontal Cortex | 12 | 35 | -20 | 2.88 | 0.005 |
| LH_SomMot_6; Left Primary Motor Cortex | -6 | -29 | 70 | LH_Default_PFC_5; Left Dorsolateral Prefrontal Cortex | -9 | 48 | 41 | 2.86 | 0.005 |
| LH_Vis_3; Left Visual Association Cortex | -17 | -60 | -7 | LH_Default_PFC_4; Left Anterior Prefrontal Cortex | -24 | 61 | -1 | 2.86 | 0.005 |
| LH_Vis_9; Left Visual Association Cortex | -6 | -82 | 26 | RH_Cont_PFCl_1; Right Anterior Prefrontal Cortex | 30 | 58 | -3 | 2.86 | 0.005 |
| RH_Vis_5; Right Primary Visual Cortex | 8 | -76 | 5 | RH_Cont_Par_2; Right Angular Gyrus | 45 | -63 | 46 | 2.86 | 0.005 |
| LH_Cont_pCun_1; Left Visual Motor Cortex | -9 | -73 | 38 | RH_Cont_Par_2; Right Angular Gyrus | 45 | -63 | 46 | 2.86 | 0.005 |

| | | | | | | | | | |
|---|---|---|---|---|---|---|---|---|---|
| LH_DorsAttn_Post_5; Left Visual Motor Cortex | -6 | -60 | 56 | LH_DorsAttn_Post_6; Left Visual Motor Cortex | -22 | -51 | 66 | 2.86 | 0.005 |
| RH_Vis_6; Right Secondary Visual Cortex | 17 | -57 | 5 | RH_DorsAttn_PrCv_1; Right Broca's Area/Operculum | 49 | 10 | 27 | 2.86 | 0.005 |
| LH_DorsAttn_Post_3; Left Visual Motor Cortex | -24 | -68 | 49 | RH_SalVentAttn_TempOccPar_2; Right Supramarginal Gyrus | 61 | -26 | 27 | 2.86 | 0.005 |
| LH_SomMot_6; Left Primary Motor Cortex | -6 | -29 | 70 | RH_Default_PFCdPFCm_2; Right Dorsolateral Prefrontal Cortex | 11 | 50 | 39 | 2.86 | 0.005 |
| LH_Vis_2; Left Visual Association Cortex | -26 | -77 | -14 | LH_Limbic_TempPole_1; Left Temporal Pole | -32 | 2 | -37 | 2.84 | 0.005 |
| LH_SomMot_5; Left Primary Motor Cortex | -39 | -23 | 59 | RH_Default_Temp_3; Right Superior Temporal Gyrus | 57 | -26 | -2 | 2.84 | 0.005 |
| LH_DorsAttn_Post_3; Left Visual Motor Cortex | -24 | -68 | 49 | LH_DorsAttn_Post_6; Left Visual Motor Cortex | -22 | -51 | 66 | 2.83 | 0.005 |
| LH_Vis_7; Left Visual Association Cortex | -47 | -71 | 11 | RH_Default_Par_1; Right Angular Gyrus | 55 | -51 | 31 | 2.82 | 0.005 |
| RH_Vis_6; Right Secondary Visual Cortex | 17 | -57 | 5 | RH_Cont_PFCl_2; Right Dorsolateral Prefrontal Cortex | 45 | 39 | 15 | 2.82 | 0.005 |
| LH_Vis_8; Left Visual Association Cortex | -25 | -88 | 20 | LH_SalVentAttn_FrOperIns_2; Left Insula | -38 | 12 | 6 | 2.82 | 0.005 |
| LH_Vis_6; Left Primary Visual Cortex | -12 | -67 | 7 | LH_Default_Par_2; Left Angular Gyrus | -48 | -63 | 35 | 2.81 | 0.006 |
| LH_SomMot_5; Left Primary Motor Cortex | -39 | -23 | 59 | RH_SomMot_1; Right Primary Auditory Cortex | 53 | -16 | 7 | 2.81 | 0.006 |
| RH_SomMot_5; Right PreMotor & Supplementary Motor Area | 47 | -11 | 48 | LH_Default_PFC_6; Left PreMotor & Supplementary Motor Area | -41 | 14 | 48 | 2.81 | 0.006 |
| RH_DorsAttn_Post_3; Right Visual Motor Cortex | 38 | -45 | 49 | RH_Default_pCunPCC_1; Right Ventral Posterior Cingulate Cortex | 12 | -54 | 14 | 2.80 | 0.006 |
| LH_Vis_7; Left Visual Association Cortex | -47 | -71 | 11 | RH_SalVentAttn_Med_1; Right Dorsal Posterior Cingulate Cortex | 11 | -31 | 45 | 2.80 | 0.006 |
| RH_Default_Par_1; Right Angular Gyrus | 55 | -51 | 31 | RH_Default_pCunPCC_2; Right Ventral Posterior Cingulate Cortex | 7 | -52 | 31 | 2.80 | 0.006 |
| LH_Vis_7; Left Visual Association Cortex | -47 | -71 | 11 | RH_Default_Temp_1; Right Medial Temporal Gyrus | 62 | -23 | -19 | 2.80 | 0.006 |
| RH_SomMot_8; Right Primary Motor Cortex | 6 | -26 | 70 | LH_Default_PFC_2; Left Pars Orbitalis | -47 | 33 | -3 | 2.79 | 0.006 |
| LH_Vis_2; Left Visual Association Cortex | -26 | -77 | -14 | LH_Default_PFC_2; Left Pars Orbitalis | -47 | 33 | -3 | 2.79 | 0.006 |
| LH_Vis_5; Left Secondary Visual Cortex | -5 | -92 | -2 | RH_Cont_Par_1; Right Supramarginal Gyrus | 57 | -39 | 44 | 2.79 | 0.006 |
| LH_SomMot_6; Left Primary Motor Cortex | -6 | -29 | 70 | RH_SomMot_6; Right Primary Motor Cortex | 41 | -22 | 60 | 2.78 | 0.006 |
| RH_Vis_6; Right Secondary Visual Cortex | 17 | -57 | 5 | RH_SalVentAttn_TempOccPar_2; Right Supramarginal Gyrus | 61 | -26 | 27 | 2.78 | 0.006 |
| LH_Vis_6; Left Primary Visual Cortex | -12 | -67 | 7 | RH_Default_pCunPCC_1; Right Ventral Posterior Cingulate Cortex | 12 | -54 | 14 | 2.78 | 0.006 |
| LH_DorsAttn_Post_6; Left Visual Motor Cortex | -22 | -51 | 66 | RH_Cont_Par_2; Right Angular Gyrus | 45 | -63 | 46 | 2.78 | 0.006 |
| LH_Vis_2; Left Visual Association Cortex | -26 | -77 | -14 | RH_Default_pCunPCC_1; Right Ventral Posterior Cingulate Cortex | 12 | -54 | 14 | 2.78 | 0.006 |
| LH_DorsAttn_Post_5; Left Visual Motor Cortex | -6 | -60 | 56 | LH_Cont_Par_1; Left Angular Gyrus | -37 | -53 | 46 | 2.77 | 0.006 |
| LH_DorsAttn_Post_5; Left Visual Motor Cortex | -6 | -60 | 56 | RH_Cont_Par_2; Right Angular Gyrus | 45 | -63 | 46 | 2.77 | 0.006 |
| LH_Vis_3; Left Visual Association Cortex | -17 | -60 | -7 | LH_Cont_Par_1; Left Angular Gyrus | -37 | -53 | 46 | 2.77 | 0.006 |
| LH_Vis_5; Left Secondary Visual Cortex | -5 | -92 | -2 | RH_DorsAttn_Post_2; Right Primary Sensory Cortex | 50 | -24 | 42 | 2.77 | 0.006 |
| LH_Vis_2; Left Visual Association Cortex | -26 | -77 | -14 | LH_Default_PFC_7; Left Frontal Eye Fields | -25 | 20 | 51 | 2.77 | 0.006 |
| LH_Vis_3; Left Visual Association Cortex | -17 | -60 | -7 | LH_DorsAttn_Post_3; Left Visual Motor Cortex | -24 | -68 | 49 | 2.77 | 0.006 |
| LH_DorsAttn_Post_5; Left Visual Motor Cortex | -6 | -60 | 56 | LH_Default_PFC_5; Left Dorsolateral Prefrontal Cortex | -9 | 48 | 41 | 2.76 | 0.006 |

| | | | | | | | | | |
|---|---|---|---|---|---|---|---|---|---|
| LH_SomMot_4; Left PreMotor & Supplementary Motor Area | -55 | -8 | 34 | LH_DorsAttn_Post_3; Left Visual Motor Cortex | -24 | -68 | 49 | 2.76 | 0.006 |
| LH_Cont_pCun_1; Left Visual Motor Cortex | -9 | -73 | 38 | RH_Default_Par_1; Right Angular Gyrus | 55 | -51 | 31 | 2.76 | 0.006 |
| RH_DorsAttn_Post_5; Right Visual Motor Cortex | 14 | -52 | 66 | LH_Default_pCunPCC_2; Left Ventral Posterior Cingulate Cortex | -6 | -53 | 33 | 2.76 | 0.007 |
| LH_Default_Par_2; Left Angular Gyrus | -48 | -63 | 35 | LH_Default_pCunPCC_1; Left Ventral Posterior Cingulate Cortex | -11 | -56 | 13 | 2.75 | 0.007 |
| RH_Vis_3; Right Fusiform Cortex | 49 | -60 | -11 | LH_Cont_Cing_1; Left Ventral Posterior Cingulate Cortex | -4 | -26 | 33 | 2.74 | 0.007 |
| LH_Vis_2; Left Visual Association Cortex | -26 | -77 | -14 | RH_DorsAttn_Post_2; Right Primary Sensory Cortex | 50 | -24 | 42 | 2.74 | 0.007 |
| LH_Default_pCunPCC_1; Left Ventral Posterior Cingulate Cortex | -11 | -56 | 13 | RH_Default_Par_1; Right Angular Gyrus | 55 | -51 | 31 | 2.74 | 0.007 |
| LH_Vis_8; Left Visual Association Cortex | -25 | -88 | 20 | RH_Vis_5; Right Primary Visual Cortex | 8 | -76 | 5 | 2.74 | 0.007 |
| RH_SalVentAttn_TempOccPar_1; Right Superior Temporal Gyrus | 58 | -42 | 13 | RH_Default_Temp_3; Right Superior Temporal Gyrus | 57 | -26 | -2 | 2.74 | 0.007 |
| RH_Vis_8; Right Visual Association Cortex | 13 | -86 | 29 | RH_Cont_pCun_1; Right Visual Motor Cortex | 9 | -66 | 43 | 2.74 | 0.007 |
| LH_Vis_2; Left Visual Association Cortex | -26 | -77 | -14 | LH_Default_pCunPCC_1; Left Ventral Posterior Cingulate Cortex | -11 | -56 | 13 | 2.73 | 0.007 |
| RH_Cont_Par_1; Right Supramarginal Gyrus | 57 | -39 | 44 | RH_Default_Temp_3; Right Superior Temporal Gyrus | 57 | -26 | -2 | 2.73 | 0.007 |
| LH_DorsAttn_FEF_1; Left PreMotor & Supplementary Motor Area | -26 | -3 | 59 | RH_Cont_Par_1; Right Supramarginal Gyrus | 57 | -39 | 44 | 2.73 | 0.007 |
| RH_Vis_3; Right Fusiform Cortex | 49 | -60 | -11 | RH_DorsAttn_Post_5; Right Visual Motor Cortex | 14 | -52 | 66 | 2.73 | 0.007 |
| RH_Vis_3; Right Fusiform Cortex | 49 | -60 | -11 | RH_SalVentAttn_Med_1; Right Dorsal Posterior Cingulate Cortex | 11 | -31 | 45 | 2.72 | 0.007 |
| LH_SomMot_5; Left Primary Motor Cortex | -39 | -23 | 59 | RH_Default_Temp_1; Right Medial Temporal Gyrus | 62 | -23 | -19 | 2.72 | 0.007 |
| LH_SomMot_2; Left Primary Sensory Cortex | -37 | -21 | 16 | LH_DorsAttn_Post_3; Left Visual Motor Cortex | -24 | -68 | 49 | 2.72 | 0.007 |
| RH_SomMot_8; Right Primary Motor Cortex | 6 | -26 | 70 | RH_DorsAttn_Post_4; Right Visual Motor Cortex | 27 | -67 | 51 | 2.72 | 0.007 |
| LH_Vis_7; Left Visual Association Cortex | -47 | -71 | 11 | LH_Default_PFC_3; Left Dorsal Posterior Cingulate Cortex | -6 | 47 | 0 | 2.72 | 0.007 |
| LH_Vis_7; Left Visual Association Cortex | -47 | -71 | 11 | RH_SomMot_7; Right Primary Sensory Cortex | 30 | -37 | 64 | 2.72 | 0.007 |
| LH_Cont_pCun_1; Left Visual Motor Cortex | -9 | -73 | 38 | RH_Default_pCunPCC_2; Right Ventral Posterior Cingulate Cortex | 7 | -52 | 31 | 2.71 | 0.007 |
| RH_SomMot_8; Right Primary Motor Cortex | 6 | -26 | 70 | LH_DorsAttn_Post_5; Left Visual Motor Cortex | -6 | -60 | 56 | 2.71 | 0.007 |
| LH_DorsAttn_Post_3; Left Visual Motor Cortex | -24 | -68 | 49 | RH_DorsAttn_Post_4; Right Visual Motor Cortex | 27 | -67 | 51 | 2.71 | 0.007 |
| RH_Vis_6; Right Secondary Visual Cortex | 17 | -57 | 5 | RH_DorsAttn_Post_2; Right Primary Sensory Cortex | 50 | -24 | 42 | 2.71 | 0.007 |
| LH_SalVentAttn_FrOperIns_2; Left Insula | -38 | 12 | 6 | LH_Cont_pCun_1; Left Visual Motor Cortex | -9 | -73 | 38 | 2.71 | 0.008 |
| LH_Cont_Par_1; Left Angular Gyrus | -37 | -53 | 46 | RH_Default_pCunPCC_2; Right Ventral Posterior Cingulate Cortex | 7 | -52 | 31 | 2.71 | 0.008 |
| LH_Vis_5; Left Secondary Visual Cortex | -5 | -92 | -2 | RH_Vis_5; Right Primary Visual Cortex | 8 | -76 | 5 | 2.70 | 0.008 |
| LH_Cont_Par_1; Left Angular Gyrus | -37 | -53 | 46 | RH_Default_pCunPCC_1; Right Ventral Posterior Cingulate Cortex | 12 | -54 | 14 | 2.70 | 0.008 |
| RH_Vis_8; Right Visual Association Cortex | 13 | -86 | 29 | LH_DorsAttn_Post_5; Left Visual Motor Cortex | -6 | -60 | 56 | 2.70 | 0.008 |
| RH_Vis_4; Right Secondary Visual Cortex | 22 | -96 | -5 | LH_DorsAttn_Post_3; Left Visual Motor Cortex | -24 | -68 | 49 | 2.70 | 0.008 |
| LH_Default_PFC_6; Left PreMotor & Supplementary Motor Area | -41 | 14 | 48 | RH_Default_Temp_1; Right Medial Temporal Gyrus | 62 | -23 | -19 | 2.70 | 0.008 |
| RH_SalVentAttn_TempOccPar_1; Right Superior Temporal Gyrus | 58 | -42 | 13 | LH_Cont_pCun_1; Left Visual Motor Cortex | -9 | -73 | 38 | 2.70 | 0.008 |

| | | | | | | | | | |
|---|---|---|---|---|---|---|---|---|---|
| RH_Vis_3; Right Fusiform Cortex | 49 | -60 | -11 | LH_DorsAttn_Post_5; Left Visual Motor Cortex | -6 | -60 | 56 | 2.70 | 0.008 |
| LH_Vis_7; Left Visual Association Cortex | -47 | -71 | 11 | RH_DorsAttn_PrCv_1; Right Broca's Area/Operculum | 49 | 10 | 27 | 2.70 | 0.008 |
| RH_Vis_2; Right Fusiform Cortex | 27 | -66 | -12 | LH_SalVentAttn_Med_2; Left Dorsal Posterior Cingulate Cortex | -11 | -34 | 45 | 2.70 | 0.008 |
| LH_Vis_2; Left Visual Association Cortex | -26 | -77 | -14 | RH_SalVentAttn_FrOperIns_1; Right Insula | 40 | 8 | 1 | 2.70 | 0.008 |
| LH_Vis_6; Left Primary Visual Cortex | -12 | -67 | 7 | RH_Vis_6; Right Secondary Visual Cortex | 17 | -57 | 5 | 2.69 | 0.008 |
| RH_DorsAttn_Post_1; Right Visual Association Cortex | 50 | -62 | 16 | RH_DorsAttn_Post_5; Right Visual Motor Cortex | 14 | -52 | 66 | 2.69 | 0.008 |
| LH_Default_Par_1; Left Angular Gyrus | -57 | -50 | 12 | RH_Default_Temp_3; Right Superior Temporal Gyrus | 57 | -26 | -2 | 2.69 | 0.008 |
| LH_DorsAttn_Post_6; Left Visual Motor Cortex | -22 | -51 | 66 | LH_Cont_pCun_1; Left Visual Motor Cortex | -9 | -73 | 38 | 2.69 | 0.008 |
| LH_Vis_6; Left Primary Visual Cortex | -12 | -67 | 7 | RH_SalVentAttn_TempOccPar_1; Right Superior Temporal Gyrus | 58 | -42 | 13 | 2.68 | 0.008 |
| RH_Cont_Par_1; Right Supramarginal Gyrus | 57 | -39 | 44 | RH_Default_Temp_1; Right Medial Temporal Gyrus | 62 | -23 | -19 | 2.68 | 0.008 |
| LH_Vis_3; Left Visual Association Cortex | -17 | -60 | -7 | RH_SalVentAttn_FrOperIns_1; Right Insula | 40 | 8 | 1 | 2.67 | 0.008 |
| LH_SomMot_6; Left Primary Motor Cortex | -6 | -29 | 70 | LH_Default_pCunPCC_2; Left Ventral Posterior Cingulate Cortex | -6 | -53 | 33 | 2.67 | 0.008 |
| RH_DorsAttn_Post_5; Right Visual Motor Cortex | 14 | -52 | 66 | LH_SalVentAttn_Med_2; Left Dorsal Posterior Cingulate Cortex | -11 | -34 | 45 | 2.67 | 0.008 |
| RH_DorsAttn_Post_5; Right Visual Motor Cortex | 14 | -52 | 66 | RH_Default_pCunPCC_1; Right Ventral Posterior Cingulate Cortex | 12 | -54 | 14 | 2.67 | 0.008 |
| RH_Vis_2; Right Fusiform Cortex | 27 | -66 | -12 | LH_SomMot_6; Left Primary Motor Cortex | -6 | -29 | 70 | 2.67 | 0.008 |
| LH_Vis_9; Left Visual Association Cortex | -6 | -82 | 26 | RH_Default_Par_1; Right Angular Gyrus | 55 | -51 | 31 | 2.67 | 0.008 |
| LH_Vis_2; Left Visual Association Cortex | -26 | -77 | -14 | LH_Cont_Par_1; Left Angular Gyrus | -37 | -53 | 46 | 2.67 | 0.008 |
| LH_Vis_6; Left Primary Visual Cortex | -12 | -67 | 7 | RH_SalVentAttn_TempOccPar_2; Right Supramarginal Gyrus | 61 | -26 | 27 | 2.67 | 0.008 |
| LH_Cont_pCun_1; Left Visual Motor Cortex | -9 | -73 | 38 | RH_Default_PFCdPFCm_2; Right Dorsolateral Prefrontal Cortex | 11 | 50 | 39 | 2.67 | 0.008 |
| RH_DorsAttn_Post_4; Right Visual Motor Cortex | 27 | -67 | 51 | RH_Default_PFCdPFCm_3; Right Frontal Eye Fields | 26 | 24 | 50 | 2.66 | 0.009 |
| LH_DorsAttn_Post_5; Left Visual Motor Cortex | -6 | -60 | 56 | LH_Default_pCunPCC_2; Left Ventral Posterior Cingulate Cortex | -6 | -53 | 33 | 2.66 | 0.009 |
| LH_DorsAttn_Post_5; Left Visual Motor Cortex | -6 | -60 | 56 | LH_Default_Temp_1; Left Medial Temporal Gyrus | -55 | -4 | -20 | 2.66 | 0.009 |
| RH_SomMot_2; Right Insula | 40 | -15 | 15 | LH_Default_pCunPCC_1; Left Ventral Posterior Cingulate Cortex | -11 | -56 | 13 | 2.65 | 0.009 |
| LH_Vis_7; Left Visual Association Cortex | -47 | -71 | 11 | LH_SalVentAttn_Med_1; Left Dorsal Anterior Cingulate Cortex | -5 | 20 | 34 | 2.65 | 0.009 |
| RH_Limbic_TempPole_1; Right Inferior Temporal Gyrus | 38 | 1 | -38 | RH_Default_PFCv_2; Right Broca's Area/ Pars Triangularis | 51 | 28 | 0 | 2.65 | 0.009 |
| RH_SomMot_7; Right Primary Sensory Cortex | 30 | -37 | 64 | RH_Default_pCunPCC_2; Right Ventral Posterior Cingulate Cortex | 7 | -52 | 31 | 2.64 | 0.009 |
| LH_DorsAttn_Post_5; Left Visual Motor Cortex | -6 | -60 | 56 | RH_Default_PFCdPFCm_2; Right Dorsolateral Prefrontal Cortex | 11 | 50 | 39 | 2.64 | 0.009 |
| RH_Vis_2; Right Fusiform Cortex | 27 | -66 | -12 | RH_Default_pCunPCC_2; Right Ventral Posterior Cingulate Cortex | 7 | -52 | 31 | 2.64 | 0.009 |
| RH_SomMot_7; Right Primary Sensory Cortex | 30 | -37 | 64 | RH_Default_pCunPCC_1; Right Ventral Posterior Cingulate Cortex | 12 | -54 | 14 | 2.64 | 0.009 |
| RH_SomMot_3; Right Primary Motor Cortex | 57 | -4 | 11 | RH_Default_PFCdPFCm_2; Right Dorsolateral Prefrontal Cortex | 11 | 50 | 39 | -2.63 | 0.009 |
| LH_Default_Par_2; Left Angular Gyrus | -48 | -63 | 35 | LH_Default_pCunPCC_2; Left Ventral Posterior Cingulate Cortex | -6 | -53 | 33 | 2.62 | 0.010 |
| RH_Vis_5; Right Primary Visual Cortex | 8 | -76 | 5 | RH_Vis_6; Right Secondary Visual Cortex | 17 | -57 | 5 | 2.62 | 0.010 |

| | | | | | | | | | |
|---|---|---|---|---|---|---|---|---|---|
| LH_SomMot_6; Left Primary Motor Cortex | -6 | -29 | 70 | RH_Default_pCunPCC_2; Right Ventral Posterior Cingulate Cortex | 7 | -52 | 31 | 2.62 | 0.010 |
| LH_DorsAttn_Post_5; Left Visual Motor Cortex | -6 | -60 | 56 | RH_DorsAttn_Post_1; Right Visual Association Cortex | 50 | -62 | 16 | 2.62 | 0.010 |
| LH_Vis_2; Left Visual Association Cortex | -26 | -77 | -14 | RH_Default_PFCv_1; Right Pars Orbitalis | 35 | 26 | -15 | 2.62 | 0.010 |
| LH_SomMot_5; Left Primary Motor Cortex | -39 | -23 | 59 | LH_Default_PFC_2; Left Pars Orbitalis | -47 | 33 | -3 | 2.62 | 0.010 |
| RH_DorsAttn_Post_5; Right Visual Motor Cortex | 14 | -52 | 66 | RH_DorsAttn_FEF_1; Right PreMotor & Supplementary Motor Area | 28 | -3 | 59 | 2.61 | 0.010 |
| LH_Vis_4; Left Secondary Visual Cortex | -27 | -95 | -4 | RH_DorsAttn_Post_3; Right Visual Motor Cortex | 38 | -45 | 49 | 2.60 | 0.010 |
| RH_SomMot_8; Right Primary Motor Cortex | 6 | -26 | 70 | LH_Default_Par_1; Left Angular Gyrus | -57 | -50 | 12 | 2.60 | 0.010 |
| LH_Vis_5; Left Secondary Visual Cortex | -5 | -92 | -2 | LH_Vis_8; Left Visual Association Cortex | -25 | -88 | 20 | 2.60 | 0.010 |
| LH_Vis_7; Left Visual Association Cortex | -47 | -71 | 11 | LH_SalVentAttn_Med_2; Left Dorsal Posterior Cingulate Cortex | -11 | -34 | 45 | 2.60 | 0.010 |
| RH_Vis_4; Right Secondary Visual Cortex | 22 | -96 | -5 | RH_DorsAttn_Post_2; Right Primary Sensory Cortex | 50 | -24 | 42 | 2.60 | 0.010 |
| LH_Vis_2; Left Visual Association Cortex | -26 | -77 | -14 | RH_SomMot_1; Right Primary Auditory Cortex | 53 | -16 | 7 | 2.59 | 0.010 |

**Eyes-Closed Gamma**

| **ROI1 Labels (Schaefer; Talairach)** | **X** | **Y** | **Z** | **ROI2 Labels (Schaefer; Talairach)** | **X** | **Y** | **Z** | **t** | **p (unc)** |
|---|---|---|---|---|---|---|---|---|---|
| LH_Vis_7; Left Visual Association Cortex | -47 | -71 | 11 | RH_Default_PFCv_2; Right Broca's Area/ Pars Triangularis | 51 | 28 | 0 | -3.59 | 0.0004 |
| LH_Vis_6; Left Primary Visual Cortex | -12 | -67 | 7 | LH_Default_PFC_4; Left Anterior Prefrontal Cortex | -24 | 61 | -1 | -3.40 | 0.0009 |
| RH_Cont_Cing_1; Right Ventral Posterior Cingulate Cortex | 5 | -27 | 33 | RH_Default_PFCdPFCm_1; Right Anterior Prefrontal Cortex | 7 | 48 | 1 | -3.38 | 0.0009 |
| LH_DorsAttn_PrCv_1; Left PreMotor & Supplementary Motor Area | -48 | 6 | 28 | LH_Default_Par_1; Left Angular Gyrus | -57 | -50 | 12 | 3.32 | 0.001 |
| LH_Vis_4; Left Secondary Visual Cortex | -27 | -95 | -4 | LH_SomMot_6; Left Primary Motor Cortex | -6 | -29 | 70 | 3.30 | 0.001 |
| LH_DorsAttn_PrCv_1; Left PreMotor & Supplementary Motor Area | -48 | 6 | 28 | RH_Default_Temp_1; Right Medial Temporal Gyrus | 62 | -23 | -19 | 3.15 | 0.002 |
| LH_DorsAttn_PrCv_1; Left PreMotor & Supplementary Motor Area | -48 | 6 | 28 | RH_Default_Temp_3; Right Superior Temporal Gyrus | 57 | -26 | -2 | 3.07 | 0.003 |
| RH_SalVentAttn_Med_1; Right Dorsal Posterior Cingulate Cortex | 11 | -31 | 45 | RH_Default_PFCdPFCm_1; Right Anterior Prefrontal Cortex | 7 | 48 | 1 | -2.95 | 0.004 |
| LH_Limbic_OFC_1; Left Orbitofrontal Cortex | -14 | 32 | -20 | LH_Limbic_TempPole_1; Left Temporal Pole | -32 | 2 | -37 | 2.87 | 0.005 |
| RH_Cont_PFCl_3; Right Dorsolateral Prefrontal Cortex | 32 | 46 | 29 | RH_Default_Par_1; Right Angular Gyrus | 55 | -51 | 31 | -2.84 | 0.005 |
| RH_SalVentAttn_Med_1; Right Dorsal Posterior Cingulate Cortex | 11 | -31 | 45 | RH_Default_PFCdPFCm_2; Right Dorsolateral Prefrontal Cortex | 11 | 50 | 39 | -2.83 | 0.005 |
| LH_DorsAttn_Post_3; Left Visual Motor Cortex | -24 | -68 | 49 | LH_Default_PFC_4; Left Anterior Prefrontal Cortex | -24 | 61 | -1 | -2.78 | 0.006 |
| RH_Vis_6; Right Secondary Visual Cortex | 17 | -57 | 5 | LH_Default_pCunPCC_2; Left Ventral Posterior Cingulate Cortex | -6 | -53 | 33 | -2.73 | 0.007 |
| RH_DorsAttn_Post_3; Right Visual Motor Cortex | 38 | -45 | 49 | LH_Default_pCunPCC_2; Left Ventral Posterior Cingulate Cortex | -6 | -53 | 33 | -2.72 | 0.007 |
| LH_Vis_5; Left Secondary Visual Cortex | -5 | -92 | -2 | LH_SomMot_6; Left Primary Motor Cortex | -6 | -29 | 70 | 2.71 | 0.008 |
| RH_Cont_pCun_1; Right Visual Motor Cortex | 9 | -66 | 43 | LH_Default_pCunPCC_1; Left Ventral Posterior Cingulate Cortex | -11 | -56 | 13 | 2.69 | 0.008 |
| LH_SomMot_5; Left Primary Motor Cortex | -39 | -23 | 59 | RH_Default_Temp_1; Right Medial Temporal Gyrus | 62 | -23 | -19 | 2.68 | 0.008 |
| RH_Cont_Par_2; Right Angular Gyrus | 45 | -63 | 46 | LH_Default_pCunPCC_2; Left Ventral Posterior Cingulate Cortex | -6 | -53 | 33 | -2.67 | 0.008 |
| RH_Vis_7; Right Visual Association Cortex | 36 | -82 | 16 | LH_Cont_Par_1; Left Angular Gyrus | -37 | -53 | 46 | -2.67 | 0.008 |

| | | | | | | | | | |
|---|---|---|---|---|---|---|---|---|---|
| RH_Limbic_OFC_1; Right Orbitofrontal Cortex | 12 | 35 | -20 | RH_Default_PFCdPFCm_1; Right Anterior Prefrontal Cortex | 7 | 48 | 1 | 2.67 | 0.008 |
| LH_Limbic_OFC_1; Left Orbitofrontal Cortex | -14 | 32 | -20 | LH_Default_pCunPCC_2; Left Ventral Posterior Cingulate Cortex | -6 | -53 | 33 | -2.61 | 0.010 |
| LH_SomMot_3; Left Primary Sensory Cortex | -54 | -12 | 13 | LH_Default_PFC_5; Left Dorsolateral Prefrontal Cortex | -9 | 48 | 41 | 2.60 | 0.010 |
| LH_Cont_Cing_1; Left Ventral Posterior Cingulate Cortex | -4 | -26 | 33 | RH_Default_PFCdPFCm_1; Right Anterior Prefrontal Cortex | 7 | 48 | 1 | -2.60 | 0.010 |
| LH_Vis_7; Left Visual Association Cortex | -47 | -71 | 11 | LH_DorsAttn_Post_6; Left Visual Motor Cortex | -22 | -51 | 66 | 2.59 | 0.010 |
| RH_DorsAttn_Post_1; Right Visual Association Cortex | 50 | -62 | 16 | LH_SalVentAttn_PFCl_1; Left Anterior Prefrontal Cortex | -30 | 44 | 30 | -2.59 | 0.010 |

$p \leq 0.01$ uncorrected

**Supplementary Table 9 Functional connectivity results for higher vs. lower menstrual pain groups at post-menarche visit 2**

| **Eyes-Open Theta** | | | | | | | | | |
|---|---|---|---|---|---|---|---|---|---|
| **ROI1 Labels (Schaefer; Talairach)** | **X** | **Y** | **Z** | **ROI2 Labels (Schaefer; Talairach)** | **X** | **Y** | **Z** | **t** | **p (unc)** |
| LH_SalVentAttn_FrOperIns_1; Left Insula | -41 | -1 | -7 | LH_Default_PFC_1; Left Pars Orbitalis | -35 | 21 | -11 | 3.38 | 0.001 |
| LH_Cont_Par_1; Left Angular Gyrus | -37 | -53 | 46 | RH_Default_PFCv_1; Right Pars Orbitalis | 35 | 26 | -15 | 3.33 | 0.001 |
| RH_Vis_7; Right Visual Association Cortex | 36 | -82 | 16 | LH_DorsAttn_PrCv_1; Left PreMotor & Supplementary Motor Area | -48 | 6 | 28 | 3.25 | 0.001 |
| RH_Vis_7; Right Visual Association Cortex | 36 | -82 | 16 | LH_Cont_Par_1; Left Angular Gyrus | -37 | -53 | 46 | 3.23 | 0.002 |
| RH_SomMot_3; Right Primary Motor Cortex | 57 | -4 | 11 | LH_DorsAttn_Post_6; Left Visual Motor Cortex | -22 | -51 | 66 | 3.15 | 0.002 |
| LH_Vis_1; Left Fusiform Cortex | -26 | -34 | -17 | RH_SomMot_4; Right Primary Motor Cortex | 58 | -5 | 31 | 3.02 | 0.003 |
| RH_Vis_5; Right Primary Visual Cortex | 8 | -76 | 5 | RH_SalVentAttn_Med_1; Right Dorsal Posterior Cingulate Cortex | 11 | -31 | 45 | 3.02 | 0.003 |
| LH_DorsAttn_Post_6; Left Visual Motor Cortex | -22 | -51 | 66 | RH_Cont_PFCmp_1; Right Frontal Eye Fields | 6 | 28 | 30 | 2.96 | 0.004 |
| LH_Vis_6; Left Primary Visual Cortex | -12 | -67 | 7 | RH_DorsAttn_PrCv_1; Right Broca's Area/Operculum | 49 | 10 | 27 | 2.93 | 0.004 |
| LH_SalVentAttn_Med_2; Left Dorsal Posterior Cingulate Cortex | -11 | -34 | 45 | RH_Cont_PFCmp_1; Right Frontal Eye Fields | 6 | 28 | 30 | 2.90 | 0.004 |
| RH_SalVentAttn_Med_1; Right Dorsal Posterior Cingulate Cortex | 11 | -31 | 45 | RH_Default_pCunPCC_1; Right Ventral Posterior Cingulate Cortex | 12 | -54 | 14 | 2.90 | 0.004 |
| LH_SomMot_3; Left Primary Sensory Cortex | -54 | -12 | 13 | LH_Default_PFC_1; Left Pars Orbitalis | -35 | 21 | -11 | 2.87 | 0.005 |
| RH_DorsAttn_PrCv_1; Right Broca's Area/Operculum | 49 | 10 | 27 | RH_SalVentAttn_Med_1; Right Dorsal Posterior Cingulate Cortex | 11 | -31 | 45 | 2.87 | 0.005 |
| LH_DorsAttn_Post_3; Left Visual Motor Cortex | -24 | -68 | 49 | RH_Cont_PFCmp_1; Right Frontal Eye Fields | 6 | 28 | 30 | 2.85 | 0.005 |
| RH_Vis_7; Right Visual Association Cortex | 36 | -82 | 16 | LH_Cont_Cing_1; Left Ventral Posterior Cingulate Cortex | -4 | -26 | 33 | 2.85 | 0.005 |
| LH_Vis_5; Left Secondary Visual Cortex | -5 | -92 | -2 | RH_Cont_Cing_1; Right Ventral Posterior Cingulate Cortex | 5 | -27 | 33 | 2.84 | 0.005 |
| LH_Vis_3; Left Visual Association Cortex | -17 | -60 | -7 | RH_SomMot_3; Right Primary Motor Cortex | 57 | -4 | 11 | 2.83 | 0.005 |
| RH_SomMot_1; Right Primary Auditory Cortex | 53 | -16 | 7 | LH_DorsAttn_Post_6; Left Visual Motor Cortex | -22 | -51 | 66 | 2.82 | 0.006 |
| RH_Cont_pCun_1; Right Visual Motor Cortex | 9 | -66 | 43 | RH_Default_PFCv_2; Right Broca's Area/ Pars Triangularis | 51 | 28 | 0 | 2.82 | 0.006 |
| RH_Vis_7; Right Visual Association Cortex | 36 | -82 | 16 | RH_Cont_Cing_1; Right Ventral Posterior Cingulate Cortex | 5 | -27 | 33 | 2.79 | 0.006 |
| LH_DorsAttn_Post_6; Left Visual Motor Cortex | -22 | -51 | 66 | RH_DorsAttn_PrCv_1; Right Broca's Area/Operculum | 49 | 10 | 27 | 2.78 | 0.006 |
| LH_Vis_3; Left Visual Association Cortex | -17 | -60 | -7 | RH_Cont_Cing_1; Right Ventral Posterior Cingulate Cortex | 5 | -27 | 33 | 2.77 | 0.006 |
| RH_Cont_Cing_1; Right Ventral Posterior Cingulate Cortex | 5 | -27 | 33 | LH_Default_pCunPCC_2; Left Ventral Posterior Cingulate Cortex | -6 | -53 | 33 | 2.77 | 0.006 |
| LH_Vis_5; Left Secondary Visual Cortex | -5 | -92 | -2 | RH_SalVentAttn_Med_1; Right Dorsal Posterior Cingulate Cortex | 11 | -31 | 45 | 2.77 | 0.006 |
| LH_DorsAttn_Post_6; Left Visual Motor Cortex | -22 | -51 | 66 | LH_Default_PFC_4; Left Anterior Prefrontal Cortex | -24 | 61 | -1 | 2.77 | 0.006 |
| LH_DorsAttn_Post_6; Left Visual Motor Cortex | -22 | -51 | 66 | RH_SalVentAttn_FrOperIns_1; Right Insula | 40 | 8 | 1 | 2.73 | 0.007 |
| LH_Cont_Par_1; Left Angular Gyrus | -37 | -53 | 46 | RH_Cont_PFCmp_1; Right Frontal Eye Fields | 6 | 28 | 30 | 2.73 | 0.007 |
| LH_Default_PFC_1; Left Pars Orbitalis | -35 | 21 | -11 | RH_Default_PFCdPFCm_2; Right Dorsolateral Prefrontal Cortex | 11 | 50 | 39 | 2.71 | 0.008 |

| | | | | | | | | | |
|---|---|---|---|---|---|---|---|---|---|
| RH_SomMot_4; Right Primary Motor Cortex | 58 | -5 | 31 | LH_Default_pCunPCC_2; Left Ventral Posterior Cingulate Cortex | -6 | -53 | 33 | 2.69 | 0.008 |
| LH_Default_PFC_1; Left Pars Orbitalis | -35 | 21 | -11 | LH_Default_PFC_2; Left Pars Orbitalis | -47 | 33 | -3 | 2.66 | 0.009 |
| RH_SalVentAttn_Med_1; Right Dorsal Posterior Cingulate Cortex | 11 | -31 | 45 | LH_Cont_Cing_1; Left Ventral Posterior Cingulate Cortex | -4 | -26 | 33 | 2.66 | 0.009 |
| LH_Cont_pCun_1; Left Visual Motor Cortex | -9 | -73 | 38 | RH_Cont_Cing_1; Right Ventral Posterior Cingulate Cortex | 5 | -27 | 33 | 2.65 | 0.009 |
| LH_Vis_3; Left Visual Association Cortex | -17 | -60 | -7 | LH_Cont_Cing_1; Left Ventral Posterior Cingulate Cortex | -4 | -26 | 33 | 2.65 | 0.009 |
| LH_Vis_2; Left Visual Association Cortex | -26 | -77 | -14 | RH_Default_Temp_2; Right Temporal Pole | 51 | 7 | -18 | 2.65 | 0.009 |
| LH_Cont_Par_1; Left Angular Gyrus | -37 | -53 | 46 | RH_Cont_PFCl_2; Right Dorsolateral Prefrontal Cortex | 45 | 39 | 15 | 2.64 | 0.009 |
| RH_SalVentAttn_FrOperIns_1; Right Insula | 40 | 8 | 1 | LH_Default_pCunPCC_2; Left Ventral Posterior Cingulate Cortex | -6 | -53 | 33 | 2.64 | 0.009 |
| LH_Vis_6; Left Primary Visual Cortex | -12 | -67 | 7 | RH_SomMot_3; Right Primary Motor Cortex | 57 | -4 | 11 | 2.63 | 0.009 |
| LH_Vis_3; Left Visual Association Cortex | -17 | -60 | -7 | RH_SalVentAttn_Med_1; Right Dorsal Posterior Cingulate Cortex | 11 | -31 | 45 | 2.63 | 0.010 |
| RH_Vis_6; Right Secondary Visual Cortex | 17 | -57 | 5 | RH_SalVentAttn_Med_1; Right Dorsal Posterior Cingulate Cortex | 11 | -31 | 45 | 2.63 | 0.010 |

**Eyes-Open Alpha**

| ROI1 Labels (Schaefer; Talairach) | X | Y | Z | ROI2 Labels (Schaefer; Talairach) | X | Y | Z | t | p (unc) |
|---|---|---|---|---|---|---|---|---|---|
| LH_Vis_4; Left Secondary Visual Cortex | -27 | -95 | -4 | LH_SomMot_6; Left Primary Motor Cortex | -6 | -29 | 70 | 3.94 | 0.0001 |
| LH_Vis_8; Left Visual Association Cortex | -25 | -88 | 20 | RH_DorsAttn_PrCv_1; Right Broca's Area/Operculum | 49 | 10 | 27 | 3.93 | 0.0001 |
| RH_Vis_8; Right Visual Association Cortex | 13 | -86 | 29 | LH_DorsAttn_Post_4; Left Primary Sensory Cortex | -42 | -34 | 48 | 3.81 | 0.0002 |
| LH_Vis_2; Left Visual Association Cortex | -26 | -77 | -14 | RH_DorsAttn_FEF_1; Right PreMotor & Supplementary Motor Area | 28 | -3 | 59 | 3.72 | 0.0003 |
| LH_DorsAttn_Post_4; Left Primary Sensory Cortex | -42 | -34 | 48 | LH_Cont_pCun_1; Left Visual Motor Cortex | -9 | -73 | 38 | 3.62 | 0.0004 |
| RH_DorsAttn_FEF_1; Right PreMotor & Supplementary Motor Area | 28 | -3 | 59 | LH_Cont_pCun_1; Left Visual Motor Cortex | -9 | -73 | 38 | 3.59 | 0.0005 |
| LH_Vis_7; Left Visual Association Cortex | -47 | -71 | 11 | RH_SomMot_8; Right Primary Motor Cortex | 6 | -26 | 70 | 3.57 | 0.001 |
| LH_SalVentAttn_ParOper_1; Left Supramarginal Gyrus | -59 | -38 | 29 | RH_Limbic_TempPole_1; Right Inferior Temporal Gyrus | 38 | 1 | -38 | 3.56 | 0.001 |
| LH_Vis_7; Left Visual Association Cortex | -47 | -71 | 11 | RH_DorsAttn_FEF_1; Right PreMotor & Supplementary Motor Area | 28 | -3 | 59 | 3.56 | 0.001 |
| LH_SomMot_1; Left Supramarginal Gyrus | -53 | -23 | 8 | RH_Default_PFCv_1; Right Pars Orbitalis | 35 | 26 | -15 | 3.49 | 0.001 |
| LH_DorsAttn_PrCv_1; Left PreMotor & Supplementary Motor Area | -48 | 6 | 28 | RH_Default_Temp_2; Right Temporal Pole | 51 | 7 | -18 | 3.46 | 0.001 |
| LH_SalVentAttn_ParOper_1; Left Supramarginal Gyrus | -59 | -38 | 29 | RH_Default_PFCv_1; Right Pars Orbitalis | 35 | 26 | -15 | 3.45 | 0.001 |
| RH_DorsAttn_PrCv_1; Right Broca's Area/Operculum | 49 | 10 | 27 | LH_SalVentAttn_ParOper_1; Left Supramarginal Gyrus | -59 | -38 | 29 | 3.40 | 0.001 |
| LH_SomMot_5; Left Primary Motor Cortex | -39 | -23 | 59 | LH_Limbic_TempPole_1; Left Temporal Pole | -32 | 2 | -37 | 3.36 | 0.001 |
| LH_Vis_8; Left Visual Association Cortex | -25 | -88 | 20 | RH_SomMot_5; Right PreMotor & Supplementary Motor Area | 47 | -11 | 48 | 3.35 | 0.001 |
| LH_DorsAttn_Post_3; Left Visual Motor Cortex | -24 | -68 | 49 | LH_DorsAttn_Post_4; Left Primary Sensory Cortex | -42 | -34 | 48 | 3.35 | 0.001 |
| LH_Vis_9; Left Visual Association Cortex | -6 | -82 | 26 | LH_DorsAttn_Post_5; Left Visual Motor Cortex | -6 | -60 | 56 | 3.33 | 0.001 |
| LH_SomMot_5; Left Primary Motor Cortex | -39 | -23 | 59 | LH_DorsAttn_Post_3; Left Visual Motor Cortex | -24 | -68 | 49 | 3.32 | 0.001 |
| RH_SomMot_8; Right Primary Motor Cortex | 6 | -26 | 70 | LH_Default_Temp_2; Left Medial Temporal Gyrus | -58 | -32 | -1 | 3.26 | 0.001 |

| | | | | | | | | | |
|---|---|---|---|---|---|---|---|---|---|
| LH_Vis_8; Left Visual Association Cortex | -25 | -88 | 20 | RH_Limbic_TempPole_1; Right Inferior Temporal Gyrus | 38 | 1 | -38 | 3.25 | 0.001 |
| RH_Cont_PFCl_4; Right Frontal Eye Fields | 43 | 16 | 45 | LH_Default_pCunPCC_1; Left Ventral Posterior Cingulate Cortex | -11 | -56 | 13 | 3.24 | 0.002 |
| LH_Vis_8; Left Visual Association Cortex | -25 | -88 | 20 | LH_SalVentAttn_PFCl_1; Left Anterior Prefrontal Cortex | -30 | 44 | 30 | 3.22 | 0.002 |
| LH_SomMot_3; Left Primary Sensory Cortex | -54 | -12 | 13 | RH_Limbic_TempPole_1; Right Inferior Temporal Gyrus | 38 | 1 | -38 | 3.22 | 0.002 |
| LH_Vis_9; Left Visual Association Cortex | -6 | -82 | 26 | RH_Default_PFCv_1; Right Pars Orbitalis | 35 | 26 | -15 | 3.22 | 0.002 |
| LH_Vis_8; Left Visual Association Cortex | -25 | -88 | 20 | RH_SomMot_4; Right Primary Motor Cortex | 58 | -5 | 31 | 3.17 | 0.002 |
| LH_Vis_7; Left Visual Association Cortex | -47 | -71 | 11 | LH_Cont_Par_1; Left Angular Gyrus | -37 | -53 | 46 | 3.16 | 0.002 |
| LH_DorsAttn_Post_1; Left Fusiform Cortex | -47 | -58 | -13 | RH_Cont_PFCl_4; Right Frontal Eye Fields | 43 | 16 | 45 | 3.16 | 0.002 |
| LH_Vis_8; Left Visual Association Cortex | -25 | -88 | 20 | LH_SomMot_6; Left Primary Motor Cortex | -6 | -29 | 70 | 3.15 | 0.002 |
| LH_SalVentAttn_ParOper_1; Left Supramarginal Gyrus | -59 | -38 | 29 | LH_Cont_pCun_1; Left Visual Motor Cortex | -9 | -73 | 38 | 3.13 | 0.002 |
| LH_Vis_9; Left Visual Association Cortex | -6 | -82 | 26 | RH_DorsAttn_FEF_1; Right PreMotor & Supplementary Motor Area | 28 | -3 | 59 | 3.12 | 0.002 |
| LH_Vis_9; Left Visual Association Cortex | -6 | -82 | 26 | RH_DorsAttn_PrCv_1; Right Broca's Area/Operculum | 49 | 10 | 27 | 3.11 | 0.002 |
| LH_Vis_8; Left Visual Association Cortex | -25 | -88 | 20 | LH_SalVentAttn_FrOperIns_2; Left Insula | -38 | 12 | 6 | 3.08 | 0.002 |
| LH_Vis_7; Left Visual Association Cortex | -47 | -71 | 11 | RH_Cont_PFCl_4; Right Frontal Eye Fields | 43 | 16 | 45 | 3.07 | 0.003 |
| LH_Default_Par_2; Left Angular Gyrus | -48 | -63 | 35 | RH_Default_PFCv_1; Right Pars Orbitalis | 35 | 26 | -15 | 3.06 | 0.003 |
| LH_SalVentAttn_FrOperIns_1; Left Insula | -41 | -1 | -7 | LH_Default_PFC_3; Left Dorsal Posterior Cingulate Cortex | -6 | 47 | 0 | 3.06 | 0.003 |
| LH_DorsAttn_Post_4; Left Primary Sensory Cortex | -42 | -34 | 48 | RH_Cont_PFCl_4; Right Frontal Eye Fields | 43 | 16 | 45 | 3.05 | 0.003 |
| LH_SomMot_3; Left Primary Sensory Cortex | -54 | -12 | 13 | RH_Default_Temp_2; Right Temporal Pole | 51 | 7 | -18 | 3.04 | 0.003 |
| LH_SalVentAttn_ParOper_1; Left Supramarginal Gyrus | -59 | -38 | 29 | LH_Limbic_OFC_1; Left Orbitofrontal Cortex | -14 | 32 | -20 | 3.03 | 0.003 |
| LH_Default_pCunPCC_2; Left Ventral Posterior Cingulate Cortex | -6 | -53 | 33 | RH_Default_Temp_2; Right Temporal Pole | 51 | 7 | -18 | 3.02 | 0.003 |
| LH_Vis_8; Left Visual Association Cortex | -25 | -88 | 20 | LH_DorsAttn_Post_5; Left Visual Motor Cortex | -6 | -60 | 56 | 3.00 | 0.003 |
| LH_Limbic_TempPole_1; Left Temporal Pole | -32 | 2 | -37 | LH_Default_Par_2; Left Angular Gyrus | -48 | -63 | 35 | 2.99 | 0.003 |
| LH_Vis_2; Left Visual Association Cortex | -26 | -77 | -14 | LH_DorsAttn_PrCv_1; Left PreMotor & Supplementary Motor Area | -48 | 6 | 28 | 2.99 | 0.003 |
| RH_Limbic_TempPole_1; Right Inferior Temporal Gyrus | 38 | 1 | -38 | LH_Default_Par_1; Left Angular Gyrus | -57 | -50 | 12 | 2.98 | 0.003 |
| LH_SalVentAttn_ParOper_1; Left Supramarginal Gyrus | -59 | -38 | 29 | RH_Cont_PFCl_3; Right Dorsolateral Prefrontal Cortex | 32 | 46 | 29 | 2.98 | 0.003 |
| LH_SomMot_1; Left Supramarginal Gyrus | -53 | -23 | 8 | RH_Default_Temp_2; Right Temporal Pole | 51 | 7 | -18 | 2.98 | 0.003 |
| LH_Vis_1; Left Fusiform Cortex | -26 | -34 | -17 | RH_Cont_PFCl_4; Right Frontal Eye Fields | 43 | 16 | 45 | 2.98 | 0.003 |
| LH_SomMot_6; Left Primary Motor Cortex | -6 | -29 | 70 | LH_SalVentAttn_FrOperIns_1; Left Insula | -41 | -1 | -7 | 2.97 | 0.004 |
| LH_Vis_4; Left Secondary Visual Cortex | -27 | -95 | -4 | RH_SomMot_8; Right Primary Motor Cortex | 6 | -26 | 70 | 2.96 | 0.004 |
| LH_Vis_9; Left Visual Association Cortex | -6 | -82 | 26 | LH_DorsAttn_Post_4; Left Primary Sensory Cortex | -42 | -34 | 48 | 2.96 | 0.004 |
| LH_SalVentAttn_ParOper_1; Left Supramarginal Gyrus | -59 | -38 | 29 | LH_Default_Par_1; Left Angular Gyrus | -57 | -50 | 12 | 2.96 | 0.004 |
| LH_DorsAttn_Post_4; Left Primary Sensory Cortex | -42 | -34 | 48 | LH_Default_pCunPCC_2; Left Ventral Posterior Cingulate Cortex | -6 | -53 | 33 | 2.96 | 0.004 |

| | | | | | | | | | |
|---|---|---|---|---|---|---|---|---|---|
| LH_Limbic_TempPole_1; Left Temporal Pole | -32 | 2 | -37 | LH_Default_PFC_5; Left Dorsolateral Prefrontal Cortex | -9 | 48 | 41 | 2.95 | 0.004 |
| LH_Vis_4; Left Secondary Visual Cortex | -27 | -95 | -4 | RH_DorsAttn_FEF_1; Right PreMotor & Supplementary Motor Area | 28 | -3 | 59 | 2.93 | 0.004 |
| LH_Vis_9; Left Visual Association Cortex | -6 | -82 | 26 | LH_Default_PFC_4; Left Anterior Prefrontal Cortex | -24 | 61 | -1 | 2.92 | 0.004 |
| LH_DorsAttn_Post_4; Left Primary Sensory Cortex | -42 | -34 | 48 | LH_DorsAttn_Post_5; Left Visual Motor Cortex | -6 | -60 | 56 | 2.91 | 0.004 |
| LH_SalVentAttn_ParOper_1; Left Supramarginal Gyrus | -59 | -38 | 29 | RH_SalVentAttn_FrOperIns_1; Right Insula | 40 | 8 | 1 | 2.89 | 0.004 |
| LH_SomMot_6; Left Primary Motor Cortex | -6 | -29 | 70 | LH_Default_Temp_2; Left Medial Temporal Gyrus | -58 | -32 | -1 | 2.89 | 0.005 |
| LH_Vis_4; Left Secondary Visual Cortex | -27 | -95 | -4 | RH_Limbic_TempPole_1; Right Inferior Temporal Gyrus | 38 | 1 | -38 | 2.88 | 0.005 |
| LH_Vis_9; Left Visual Association Cortex | -6 | -82 | 26 | RH_Limbic_TempPole_1; Right Inferior Temporal Gyrus | 38 | 1 | -38 | 2.88 | 0.005 |
| RH_SalVentAttn_TempOccPar_2; Right Supramarginal Gyrus | 61 | -26 | 27 | RH_Default_Temp_3; Right Superior Temporal Gyrus | 57 | -26 | -2 | 2.88 | 0.005 |
| LH_Default_PFC_1; Left Pars Orbitalis | -35 | 21 | -11 | LH_Default_PFC_3; Left Dorsal Posterior Cingulate Cortex | -6 | 47 | 0 | 2.87 | 0.005 |
| LH_Vis_2; Left Visual Association Cortex | -26 | -77 | -14 | LH_DorsAttn_Post_5; Left Visual Motor Cortex | -6 | -60 | 56 | 2.87 | 0.005 |
| LH_Cont_pCun_1; Left Visual Motor Cortex | -9 | -73 | 38 | RH_Default_Temp_2; Right Temporal Pole | 51 | 7 | -18 | 2.87 | 0.005 |
| RH_DorsAttn_PrCv_1; Right Broca's Area/Operculum | 49 | 10 | 27 | LH_Cont_pCun_1; Left Visual Motor Cortex | -9 | -73 | 38 | 2.86 | 0.005 |
| LH_Vis_2; Left Visual Association Cortex | -26 | -77 | -14 | RH_Limbic_TempPole_1; Right Inferior Temporal Gyrus | 38 | 1 | -38 | 2.86 | 0.005 |
| LH_SomMot_6; Left Primary Motor Cortex | -6 | -29 | 70 | LH_Default_PFC_4; Left Anterior Prefrontal Cortex | -24 | 61 | -1 | 2.85 | 0.005 |
| LH_Vis_4; Left Secondary Visual Cortex | -27 | -95 | -4 | LH_Default_PFC_5; Left Dorsolateral Prefrontal Cortex | -9 | 48 | 41 | 2.84 | 0.005 |
| LH_Vis_1; Left Fusiform Cortex | -26 | -34 | -17 | RH_DorsAttn_PrCv_1; Right Broca's Area/Operculum | 49 | 10 | 27 | 2.83 | 0.005 |
| LH_SalVentAttn_ParOper_1; Left Supramarginal Gyrus | -59 | -38 | 29 | RH_Cont_pCun_1; Right Visual Motor Cortex | 9 | -66 | 43 | 2.82 | 0.005 |
| LH_Vis_3; Left Visual Association Cortex | -17 | -60 | -7 | RH_DorsAttn_FEF_1; Right PreMotor & Supplementary Motor Area | 28 | -3 | 59 | 2.82 | 0.006 |
| LH_Vis_7; Left Visual Association Cortex | -47 | -71 | 11 | RH_SalVentAttn_TempOccPar_2; Right Supramarginal Gyrus | 61 | -26 | 27 | 2.82 | 0.006 |
| RH_DorsAttn_FEF_1; Right PreMotor & Supplementary Motor Area | 28 | -3 | 59 | LH_Limbic_TempPole_2; Left Inferior Temporal Gyrus | -57 | -33 | -21 | 2.81 | 0.006 |
| LH_Vis_7; Left Visual Association Cortex | -47 | -71 | 11 | LH_SomMot_6; Left Primary Motor Cortex | -6 | -29 | 70 | 2.81 | 0.006 |
| LH_Vis_8; Left Visual Association Cortex | -25 | -88 | 20 | RH_DorsAttn_FEF_1; Right PreMotor & Supplementary Motor Area | 28 | -3 | 59 | 2.81 | 0.006 |
| LH_SomMot_1; Left Supramarginal Gyrus | -53 | -23 | 8 | RH_Limbic_TempPole_1; Right Inferior Temporal Gyrus | 38 | 1 | -38 | 2.81 | 0.006 |
| LH_SomMot_5; Left Primary Motor Cortex | -39 | -23 | 59 | LH_Default_pCunPCC_2; Left Ventral Posterior Cingulate Cortex | -6 | -53 | 33 | 2.81 | 0.006 |
| RH_SomMot_8; Right Primary Motor Cortex | 6 | -26 | 70 | LH_Default_Par_1; Left Angular Gyrus | -57 | -50 | 12 | 2.80 | 0.006 |
| LH_Cont_pCun_1; Left Visual Motor Cortex | -9 | -73 | 38 | RH_Default_PFCv_1; Right Pars Orbitalis | 35 | 26 | -15 | 2.80 | 0.006 |
| RH_Vis_5; Right Primary Visual Cortex | 8 | -76 | 5 | LH_DorsAttn_Post_5; Left Visual Motor Cortex | -6 | -60 | 56 | 2.79 | 0.006 |
| LH_Vis_3; Left Visual Association Cortex | -17 | -60 | -7 | LH_SalVentAttn_Med_2; Left Dorsal Posterior Cingulate Cortex | -11 | -34 | 45 | 2.79 | 0.006 |
| LH_Vis_8; Left Visual Association Cortex | -25 | -88 | 20 | LH_Default_PFC_4; Left Anterior Prefrontal Cortex | -24 | 61 | -1 | 2.79 | 0.006 |
| LH_SalVentAttn_ParOper_1; Left Supramarginal Gyrus | -59 | -38 | 29 | RH_SalVentAttn_TempOccPar_1; Right Superior Temporal Gyrus | 58 | -42 | 13 | 2.77 | 0.006 |
| LH_Vis_4; Left Secondary Visual Cortex | -27 | -95 | -4 | LH_DorsAttn_Post_4; Left Primary Sensory Cortex | -42 | -34 | 48 | 2.77 | 0.006 |

| | | | | | | | | | |
|---|---|---|---|---|---|---|---|---|---|
| LH_SomMot_1; Left Supramarginal Gyrus | -53 | -23 | 8 | RH_SomMot_5; Right PreMotor & Supplementary Motor Area | 47 | -11 | 48 | 2.77 | 0.006 |
| LH_Vis_2; Left Visual Association Cortex | -26 | -77 | -14 | LH_SalVentAttn_Med_2; Left Dorsal Posterior Cingulate Cortex | -11 | -34 | 45 | 2.76 | 0.007 |
| LH_Vis_7; Left Visual Association Cortex | -47 | -71 | 11 | RH_SomMot_3; Right Primary Motor Cortex | 57 | -4 | 11 | 2.75 | 0.007 |
| LH_SomMot_2; Left Primary Sensory Cortex | -37 | -21 | 16 | RH_Default_PFCv_1; Right Pars Orbitalis | 35 | 26 | -15 | 2.75 | 0.007 |
| LH_Default_Par_2; Left Angular Gyrus | -48 | -63 | 35 | RH_Default_PFCdPFCm_2; Right Dorsolateral Prefrontal Cortex | 11 | 50 | 39 | 2.75 | 0.007 |
| RH_Vis_3; Right Fusiform Cortex | 49 | -60 | -11 | LH_SomMot_6; Left Primary Motor Cortex | -6 | -29 | 70 | 2.75 | 0.007 |
| RH_Limbic_TempPole_1; Right Inferior Temporal Gyrus | 38 | 1 | -38 | LH_Cont_pCun_1; Left Visual Motor Cortex | -9 | -73 | 38 | 2.74 | 0.007 |
| LH_DorsAttn_Post_1; Left Fusiform Cortex | -47 | -58 | -13 | RH_Limbic_TempPole_1; Right Inferior Temporal Gyrus | 38 | 1 | -38 | 2.74 | 0.007 |
| LH_Vis_6; Left Primary Visual Cortex | -12 | -67 | 7 | LH_SalVentAttn_ParOper_1; Left Supramarginal Gyrus | -59 | -38 | 29 | 2.74 | 0.007 |
| RH_DorsAttn_PrCv_1; Right Broca's Area/Operculum | 49 | 10 | 27 | LH_Default_Par_2; Left Angular Gyrus | -48 | -63 | 35 | 2.73 | 0.007 |
| RH_SomMot_5; Right PreMotor & Supplementary Motor Area | 47 | -11 | 48 | LH_Cont_pCun_1; Left Visual Motor Cortex | -9 | -73 | 38 | 2.73 | 0.007 |
| LH_Vis_8; Left Visual Association Cortex | -25 | -88 | 20 | RH_Cont_pCun_1; Right Visual Motor Cortex | 9 | -66 | 43 | 2.73 | 0.007 |
| RH_Vis_1; Right Fusiform Cortex | 32 | -31 | -22 | LH_SalVentAttn_ParOper_1; Left Supramarginal Gyrus | -59 | -38 | 29 | 2.72 | 0.007 |
| LH_Vis_6; Left Primary Visual Cortex | -12 | -67 | 7 | LH_DorsAttn_Post_5; Left Visual Motor Cortex | -6 | -60 | 56 | 2.72 | 0.007 |
| LH_Vis_3; Left Visual Association Cortex | -17 | -60 | -7 | RH_Cont_PFCl_4; Right Frontal Eye Fields | 43 | 16 | 45 | 2.72 | 0.007 |
| LH_Vis_1; Left Fusiform Cortex | -26 | -34 | -17 | RH_DorsAttn_FEF_1; Right PreMotor & Supplementary Motor Area | 28 | -3 | 59 | 2.72 | 0.008 |
| LH_Vis_2; Left Visual Association Cortex | -26 | -77 | -14 | LH_Default_PFC_7; Left Frontal Eye Fields | -25 | 20 | 51 | 2.71 | 0.008 |
| LH_SomMot_3; Left Primary Sensory Cortex | -54 | -12 | 13 | RH_Default_PFCv_1; Right Pars Orbitalis | 35 | 26 | -15 | 2.71 | 0.008 |
| LH_DorsAttn_Post_5; Left Visual Motor Cortex | -6 | -60 | 56 | LH_Cont_pCun_1; Left Visual Motor Cortex | -9 | -73 | 38 | 2.71 | 0.008 |
| LH_Vis_3; Left Visual Association Cortex | -17 | -60 | -7 | RH_Default_Temp_2; Right Temporal Pole | 51 | 7 | -18 | 2.71 | 0.008 |
| LH_Cont_pCun_1; Left Visual Motor Cortex | -9 | -73 | 38 | RH_Cont_PFCl_4; Right Frontal Eye Fields | 43 | 16 | 45 | 2.71 | 0.008 |
| LH_Vis_4; Left Secondary Visual Cortex | -27 | -95 | -4 | LH_Cont_Par_1; Left Angular Gyrus | -37 | -53 | 46 | 2.70 | 0.008 |
| RH_DorsAttn_FEF_1; Right PreMotor & Supplementary Motor Area | 28 | -3 | 59 | LH_Default_pCunPCC_2; Left Ventral Posterior Cingulate Cortex | -6 | -53 | 33 | 2.70 | 0.008 |
| RH_DorsAttn_PrCv_1; Right Broca's Area/Operculum | 49 | 10 | 27 | LH_Default_pCunPCC_2; Left Ventral Posterior Cingulate Cortex | -6 | -53 | 33 | 2.69 | 0.008 |
| LH_SalVentAttn_Med_2; Left Dorsal Posterior Cingulate Cortex | -11 | -34 | 45 | LH_Default_pCunPCC_2; Left Ventral Posterior Cingulate Cortex | -6 | -53 | 33 | 2.68 | 0.008 |
| LH_DorsAttn_PrCv_1; Left PreMotor & Supplementary Motor Area | -48 | 6 | 28 | RH_Default_Temp_1; Right Medial Temporal Gyrus | 62 | -23 | -19 | 2.68 | 0.008 |
| LH_Vis_7; Left Visual Association Cortex | -47 | -71 | 11 | RH_DorsAttn_Post_5; Right Visual Motor Cortex | 14 | -52 | 66 | 2.67 | 0.008 |
| LH_SomMot_1; Left Supramarginal Gyrus | -53 | -23 | 8 | RH_Default_PFCv_2; Right Broca's Area/ Pars Triangularis | 51 | 28 | 0 | 2.67 | 0.008 |
| RH_DorsAttn_FEF_1; Right PreMotor & Supplementary Motor Area | 28 | -3 | 59 | LH_Default_pCunPCC_1; Left Ventral Posterior Cingulate Cortex | -11 | -56 | 13 | 2.66 | 0.009 |
| LH_SalVentAttn_ParOper_1; Left Supramarginal Gyrus | -59 | -38 | 29 | RH_Cont_PFCl_4; Right Frontal Eye Fields | 43 | 16 | 45 | 2.66 | 0.009 |
| RH_SomMot_5; Right PreMotor & Supplementary Motor Area | 47 | -11 | 48 | LH_DorsAttn_Post_2; Left Supramarginal Gyrus | -57 | -25 | 39 | 2.66 | 0.009 |
| LH_Default_Par_1; Left Angular Gyrus | -57 | -50 | 12 | RH_Default_PFCv_1; Right Pars Orbitalis | 35 | 26 | -15 | 2.66 | 0.009 |

| | | | | | | | | | |
|---|---|---|---|---|---|---|---|---|---|
| RH_SalVentAttn_TempOccPar_2; Right Supramarginal Gyrus | 61 | -26 | 27 | RH_Default_PFCv_2; Right Broca's Area/ Pars Triangularis | 51 | 28 | 0 | 2.65 | 0.009 |
| RH_SomMot_3; Right Primary Motor Cortex | 57 | -4 | 11 | LH_SalVentAttn_ParOper_1; Left Supramarginal Gyrus | -59 | -38 | 29 | 2.65 | 0.009 |
| RH_DorsAttn_FEF_1; Right PreMotor & Supplementary Motor Area | 28 | -3 | 59 | LH_Cont_Cing_1; Left Ventral Posterior Cingulate Cortex | -4 | -26 | 33 | 2.65 | 0.009 |
| LH_Vis_8; Left Visual Association Cortex | -25 | -88 | 20 | RH_Cont_PFCl_4; Right Frontal Eye Fields | 43 | 16 | 45 | 2.65 | 0.009 |
| LH_Vis_8; Left Visual Association Cortex | -25 | -88 | 20 | RH_Limbic_OFC_1; Right Orbitofrontal Cortex | 12 | 35 | -20 | 2.63 | 0.009 |
| LH_SalVentAttn_ParOper_1; Left Supramarginal Gyrus | -59 | -38 | 29 | LH_Limbic_TempPole_1; Left Temporal Pole | -32 | 2 | -37 | 2.63 | 0.010 |
| LH_Vis_9; Left Visual Association Cortex | -6 | -82 | 26 | LH_SomMot_6; Left Primary Motor Cortex | -6 | -29 | 70 | 2.62 | 0.010 |
| LH_DorsAttn_Post_4; Left Primary Sensory Cortex | -42 | -34 | 48 | RH_Cont_pCun_1; Right Visual Motor Cortex | 9 | -66 | 43 | 2.62 | 0.010 |
| LH_DorsAttn_Post_1; Left Fusiform Cortex | -47 | -58 | -13 | RH_Limbic_OFC_1; Right Orbitofrontal Cortex | 12 | 35 | -20 | 2.62 | 0.010 |
| LH_Vis_7; Left Visual Association Cortex | -47 | -71 | 11 | RH_SomMot_4; Right Primary Motor Cortex | 58 | -5 | 31 | 2.62 | 0.010 |
| LH_SomMot_1; Left Supramarginal Gyrus | -53 | -23 | 8 | RH_SomMot_8; Right Primary Motor Cortex | 6 | -26 | 70 | 2.61 | 0.010 |
| LH_Vis_5; Left Secondary Visual Cortex | -5 | -92 | -2 | LH_DorsAttn_Post_5; Left Visual Motor Cortex | -6 | -60 | 56 | 2.61 | 0.010 |
| LH_SalVentAttn_ParOper_1; Left Supramarginal Gyrus | -59 | -38 | 29 | LH_Default_pCunPCC_1; Left Ventral Posterior Cingulate Cortex | -11 | -56 | 13 | 2.61 | 0.010 |
| LH_Vis_8; Left Visual Association Cortex | -25 | -88 | 20 | LH_Default_PFC_7; Left Frontal Eye Fields | -25 | 20 | 51 | 2.60 | 0.010 |
| RH_Vis_8; Right Visual Association Cortex | 13 | -86 | 29 | LH_SalVentAttn_ParOper_1; Left Supramarginal Gyrus | -59 | -38 | 29 | 2.60 | 0.010 |
| RH_SomMot_1; Right Primary Auditory Cortex | 53 | -16 | 7 | LH_SalVentAttn_ParOper_1; Left Supramarginal Gyrus | -59 | -38 | 29 | 2.60 | 0.010 |

**Eyes-Open Beta**

| **ROI1 Labels (Schaefer; Talairach)** | **X** | **Y** | **Z** | **ROI2 Labels (Schaefer; Talairach)** | **X** | **Y** | **Z** | **t** | **p (unc)** |
|---|---|---|---|---|---|---|---|---|---|
| RH_SomMot_8; Right Primary Motor Cortex | 6 | -26 | 70 | LH_Cont_Par_1; Left Angular Gyrus | -37 | -53 | 46 | 4.20 | 5.01E-05 |
| RH_SomMot_8; Right Primary Motor Cortex | 6 | -26 | 70 | LH_DorsAttn_Post_3; Left Visual Motor Cortex | -24 | -68 | 49 | 3.98 | 0.0001 |
| RH_Cont_PFCl_1; Right Anterior Prefrontal Cortex | 30 | 58 | -3 | RH_Cont_PFCmp_1; Right Frontal Eye Fields | 6 | 28 | 30 | 3.92 | 0.0001 |
| RH_SomMot_8; Right Primary Motor Cortex | 6 | -26 | 70 | LH_Default_PFC_4; Left Anterior Prefrontal Cortex | -24 | 61 | -1 | 3.84 | 0.0002 |
| RH_SomMot_8; Right Primary Motor Cortex | 6 | -26 | 70 | LH_DorsAttn_Post_5; Left Visual Motor Cortex | -6 | -60 | 56 | 3.56 | 0.0005 |
| RH_Default_PFCdPFCm_2; Right Dorsolateral Prefrontal Cortex | 11 | 50 | 39 | RH_Default_PFCdPFCm_3; Right Frontal Eye Fields | 26 | 24 | 50 | 3.52 | 0.0006 |
| LH_SomMot_6; Left Primary Motor Cortex | -6 | -29 | 70 | LH_DorsAttn_Post_3; Left Visual Motor Cortex | -24 | -68 | 49 | 3.45 | 0.0007 |
| LH_SomMot_6; Left Primary Motor Cortex | -6 | -29 | 70 | RH_Default_Temp_2; Right Temporal Pole | 51 | 7 | -18 | 3.40 | 0.0009 |
| RH_Vis_8; Right Visual Association Cortex | 13 | -86 | 29 | RH_Cont_PFCl_3; Right Dorsolateral Prefrontal Cortex | 32 | 46 | 29 | 3.39 | 0.0009 |
| RH_Cont_PFCmp_1; Right Frontal Eye Fields | 6 | 28 | 30 | RH_Default_PFCdPFCm_3; Right Frontal Eye Fields | 26 | 24 | 50 | 3.39 | 0.0009 |
| RH_SomMot_8; Right Primary Motor Cortex | 6 | -26 | 70 | LH_Default_pCunPCC_1; Left Ventral Posterior Cingulate Cortex | -11 | -56 | 13 | 3.38 | 0.0009 |
| RH_SomMot_8; Right Primary Motor Cortex | 6 | -26 | 70 | RH_Default_Temp_2; Right Temporal Pole | 51 | 7 | -18 | 3.37 | 0.0010 |
| LH_SomMot_6; Left Primary Motor Cortex | -6 | -29 | 70 | LH_Cont_Par_1; Left Angular Gyrus | -37 | -53 | 46 | 3.36 | 0.001 |

| | | | | | | | | | |
|---|---|---|---|---|---|---|---|---|---|
| RH_SomMot_8; Right Primary Motor Cortex | 6 | -26 | 70 | LH_DorsAttn_Post_6; Left Visual Motor Cortex | -22 | -51 | 66 | 3.27 | 0.001 |
| LH_Default_PFC_7; Left Frontal Eye Fields | -25 | 20 | 51 | RH_Default_PFCv_2; Right Broca's Area/ Pars Triangularis | 51 | 28 | 0 | 3.24 | 0.002 |
| LH_SomMot_2; Left Primary Sensory Cortex | -37 | -21 | 16 | RH_DorsAttn_FEF_1; Right PreMotor & Supplementary Motor Area | 28 | -3 | 59 | 3.23 | 0.002 |
| LH_Vis_7; Left Visual Association Cortex | -47 | -71 | 11 | LH_SomMot_2; Left Primary Sensory Cortex | -37 | -21 | 16 | 3.18 | 0.002 |
| RH_Cont_PFCl_1; Right Anterior Prefrontal Cortex | 30 | 58 | -3 | RH_Default_PFCdPFCm_2; Right Dorsolateral Prefrontal Cortex | 11 | 50 | 39 | 3.17 | 0.002 |
| RH_SomMot_8; Right Primary Motor Cortex | 6 | -26 | 70 | RH_Default_PFCv_2; Right Broca's Area/ Pars Triangularis | 51 | 28 | 0 | 3.12 | 0.002 |
| LH_DorsAttn_Post_4; Left Primary Sensory Cortex | -42 | -34 | 48 | LH_DorsAttn_Post_6; Left Visual Motor Cortex | -22 | -51 | 66 | 3.12 | 0.002 |
| LH_Vis_9; Left Visual Association Cortex | -6 | -82 | 26 | LH_Default_pCunPCC_1; Left Ventral Posterior Cingulate Cortex | -11 | -56 | 13 | 3.09 | 0.002 |
| RH_Cont_PFCl_3; Right Dorsolateral Prefrontal Cortex | 32 | 46 | 29 | RH_Cont_PFCmp_1; Right Frontal Eye Fields | 6 | 28 | 30 | 3.04 | 0.003 |
| LH_SomMot_6; Left Primary Motor Cortex | -6 | -29 | 70 | LH_DorsAttn_Post_5; Left Visual Motor Cortex | -6 | -60 | 56 | 3.03 | 0.003 |
| RH_DorsAttn_FEF_1; Right PreMotor & Supplementary Motor Area | 28 | -3 | 59 | LH_SalVentAttn_Med_1; Left Dorsal Anterior Cingulate Cortex | -5 | 20 | 34 | 3.03 | 0.003 |
| LH_DorsAttn_Post_2; Left Supramarginal Gyrus | -57 | -25 | 39 | LH_Default_PFC_7; Left Frontal Eye Fields | -25 | 20 | 51 | 3.03 | 0.003 |
| LH_SomMot_1; Left Supramarginal Gyrus | -53 | -23 | 8 | RH_SomMot_2; Right Insula | 40 | -15 | 15 | 3.01 | 0.003 |
| RH_DorsAttn_FEF_1; Right PreMotor & Supplementary Motor Area | 28 | -3 | 59 | LH_Cont_Cing_1; Left Ventral Posterior Cingulate Cortex | -4 | -26 | 33 | 3.01 | 0.003 |
| LH_DorsAttn_Post_3; Left Visual Motor Cortex | -24 | -68 | 49 | RH_Default_PFCdPFCm_2; Right Dorsolateral Prefrontal Cortex | 11 | 50 | 39 | 3.01 | 0.003 |
| LH_SomMot_6; Left Primary Motor Cortex | -6 | -29 | 70 | LH_Default_PFC_4; Left Anterior Prefrontal Cortex | -24 | 61 | -1 | 2.95 | 0.004 |
| LH_SomMot_6; Left Primary Motor Cortex | -6 | -29 | 70 | LH_DorsAttn_Post_6; Left Visual Motor Cortex | -22 | -51 | 66 | 2.95 | 0.004 |
| RH_Vis_3; Right Fusiform Cortex | 49 | -60 | -11 | RH_Vis_8; Right Visual Association Cortex | 13 | -86 | 29 | 2.94 | 0.004 |
| RH_Vis_3; Right Fusiform Cortex | 49 | -60 | -11 | LH_Default_Temp_2; Left Medial Temporal Gyrus | -58 | -32 | -1 | 2.92 | 0.004 |
| RH_DorsAttn_FEF_1; Right PreMotor & Supplementary Motor Area | 28 | -3 | 59 | RH_Cont_Cing_1; Right Ventral Posterior Cingulate Cortex | 5 | -27 | 33 | 2.92 | 0.004 |
| LH_Vis_6; Left Primary Visual Cortex | -12 | -67 | 7 | RH_SomMot_8; Right Primary Motor Cortex | 6 | -26 | 70 | 2.91 | 0.004 |
| RH_DorsAttn_FEF_1; Right PreMotor & Supplementary Motor Area | 28 | -3 | 59 | LH_Default_PFC_1; Left Pars Orbitalis | -35 | 21 | -11 | 2.90 | 0.004 |
| LH_DorsAttn_Post_4; Left Primary Sensory Cortex | -42 | -34 | 48 | LH_SalVentAttn_Med_2; Left Dorsal Posterior Cingulate Cortex | -11 | -34 | 45 | 2.88 | 0.005 |
| RH_DorsAttn_FEF_1; Right PreMotor & Supplementary Motor Area | 28 | -3 | 59 | RH_Cont_PFCmp_1; Right Frontal Eye Fields | 6 | 28 | 30 | 2.88 | 0.005 |
| LH_Vis_7; Left Visual Association Cortex | -47 | -71 | 11 | LH_DorsAttn_Post_6; Left Visual Motor Cortex | -22 | -51 | 66 | 2.88 | 0.005 |
| LH_Vis_4; Left Secondary Visual Cortex | -27 | -95 | -4 | RH_Vis_3; Right Fusiform Cortex | 49 | -60 | -11 | 2.88 | 0.005 |
| LH_Vis_2; Left Visual Association Cortex | -26 | -77 | -14 | RH_DorsAttn_Post_2; Right Primary Sensory Cortex | 50 | -24 | 42 | 2.85 | 0.005 |
| RH_SomMot_8; Right Primary Motor Cortex | 6 | -26 | 70 | LH_Cont_pCun_1; Left Visual Motor Cortex | -9 | -73 | 38 | 2.84 | 0.005 |
| LH_DorsAttn_Post_4; Left Primary Sensory Cortex | -42 | -34 | 48 | RH_DorsAttn_FEF_1; Right PreMotor & Supplementary Motor Area | 28 | -3 | 59 | 2.83 | 0.005 |
| LH_Vis_2; Left Visual Association Cortex | -26 | -77 | -14 | RH_SalVentAttn_TempOccPar_2; Right Supramarginal Gyrus | 61 | -26 | 27 | 2.82 | 0.005 |
| LH_Vis_1; Left Fusiform Cortex | -26 | -34 | -17 | LH_Cont_pCun_1; Left Visual Motor Cortex | -9 | -73 | 38 | 2.82 | 0.006 |
| LH_Vis_7; Left Visual Association Cortex | -47 | -71 | 11 | LH_DorsAttn_Post_4; Left Primary Sensory Cortex | -42 | -34 | 48 | 2.82 | 0.006 |

| | | | | | | | | | |
|---|---|---|---|---|---|---|---|---|---|
| RH_DorsAttn_Post_2; Right Primary Sensory Cortex | 50 | -24 | 42 | RH_Default_Par_1; Right Angular Gyrus | 55 | -51 | 31 | 2.81 | 0.006 |
| RH_Cont_PFCl_2; Right Dorsolateral Prefrontal Cortex | 45 | 39 | 15 | RH_Cont_PFCmp_1; Right Frontal Eye Fields | 6 | 28 | 30 | 2.81 | 0.006 |
| LH_SomMot_1; Left Supramarginal Gyrus | -53 | -23 | 8 | RH_SomMot_3; Right Primary Motor Cortex | 57 | -4 | 11 | 2.79 | 0.006 |
| LH_DorsAttn_Post_3; Left Visual Motor Cortex | -24 | -68 | 49 | RH_SalVentAttn_Med_2; Right PreMotor & Supplementary Motor Area | 7 | 6 | 52 | 2.79 | 0.006 |
| LH_SalVentAttn_ParOper_1; Left Supramarginal Gyrus | -59 | -38 | 29 | RH_Cont_PFCl_3; Right Dorsolateral Prefrontal Cortex | 32 | 46 | 29 | -2.78 | 0.006 |
| LH_DorsAttn_FEF_1; Left PreMotor & Supplementary Motor Area | -26 | -3 | 59 | RH_SalVentAttn_TempOccPar_2; Right Supramarginal Gyrus | 61 | -26 | 27 | -2.76 | 0.007 |
| RH_DorsAttn_Post_2; Right Primary Sensory Cortex | 50 | -24 | 42 | RH_Cont_Par_1; Right Supramarginal Gyrus | 57 | -39 | 44 | 2.76 | 0.007 |
| RH_SalVentAttn_Med_1; Right Dorsal Posterior Cingulate Cortex | 11 | -31 | 45 | RH_Cont_Cing_1; Right Ventral Posterior Cingulate Cortex | 5 | -27 | 33 | 2.76 | 0.007 |
| LH_DorsAttn_FEF_1; Left PreMotor & Supplementary Motor Area | -26 | -3 | 59 | LH_Default_PFC_6; Left PreMotor & Supplementary Motor Area | -41 | 14 | 48 | 2.75 | 0.007 |
| LH_SomMot_6; Left Primary Motor Cortex | -6 | -29 | 70 | RH_SomMot_8; Right Primary Motor Cortex | 6 | -26 | 70 | 2.75 | 0.007 |
| LH_DorsAttn_Post_4; Left Primary Sensory Cortex | -42 | -34 | 48 | RH_SalVentAttn_Med_1; Right Dorsal Posterior Cingulate Cortex | 11 | -31 | 45 | 2.75 | 0.007 |
| LH_Cont_pCun_1; Left Visual Motor Cortex | -9 | -73 | 38 | RH_Default_PFCdPFCm_2; Right Dorsolateral Prefrontal Cortex | 11 | 50 | 39 | 2.74 | 0.007 |
| RH_DorsAttn_PrCv_1; Right Broca's Area/Operculum | 49 | 10 | 27 | RH_Default_PFCdPFCm_2; Right Dorsolateral Prefrontal Cortex | 11 | 50 | 39 | 2.73 | 0.007 |
| LH_DorsAttn_Post_5; Left Visual Motor Cortex | -6 | -60 | 56 | RH_DorsAttn_FEF_1; Right PreMotor & Supplementary Motor Area | 28 | -3 | 59 | 2.72 | 0.007 |
| RH_SomMot_8; Right Primary Motor Cortex | 6 | -26 | 70 | RH_SalVentAttn_TempOccPar_1; Right Superior Temporal Gyrus | 58 | -42 | 13 | 2.71 | 0.008 |
| LH_SomMot_4; Left PreMotor & Supplementary Motor Area | -55 | -8 | 34 | LH_SalVentAttn_Med_1; Left Dorsal Anterior Cingulate Cortex | -5 | 20 | 34 | 2.70 | 0.008 |
| LH_Vis_3; Left Visual Association Cortex | -17 | -60 | -7 | RH_SalVentAttn_Med_2; Right PreMotor & Supplementary Motor Area | 7 | 6 | 52 | 2.70 | 0.008 |
| RH_Vis_8; Right Visual Association Cortex | 13 | -86 | 29 | RH_SomMot_8; Right Primary Motor Cortex | 6 | -26 | 70 | 2.70 | 0.008 |
| LH_DorsAttn_Post_3; Left Visual Motor Cortex | -24 | -68 | 49 | LH_SalVentAttn_Med_3; Left PreMotor & Supplementary Motor Area | -6 | 4 | 62 | 2.69 | 0.008 |
| RH_SomMot_8; Right Primary Motor Cortex | 6 | -26 | 70 | LH_Default_Temp_1; Left Medial Temporal Gyrus | -55 | -4 | -20 | 2.69 | 0.008 |
| LH_DorsAttn_Post_6; Left Visual Motor Cortex | -22 | -51 | 66 | LH_Cont_Par_1; Left Angular Gyrus | -37 | -53 | 46 | 2.68 | 0.008 |
| LH_DorsAttn_Post_4; Left Primary Sensory Cortex | -42 | -34 | 48 | RH_Default_pCunPCC_2; Right Ventral Posterior Cingulate Cortex | 7 | -52 | 31 | 2.67 | 0.009 |
| RH_Vis_8; Right Visual Association Cortex | 13 | -86 | 29 | LH_DorsAttn_Post_1; Left Fusiform Cortex | -47 | -58 | -13 | 2.66 | 0.009 |
| LH_SomMot_4; Left PreMotor & Supplementary Motor Area | -55 | -8 | 34 | LH_Default_PFC_7; Left Frontal Eye Fields | -25 | 20 | 51 | 2.66 | 0.009 |
| LH_SomMot_4; Left PreMotor & Supplementary Motor Area | -55 | -8 | 34 | RH_Cont_Cing_1; Right Ventral Posterior Cingulate Cortex | 5 | -27 | 33 | 2.65 | 0.009 |
| RH_Vis_7; Right Visual Association Cortex | 36 | -82 | 16 | RH_Vis_8; Right Visual Association Cortex | 13 | -86 | 29 | 2.65 | 0.009 |
| RH_Cont_PFCl_4; Right Frontal Eye Fields | 43 | 16 | 45 | RH_Default_PFCdPFCm_2; Right Dorsolateral Prefrontal Cortex | 11 | 50 | 39 | 2.64 | 0.009 |
| LH_Vis_7; Left Visual Association Cortex | -47 | -71 | 11 | RH_DorsAttn_Post_1; Right Visual Association Cortex | 50 | -62 | 16 | 2.64 | 0.009 |
| LH_SalVentAttn_ParOper_1; Left Supramarginal Gyrus | -59 | -38 | 29 | LH_Cont_PFCl_1; Left Dorsolateral Prefrontal Cortex | -43 | 33 | 21 | 2.64 | 0.009 |
| RH_Cont_PFCmp_1; Right Frontal Eye Fields | 6 | 28 | 30 | RH_Default_PFCdPFCm_1; Right Anterior Prefrontal Cortex | 7 | 48 | 1 | 2.63 | 0.010 |
| LH_Vis_7; Left Visual Association Cortex | -47 | -71 | 11 | RH_DorsAttn_FEF_1; Right PreMotor & Supplementary Motor Area | 28 | -3 | 59 | 2.63 | 0.010 |
| LH_Vis_8; Left Visual Association Cortex | -25 | -88 | 20 | RH_SalVentAttn_Med_2; Right PreMotor & Supplementary Motor Area | 7 | 6 | 52 | 2.63 | 0.010 |

| | | | | | | | | | |
|---|---|---|---|---|---|---|---|---|---|
| RH_SomMot_3; Right Primary Motor Cortex | 57 | -4 | 11 | LH_Default_Par_2; Left Angular Gyrus | -48 | -63 | 35 | 2.63 | 0.010 |
| LH_SalVentAttn_Med_1; Left Dorsal Anterior Cingulate Cortex | -5 | 20 | 34 | RH_Cont_PFCl_1; Right Anterior Prefrontal Cortex | 30 | 58 | -3 | 2.62 | 0.010 |
| RH_Vis_3; Right Fusiform Cortex | 49 | -60 | -11 | LH_Default_Par_1; Left Angular Gyrus | -57 | -50 | 12 | 2.62 | 0.010 |
| LH_SomMot_6; Left Primary Motor Cortex | -6 | -29 | 70 | LH_DorsAttn_Post_1; Left Fusiform Cortex | -47 | -58 | -13 | 2.60 | 0.010 |
| **Eyes-Open Gamma** | | | | | | | | | |
| **ROI1 Labels (Schaefer; Talairach)** | **X** | **Y** | **Z** | **ROI2 Labels (Schaefer; Talairach)** | **X** | **Y** | **Z** | **t** | **p (unc)** |
| RH_Cont_Par_1; Right Supramarginal Gyrus | 57 | -39 | 44 | RH_Cont_PFCl_1; Right Anterior Prefrontal Cortex | 30 | 58 | -3 | 3.90 | 0.0002 |
| RH_Cont_pCun_1; Right Visual Motor Cortex | 9 | -66 | 43 | RH_Default_Temp_2; Right Temporal Pole | 51 | 7 | -18 | 3.57 | 0.0005 |
| RH_Cont_Par_1; Right Supramarginal Gyrus | 57 | -39 | 44 | LH_Default_PFC_4; Left Anterior Prefrontal Cortex | -24 | 61 | -1 | 3.43 | 0.0008 |
| RH_Cont_PFCl_1; Right Anterior Prefrontal Cortex | 30 | 58 | -3 | RH_Default_Par_1; Right Angular Gyrus | 55 | -51 | 31 | 3.42 | 0.0008 |
| LH_SomMot_4; Left PreMotor & Supplementary Motor Area | -55 | -8 | 34 | LH_Default_Temp_1; Left Medial Temporal Gyrus | -55 | -4 | -20 | 3.24 | 0.002 |
| RH_SomMot_4; Right Primary Motor Cortex | 58 | -5 | 31 | LH_SalVentAttn_Med_1; Left Dorsal Anterior Cingulate Cortex | -5 | 20 | 34 | 3.21 | 0.002 |
| LH_SomMot_1; Left Supramarginal Gyrus | -53 | -23 | 8 | RH_SalVentAttn_TempOccPar_2; Right Supramarginal Gyrus | 61 | -26 | 27 | 3.19 | 0.002 |
| LH_SalVentAttn_PFCl_1; Left Anterior Prefrontal Cortex | -30 | 44 | 30 | LH_Limbic_OFC_1; Left Orbitofrontal Cortex | -14 | 32 | -20 | -3.15 | 0.002 |
| LH_SomMot_2; Left Primary Sensory Cortex | -37 | -21 | 16 | RH_SalVentAttn_TempOccPar_2; Right Supramarginal Gyrus | 61 | -26 | 27 | 3.13 | 0.002 |
| RH_DorsAttn_Post_5; Right Visual Motor Cortex | 14 | -52 | 66 | RH_SalVentAttn_Med_1; Right Dorsal Posterior Cingulate Cortex | 11 | -31 | 45 | -3.10 | 0.002 |
| RH_SalVentAttn_TempOccPar_2; Right Supramarginal Gyrus | 61 | -26 | 27 | LH_Default_Temp_2; Left Medial Temporal Gyrus | -58 | -32 | -1 | 3.09 | 0.002 |
| LH_DorsAttn_PrCv_1; Left PreMotor & Supplementary Motor Area | -48 | 6 | 28 | LH_Default_PFC_5; Left Dorsolateral Prefrontal Cortex | -9 | 48 | 41 | 3.08 | 0.003 |
| LH_SalVentAttn_PFCl_1; Left Anterior Prefrontal Cortex | -30 | 44 | 30 | RH_Cont_PFCl_1; Right Anterior Prefrontal Cortex | 30 | 58 | -3 | -3.08 | 0.003 |
| LH_Cont_Cing_1; Left Ventral Posterior Cingulate Cortex | -4 | -26 | 33 | RH_Cont_PFCl_4; Right Frontal Eye Fields | 43 | 16 | 45 | 3.07 | 0.003 |
| LH_Default_PFC_4; Left Anterior Prefrontal Cortex | -24 | 61 | -1 | RH_Default_Par_1; Right Angular Gyrus | 55 | -51 | 31 | 3.06 | 0.003 |
| RH_DorsAttn_Post_1; Right Visual Association Cortex | 50 | -62 | 16 | RH_Default_Temp_2; Right Temporal Pole | 51 | 7 | -18 | 3.03 | 0.003 |
| LH_DorsAttn_FEF_1; Left PreMotor & Supplementary Motor Area | -26 | -3 | 59 | LH_Limbic_TempPole_2; Left Inferior Temporal Gyrus | -57 | -33 | -21 | 3.03 | 0.003 |
| RH_Vis_8; Right Visual Association Cortex | 13 | -86 | 29 | LH_SalVentAttn_ParOper_1; Left Supramarginal Gyrus | -59 | -38 | 29 | 3.03 | 0.003 |
| LH_SomMot_3; Left Primary Sensory Cortex | -54 | -12 | 13 | RH_Cont_Par_1; Right Supramarginal Gyrus | 57 | -39 | 44 | 3.02 | 0.003 |
| LH_SomMot_3; Left Primary Sensory Cortex | -54 | -12 | 13 | RH_DorsAttn_Post_2; Right Primary Sensory Cortex | 50 | -24 | 42 | 2.99 | 0.003 |
| RH_Cont_Par_1; Right Supramarginal Gyrus | 57 | -39 | 44 | LH_Default_PFC_5; Left Dorsolateral Prefrontal Cortex | -9 | 48 | 41 | 2.98 | 0.003 |
| RH_DorsAttn_PrCv_1; Right Broca's Area/Operculum | 49 | 10 | 27 | LH_Default_Temp_2; Left Medial Temporal Gyrus | -58 | -32 | -1 | 2.97 | 0.004 |
| RH_Cont_Par_2; Right Angular Gyrus | 45 | -63 | 46 | RH_Default_PFCv_1; Right Pars Orbitalis | 35 | 26 | -15 | 2.96 | 0.004 |
| RH_Default_Par_1; Right Angular Gyrus | 55 | -51 | 31 | RH_Default_PFCv_1; Right Pars Orbitalis | 35 | 26 | -15 | 2.95 | 0.004 |
| RH_Vis_8; Right Visual Association Cortex | 13 | -86 | 29 | LH_DorsAttn_Post_2; Left Supramarginal Gyrus | -57 | -25 | 39 | 2.95 | 0.004 |
| RH_SalVentAttn_TempOccPar_1; Right Superior Temporal Gyrus | 58 | -42 | 13 | RH_Cont_Par_2; Right Angular Gyrus | 45 | -63 | 46 | 2.91 | 0.004 |

| | | | | | | | | | |
|---|---|---|---|---|---|---|---|---|---|
| LH_SalVentAttn_PFCl_1; Left Anterior Prefrontal Cortex | -30 | 44 | 30 | RH_Cont_Par_1; Right Supramarginal Gyrus | 57 | -39 | 44 | 2.91 | 0.004 |
| RH_DorsAttn_Post_2; Right Primary Sensory Cortex | 50 | -24 | 42 | RH_SalVentAttn_FrOperIns_1; Right Insula | 40 | 8 | 1 | 2.89 | 0.004 |
| RH_Cont_pCun_1; Right Visual Motor Cortex | 9 | -66 | 43 | RH_Default_PFCv_2; Right Broca's Area/ Pars Triangularis | 51 | 28 | 0 | 2.87 | 0.005 |
| LH_DorsAttn_Post_2; Left Supramarginal Gyrus | -57 | -25 | 39 | LH_Cont_Par_1; Left Angular Gyrus | -37 | -53 | 46 | 2.87 | 0.005 |
| RH_DorsAttn_Post_2; Right Primary Sensory Cortex | 50 | -24 | 42 | LH_Default_Temp_2; Left Medial Temporal Gyrus | -58 | -32 | -1 | 2.85 | 0.005 |
| LH_SalVentAttn_Med_2; Left Dorsal Posterior Cingulate Cortex | -11 | -34 | 45 | RH_Cont_PFCl_4; Right Frontal Eye Fields | 43 | 16 | 45 | 2.83 | 0.005 |
| RH_SalVentAttn_TempOccPar_1; Right Superior Temporal Gyrus | 58 | -42 | 13 | RH_Default_Par_1; Right Angular Gyrus | 55 | -51 | 31 | 2.82 | 0.006 |
| RH_Vis_7; Right Visual Association Cortex | 36 | -82 | 16 | RH_Default_Temp_2; Right Temporal Pole | 51 | 7 | -18 | 2.78 | 0.006 |
| RH_Default_Par_1; Right Angular Gyrus | 55 | -51 | 31 | RH_Default_Temp_2; Right Temporal Pole | 51 | 7 | -18 | 2.77 | 0.006 |
| RH_DorsAttn_Post_1; Right Visual Association Cortex | 50 | -62 | 16 | RH_Limbic_OFC_1; Right Orbitofrontal Cortex | 12 | 35 | -20 | 2.76 | 0.007 |
| LH_DorsAttn_Post_2; Left Supramarginal Gyrus | -57 | -25 | 39 | LH_Default_pCunPCC_2; Left Ventral Posterior Cingulate Cortex | -6 | -53 | 33 | 2.73 | 0.007 |
| LH_Vis_5; Left Secondary Visual Cortex | -5 | -92 | -2 | LH_SalVentAttn_ParOper_1; Left Supramarginal Gyrus | -59 | -38 | 29 | 2.73 | 0.007 |
| RH_SalVentAttn_FrOperIns_1; Right Insula | 40 | 8 | 1 | RH_Cont_Par_1; Right Supramarginal Gyrus | 57 | -39 | 44 | 2.72 | 0.007 |
| RH_SomMot_2; Right Insula | 40 | -15 | 15 | LH_DorsAttn_PrCv_1; Left PreMotor & Supplementary Motor Area | -48 | 6 | 28 | 2.69 | 0.008 |
| RH_Vis_2; Right Fusiform Cortex | 27 | -66 | -12 | RH_Default_Temp_1; Right Medial Temporal Gyrus | 62 | -23 | -19 | 2.67 | 0.009 |
| LH_Vis_4; Left Secondary Visual Cortex | -27 | -95 | -4 | LH_SomMot_1; Left Supramarginal Gyrus | -53 | -23 | 8 | 2.65 | 0.009 |
| RH_SomMot_4; Right Primary Motor Cortex | 58 | -5 | 31 | LH_Default_Temp_2; Left Medial Temporal Gyrus | -58 | -32 | -1 | 2.64 | 0.009 |
| LH_Limbic_OFC_1; Left Orbitofrontal Cortex | -14 | 32 | -20 | RH_Cont_Cing_1; Right Ventral Posterior Cingulate Cortex | 5 | -27 | 33 | 2.64 | 0.009 |
| LH_SomMot_1; Left Supramarginal Gyrus | -53 | -23 | 8 | RH_SomMot_2; Right Insula | 40 | -15 | 15 | 2.63 | 0.010 |
| LH_SalVentAttn_PFCl_1; Left Anterior Prefrontal Cortex | -30 | 44 | 30 | RH_Default_Par_1; Right Angular Gyrus | 55 | -51 | 31 | 2.63 | 0.010 |
| RH_DorsAttn_Post_2; Right Primary Sensory Cortex | 50 | -24 | 42 | RH_Default_PFCv_1; Right Pars Orbitalis | 35 | 26 | -15 | 2.62 | 0.010 |
| LH_SalVentAttn_ParOper_1; Left Supramarginal Gyrus | -59 | -38 | 29 | RH_Default_pCunPCC_1; Right Ventral Posterior Cingulate Cortex | 12 | -54 | 14 | 2.62 | 0.010 |
| LH_SomMot_3; Left Primary Sensory Cortex | -54 | -12 | 13 | RH_DorsAttn_Post_3; Right Visual Motor Cortex | 38 | -45 | 49 | 2.61 | 0.010 |
| RH_SalVentAttn_Med_1; Right Dorsal Posterior Cingulate Cortex | 11 | -31 | 45 | LH_Default_PFC_6; Left PreMotor & Supplementary Motor Area | -41 | 14 | 48 | 2.60 | 0.010 |
| LH_SomMot_5; Left Primary Motor Cortex | -39 | -23 | 59 | RH_DorsAttn_FEF_1; Right PreMotor & Supplementary Motor Area | 28 | -3 | 59 | 2.60 | 0.010 |
| **Eyes-Closed Theta** | | | | | | | | | |
| **ROI1 Labels (Schaefer; Talairach)** | **X** | **Y** | **Z** | **ROI2 Labels (Schaefer; Talairach)** | **X** | **Y** | **Z** | **t** | **p (unc)** |
| LH_Limbic_TempPole_2; Left Inferior Temporal Gyrus | -57 | -33 | -21 | RH_Limbic_TempPole_1; Right Inferior Temporal Gyrus | 38 | 1 | -38 | 3.40 | 0.0009 |
| RH_SomMot_1; Right Primary Auditory Cortex | 53 | -16 | 7 | LH_Limbic_TempPole_1; Left Temporal Pole | -32 | 2 | -37 | 3.14 | 0.002 |
| RH_SomMot_2; Right Insula | 40 | -15 | 15 | LH_Limbic_TempPole_1; Left Temporal Pole | -32 | 2 | -37 | 2.96 | 0.004 |
| RH_Default_Temp_1; Right Medial Temporal Gyrus | 62 | -23 | -19 | RH_Default_pCunPCC_1; Right Ventral Posterior Cingulate Cortex | 12 | -54 | 14 | 2.91 | 0.004 |
| LH_Vis_6; Left Primary Visual Cortex | -12 | -67 | 7 | RH_Default_Temp_2; Right Temporal Pole | 51 | 7 | -18 | 2.90 | 0.004 |

| | | | | | | | | | |
|---|---|---|---|---|---|---|---|---|---|
| RH_Limbic_TempPole_1; Right Inferior Temporal Gyrus | 38 | 1 | -38 | LH_Default_Temp_2; Left Medial Temporal Gyrus | -58 | -32 | -1 | 2.83 | 0.005 |
| RH_Vis_7; Right Visual Association Cortex | 36 | -82 | 16 | RH_Default_pCunPCC_2; Right Ventral Posterior Cingulate Cortex | 7 | -52 | 31 | 2.79 | 0.006 |
| RH_SalVentAttn_TempOccPar_1; Right Superior Temporal Gyrus | 58 | -42 | 13 | LH_Default_pCunPCC_1; Left Ventral Posterior Cingulate Cortex | -11 | -56 | 13 | 2.76 | 0.007 |
| LH_Vis_6; Left Primary Visual Cortex | -12 | -67 | 7 | RH_SalVentAttn_TempOccPar_1; Right Superior Temporal Gyrus | 58 | -42 | 13 | 2.73 | 0.007 |
| LH_SomMot_4; Left PreMotor & Supplementary Motor Area | -55 | -8 | 34 | LH_DorsAttn_Post_6; Left Visual Motor Cortex | -22 | -51 | 66 | -2.70 | 0.008 |
| LH_Vis_7; Left Visual Association Cortex | -47 | -71 | 11 | RH_Cont_PFCl_2; Right Dorsolateral Prefrontal Cortex | 45 | 39 | 15 | 2.66 | 0.009 |
| RH_Vis_7; Right Visual Association Cortex | 36 | -82 | 16 | RH_SalVentAttn_FrOperIns_1; Right Insula | 40 | 8 | 1 | 2.63 | 0.010 |
| RH_SalVentAttn_Med_2; Right PreMotor & Supplementary Motor Area | 7 | 6 | 52 | LH_Limbic_TempPole_1; Left Temporal Pole | -32 | 2 | -37 | 2.60 | 0.010 |
| **Eyes-Closed Alpha** | | | | | | | | | |
| **ROI1 Labels (Schaefer; Talairach)** | **X** | **Y** | **Z** | **ROI2 Labels (Schaefer; Talairach)** | **X** | **Y** | **Z** | **t** | **p (unc)** |
| RH_Vis_4; Right Secondary Visual Cortex | 22 | -96 | -5 | LH_Default_Par_2; Left Angular Gyrus | -48 | -63 | 35 | 3.27 | 0.001 |
| RH_Vis_6; Right Secondary Visual Cortex | 17 | -57 | 5 | RH_SalVentAttn_Med_2; Right PreMotor & Supplementary Motor Area | 7 | 6 | 52 | 3.15 | 0.002 |
| LH_Cont_PFCl_1; Left Dorsolateral Prefrontal Cortex | -43 | 33 | 21 | RH_Cont_Par_2; Right Angular Gyrus | 45 | -63 | 46 | -3.11 | 0.002 |
| RH_SomMot_4; Right Primary Motor Cortex | 58 | -5 | 31 | RH_SalVentAttn_Med_2; Right PreMotor & Supplementary Motor Area | 7 | 6 | 52 | 3.00 | 0.003 |
| RH_Vis_2; Right Fusiform Cortex | 27 | -66 | -12 | LH_Default_pCunPCC_1; Left Ventral Posterior Cingulate Cortex | -11 | -56 | 13 | 2.87 | 0.005 |
| RH_Vis_6; Right Secondary Visual Cortex | 17 | -57 | 5 | LH_Default_Par_2; Left Angular Gyrus | -48 | -63 | 35 | 2.84 | 0.005 |
| RH_DorsAttn_Post_5; Right Visual Motor Cortex | 14 | -52 | 66 | LH_Default_pCunPCC_1; Left Ventral Posterior Cingulate Cortex | -11 | -56 | 13 | 2.81 | 0.006 |
| RH_Default_Par_1; Right Angular Gyrus | 55 | -51 | 31 | RH_Default_pCunPCC_2; Right Ventral Posterior Cingulate Cortex | 7 | -52 | 31 | 2.73 | 0.007 |
| RH_SomMot_6; Right Primary Motor Cortex | 41 | -22 | 60 | LH_Default_PFC_1; Left Pars Orbitalis | -35 | 21 | -11 | 2.72 | 0.007 |
| RH_Vis_5; Right Primary Visual Cortex | 8 | -76 | 5 | LH_Default_Par_2; Left Angular Gyrus | -48 | -63 | 35 | 2.71 | 0.008 |
| LH_SomMot_3; Left Primary Sensory Cortex | -54 | -12 | 13 | LH_Cont_PFCl_1; Left Dorsolateral Prefrontal Cortex | -43 | 33 | 21 | -2.68 | 0.008 |
| LH_Vis_3; Left Visual Association Cortex | -17 | -60 | -7 | LH_DorsAttn_PrCv_1; Left PreMotor & Supplementary Motor Area | -48 | 6 | 28 | 2.66 | 0.009 |
| LH_Default_Par_2; Left Angular Gyrus | -48 | -63 | 35 | RH_Default_pCunPCC_1; Right Ventral Posterior Cingulate Cortex | 12 | -54 | 14 | 2.65 | 0.009 |
| RH_DorsAttn_Post_4; Right Visual Motor Cortex | 27 | -67 | 51 | LH_Cont_pCun_1; Left Visual Motor Cortex | -9 | -73 | 38 | 2.65 | 0.009 |
| RH_SomMot_7; Right Primary Sensory Cortex | 30 | -37 | 64 | LH_Default_pCunPCC_1; Left Ventral Posterior Cingulate Cortex | -11 | -56 | 13 | 2.64 | 0.009 |
| LH_SomMot_1; Left Supramarginal Gyrus | -53 | -23 | 8 | RH_SalVentAttn_TempOccPar_1; Right Superior Temporal Gyrus | 58 | -42 | 13 | 2.62 | 0.010 |
| RH_SomMot_8; Right Primary Motor Cortex | 6 | -26 | 70 | LH_SalVentAttn_Med_2; Left Dorsal Posterior Cingulate Cortex | -11 | -34 | 45 | 2.62 | 0.010 |
| LH_Vis_7; Left Visual Association Cortex | -47 | -71 | 11 | LH_DorsAttn_Post_5; Left Visual Motor Cortex | -6 | -60 | 56 | 2.61 | 0.010 |
| **Eyes-Closed Beta** | | | | | | | | | |
| **ROI1 Labels (Schaefer; Talairach)** | **X** | **Y** | **Z** | **ROI2 Labels (Schaefer; Talairach)** | **X** | **Y** | **Z** | **t** | **p (unc)** |
| LH_Default_PFC_4; Left Anterior Prefrontal Cortex | -24 | 61 | -1 | RH_Default_Temp_2; Right Temporal Pole | 51 | 7 | -18 | 3.22 | 0.002 |
| RH_Cont_Par_2; Right Angular Gyrus | 45 | -63 | 46 | LH_Default_Temp_1; Left Medial Temporal Gyrus | -55 | -4 | -20 | 3.17 | 0.002 |

| | | | | | | | | | |
|---|---|---|---|---|---|---|---|---|---|
| LH_Cont_PFCl_1; Left Dorsolateral Prefrontal Cortex | -43 | 33 | 21 | RH_Default_Temp_2; Right Temporal Pole | 51 | 7 | -18 | 3.09 | 0.002 |
| LH_DorsAttn_Post_6; Left Visual Motor Cortex | -22 | -51 | 66 | LH_SalVentAttn_FrOperIns_1; Left Insula | -41 | -1 | -7 | 2.99 | 0.003 |
| LH_Cont_PFCl_1; Left Dorsolateral Prefrontal Cortex | -43 | 33 | 21 | LH_Cont_pCun_1; Left Visual Motor Cortex | -9 | -73 | 38 | 2.85 | 0.005 |
| LH_DorsAttn_FEF_1; Left PreMotor & Supplementary Motor Area | -26 | -3 | 59 | RH_Default_PFCdPFCm_1; Right Anterior Prefrontal Cortex | 7 | 48 | 1 | 2.77 | 0.006 |
| LH_DorsAttn_Post_2; Left Supramarginal Gyrus | -57 | -25 | 39 | LH_DorsAttn_Post_3; Left Visual Motor Cortex | -24 | -68 | 49 | -2.73 | 0.007 |
| LH_Vis_3; Left Visual Association Cortex | -17 | -60 | -7 | RH_Cont_PFCl_3; Right Dorsolateral Prefrontal Cortex | 32 | 46 | 29 | -2.71 | 0.008 |
| LH_DorsAttn_Post_6; Left Visual Motor Cortex | -22 | -51 | 66 | LH_Default_Temp_1; Left Medial Temporal Gyrus | -55 | -4 | -20 | 2.67 | 0.009 |
| LH_SalVentAttn_Med_2; Left Dorsal Posterior Cingulate Cortex | -11 | -34 | 45 | LH_Limbic_TempPole_2; Left Inferior Temporal Gyrus | -57 | -33 | -21 | 2.67 | 0.009 |
| LH_Cont_PFCl_1; Left Dorsolateral Prefrontal Cortex | -43 | 33 | 21 | RH_Default_PFCdPFCm_2; Right Dorsolateral Prefrontal Cortex | 11 | 50 | 39 | 2.67 | 0.009 |
| RH_Limbic_TempPole_1; Right Inferior Temporal Gyrus | 38 | 1 | -38 | RH_Default_pCunPCC_1; Right Ventral Posterior Cingulate Cortex | 12 | -54 | 14 | 2.65 | 0.009 |
| RH_SalVentAttn_Med_2; Right PreMotor & Supplementary Motor Area | 7 | 6 | 52 | LH_Cont_PFCl_1; Left Dorsolateral Prefrontal Cortex | -43 | 33 | 21 | 2.64 | 0.009 |
| RH_DorsAttn_Post_5; Right Visual Motor Cortex | 14 | -52 | 66 | LH_Default_pCunPCC_1; Left Ventral Posterior Cingulate Cortex | -11 | -56 | 13 | 2.63 | 0.009 |
| LH_SalVentAttn_Med_3; Left PreMotor & Supplementary Motor Area | -6 | 4 | 62 | RH_Cont_PFCl_3; Right Dorsolateral Prefrontal Cortex | 32 | 46 | 29 | 2.63 | 0.010 |
| RH_Vis_1; Right Fusiform Cortex | 32 | -31 | -22 | LH_DorsAttn_Post_3; Left Visual Motor Cortex | -24 | -68 | 49 | 2.63 | 0.010 |
| RH_DorsAttn_Post_4; Right Visual Motor Cortex | 27 | -67 | 51 | LH_SalVentAttn_Med_3; Left PreMotor & Supplementary Motor Area | -6 | 4 | 62 | -2.62 | 0.010 |
| LH_SomMot_2; Left Primary Sensory Cortex | -37 | -21 | 16 | LH_Limbic_OFC_1; Left Orbitofrontal Cortex | -14 | 32 | -20 | -2.60 | 0.010 |

**Eyes-Closed Gamma**

| ROI1 Labels (Schaefer; Talairach) | X | Y | Z | ROI2 Labels (Schaefer; Talairach) | X | Y | Z | t | p (unc) |
|---|---|---|---|---|---|---|---|---|---|
| LH_Vis_5; Left Secondary Visual Cortex | -5 | -92 | -2 | RH_Cont_PFCl_3; Right Dorsolateral Prefrontal Cortex | 32 | 46 | 29 | -4.07 | 8.18E-05 |
| LH_Cont_pCun_1; Left Visual Motor Cortex | -9 | -73 | 38 | RH_Cont_PFCl_2; Right Dorsolateral Prefrontal Cortex | 45 | 39 | 15 | -3.60 | 0.0005 |
| LH_SomMot_3; Left Primary Sensory Cortex | -54 | -12 | 13 | RH_DorsAttn_Post_1; Right Visual Association Cortex | 50 | -62 | 16 | -3.55 | 0.0005 |
| LH_Default_PFC_5; Left Dorsolateral Prefrontal Cortex | -9 | 48 | 41 | RH_Default_PFCv_1; Right Pars Orbitalis | 35 | 26 | -15 | -3.52 | 0.0006 |
| LH_Vis_2; Left Visual Association Cortex | -26 | -77 | -14 | RH_SomMot_2; Right Insula | 40 | -15 | 15 | -3.40 | 0.0009 |
| LH_Vis_5; Left Secondary Visual Cortex | -5 | -92 | -2 | RH_Default_PFCdPFCm_1; Right Anterior Prefrontal Cortex | 7 | 48 | 1 | -3.40 | 0.0009 |
| RH_Vis_8; Right Visual Association Cortex | 13 | -86 | 29 | RH_Cont_PFCl_2; Right Dorsolateral Prefrontal Cortex | 45 | 39 | 15 | -3.27 | 0.001 |
| RH_Cont_pCun_1; Right Visual Motor Cortex | 9 | -66 | 43 | RH_Default_PFCv_2; Right Broca's Area/ Pars Triangularis | 51 | 28 | 0 | -3.23 | 0.002 |
| RH_Vis_8; Right Visual Association Cortex | 13 | -86 | 29 | RH_Default_PFCv_2; Right Broca's Area/ Pars Triangularis | 51 | 28 | 0 | -3.21 | 0.002 |
| LH_Limbic_TempPole_2; Left Inferior Temporal Gyrus | -57 | -33 | -21 | RH_Default_PFCv_2; Right Broca's Area/ Pars Triangularis | 51 | 28 | 0 | -3.19 | 0.002 |
| LH_SomMot_4; Left PreMotor & Supplementary Motor Area | -55 | -8 | 34 | RH_SomMot_7; Right Primary Sensory Cortex | 30 | -37 | 64 | -3.18 | 0.002 |
| RH_DorsAttn_FEF_1; Right PreMotor & Supplementary Motor Area | 28 | -3 | 59 | RH_SalVentAttn_TempOccPar_2; Right Supramarginal Gyrus | 61 | -26 | 27 | 3.16 | 0.002 |
| RH_Cont_PFCl_2; Right Dorsolateral Prefrontal Cortex | 45 | 39 | 15 | RH_Default_pCunPCC_2; Right Ventral Posterior Cingulate Cortex | 7 | -52 | 31 | -3.14 | 0.002 |

| | | | | | | | | | |
|---|---|---|---|---|---|---|---|---|---|
| LH_Vis_4; Left Secondary Visual Cortex | -27 | -95 | -4 | RH_Default_PFCdPFCm_1; Right Anterior Prefrontal Cortex | 7 | 48 | 1 | -3.13 | 0.002 |
| LH_Limbic_TempPole_2; Left Inferior Temporal Gyrus | -57 | -33 | -21 | RH_Default_PFCv_1; Right Pars Orbitalis | 35 | 26 | -15 | -3.12 | 0.002 |
| LH_SomMot_4; Left PreMotor & Supplementary Motor Area | -55 | -8 | 34 | RH_DorsAttn_Post_5; Right Visual Motor Cortex | 14 | -52 | 66 | -3.12 | 0.002 |
| LH_Limbic_TempPole_2; Left Inferior Temporal Gyrus | -57 | -33 | -21 | RH_Cont_PFCl_1; Right Anterior Prefrontal Cortex | 30 | 58 | -3 | -3.11 | 0.002 |
| LH_SalVentAttn_Med_3; Left PreMotor & Supplementary Motor Area | -6 | 4 | 62 | LH_Default_PFC_1; Left Pars Orbitalis | -35 | 21 | -11 | 3.09 | 0.002 |
| LH_Vis_5; Left Secondary Visual Cortex | -5 | -92 | -2 | RH_Cont_PFCl_2; Right Dorsolateral Prefrontal Cortex | 45 | 39 | 15 | -3.04 | 0.003 |
| LH_Vis_5; Left Secondary Visual Cortex | -5 | -92 | -2 | RH_Default_PFCdPFCm_2; Right Dorsolateral Prefrontal Cortex | 11 | 50 | 39 | -3.02 | 0.003 |
| RH_DorsAttn_Post_4; Right Visual Motor Cortex | 27 | -67 | 51 | RH_Default_PFCv_2; Right Broca's Area/ Pars Triangularis | 51 | 28 | 0 | -3.01 | 0.003 |
| LH_SomMot_1; Left Supramarginal Gyrus | -53 | -23 | 8 | RH_Default_PFCdPFCm_3; Right Frontal Eye Fields | 26 | 24 | 50 | -2.98 | 0.003 |
| LH_SomMot_1; Left Supramarginal Gyrus | -53 | -23 | 8 | RH_Cont_PFCl_3; Right Dorsolateral Prefrontal Cortex | 32 | 46 | 29 | -2.97 | 0.004 |
| LH_Vis_5; Left Secondary Visual Cortex | -5 | -92 | -2 | RH_Limbic_OFC_1; Right Orbitofrontal Cortex | 12 | 35 | -20 | -2.91 | 0.004 |
| RH_Limbic_OFC_1; Right Orbitofrontal Cortex | 12 | 35 | -20 | LH_Default_PFC_6; Left PreMotor & Supplementary Motor Area | -41 | 14 | 48 | -2.90 | 0.004 |
| RH_Vis_3; Right Fusiform Cortex | 49 | -60 | -11 | LH_SalVentAttn_ParOper_1; Left Supramarginal Gyrus | -59 | -38 | 29 | -2.88 | 0.005 |
| LH_SomMot_3; Left Primary Sensory Cortex | -54 | -12 | 13 | RH_SalVentAttn_TempOccPar_1; Right Superior Temporal Gyrus | 58 | -42 | 13 | -2.87 | 0.005 |
| LH_SomMot_6; Left Primary Motor Cortex | -6 | -29 | 70 | LH_Default_PFC_6; Left PreMotor & Supplementary Motor Area | -41 | 14 | 48 | -2.87 | 0.005 |
| LH_SomMot_4; Left PreMotor & Supplementary Motor Area | -55 | -8 | 34 | RH_SalVentAttn_Med_1; Right Dorsal Posterior Cingulate Cortex | 11 | -31 | 45 | -2.86 | 0.005 |
| LH_Vis_6; Left Primary Visual Cortex | -12 | -67 | 7 | RH_Default_PFCdPFCm_1; Right Anterior Prefrontal Cortex | 7 | 48 | 1 | -2.85 | 0.005 |
| LH_Vis_5; Left Secondary Visual Cortex | -5 | -92 | -2 | RH_Default_PFCv_2; Right Broca's Area/ Pars Triangularis | 51 | 28 | 0 | -2.85 | 0.005 |
| RH_Cont_PFCmp_1; Right Frontal Eye Fields | 6 | 28 | 30 | RH_Default_PFCv_2; Right Broca's Area/ Pars Triangularis | 51 | 28 | 0 | -2.84 | 0.005 |
| LH_DorsAttn_Post_2; Left Supramarginal Gyrus | -57 | -25 | 39 | RH_Cont_PFCl_4; Right Frontal Eye Fields | 43 | 16 | 45 | -2.84 | 0.005 |
| LH_SalVentAttn_FrOperIns_1; Left Insula | -41 | -1 | -7 | RH_SalVentAttn_TempOccPar_1; Right Superior Temporal Gyrus | 58 | -42 | 13 | -2.83 | 0.005 |
| LH_SomMot_4; Left PreMotor & Supplementary Motor Area | -55 | -8 | 34 | RH_SomMot_8; Right Primary Motor Cortex | 6 | -26 | 70 | -2.83 | 0.005 |
| LH_SomMot_3; Left Primary Sensory Cortex | -54 | -12 | 13 | RH_SalVentAttn_TempOccPar_2; Right Supramarginal Gyrus | 61 | -26 | 27 | -2.81 | 0.006 |
| LH_SomMot_5; Left Primary Motor Cortex | -39 | -23 | 59 | LH_SalVentAttn_FrOperIns_1; Left Insula | -41 | -1 | -7 | 2.81 | 0.006 |
| LH_DorsAttn_Post_6; Left Visual Motor Cortex | -22 | -51 | 66 | RH_Default_pCunPCC_2; Right Ventral Posterior Cingulate Cortex | 7 | -52 | 31 | 2.81 | 0.006 |
| RH_Vis_5; Right Primary Visual Cortex | 8 | -76 | 5 | RH_Cont_PFCl_2; Right Dorsolateral Prefrontal Cortex | 45 | 39 | 15 | -2.80 | 0.006 |
| LH_DorsAttn_Post_1; Left Fusiform Cortex | -47 | -58 | -13 | RH_Cont_PFCmp_1; Right Frontal Eye Fields | 6 | 28 | 30 | -2.79 | 0.006 |
| LH_SomMot_5; Left Primary Motor Cortex | -39 | -23 | 59 | LH_Default_Temp_1; Left Medial Temporal Gyrus | -55 | -4 | -20 | 2.79 | 0.006 |
| LH_Cont_pCun_1; Left Visual Motor Cortex | -9 | -73 | 38 | RH_Cont_PFCl_3; Right Dorsolateral Prefrontal Cortex | 32 | 46 | 29 | -2.78 | 0.006 |
| LH_SomMot_2; Left Primary Sensory Cortex | -37 | -21 | 16 | RH_Cont_PFCl_2; Right Dorsolateral Prefrontal Cortex | 45 | 39 | 15 | -2.78 | 0.006 |
| RH_DorsAttn_Post_2; Right Primary Sensory Cortex | 50 | -24 | 42 | RH_DorsAttn_FEF_1; Right PreMotor & Supplementary Motor Area | 28 | -3 | 59 | 2.78 | 0.006 |
| LH_SomMot_3; Left Primary Sensory Cortex | -54 | -12 | 13 | RH_Cont_PFCl_2; Right Dorsolateral Prefrontal Cortex | 45 | 39 | 15 | -2.78 | 0.006 |

| | | | | | | | | | |
|---|---|---|---|---|---|---|---|---|---|
| LH_Vis_5; Left Secondary Visual Cortex | -5 | -92 | -2 | RH_Cont_PFCl_1; Right Anterior Prefrontal Cortex | 30 | 58 | -3 | -2.78 | 0.006 |
| LH_Vis_1; Left Fusiform Cortex | -26 | -34 | -17 | RH_Default_PFCdPFCm_1; Right Anterior Prefrontal Cortex | 7 | 48 | 1 | -2.77 | 0.006 |
| LH_SomMot_4; Left PreMotor & Supplementary Motor Area | -55 | -8 | 34 | RH_Limbic_OFC_1; Right Orbitofrontal Cortex | 12 | 35 | -20 | -2.76 | 0.007 |
| LH_Vis_9; Left Visual Association Cortex | -6 | -82 | 26 | RH_Cont_PFCl_3; Right Dorsolateral Prefrontal Cortex | 32 | 46 | 29 | -2.76 | 0.007 |
| LH_Default_PFC_5; Left Dorsolateral Prefrontal Cortex | -9 | 48 | 41 | RH_Default_Temp_1; Right Medial Temporal Gyrus | 62 | -23 | -19 | -2.76 | 0.007 |
| RH_Cont_PFCl_2; Right Dorsolateral Prefrontal Cortex | 45 | 39 | 15 | LH_Default_Temp_2; Left Medial Temporal Gyrus | -58 | -32 | -1 | -2.74 | 0.007 |
| LH_SalVentAttn_FrOperIns_1; Left Insula | -41 | -1 | -7 | RH_Default_PFCv_1; Right Pars Orbitalis | 35 | 26 | -15 | -2.74 | 0.007 |
| RH_Vis_7; Right Visual Association Cortex | 36 | -82 | 16 | RH_Cont_PFCl_4; Right Frontal Eye Fields | 43 | 16 | 45 | -2.73 | 0.007 |
| RH_Limbic_TempPole_1; Right Inferior Temporal Gyrus | 38 | 1 | -38 | RH_Cont_PFCl_4; Right Frontal Eye Fields | 43 | 16 | 45 | -2.73 | 0.007 |
| RH_SomMot_6; Right Primary Motor Cortex | 41 | -22 | 60 | LH_Default_PFC_6; Left PreMotor & Supplementary Motor Area | -41 | 14 | 48 | -2.73 | 0.007 |
| RH_SalVentAttn_FrOperIns_1; Right Insula | 40 | 8 | 1 | LH_Default_PFC_5; Left Dorsolateral Prefrontal Cortex | -9 | 48 | 41 | -2.72 | 0.007 |
| RH_Cont_PFCl_2; Right Dorsolateral Prefrontal Cortex | 45 | 39 | 15 | RH_Cont_pCun_1; Right Visual Motor Cortex | 9 | -66 | 43 | -2.71 | 0.008 |
| RH_DorsAttn_FEF_1; Right PreMotor & Supplementary Motor Area | 28 | -3 | 59 | RH_Cont_Par_1; Right Supramarginal Gyrus | 57 | -39 | 44 | 2.70 | 0.008 |
| RH_DorsAttn_Post_5; Right Visual Motor Cortex | 14 | -52 | 66 | RH_Default_PFCdPFCm_3; Right Frontal Eye Fields | 26 | 24 | 50 | -2.70 | 0.008 |
| RH_DorsAttn_Post_4; Right Visual Motor Cortex | 27 | -67 | 51 | RH_Cont_PFCl_2; Right Dorsolateral Prefrontal Cortex | 45 | 39 | 15 | -2.68 | 0.008 |
| LH_Vis_1; Left Fusiform Cortex | -26 | -34 | -17 | RH_Default_Temp_2; Right Temporal Pole | 51 | 7 | -18 | -2.68 | 0.008 |
| LH_SomMot_2; Left Primary Sensory Cortex | -37 | -21 | 16 | RH_Cont_PFCl_4; Right Frontal Eye Fields | 43 | 16 | 45 | -2.67 | 0.009 |
| LH_SomMot_1; Left Supramarginal Gyrus | -53 | -23 | 8 | RH_DorsAttn_Post_1; Right Visual Association Cortex | 50 | -62 | 16 | -2.66 | 0.009 |
| LH_SomMot_1; Left Supramarginal Gyrus | -53 | -23 | 8 | RH_Default_Par_1; Right Angular Gyrus | 55 | -51 | 31 | -2.65 | 0.009 |
| LH_SomMot_5; Left Primary Motor Cortex | -39 | -23 | 59 | RH_SomMot_8; Right Primary Motor Cortex | 6 | -26 | 70 | -2.65 | 0.009 |
| LH_SalVentAttn_PFCl_1; Left Anterior Prefrontal Cortex | -30 | 44 | 30 | RH_SalVentAttn_Med_1; Right Dorsal Posterior Cingulate Cortex | 11 | -31 | 45 | -2.64 | 0.009 |
| RH_SalVentAttn_FrOperIns_1; Right Insula | 40 | 8 | 1 | RH_Cont_PFCmp_1; Right Frontal Eye Fields | 6 | 28 | 30 | -2.64 | 0.009 |
| LH_SalVentAttn_ParOper_1; Left Supramarginal Gyrus | -59 | -38 | 29 | RH_Cont_PFCl_3; Right Dorsolateral Prefrontal Cortex | 32 | 46 | 29 | -2.64 | 0.009 |
| LH_Vis_6; Left Primary Visual Cortex | -12 | -67 | 7 | RH_Cont_PFCl_2; Right Dorsolateral Prefrontal Cortex | 45 | 39 | 15 | -2.64 | 0.009 |
| LH_SomMot_4; Left PreMotor & Supplementary Motor Area | -55 | -8 | 34 | RH_Cont_PFCl_4; Right Frontal Eye Fields | 43 | 16 | 45 | -2.63 | 0.010 |
| LH_SomMot_5; Left Primary Motor Cortex | -39 | -23 | 59 | LH_SomMot_6; Left Primary Motor Cortex | -6 | -29 | 70 | -2.63 | 0.010 |
| LH_SomMot_1; Left Supramarginal Gyrus | -53 | -23 | 8 | RH_SalVentAttn_FrOperIns_1; Right Insula | 40 | 8 | 1 | -2.62 | 0.010 |
| LH_SalVentAttn_PFCl_1; Left Anterior Prefrontal Cortex | -30 | 44 | 30 | RH_SalVentAttn_TempOccPar_2; Right Supramarginal Gyrus | 61 | -26 | 27 | -2.61 | 0.010 |
| LH_SalVentAttn_Med_2; Left Dorsal Posterior Cingulate Cortex | -11 | -34 | 45 | LH_Default_PFC_6; Left PreMotor & Supplementary Motor Area | -41 | 14 | 48 | -2.61 | 0.010 |
| LH_Vis_2; Left Visual Association Cortex | -26 | -77 | -14 | RH_Default_Temp_3; Right Superior Temporal Gyrus | 57 | -26 | -2 | -2.61 | 0.010 |
| RH_DorsAttn_Post_4; Right Visual Motor Cortex | 27 | -67 | 51 | RH_Default_PFCv_1; Right Pars Orbitalis | 35 | 26 | -15 | -2.60 | 0.010 |
| LH_SomMot_6; Left Primary Motor Cortex | -6 | -29 | 70 | RH_Default_pCunPCC_2; Right Ventral Posterior Cingulate Cortex | 7 | -52 | 31 | 2.60 | 0.010 |

$p \leq 0.01$ uncorrected
